# Development and validation of a high-throughput qPCR platform for the detection of soil-transmitted helminth infections

**DOI:** 10.1101/2023.11.27.23299079

**Authors:** Nils Pilotte, Victor Omballa, Monica Voss, Leah Padgett, Malathi Manuel, Jeanne L. Goodman, Tim Littlewood, Zayina Zondervenni Manoharan, Lisette van Lieshout, Jaco Verweij, Manigandan Sekar, Ajith Kumar Muthukumar, Gretchen Walch, Andrew Gonzalez, Sean R Galagan, Sitara Swarna Rao Ajjampur, Moudachirou Ibikounlé, Steven A Williams, Doug Rains, Ushashi Dadwal, Judd L Walson

## Abstract

**Background:** Historically, soil-transmitted helminth (STH) control and prevention strategies have relied on mass drug administration efforts targeting preschool and school-aged children. While these efforts have succeeded in reducing morbidity associated with STH infection, recent modeling efforts have suggested that expanding intervention to treatment of the entire community could achieve transmission interruption in some settings. Testing the feasibility of such an approach requires large-scale clinical trials, such as the DeWorm3 cluster randomized trial. In addition, accurate interpretation of trial outcomes will require diagnostic platforms capable of accurately determining infection prevalence, particularly as infection intensity is reduced, at large population scale and with significant throughput. Here, we describe the development and validation of a multi-site, high-throughput molecular testing platform.

**Methodology/Principal Findings:** Through the development, selection, and validation of appropriate controls, we have successfully created and evaluated the performance of a testing platform capable of the semi-automated, high-throughput detection of four species of STH in human stool samples. Comparison of this platform with singleplex reference assays for the detection of these same pathogens has demonstrated comparable performance metrics across multiple testing locations, with index assay accuracy measuring at or above 99.5% and 98.1% for each target species at the level of the technical replicate and individual extraction respectively. Through the implementation of a rigorous validation program, we have developed a diagnostic platform capable of providing the necessary throughput and performance needed to meet the needs of the DeWorm3 cluster randomized trial and other large-scale operational research efforts for STH.

**Importance:** Current models predict that an expansion of intervention efforts to include community-wide treatment strategies could, in certain geographies, result in the elimination of soil-transmitted helminths as a public health problem. Large-scale operational research efforts to reduce the burden of STH such as the DeWorm3 cluster randomized trial have been organized to evaluate these predictions, but such evaluations require a high throughput testing strategy capable of providing sensitive, specific, and accurate results. To meet this need, we have developed a multiplexed diagnostic platform that was rigorously evaluated across multiple testing locations using a novel validation plan with pre-defined performance characteristics. Our successful validation of this platform has provided the community with a tool capable of meeting the diagnostic requirements of large-scale operational research efforts. Furthermore, this strategy provides a blueprint for the development of similar platforms adaptable for use with other neglected tropical disease programs evaluating other intervention strategies.

## Introduction

Soil-transmitted helminths (STH) are a leading cause of human disease, infecting over 1.5 billion individuals worldwide, with the majority of infections occurring in low and middle-income countries [1,2]. Large-scale programs designed to control these infections have relied heavily on mass drug administration (MDA)-based deworming efforts, targeting populations at high risk of morbidity, namely preschool and school-aged children (PSAC and SAC) and women of reproductive age. Historically, these programs have utilized coproscopy-based techniques (such as the Kato-Katz method) to determine the prevalence and intensity of infection within a population, in turn informing decisions related to intervention frequency and duration [3]. This intervention and monitoring strategy has been shown to effectively reduce morbidity in multiple geographies [4]. However, such microscopy-based diagnostics are known to lack sensitivity. This shortcoming is particularly pronounced when infection intensities are low, as the sensitivity of detection may drop below 50% [3]. Additionally, with programmatic consideration being given to the possible transition from morbidity control strategies to strategies aimed at transmission interruption and/or the elimination of disease as a public health problem, diagnostics with high-throughput and improved performance characteristics are urgently needed. Although molecular assays such as quantitative polymerase chain reaction (qPCR) have been designed, tested, and used in research settings [3,4], the development and validation of a large-scale, high-throughput testing platform, exportable for use in multiple laboratories across both high-and low-infection intensity settings, has not been previously described.

The DeWorm3 project is a multi-country, community, cluster-randomized trial designed to determine the feasibility of successful transmission interruption of soil-transmitted helminth (STH) infections following sequential, high-coverage, community-wide mass-drug administration (cMDA) interventions [5]. This large-scale project includes study sites in Benin, India, and Malawi. As the primary discriminator of infection for this study, we developed and validated a large-scale, high-throughput, multi-site molecular testing platform for the detection of STH in human stool. In all three participating countries, a geographically defined administrative unit that included a population of approximately 80,000 – 140,000 individuals was censused and mapped. Each study site was divided into 40 clusters, which were randomized (1:1) to receive either the intervention (biannual cMDA with albendazole) or control treatment (standard of care MDA delivered to PSAC & SAC annually) for three years [5]. In total, approximately 300,000 stool samples were collected during the five-year study period.

To meet the testing needs of the DeWorm3 project, and to provide a diagnostic design strategy for future studies and for potential future programmatic use, we developed a high-throughput, molecular testing platform for the detection of *Ascaris lumbricoides*, *Necator americanus*, *Ancylostoma duodenale*, and *Trichuris trichiura*. To ensure the integrity of our approach, an a priori validation plan was developed in consultation with an external panel of independent evaluators. The high-throughput testing approach was standardized and validated at laboratories in the United States (Quantigen, Fishers, IN), India (Christian Medical College, Vellore), and Benin (Institut de Recherche Clinique du Bénin, Abomey-Calavi) through the incorporation of rigorous pre-validation steps (run controls) and quality control (QC) measures. Here we describe the validation and quality assurance process of a high-throughput qPCR testing platform for the detection of these STH targets. This detailed description of our validation methods demonstrates the robust approach used to test the DeWorm3 trial samples and provides a framework for the deployment of this or other similar large-scale, community-level testing platforms in the future.

## Methods

All experiments in the validation of this high-throughput molecular system utilized the standard set of extraction and qPCR methods described below. The full report of the validation plan is available in the supplementary materials (S1 appendix).

### Sample disruption and nucleic acid isolation

Isolation of DNA from STH eggs requires both physical disruption and chemical digestion. DNA extraction from *T. trichiura* eggs is particularly challenging due to their multi-layer structure: an outer vitelline layer, a middle chitinous layer that resists physical disruption, and an inner lipid layer that provides resistance to chemical breakdown [6]. In an attempt to overcome these challenges, we optimized a previously described bead-beating protocol to disrupt eggs within stool samples using a 96-well plate shaking system (OMNI International, Kennesaw, GA) [7,8]. After disruption, we isolated DNA using the semi-automated KingFisher Flex 96-well sample extractor (Thermo Fisher Scientific, Waltham, MA) with the MagMAX Microbiome Ultra Nucleic Acid Isolation Kit following the manufacturer’s suggested protocol (Thermo Fisher Scientific) (Fig 1) with a single modification. Namely, MVP II Binding Beads (Thermo Fisher Scientific) were used in place of those included with the MagMAX kit.

**Fig 1.**
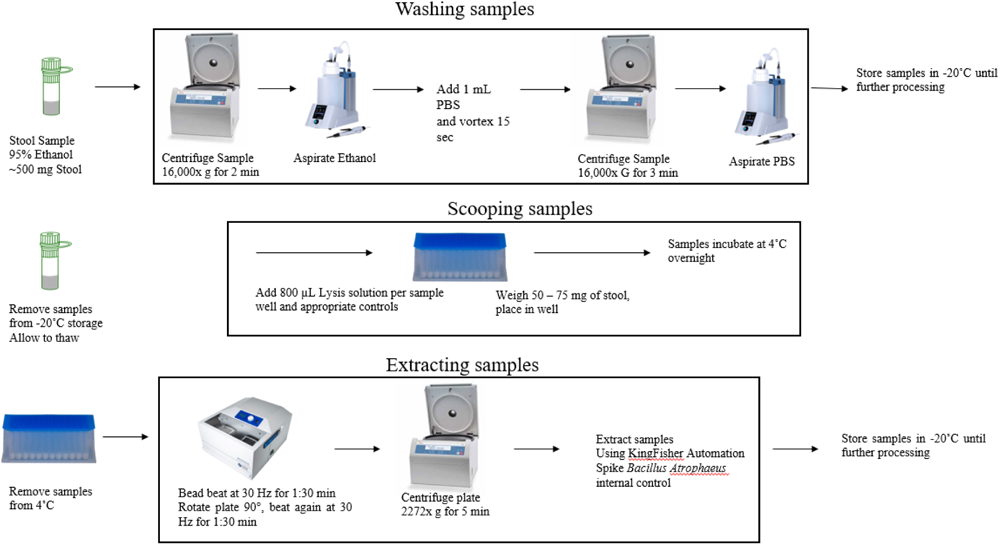
Procedure for isolation of DNA from stool samples. After sample collection, isolation of DNA occurred in three steps. Samples were first washed, to remove the ethanol used for storage and preservation. Samples were then scooped into OMNI 96-well 1.4 mm ceramic bead-beating plates. Following overnight incubation, DNA extraction was performed.

### Multiplexing of qPCR assays

Four previously developed primer/probe-based assays, each targeting a highly repetitive genomic element unique to a single STH species, were utilized in the development of our index assays [8,9]. Based on initial oligo optimization experiments, we selected primer and probe concentrations that yielded comparable C_T_ and ΔRn values between singleplex and multiplex formats when used on contrived mixed-infection controls (Table 1). Hydrolysis probes for each assay were labeled with either 6-FAM or Yakima Yellow reporters (IDT, Coralville, IA) and double-quenched with ZEN-IABkFQ chemistries (IDT). These fluorophore-quencher pairings were chosen due to the reduced background fluorescence and improved sensitivity which they offer relative to single-quenched probes [8]. Assays were then duplexed by pairing the *N. americanus* assay with the *T. trichiura* assay, and the *A. lumbricoides* assay with the *A. duodenale* assay. These pairings were chosen as *N. americanus* and *A. lumbricoides* were expected to represent the most common sources of infection across study sites. For this reason, it was decided that they should be separate, such that the likelihood of competition for reagents within a single qPCR reaction well would be reduced. Duplexed assays were then tested against a characterized panel of DNA samples previously extracted from stool collected in India. Results were compared to singleplex results obtained for the same samples (S1 Table). Through the process of duplexing, a single sample could be tested for the presence of all four STH targets in only two qPCR wells. To each of the two STH-specific duplexed assays, we added a third, primer-limited (50 nM each) ABY-labeled primer/probe-based assay designed to amplify an exogenous extraction/qPCR internal positive control (IPC): *Bacillus atrophaeus* bacteria (ZeptoMetrix, Buffalo, NY) [10]. The selection of this control is discussed in detail below. Details for all primers and probes used in the index assays are listed in Table 1.

**Table 1:**
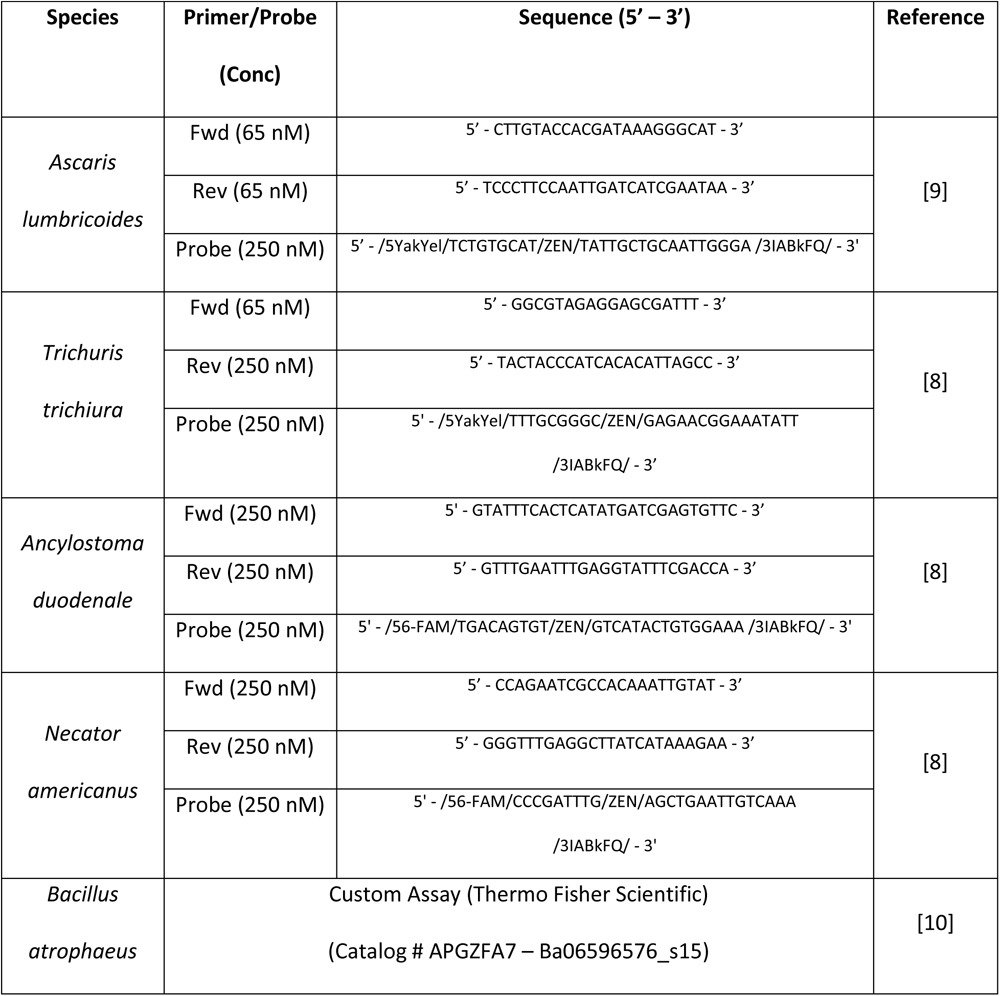
Primers and probes used in index assays.

### Selection of qPCR reagents and lyophilization

Sample-derived qPCR inhibitors can limit amplification of target STH gDNA [11]. To evaluate master mix performance and select an appropriate chemistry, naive stool samples, collected in Benin, were spiked with STH gDNA to generate qPCR sensitivity and amplification curves. Various commercially available master mixes were tested on the QuantStudio 7 Real-Time PCR System (Life Technologies, Carlsbad, CA) at Quantigen. All tested mixes were obtained from Thermo Fisher Scientific and included TaqMan Environmental Master Mix 2.0, Ambion Path-ID qPCR Master Mix, and TaqPath 1-Step RT-qPCR Master Mix, CG. As our goal was to lyophilize all qPCR reagents – master mix and assays – to ensure reagent stability during international shipment and subsequent storage, and to improve workflow efficiency and accuracy in the laboratory through the reduction of pipetting steps, Thermo Fisher customized a comparable formulation of the TaqPath 1-Step RT-qPCR Master Mix, CG as a TaqMan 5X Lyo-ready qPCR Master Mix with ROX and excipient. Testing identified no significant differences in detection sensitivity as a function of master mix tested across target species (S2 Table). Argonaut Manufacturing Services (Carlsbad, CA) produced 384-well custom-designed lyophilized qPCR plates (lyo-plates) in bulk. Following batch testing at Quantigen, these plates were disseminated to all testing laboratories for use with all DeWorm3 sample testing. Subsequently, a second batch of plates was required, which was produced and tested in an identical manner.

### Assay validation

#### Control selection and validation

Assay validation first requires control validation. Accordingly, appropriate controls were identified and selected for use with our high-throughput testing platform. Following selection, control validation focused on the performance of positive extraction controls (using both external and internal targets), and of appropriate qPCR controls.

An optimal external extraction control should reflect the performance characteristics and specific challenges of the real-world samples and targets being studied. *Ascaris suum* was selected as our platform’s external extraction control, as it closely mirrors one of the target organisms (*Ascaris lumbricoides)* and is commercially available (Excelsior Sentinel, Inc., Ithica, NY). *Ascaris lumbricoides* and *Ascaris suum* are genetically similar organisms, thought to potentially represent sub-species rather than unique species [12]. Furthermore, the previously described assay target used in our index assay has been shown to amplify both species equivalently [13,14]. For these reasons, batches of *A. suum* control aliquots were prepared by adding approximately 35,000 eggs to 6 g of commercially purchased STH-negative stool (BioIVT, Westbury, NY). These batches were combined with 21 mL of molecular grade ethanol and were thoroughly mixed using a handheld immersion homogenizer (OMNI International, Kennesaw, GA). Homogenization yielded an egg concentration of approximately 1.7 eggs/µL. From this initial batch, 25 *A. suum* controls were randomly selected, extracted (as described previously [10]), and tested in duplicate using qPCR to determine the expected C_T_ range (S3 Table). Based on these results, passing qualifications for validation of the *A. suum* control were defined as positive *A. suum* amplification with a C_T_ value of ≤ 23, a value approximating the upper bound of those seen during this initial testing.

Arguably, the most critical control is the internal positive control (IPC). When properly selected, this control serves as a per-sample extraction, recovery, and amplification control. For use with our testing platform, commercially available *Bacillus atrophaeus* was selected as the IPC. *B. atrophaeus* is a spore forming bacteria whose structure provides some resistance to lysis [15], making it an appropriate choice for use with STH eggs whose lysis requires mechanical disruption and chemical digestion. This bacterium is well-validated as a control for DNA extraction [10]. Given these characteristics, *B. atrophaeus* was commercially purchased (ZeptoMetrix, Buffalo, NY) for use with our platform [15]. Bacterial spores were spiked into 200 naive stool samples, and the *B. atrophaeus* IPC was validated using both index Assay 1 (*B. atrophaeus*, *N. americanus*, *T. trichiura*) and index Assay 2 (*B. atrophaeus*, *A. lumbricoides*, *A. duodenale*). Passing qualifications for validation of this control were defined as positive *B. atrophaeus* amplification with a C_T_ score ≤ 34 in both Assay 1 & Assay 2 in the absence of STH amplification.

Plasmid DNA was used as a positive PCR control to ensure reagent integrity and to act as a qPCR process control. Stocks of four plasmids, each containing a single copy of the target sequence for one of the four pathogens of interest, were prepared as previously described [9]. Plasmids were then combined into a single pool and diluted for testing at two masses (2 pg and 200 fg). Twenty-five replicates of each dilution were then tested using the index assays to verify that all plasmid targets performed consistently and allowed for predictable and reproducible amplification within an expected C_T_ value range (S4 Table). Passing qualifications for validation of this control were defined as < 2 standard deviations from the validation-defined mean C_T_ for each assay run with 100 fg of template. Thus, the C_T_ score cut-off for each species were determined as follows: *N. americanus* ≤ 25.2, *A. lumbricoides* ≤ 23.4, *A. duodenale* ≤ 23.5, and *T. trichiura* ≤ 21.95.

#### Preparation of spiked-stool standards

A panel of controls comprised of twenty STH-positive and five STH-negative standards was prepared at Smith College (Northampton, MA). Standards were created by spiking helminth-naive stool with field samples of known STH infection status for each species of STH. To create each standard, 500 mg aliquots of STH-containing stool, previously characterized in an unrelated study [3], were inoculated into separate 6-gram aliquots of commercially available, STH-negative bulk stool (BioIVT). Each spiked stool sample was then suspended in 21 mL of molecular grade ethanol and thoroughly mixed using a hand-held homogenizer to increase the uniformity of egg distribution. For each standard, a total of 75 aliquots averaging 85 mg of stool homogenate in 300 µL of ethanol were generated. Randomly selected aliquots of each standard were then tested at Smith College using singleplex qPCR assays to ensure the expected presence/absence of each target in each tested aliquot.

#### Characterization of spiked-stool standards

Three blinded aliquots of each standard (∼85 mg per aliquot) were extracted and qPCR tested in quadruplicate at Quantigen using the multiplexed index assays (25 standards x 3 aliquots x 4 technical replicates). An individual test result was considered positive if amplification occurred with a C_T_ value < 40, while a test was considered negative if amplification did not occur. Following testing, results generated at Quantigen were compared with singleplex assay results generated at Smith College. This comparison was performed to ensure that all standards were of sufficient quality to allow for consistent performance. With two exceptions (samples 15 and 18), samples producing discordant results either (1) across technical replicates tested by the index assays at Quantigen and/or (2) between aliquots tested by the multiplexed index assays and the original singleplex assays (hereafter the reference assays) were eliminated. Samples 15 and 18 were retained despite inconsistent detection of *A. lumbricoides* and *N. americanus* due to their consistent performance when tested with the *T. trichiura* assay and the desire to retain *T. trichiura* containing standards. Elimination of poorly performing samples resulted in a panel of 18 standards that were used in all further testing/validation (Table 2).

**Table 2.** Infection characteristics for spiked stool standards.

| Standard # | STH Status |  |  |  |
| --- | --- | --- | --- | --- |
|  | *T. trichiura* | *A. lumbricoides* | *N. americanus* | *A. duodenale* |
| 1 |  | X | X |  |
| 2 |  |  | X |  |
| 3 |  | X |  |  |
| 4 |  |  | X |  |
| 5 |  | X |  |  |
| 6 |  | X | X |  |
| 7 |  |  | X |  |
| 8 | X | X | X |  |
| 9 |  |  |  |  |
| 10 |  |  |  |  |
| 11 |  |  |  |  |
| 12 |  |  |  |  |
| 13 |  |  |  |  |
| 14 |  | X | X | X |
| 15 | X | (X) | X |  |
| 16 | X | X | X |  |
| 17 | X | X | X |  |
| 18 | X | X | (X) |  |

(X) indicate the presence of trace amounts of target that were inconsistently identified. For these samples, either a “positive” or a “negative” result was accepted as target concentrations fall at/near the limit of detection of the assay and therefore do not allow for consistently reproducible results.

#### Evaluation of assay accuracy

Assay accuracy was evaluated on the technical replicate level and the aliquot level using the above-described panel of contrived standards. Accuracy was calculated using the sum of true positives and true negatives as the numerator, and the total number of samples tested as the denominator (Equation 1). Three blinded aliquots (∼85 mg) of each of the 18 contrived standards were extracted and qPCR-tested in quadruplicate using the index assays (18 standards x 3 aliquots x 4 technical replicates). The true positive or negative status of each standard was defined by the initial characterization performed at Smith College using the singleplex reference assays (see “Preparation of spiked-stool standards” above). Evaluation of STH positivity at the aliquot level was defined as at least one positive amplification out of all technical replicates tested by the index assay. Well failures were not included in calculations of assay accuracy.

Equation 1. Accuracy calculations of index assay performance when testing spiked-stool standards.

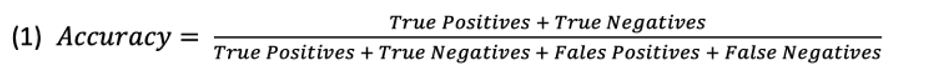

#### Evaluation of assay specificity

Assay specificity was assessed using 413 stool samples from Benin (213) and India (200) which initially tested negative for all species using the index assay. These samples were subsequently retested utilizing the reference assays, and specificity was calculated through comparison of these results.

#### Evaluation of assay limits of detection

A subset comprised of four spiked-stool standards was used to determine the limits of detection (LOD) for the index assays. LOD testing was possible, as matched microscopy data (Kato-Katz) were available for the field samples used in contrived sample creation. Using this information, the concentration of eggs per gram (EPG) of stool in the selected spiked stool standards could be estimated (Table 3). Extracted DNA for each standard was serially diluted 1:10 for a total of six dilution points. Each dilution point was tested by the index assays in 10 technical replicates (4 standards x 6 dilution points x 10 replicates), and the lowest concentration in which all 10 replicates generated a positive signal was identified. Starting at a concentration 2-fold higher than this concentration, five 1:2 serial dilutions were created, and 20 technical replicates of each dilution point were tested by qPCR (4 standards x 5 dilution points x 20 replicates). The LOD was determined as the lowest concentration at which at least 19/20 technical replicates amplified for the target STH species using Equation 2 below (S5 Table).

**Table 3.**
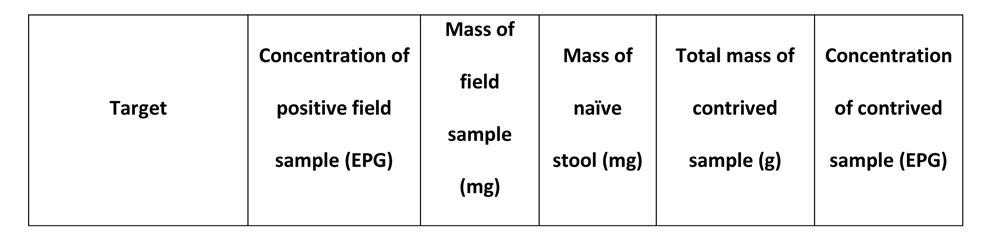
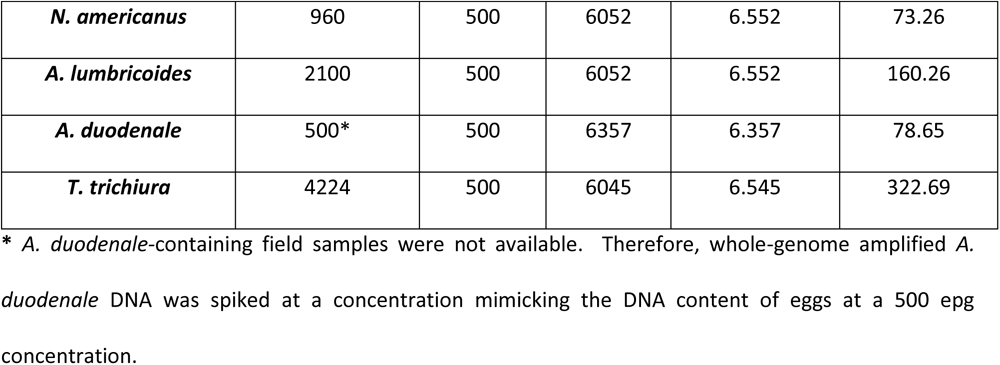
Characteristics of contrived standards used for limit of detection testing.

| Target | Concentration of<br>positive field<br>sample (EPG) | Mass of<br>field<br>sample<br>(mg) | Mass of<br>naïve<br>stool (mg) | Total mass of<br>contrived<br>sample (g) | Concentration<br>of contrived<br>sample (EPG) |
| --- | --- | --- | --- | --- | --- |
| ***N. americanus*** | 960 | 500 | 6052 | 6.552 | 73.26 |
| ***A. lumbricoides*** | 2100 | 500 | 6052 | 6.552 | 160.26 |
| ***A. duodenale*** | 500* | 500 | 6357 | 6.357 | 78.65 |
| ***T. trichiura*** | 4224 | 500 | 6045 | 6.545 | 322.69 |

\* *A. duodenale*-containing field samples were not available. Therefore, whole-genome amplified *A. duodenale* DNA was spiked at a concentration mimicking the DNA content of eggs at a 500 epg concentration.

Equation 2. Calculation of index assay limit of detection as determined by the testing of DNA extracts isolated from contrived stool standards.

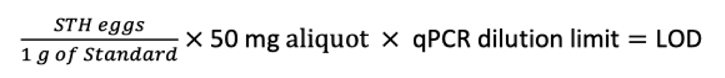

#### Assessment of sample heterogeneity

Sample heterogeneity was explored by selecting a random 10% of the first 2,000 tested baseline samples from Benin and extracting four aliquots of each sample. To test for site-dependent extraction differences, for each sample, two extractions were completed using the index assay protocol and two independent extractions were completed via the reference assay protocol. Each extraction was then tested using both index and reference assays. Testing generated a total of two data points per extraction (eight data points for each sample). (Fig 2).

**Fig 2.**
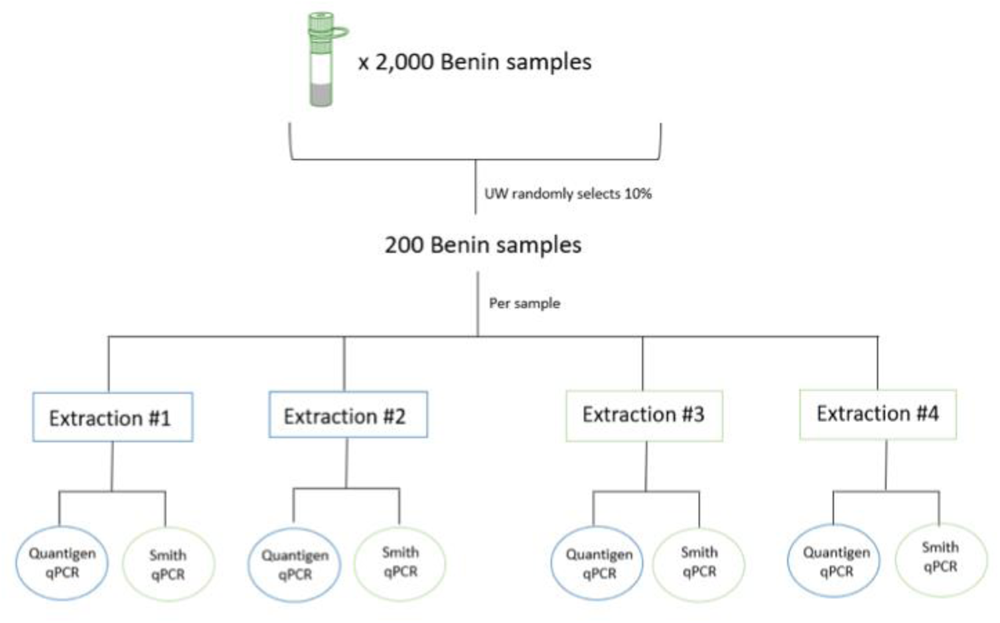
Assessment of sample heterogeneity. Illustration depicting how each sample used in the assessment of sample heterogeneity generated eight points of data. Blue outlines indicate the index protocol and green outlines indicate the reference protocol.

**Fig 3.**
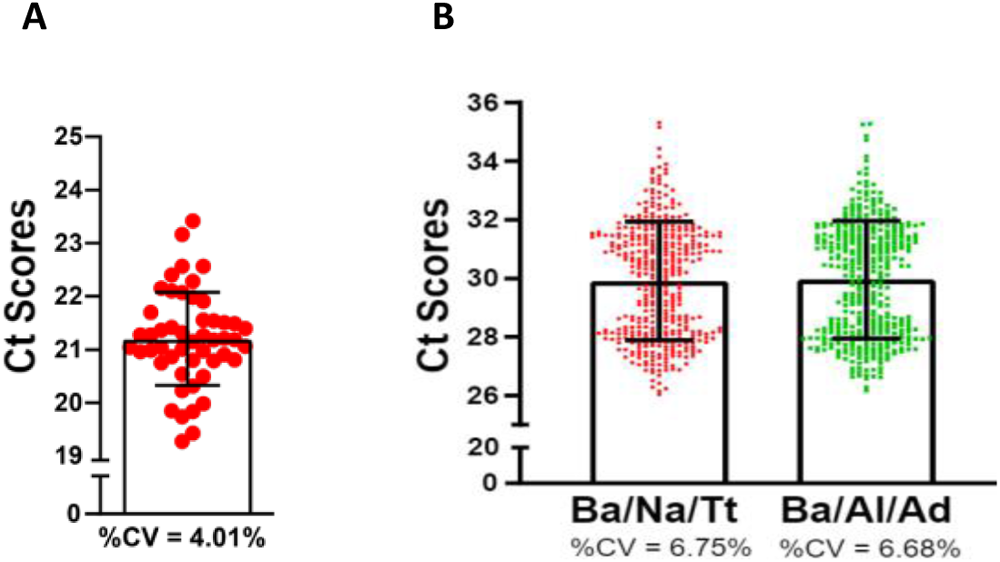
Results of control validation. **(A)** Dot plot depicting the range of C_T_ values for *A. suum* detection. The error bars represent the standard deviation. **(B)** Dot plot depicting C_T_ values for all *B. atrophaeus* results obtained when running both index assays on a panel of STH-negative, *B. atrophaeus-*positive samples. Error bars represent the standard deviation.

### Data description

All supporting data is included in the body of the manuscript or as part of the supplemental material.

## Results

### Control selection and validation

All run controls (internal and external extraction controls as well as qPCR controls) passed the predefined thresholds for validation. Passing qualifications to validate the performance of the *A. suum* external extraction control were defined as positive *A. suum* amplification with a C_T_ value of ≤ 23. The coefficient of variation (CV) for the *A. suum* positive extraction control was 4% (n = 50) with an average C_T_ of 21.14 (±0.85) (Fig 2a). Validation testing for the performance of the *B. atrophaeus* IPC resulted in a CV of 7% with an average C_T_ value of 29.96 (±2.02) for Assay 1 and an average C_T_ value of 30.00 (±2.00) for Assay 2 (Fig 2b), meeting the minimum requirements for validation. All qPCR positive controls also passed the predefined validation threshold, with C_T_ values falling below required values for all target species in all replicate reactions.

### Evaluation of assay accuracy

Extraction and testing of three independent aliquots of all contrived spiked-stool standards resulted in positive detection for each expected species in at least three out of the four qPCR replicates. In total, <1% of all qPCR wells failed to amplify as expected (Table 4). Assay accuracy at the technical replicate level met the minimum target with >99% accuracy for all assays. The *A. lumbricoides*, *N. americanus*, and *A. duodenale* assays met the minimum standard of 100% accuracy at the individual extraction level. However, one *T. trichiura* false positive result was seen among the 12 technical replicates tested, producing an individual extraction level accuracy for *T. trichiura* of 98.1% (Table 4, S6 Table).

**Table 4.**
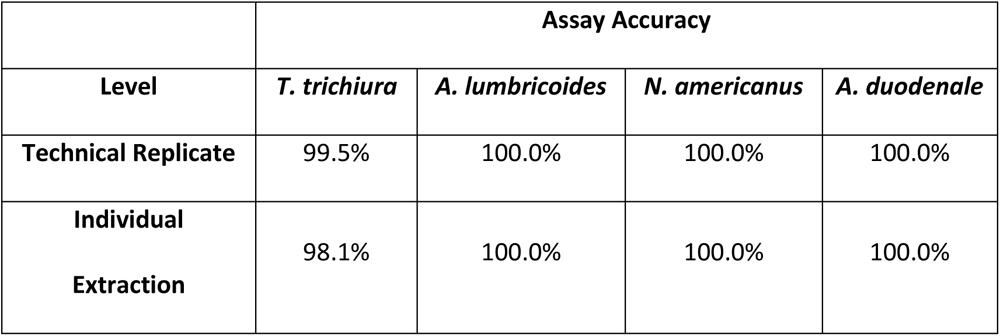
Index assay accuracy at the individual extraction and technical replicate levels.

Upon quadruplicate repeat testing at Quantigen, using both the index multiplex and reference singleplex assays (n = 8), the one false positive *T. trichiura* amplification event could not be reproduced. None of the STH-negative stool standards (n = 5) produced a positive result for any erroneous/unexpected target.

### Evaluation of assay specificity

Assay specificity was evaluated through testing, using the singleplex reference assays, of 413 samples that initially tested negative for all STH species at Quantigen. Reference assay testing of 213 samples from Benin produced three positive results (two *N. americanus* positive results and one *A. lumbricoides* positive result). To evaluate these discordant results, DNA extracts from all three discordant samples were retested using both liquid and lyophilized assays. On repeat testing, all three samples were confirmed to be positive in liquid and/or lyophilized testing formats. To further investigate aliquot-to-aliquot variability, three new aliquots of each discordant sample (three samples x three aliquots) were re-extracted using the index platform extraction procedure and tested by the index liquid assay in single qPCR replicates. All previously detected species were detected in at least two out of three aliquots.

Reference assay testing of 200 samples from India produced five *N. americanus* positive samples previously identified as negative during Quantigen’s initial screening efforts using the index assays. The DNA extracts for all five discordant samples were retested, in triplicate, using both the lyophilized index assay and the reference liquid assay. Results were again negative using the index procedures, but positive by reference methods. To investigate aliquot-to-aliquot variability, additional aliquots of the five discordant samples were then re-extracted. Three of the five samples were re-extracted in duplicate, and two samples underwent re-extraction of a single aliquot due to limitations in sample volume. All eight re-extraction products ([three samples x two extractions] + [two samples x one extraction]) were tested via the lyophilized index assay in duplicate, and all samples confirmed the detection of *N. americanus* in at least one retested extract. Based on these data, we calculated the index assay specificity at 98%.

### Evaluation of assay limits of detection

Through the testing of a dilution series of characterized samples, the lowest concentration at which at least 19/20 technical replicates amplified for each target STH species was identified. These concentrations were then used to calculate LODs, and the LOD for each assay was determined to be <1 EPG (Table 5).

**Table 5.**
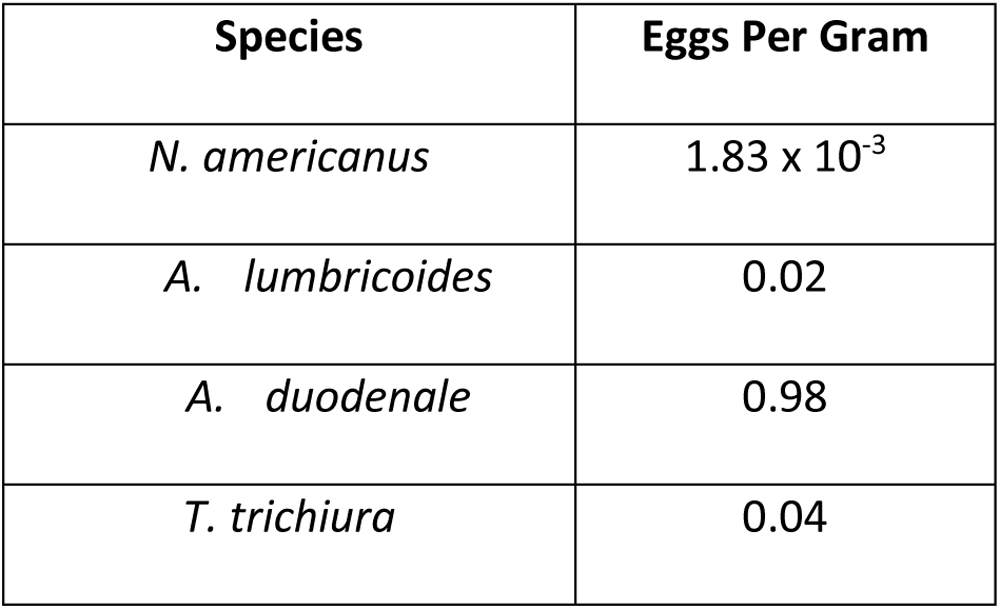
By species results of limit of detection testing.

| Species | Eggs Per Gram |
| --- | --- |
| *N. americanus* | $1.83 \times 10^{-3}$ |
| *A. lumbricoides* | 0.02 |
| *A. duodenale* | 0.98 |
| *T. trichiura* | 0.04 |

### Assessment of sample heterogeneity

To determine the potential impact of aliquot heterogeneity on the detection of target species from a single sample, 200 randomly selected samples (10%), from among the first 2,000 samples tested in Benin, were subjected to further testing. Four aliquots were taken from each of these 200 samples and DNA was independently extracted from each aliquot with two extractions occurring using reference procedures and two extractions occurring using index procedures. Follow up qPCR analysis then occurred on each extraction product using both the index and reference assays. Minimal discordance was observed in three of the four target species; 8/200 *N. americanus* results discordant, 3/200 *T. trichiura* results discordant, and 10/200 A. *lumbricoides* results discordant. All testing for *A. duodenale* was concordant negative. The rate of discordance was similar across laboratories, assay methodologies, and extraction techniques (S7 Table). Extracts one, three, and four each detected STH species in 36/200 samples while extract two detected STH species in 43/200 samples. To further assess the performance of the reference assay and the index assay, concordance of STH detection was calculated across all extracts and assays. Overall concordance for each species was as follows: *N. americanus* – 98.5% concordant, *T. trichiura* – 100% concordant, *A. duodenale* – 100% concordant, and *A. lumbricoides* – 98.1% concordant.

## Discussion

As the global burden of disease due to STH infections declines, largely because of both improvements in sanitation infrastructure and increasing coverage of mass drug administration programs, accurate estimates of helminth prevalence are critical to inform policy and programs. Multiple studies have demonstrated that molecular assays are superior for the detection of infection and the quantification of prevalence and intensity across geographic settings and epidemiologic contexts [16]. As the prevalence and intensity of STH infections declines globally, programs shift towards the elimination of STH as a public health problem, or to the interruption of disease transmission; the improved sensitivity and accuracy of molecular assays become critical for programs to achieve and confirm success. Unfortunately, platforms for the use of molecular diagnostics for STH at scale have not been previously developed or commercialized and these assays have largely been confined to research use. The small-scale use of these assays has meant that there has been little efficiency of scale, allowing costs to remain high and methods, operating procedures, and QA/QC methodologies to remain unstandardized.

Here we describe a well-validated, standardized, and scalable high-throughput platform for qPCR-based detection of STH at the population level. Importantly, validation practices for an assay designed for the diagnostic testing of an individual patient are fundamentally different from an assay designed strictly for the assessment of population-level infection prevalence and/or intensity for purposes of informing programmatic decision making. It is therefore critical that an assay be evaluated in the context of its expected utility. Accordingly, appropriate criteria for diagnostic platform validation were defined a priori, and the assay was validated in multiple laboratories. The results of this validation study demonstrate reliable, reproducible, and accurate detection of STH targets with high sensitivity and specificity.

The poor diagnostic performance of existing, commonly employed, coproscopic techniques limits the ability to define a true gold standard for detection of STH. Given that properly designed molecular assays are significantly more sensitive and specific than microscopy-based methods, both reference and index assays employed in this validation study would be expected to detect more true infections than would microscopy-based approaches. As a result, validation requires testing against a contrived panel, created by the spiking of an appropriate, naive matrix material with various combinations/concentrations of target material (eggs, DNA, etc.). Through the creation and testing of 18 such samples, each containing combinations of none/some/all the targets of interest at various concentrations, we were able to validate the performance of our index assays relative to the corresponding reference assays, and to assess transferability of technology through assay use at multiple laboratories. In addition, owing to their well-characterized nature, these samples were also used to create a bank of material appropriate for troubleshooting, technician qualification, and periodic proficiency testing. Coupled with additional controls including a *B. atrophaeus* IPC, an *A. suum*-containing external extraction control, and appropriate PCR controls, we were able to overcome the challenges associated with a lack of properly characterized control material in the absence of a true gold standard.

The approach to assay development, validation, and verification described here has several strengths. Our index platform is intended for use with samples collected as part of a rigorous, large-scale clinical trial (the DeWorm3 cluster randomized trial) conducted across multiple geographic and epidemiologic settings and involving hundreds of thousands of individuals. Given this intended use case, the a priori establishment of a validation protocol and the inclusion of a group of external evaluators increases the rigor of the assessment, which is critical for inspiring confidence in study results. In addition, the demonstration of reproducibility across laboratories suggests that this assay can be successfully transferred while maintaining diagnostic performance. This is critical in results interpretation for multi-site studies at scale and for future potential programmatic use. Finally, the creation of standard operating procedures, training documents, data collection instruments, and QA/QC procedures and panels can be used to support the establishment of large-scale laboratory platforms capable of supporting STH programs globally. These procedures and materials complement currently available resources, such as the STH community’s external quality assessment scheme, which is a valuable tool for validating and assessing the periodic performance of laboratories but does not support the frequency of quality assurance testing required by an effort such as the DeWorm3 trial [17].

While the development and validation pipelines that we employed allowed us to create a testing platform that is well-suited to its intended task, some limitations remain. Most critically, assessment of sample heterogeneity revealed some discrepancies in detection of targets across aliquots created from individual samples. These discrepancies were likely the result of the heterogenous distribution of target material within each sample, despite efforts to optimize homogenization procedures prior to DNA extraction. The inability to generate completely homogeneous samples means that the potential for some number of false negative results remains. Additional work will determine the frequency of false negativity, the likelihood of obtaining a false-negative result as a function of target mass/copy number in a sample, and the ability to buffer against false-negative results through procedural modifications (replicate testing, etc.). An additional limitation involved an element of inefficiency in DNA recovery because of imperfect methods for DNA extraction from fecal samples.

In addition to its intended purpose as a validation procedure for a high-throughput molecular testing platform for large-scale population-level studies of STH prevalence, the methodologies described here have utility beyond their primary use case. Through the adaptation of the described procedures, we have provided a blueprint for platform development capable of aiding programmatic efforts to evaluate other pathogens of interest. As global programs aimed at control/elimination of numerous neglected tropical diseases (NTD)s continue to realize varying degrees of success, there is increased need for sensitive and specific tools capable of accurately assessing infection prevalence at scale [18]. As NTD programs pivot towards elimination of disease as a public health problem and, in some cases, to elimination of transmission, such tools are needed to inform intervention stopping decisions and to monitor for disease recrudescence in the post-intervention setting. As such, guidelines facilitating the development of platforms capable of providing these answers, in the absence of reliable gold-standard methods, will be critical.

The success of current STH programs has led to substantial reductions in infection prevalence in target populations, primarily among children, and to a lesser extent, among women of reproductive age. Novel diagnostics are needed to accurately monitor the progress of STH programs given the limitations of existing tools. The platform described here is a well validated quantitative PCR-based approach that could be readily scaled for broad programmatic use in support of global morbidity management and STH transmission interruption efforts.

## Data Availability

All data produced in the present work are contained in the manuscript and supplement.

## Acknowledgements

The authors wish to thank all the study participants, communities and community leaders, national NTD program staff and other local, regional and national partners who have participated or supported the DeWorm3 study. We would like to acknowledge the work of all members of the DeWorm3 study teams and affiliated institutions, in particular the laboratory staff at Christian Medical College, Vellore, Versiti Inc, and the Institut de recherche Clinique du Benin (IRCB). We also wish to acknowledge the contributions of the DeWorm3 Data Safety Monitoring Board, including Richard Hayes, Harriet Mpairwe, Purushothaman Jambulingam and Jimmy Whitworth.

This work was supported by a Bill and Melinda Gates Foundation investment to the University of Washington (OPP1129535, PI JLW). A prior award to the Natural History Museum, London, also from the Bill and Melinda Gates Foundation (INV-030049), funded a portion of the work described. There is no involvement of the funders in final decisions regarding the study design and trial procedures or decision to publish the manuscript or in data collection, analysis or publication of study results. The funders reviewed the final decisions regarding study design and trial procedures.

