## Supplementary figures and images for "Development and validation of a high-throughput qPCR platform for the detection of soil-transmitted helminth infections"

### S6 Table

**S6 Table. Assay accuracy at the technical replicate and individual extraction level.**

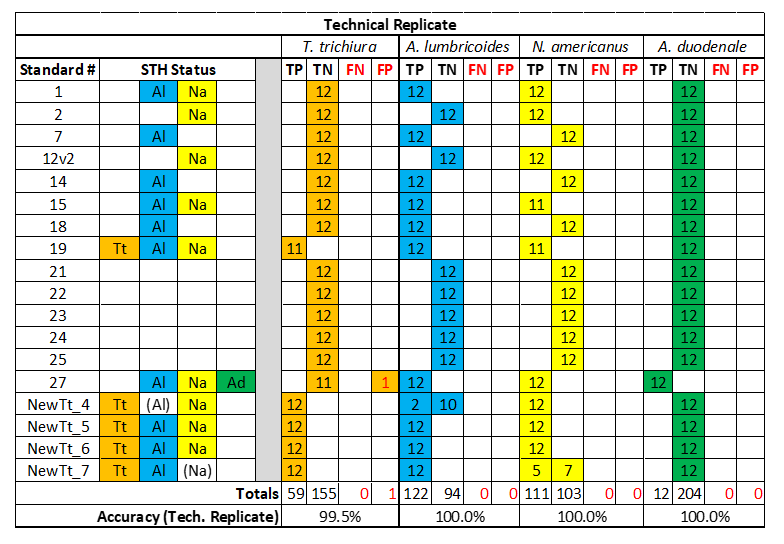

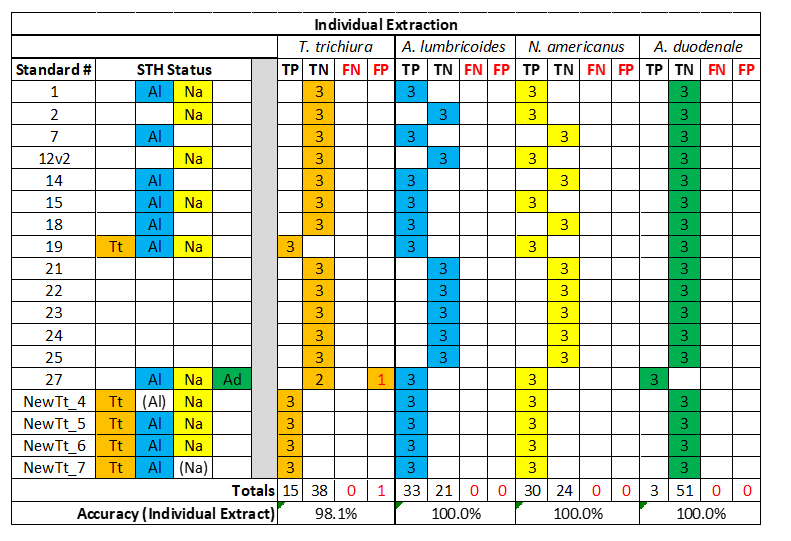
