## Supplementary material for "Development and validation of a high-throughput qPCR platform for the detection of soil-transmitted helminth infections": S1 Appendix

### DeWorm3 Assay Validation Report

Appendices ........ i

A1: Raw Data: *Ascaris suum*-spiked stool Validation (II.A.iii) ........ i

A2: Raw Data: *B. atrophaeus* Validation (II.A.iv) ........ ii

A3: Raw Data: WGA STH nucleic acid (II.A.v) ........ x

A4: Raw Data: Assay Accuracy (II.B.ii) ........ xi

A5: Raw Data: Assay Sensitivity/LOD (II.B.v) ........ xvi

A6: Raw Data: Multi-site Characterization of Spiked Stool Standards (II.B.iii) ........ xxv

A7: Evaluation of Assay Specificity (II.B.iv) ........ xxxv

A8: 200 Sample Experiment Data ........ xxxix

Figure A8.1: *The 200 sample experiment found 48 positive samples across 4 extractions. Within these samples, there was discordance, both between sites and within sites.* ........ xxxix

Figure A8.2: *All line data for the 200 sample experiment extractions* ........ xliii

Figure A8.3: *DNA line data for N. americanus at Quantigen and Smith* ........ lix

Figure A8.4: *DNA line data for T. trichiura at Quantigen and Smith* ........ lxvi

Figure A8.4: *DNA line data for A. lumbricoides at Quantigen and Smith* ........ lxxiii

A9: Control Proposal ........ lxxx
Purpose ........ lxxx
Internal Process Control: *Bacillus atrophaeus* ........ lxxx
*Data in support of B. atrophaeus Proposal* ........ lxxxi
*Supporting Material for the B. atrophaeus Proposal* ........ lxxxiii
Extraction Control: *A. suum* ........ lxxxiv
*Current A. suum Control* ........ lxxxiv
*A. suum Proposal* ........ lxxxiv
*Data in support of A. suum Proposal* ........ lxxxv
*Supporting Material for the A. suum Proposal* ........ lxxxvii
Negative Controls: No Sample Control and No Template Control ........ lxxxviii
*No Template Control (NTC)* ........ lxxxviii
*NSC/NTC Proposal* ........ lxxxviii
*Data to support NSC/NTC Proposal* ........ lxxxviii
*Evidence to support NSC/NTC Proposal* ........ xci
Positive PCR ........ xci
*Current Positive PCR Standard* ........ xci
*Positive PCR Proposal* ........ xci
*Transition to Plasmids* ........ xci
Continue with WGA, with altered passing qualifications. ........ xcii
*Data to Support Positive PCR Proposal* ........ xcii
*Evidence to Support Positive PCR Proposal* ........ xciv

### Introduction

The *DeWorm3 Assay Validation Plan* outlined a novel approach to assay validation in the absence of both standardized Soil Transmitted Helminth (STH) control material and an accurate "gold-standard" for STH diagnostics (I.H). This plan was executed by DeWorm3 collaborators at Smith College, Quantigen LLC, and Christian Medical College Vellore (CMC) from September 2020 through August 2021. The results demonstrate high performance of the DeWorm3 Assay and point to the potential for improved STH diagnostics.

Results presented below are organized by sections of the *DeWorm3 Assay Validation Plan*: Run Control Validation (II.A) and Assay Validation (II.B). Each verification checkpoint outlined in the *DeWorm3 Assay Validation Plan* is denoted with "***Verification***" throughout the report.

### Background

STH experts at Smith College developed and characterized (1) run controls (I.K) and (2) a panel of spiked-stool standards for use in evaluating the DeWorm3 qPCR Assay (I.I). Through a generative process of standards development and assay validation, DeWorm3 teams at Quantigen and CMC further characterized these spiked stool standards and run controls. A final panel of 18 well-characterized spiked stool standards representing various species and intensities of STH infection were developed and validated (**Figure 4**). The final panel of standards, in addition to field-collected samples from DeWorm3 collection sites, were utilized to measure target performance specifications of the DeWorm3 Assay (II.B.ii – II.B.vi). Finally, a large-scale additional investigation was included to resolve any Assay performance concerns.

### II.A. Run Controls Validation

#### II.A.iii. Validation of Run Control: *Ascaris suum*-spiked stool

##### Control Preparation (II.A.i)

*A. suum*-spiked stool will serve as the positive extraction control in the DeWorm3 extraction procedure. A fixed number of *A. suum* eggs were spiked into 5g of commercially purchased, helminth-naïve stool to achieve an eggs-per-gram (epg) concentration indicative of a moderate-level infection (5,000 to 49,999 epg). 280 aliquots (70 mg) of *A. suum*-spiked stool were prepared. Smith College extracted DNA from 10 aliquots and tested by qPCR in duplicate for quality control.

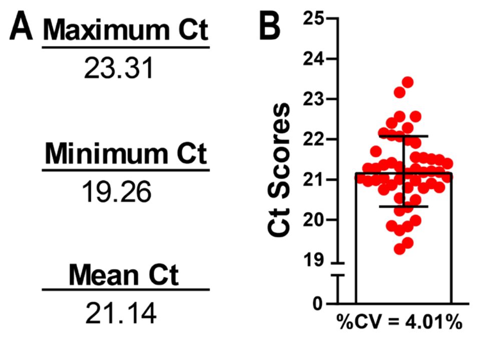

**Figure 1:** *A. suum* run control validation. A) The minimum, maximum, and average Ct of the validation. B) Dot plot depicting the range of the Ct's from the assay. The error bar in B is the standard deviation.

##### Methods Summary

Quantigen extracted DNA from 25 *A. suum*-spiked aliquots and tested them in duplicate by qPCR using DeWorm3 Lyophilized qPCR Plates.

##### Results

*A. suum* Ct scores (n=50) generated a composite CV of 4%, meeting the minimum target specification outlined in the *DeWorm3 Assay Validation Plan (**Verification II.A.iii.15**)*, with a mean of 21.14 (± 0.86) (**Figure 1**). The raw data are presented in the appendix (**Table A1. 1**).

##### Defining Ct Score Cut-off

Passing qualifications for this control are defined as positive *A. suum* amplification with a Ct score of ≤ 23.

#### II.A.iv. Validation of Run Control: *Bacillus atrophaeus* bacteria

##### Control Preparation

*Bacillus atrophaeus* will serve as the internal extraction control spiked into each DeWorm3 sample at a fixed concentration. *B. atrophaeus* stock were commercially purchased and diluted prior to use.

##### Methods Summary

*B. atrophaeus* spores were spiked into 200 STH-negative stool samples each from Benin and India[1]. *B. atrophaeus* was validated on both Assay 1 (*B. atrophaeus*, *N. americanus*, *T. trichiura*) and Assay 2 (*B. atrophaeus, A. lumbricoides, A. duodenale*).

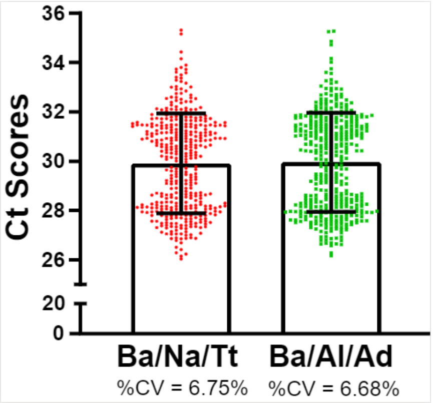

**Figure 2:** *B. atrophaeus* validation. A dot plot displaying all the Ct scores for Benin and India samples for both assays.

##### Results

Across samples from both Benin and India (n=413)[2] *B. atrophaeus* Ct scores generated a composite CV of < 7% (**Figure 2**), meeting the minimum target specification outlined in the *DeWorm3 Assay Validation Plan (**Verification II.A.iv.20**)*. The mean *B. atrophaeus* Ct score for Assay 1 and Assay 2 were 29.96 (±2.02) and 30.00 (±2.00), respectively. The raw data are presented in the **Table A2. 1**.

##### Defining Ct Score Cut-off

The acceptable *B. atrophaeus* Ct score range for the DeWorm3 study will be informed by results from DeWorm3 baseline samples. In the interim, passing qualifications for this control are defined as positive *B. atrophaeus* amplification with a Ct score ≤ 34 in both Assay 1 & Assay 2 in the absence of STH-amplification.

[1] STH-negative status was determined by the DeWorm3 Assay as part of evaluation of Assay Specificity (section II.B.iv). DeWorm3 samples collected in the Malawi site are not included in this report due to delays in sample shipment. Once received, *B. atrophaeus*, will be validated in samples from this site.

[2] An additional 13 STH negative stool samples were utilized from Benin for a total of 213 Benin samples.

#### II.A.v. Validation of Run Control: Whole genome-amplified (WGA) STH nucleic acid

##### Control Preparation (II.A.ii)

WGA STH nucleic acid will serve as a positive PCR control for the DeWorm3 Assay. Purified genomic DNA for each of the four helminth species was subjected to whole-genome amplification (WGA), diluted, and combined into a single pool targeting ±2 Ct's for each species.

**Table 1:** WGA validation.

| Species | Min Ct | Max Ct | Mean Ct | %CV |
|---|---|---|---|---|
| *N. americanus* | 27.74 | 31.27 | 29.67 | 3.32% |
| *A. lumbricoides* | 28.97 | 37.84 | 32.03 | 5.90% |
| *A. duodenale* | 28.26 | 37.47 | 29.41 | 6.21% |
| *T. trichiura* | 26.94 | 29.20 | 27.85 | 1.80% |

##### Methods Summary

Twenty-five aliquots of the final WGA STH nucleic acid pool were tested by the by qPCR using DeWorm3 Lyophilized qPCR Plates.

##### Results

Ct scores for each STH species in all aliquots resulted in < 10% CV meeting the minimum target specification outlined in the *DeWorm3 Assay Validation Plan (**Verification II.A.v.23**)*. The raw data are presented in the appendix (**Table A3. 1**).

##### Defining Ct Score Cut-off

Passing qualifications for WGA STH nucleic acid will be defined as < 2 standard deviations from the validation-defined mean Ct for each Assay. Thus, the Ct score cut-off for each species are as follows: *N. americanus* ≤ 31.6, *A. lumbricoides* ≤ 35.8, *A. duodenale* ≤ 33.1, and *T. trichiura* ≤ 28.9.

#### II.A. Run Control Validation Target Performance Specifications Go/No-Go Chart

**II.A. Run Controls Validation**

| | | Variance | | Results | |
|---|---|---|---|---|---|
| **Section** | **Validation** | **Minimum** | **Optimistic** | **Observed** | **Pass/Fail** |
| II.A.iii. | *Ascaris suum*-spiked stool | ≤ 10% CV | ≤ 2% CV | 4.01% CV | Pass |
| II.A.iv. | *Bacillus atrophaeus* | ≤ 10% CV | ≤ 2% CV | <7% CV | Pass |
| II.A.v. | WGA STH nucleic acid | ≤ 10% CV | ≤ 2% CV | 2-6% CV | Pass |

**Table 2**: Go/No-go table for validation II.A. Section II.A.iv is awaiting *B. atrophaeus* data from Malawi samples, however, all run controls meet the minimum target.

#### II.B. Assay Validation

Smith College prepared a panel of twenty-five candidate standards containing twenty STH-positive and five STH-negative standards by spiking helminth naïve stool with field samples of known STH infection status (**Figure 3**). Standards were initially characterized by triplicate DNA extraction and quadruplicate qPCR (3 aliquots x 4 replicates), according to Smith College's procedures. All standards maintained consistent amplification across all four qPCR technical replicates tested at Smith College. For assay validation, the STH-status of each standard was defined by the initial characterization by the Smith College Assay.

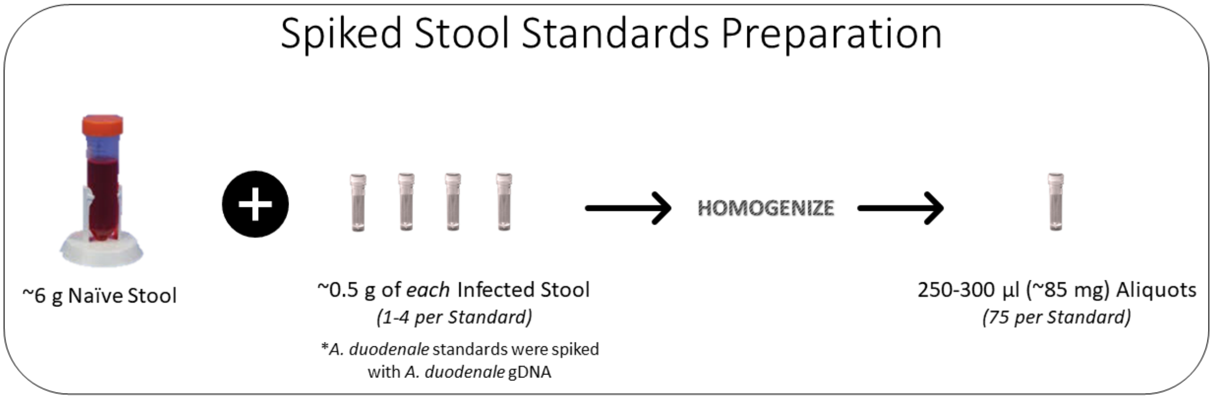

**Figure 3***: Smith College procedure for preparation of spiked-stool standards*

##### II.B.iii Multi-Site Characterization of Spiked-Stool Standards

**Methods Summary**

*Characterization of Initial Panel*

Three blinded aliquots of each standard (~85 mg per aliquot) were extracted and qPCR tested in quadruplicate at Quantigen by the DeWorm3 Assay (25 standards x 3 aliquots x 4 technical replicates). In the following summary, technical replicates refer to the 300 qPCR replicates tested at Quantigen (i.e., twelve qPCR data points for each of the twenty-five standards).

Discordant results across (1) technical replicates tested by the DeWorm3 Assay and (2) between aliquots tested by the DeWorm3 Assay and the Smith College Assay were investigated to determine the source of discordance. Experiments interrogated (1) Differences in qPCR Assay (i.e., Smith College Assay vs. DeWorm3 Assay), (2) Differences in DNA extraction procedures (i.e., Smith College DNA Extraction vs. DeWorm3 DNA extraction), and (3) Aliquot-to-aliquot variability of the contrived spiked-stool standards. As outlined in the Validation Plan, multi-site characterization interrogated all standards to ensure a robust panel of standards for utilization in DeWorm3 Assay Validation.

#### Results

*Characterization of Initial Panel*

Quantigen observed dropout of technical replicates in 11 of the 25 standards during initial testing by the DeWorm3 Assay (**Table 9**)**.** The DeWorm3 Assay failed to detect *T. trichiura* in at least 1 of 12 technical replicates for 8 standards (#3, #4.2, #8, #9, #11, #13, #16, #20, **Appendix A6**, **Table A6. 5**) including 3 standards for which Quantigen failed to detect *T. trichiura* for at least 1 of 3 aliquots (Standards #4.2, #13, and #16). The DeWorm3 Assay failed to detect *N. americanus* in at least 1 of 12 technical replicates for 2 standards (#12 and #17) (**Appendix A6, Table A6. 7**), none of which failed detection at the aliquot level. Finally, the DeWorm3 Assay failed to detect *A. duodenale* in at least 1 of 12 technical replicates for 1 standard (#10.2) (**Appendix A6, Table A6. 8**), which did not fail detection at the aliquot level.

**Table 9:** Failed Detection of STH in Initial Panel Standards by the DeWorm3 Assay. 3 aliquots x 4 technical replicates (n=12) were tested per standard.

| Standard # | STH Target | Aliquot Failures | Technical Replicate Failures |
|---|---|---|---|
| 3 | Tt | | 1 |
| 4.2 | Tt | 1 | 7 |
| 8 | Tt | | 1 |
| 9 | Tt | | 2 |
| 10.2 | Ad | | 4 |
| 11 | Tt | | 2 |
| 12 | Na | | 2 |
| 13 | Tt | 2 | 9 |
| 16 | Tt | 1 | 4 |
| 17 | Na | | 5 |
| 20 | Tt | | 2 |

*(1) Comparison of qPCR Assay: DeWorm3 vs. Smith College*

The failed detection of *T. trichiura* by the DeWorm3 Assay was investigated first by comparison of qPCR Assays. Quantigen-extracted DNA from all three aliquots of Standards #4.2, #13, and #16 were shipped to Smith College for testing by the Smith College qPCR Assay. The Smith College Assay produced nearly identical results to the DeWorm3 Assay at the aliquot and technical replicate level, suggesting either (a) sub-optimal DNA extraction by the DeWorm3 procedure or (b) aliquot-to-aliquot variability in the spiked-stool standards created by Smith College (**Appendix A6, Table A6. 9**).

*(2) Comparison of DNA Extraction Procedures: DeWorm3 vs. Smith College*

Due to the robust nature of *T. trichiura* eggs, sub-optimal lysis by the DeWorm3 DNA extraction procedure was suspected and investigated. Increased bead-beating (lysis) times and different bead mixtures (i.e., Omni International beads utilized in the DeWorm3 extraction vs. MP Bio beads utilized in the Smith College extraction) were compared across a sub-set of standard aliquots, but no clear differences were observed (**Appendix A6, Table A6. 10**).

*(3) Aliquot-to-Aliquot Variability of Standards*

Finally, aliquot-to-aliquot variability was investigated through multiple extractions of Standard #11 with the Smith College extraction procedure (utilizing both Omni International beads and MP Bio beads) and the Smith College qPCR assay. Smith College observed dropout of *T. trichiura* detection across aliquots extracted with both bead types, suggesting that inconsistent detection of *T. trichiura* by the DeWorm3 Assay was likely due to variability across aliquots of the standards (**Appendix A4, Table A6. 11**).

It is important to note that *T. trichiura* material utilized in the creation of spiked-stool standards was limited, as exhibited by the high Ct Scores (Mean Ct of 30) observed across Smith and Quantigen during initial characterization (**Appendix A6, Table A6. 1**). Further, since the exact STH egg-count spiked into each standard is unknown, it is impossible to determine whether eggs were poorly dispersed across aliquots, or if the concentration of *T. trichiura* material is skirting the limit of detection for both the Smith College and DeWorm3 Assays.

Following the above outlined experiments, Smith College created new *T. trichiura* standards by spiking-in more than one *T. trichiura*-positive infected stool sample (3-4 total samples) into helminth naïve stool. These new standards (NewTt_4, NewTt_5, NewTt_6, and NewTt_7) performed consistently across aliquots and technical replicates tested by both the Smith College Assay and the DeWorm3 Assay (**Appendix A4, Table A4. 2**).

Three additional standards (#10.2, #12, and #17) produced inconsistent results and were removed from the panel. Standard #12 and #17 demonstrated a large deviation in Ct score across Smith aliquots, in addition to technical replicate drop out at Quantigen (**Appendix A6, Table A6. 3** & **Table A6. 7**). Standard #10.2 demonstrated technical replicate dropout for *A. duodenale* at Quantigen (**Appendix A6, Table A6. 8**).

*Characterization of Final Panel*

The final panel (**Figure 4**) performed consistently (at the technical replicate and aliquot level) at Smith College and Quantigen with all standards meeting the target of 100% Inter-laboratory concordance (***Verification II.B.iii.19***). Raw data is presented in **Appendix A4** (**Table A4. 2, Table A4. 3, Table A4. 4, Table A4. 5**).

| Species | Count |
|---|---|
| Tt | 5 |
| Al | 11 |
| Na | 10 |
| Ad | 1 |
| **Total** | **18** |

| Standard # | STH Status | | | |
|---|---|---|---|---|
| 1 | | Al | Na | |
| 2 | | | Na | |
| 7 | | Al | | |
| 12v2 | | | Na | |
| 14 | | Al | | |
| 15 | | Al | Na | |
| 18 | | Al | | |
| 19 | Tt | Al | Na | |
| 21 | | | | |
| 22 | | | | |
| 23 | | | | |
| 24 | | | | |
| 25 | | | | |
| 27 | | Al | Na | Ad |
| NewTt_4 | Tt | (Al) | Na | |
| NewTt_5 | Tt | Al | Na | |
| NewTt_6 | Tt | Al | Na | |
| NewTt_7 | Tt | Al | (Na) | |

**Figure 4:** DeWorm3 Spiked Stool Standards. STH-status represented in parentheses denotes STH which were shown to be detected inconsistently at both Smith and Quantigen.

Of note, standards NewTt_4 and NewTt_7 are suspected to include low quantities of *A. lumbricoides* and *N. americanus*, respectively, due to sporadic detection of these species across technical replicates at both Smith and Quantigen (**Figure 5**). These "weak positives" are likely due to a low-intensity *A. lumbricoides* or *N. americanus* infection in one of the 3-4 infected stool samples utilized in the preparation of these standards (**Figure 3**). Thus, for the purposes of validation and use of the Standards Panel for proficiency testing either positive or negative amplification may be acceptable for these species in these standards; this is denoted by parentheses in **Figure 4**.

| | | Positive Amplification (Tech. Replicates) | | | |
|---|---|---|---|---|---|
| | | Tt | (Al) | Na | Ad |
| **NewTt_4** | **Smith** | 12 | 2 | 12 | 0 |
| | **Quantigen** | 12 | 2 | 12 | 0 |

| | | Positive Amplification (Tech. Replicates) | | | |
|---|---|---|---|---|---|
| | | Tt | Al | (Na) | Ad |
| **NewTt_7** | **Smith** | 12 | 12 | 1 | 0 |
| | **Quantigen** | 12 | 12 | 5 | 0 |

**Figure 5**: Technical Replicate Positive STH status Smith vs. Quantigen for NewTt_4 and NewTt_7.

#### Conclusion

Ultimately, a final panel of 18 consistent Spiked-Stool Standards (13 STH-positive and 5 STH-negative, **Figure 4**) was utilized for DeWorm3 Assay Validation. This final panel will be validated at CMC for site qualification.[3] The final data from all three sites will be compared to ensure 100% interlaboratory concordance, but the current results point to strong interlaboratory concordance.

#### II.B.ii Evaluation of Assay Accuracy

##### Methods Summary

Assay accuracy was evaluated on the technical replicate level and the aliquot level using the final panel of 18 standards. Three blinded aliquots (~85 mg) of each of the 18 standards were extracted and qPCR tested in quadruplicate by the DeWorm3 Assay (18 standards x 3 aliquots x 4 technical replicates). Accuracy was calculated using **Formula 1**. The true positive or negative status of each standard was defined by Smith College's initial characterization (**Figure 4**). Evaluation of STH positivity at the aliquot level was defined as at least one positive amplification out of all technical replicates tested by the DeWorm3 Assay. Well failures were not included in calculations of assay accuracy.

$$(1)\quad Accuracy = \frac{True\ Positives + True\ Negatives}{True\ Positives + True\ Negatives + Fales\ Positives + False\ Negatives}$$

##### Results

**Table 3:** DeWorm3 Assay Accuracy: Individual Extraction and Technical replicate level.

| | Assay Accuracy | | | |
|---|---|---|---|---|
| **Level** | *T. trichiura* | *A. lumbricoides* | *N. americanus* | *A. duodenale* |
| Technical Replicate | 99.5% | 100.0% | 100.0% | 100.0% |
| Individual Extraction | 98.1% | 100.0% | 100.0% | 100.0% |

Assay Accuracy on the technical replicate level met the minimum target with > 99% accuracy for all assays (**Table 3**). *A. lumbricoides, N. americanus,* and *A. duodenale* assays met the minimum 100% accuracy at the individual extraction level. One *T. trichiura* false positive present in standard #27, however, decreased the individual extraction level accuracy for *T. trichiura* to 98.1% (**Table 3**). See Appendix A4 for raw data.

The above-mentioned false positive was due to erroneous *T. trichiura* amplification in 1 out of 12 technical replicates of standard #27 (36.8 Ct, **Table A4. 2**); this is the sole exception to meeting ***Verification II.B.ii.14***. Upon quadruplicate repeat testing at Quantigen in DeWorm3 multiplex and singleplex assays (n=8), this *T. trichiura* amplification was not reproducible.

All the species present in the stool standards were amplified in all three aliquots (***Verification II.B.ii.10***). All the standard aliquots amplified helminth targets ≥ 3 out of the 4 qPCR wells and < 1% of all qPCR wells failed to amplify as expected (***Verification II.B.ii.*11**, **Table A4. 1**). None of the STH negative stool standards (n=5) amplified erroneous targets (***Verification II.B.ii.13****)*.

All replicate wells in which at least one of the expected STH targets did not amplify, *B. atrophaeus* (internal extraction control) also failed to amplify (***Verification II.B.ii.12***). No false negatives were observed, meeting ***Verification II.B.ii.15***.

[3] CMC has yet to complete testing due to delays in receipt of supplies. Results will be reported when testing is completed at this site.

##### II.B.iv. Evaluation of Assay Specificity

#### Methods Summary

Assay specificity was assessed using 200 confirmed STH-negative stool samples per DeWorm3 site (n=413)[4]. Quantigen identified 213 and 200 STH-negative samples from the Benin and India sites, respectively, using the DeWorm3 Assay. All 413 stool samples found STH-negative by the Deworm3 Assay were subsequently tested utilizing the Smith College assay. The panel of Benin negatives, the "Benin Specificity Panel" (n=213) were tested at Smith College[5] and the panel of India negatives, the "India Specificity Panel" (n=200) were tested at CMC; both Smith College and CMC utilized the Smith College extraction and qPCR procedure.

#### Results

*Benin Specificity Panel*

Smith College identified 3 samples from the Benin Specificity Panel to be positive for *N. americanus* (n=2) and *A. lumbricoides* (n=1) (**Appendix A7, Figure A7. 1**). As outlined in the validation plan, these inconsistencies were investigated. First, DNA extracted by Smith College and Quantigen from all three discordant samples (DNA from initial testing at each site) were tested by the liquid and lyophilized DeWorm3 Assay at Quantigen. The STH status of the Smith College extracts were confirmed by the DeWorm3 Assay in liquid and lyophilized form (**Appendix A7, Figure A7. 2**). However, one of the three Quantigen extracts were found positive for *A. lumbricoides* (n=1) by testing in both the liquid and lyophilized DeWorm3 Assay (**Appendix A7, Figure A7. 3**).

To investigate aliquot-to-aliquot variability, three new aliquots of each discordant sample (3 samples x 3 aliquots) were re-extracted by the DeWorm3 procedure and tested by the DeWorm3 liquid Assay in single qPCR replicates. All previously detected STH were detected in at least 2 out of 3 aliquots extracted. (**Appendix A7, Figure A7. 4**).

Discordance among different aliquots from the same sample, tested by the same procedure and assay, suggests the discordance between Smith and Quantigen results lies in the heterogenous distribution of STH eggs throughout stool sample aliquots.

*India Specificity Panel*

CMC identified 5 *N. americanus*-positive samples among those identified as negative in Quantigen's initial screening. Further investigation into these inconsistencies was conducted. Per the validation plan, these inconsistencies were interrogated. DNA extracted by CMC and Quantigen from the 5 discordant samples were tested in triplicate by the lyophilized DeWorm3 Assay at Quantigen (**Appendix A7, Figure A7. 5**, **Figure A7. 6**). All original STH status of the extracts were confirmed.

---

[4] DeWorm3 samples collected in the Malawi site are not included in this report due to delays in sample shipment. Once received, assay specificity will be validated by the same methods.

[5] The 13 additional Benin samples were shipped to Smith because some samples were low volume. However, all 213 samples had sufficient volume upon extraction at Smith, thus, all results are reported here.

To investigate aliquot-to-aliquot variability, the 5 discordant samples were re-extracted. Three of 5 samples were re-extracted in duplicate, 2 of 5 samples were re-extracted once.[6] These 8 extracts (3 samples x 2 extractions + 2 samples x 1 extraction) were tested via the lyophilized DeWorm3 Assay in duplicate. The samples that were re-extracted once detected STH in all qPCR replicates. One sample extracted in duplicate detected STH across both aliquots. Two samples that extracted in duplicate were discordant in STH detections between extractions (**Appendix A7,Figure A7. 7).**

Following the investigation into India Specificity discordance, the data suggests that the source of discordance between CMC and Quantigen is the uneven distribution of STH eggs throughout stool sample aliquots.

*Specificity Go/No-Go*

With Benin and India Specificity Panel results, the DeWorm3 Assay Specificity is 98%; this does not meet the minimum target of 99% (***Verification II.B.iv.25***). As demonstrated by the investigation of the discordant samples from the Benin and India Specificity Panels, however, it is likely this variability is due to the heterogeneity of stool samples.

#### II.B.v. Evaluation of Assay Sensitivity/LOD

##### Methods Summary

Spiked stool standards were selected for the determination of the limit of detection (LOD) due to the availability of Kato-Katz data from the field samples used in their creation[7] (**Table 4**).

Approximately 500mg of each field sample was spiked into naïve feces for the development of all standards. **Formula 2** was used to calculate the concentration of STH (epg) in each standard. Estimated STH concentrations (epg) for each standard are listed in **Table 5**.

**Table 4:** Concentration of STH eggs in field samples used to make the validation standards.

| Species | Standard | Field Sample Concentration |
|---|---|---|
| *N. americanus* | 1 | 960 epg |
| *A. lumbricoides* | 1 | 2100 epg |
| *A. duodenale* | 27 | 500 epg* |
| *T. trichiura* | 19 | 4224 epg |

*Standard 27 was spiked with 13 ng of *A. duodenale* WGA material which equates to 500 eggs.

$$(2)\quad \left(\frac{\text{eggs in field sample}}{1\ g}\right) \times \left(\frac{\text{Spiked Field Sample Mass}}{\text{Total Standard Mass}}\right) = STH\ eggs\ per\ gram\ of\ Standard$$

[6] The two samples that were extracted once were done so due to exhaustion.
[7] Note that no *A. duodenale* egg material was available for standard development, thus Standard #27 was spiked with 13ng of *A. duodenale* WGA DNA, which equates to approximately 500 eggs.

Each standard was extracted by the DeWorm3 procedure. Extracted DNA for each standard was serially diluted 1:10 for a total of 6 dilution points. Each dilution point was tested by the DeWorm3 Assay in ten technical replicates (4 standards x 6 dilution points x 10 replicates) and the lowest concentration in which all ten replicates generated a positive signal was identified. Starting at a concentration 2-fold higher than this concentration, five 1:2 serial dilutions were created. Twenty technical replicates of each dilution point were tested by qPCR (4 standards x 5 dilution points x 20 replicates). The LOD was determined as the lowest concentration at which at least 19/20 technical replicates amplified for the target STH species (**Table A5. 1**).

**Table 5**: Concentration of STH+ in stool standards.

| Species | Field Sample Mass | Naive Stool Mass | Conc. per gram** |
|---|---|---|---|
| *N. americanus* | ~500 mg | 6.052 g | 73.26 epg |
| *A. lumbricoides* | ~500 mg | 6.052 g | 160.26 epg |
| *A. duodenale* | 500 eggs* | 6.357 g | 78.65 epg |
| *T. trichiura* | ~500 mg | 6.045 g | 322.69 epg |

*13 ng/500 eggs were spiked into naive stool due to lack of egg material

**Final concentration of STH eggs or DNA per gram of stool

The assay LOD was calculated using **Formula 3.** However, several assumptions are made in these calculations. First, it is assumed that the STH eggs are evenly distributed throughout the feces. Second, it is assumed that 500 mg of spiked stool is added to naïve stool, as the initial spiked amount was not recorded. Third, between 50 and 75 mg of feces is extracted and used to test for STH. To measure the LOD, it was assumed that the lowest allowable mass was extracted (50 mg).

*(3)* $\frac{STH\ eggs}{1\ g\ of\ Standard} \times 50\text{ mg aliquot} \times \text{qPCR dilution limit} = \text{LOD}$

#### Results

Quantigen determined the lowest concentration at which at least 19/20 technical replicates amplified for the target STH species (See **Table A5. 2** & **Table A5. 3** for *N. americanus* data, **Table A5. 4** & **Table A5. 5** for *A. lumbricoides* data, **Table A5. 6** & **Table A5. 7** for *A. duodenale* data, and **Table A5. 8** & **Table A5. 9** for *T. trichiura* data). Using **Formula 3**, Quantigen determined that the LOD for all standards was <1 epg for each species (**Table 6**), meeting the minimum target (**Table 8**).

**Table 6**: Limit of detection for each STH.

| Species | Dilution Limit | LOD |
|---|---|---|
| *N. americanus* | $2.50 \times 10^{-5}$ | $9.16 \times 10^{-5}$ |
| *A. lumbricoides* | $1.25 \times 10^{-4}$ | $1.00 \times 10^{-3}$ |
| *A. duodenale* | $1.25 \times 10^{-2}$ | $4.92 \times 10^{-2}$ |
| *T. trichiura* | $1.25 \times 10^{-4}$ | $2.02 \times 10^{-3}$ |

#### II.B.vi. Evaluation of Assay Precision

##### Methods Summary

Three independent operators will extract DNA from all standards and test on qPCR using 2 separate QuantStudio qPCR machines in duplicate.

**Table 7:** Assay precision for technical and aliquot extractions.

| Species | Max Ct | |
|---|---|---|
| | TR* | AQ** |
| *N. americanus* | 1.18% | 0.62% |
| *A. lumbricoides* | 24.37% | 8.99% |
| *A. duodenale* | 1.90% | 0.95% |
| *T. trichiura* | 6.89% | 3.09% |

*Technical replicate
**Aliquot

##### Validation of protocol

The precision of the assay failed at the level of technical replicates but passed at the level of extraction (aliquots) (***Verification II.B.vi.35,* Table 7**). One technical replicate out of 4 from standard 18 had an *A. lumbricoides* Ct score that was uncharacteristically higher than that of the other technical replicates for this aliquot. We speculate that this could be a loading error. It is important to note that the "clinical call" for this replicate would still be accurate.

#### II.B Assay Validation Target Performance Specifications Go/No-Go Chart

**II.B. Assay Validation**

| Section | Validation | Level | Variance | | Results | |
|---|---|---|---|---|---|---|
| | | | Minimum | Optimistic | Observed | Pass/Fail |
| II.B.ii. | Accuracy | Technical Replicate | ≥ 95% | 100% | ≥ 99.5% | Pass |
| | | Individual Extraction | 100% | 100% | ≥ 98.1% | Fail |
| II.B.iii. | Multi-Site Characterization | - | 100% | 100% | TBD | - |
| II.B.iv. | Specificity | - | ≥ 99% | 100% | 98% | Fail |
| II.B.v. | Sensitivity/LOD | - | ≤ 1 epg | ≤ 1 epg | ≤ 1 epg | Pass |
| II.B.vi. | Precision | Technical Replicate | CV ≤ 10% | CV ≤ 5% | ≤ 24.4%* | Fail |
| | | Individual Extraction | CV ≤ 15% | CV ≤ 10% | ≤ 9% | Pass |

*Precision for all but one technical replicate of Standard #18 produced < 9.3% CV.

**Table 8**: Assay Validation Go/No-Go Table.

### Additional Investigation

##### 200 Sample Experiment

##### Methods Summary

The goal of the 200 Sample Experiment was to determine whether discordance seen during Assay Validation was due to sample heterogeneity or suboptimal assay performance. This experiment was done by taking a random 10% of the first 2,000 extracted samples and extracting four (4) aliquots of each sample. Each extraction was then run on both the DeWorm3 Assay and the comparator Smith College Assay. This generated a total of 2 data points per extraction, and a total of 8 data points for each sample.  To test for site extraction differences, two extractions per sample were completed via the DeWorm3 protocol and two extractions per sample were completed via the Smith College protocol. A diagram illustrating the workflow for one sample is below in Figure 6.

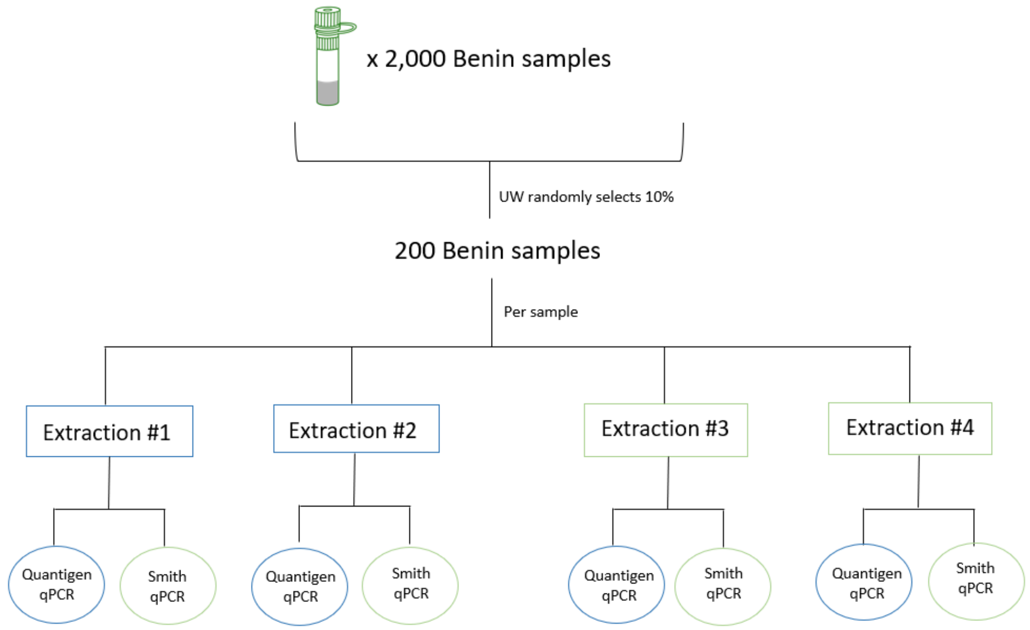

*Figure 6: Sample workflow for the 200-sample experiment. This diagram shows how one sample generates eight points of data*

#### Results

Between all four extractions, discordance was seen for all but one target, *A. duodenale*, which was negative for all extractions. 8/200 *N. americanus*, 3/200 *T. trichiura*, and 10/200 *A. lumbricoides* were found to be discordant.

When looking at the data on a site-by-site extraction basis, Quantigen saw discordance for *N. americanus* (6/200) and *A. lumbricoides* (8/200), while Smith College saw discordance for *N. americanus* (2/200), *T. trichiura* (1/200), and *A. lumbricoides* (3/200). Further looking at site-by-site performance, Quantigen and Smith found similar numbers of overall positivity (200 samples x 2 extractions = 400).

Extraction 1, 3, and 4 found 36 positives on a population level, while extraction 2 found 43. The remaining difference between extraction 2 and the other extractions can be reduced to unavoidable stochastic variability. These data are illustrated in Figure 7 and show that despite the procedural differences, the DeWorm3 Assay and Smith College Assay have equivalent performance.

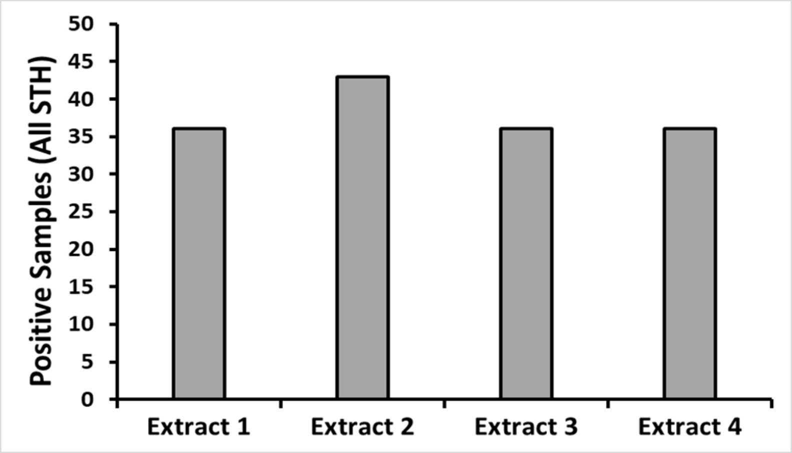

*Figure 7: Positive Samples on a Population per extraction. Each extraction had an (n) of 200. Extract 1, Extract 3, and Extract 4 found (n)=36 positives and Extract 2 found (n)=43 positives*

Further comparisons were made between the Smith College qPCR Assay and the Quantigen DeWorm3 Assay. This was done by comparing the results of one extract on the two assays. The two assays produced high amount of concordance between the labs, indicating that both assays are equivalent. Overall concordance for each species is as follows: *N. americanus* 98.5% concordance, *T. trichiura* 100% concordance, *A. duodenale* 100% concordance, and *A. lumbricoides* 98.1% concordance.

All line data for this experiment can be found in **Figure A8.2**

#### Conclusion

Looking at all these data, there is no clear assay performance differences between the DeWorm3 Assay and the Smith College Assay, leading to the conclusion that the Quantigen DeWorm3 Assay is performing at the necessary level for final validation.

This experiment also shows that the previous specificity inaccuracies were due to stool sample heterogeneity that cannot be avoided, regardless of the procedure used. The data confirms that the sample heterogeneity that is observed is not a function of the DeWorm3 assay itself. Additionally, the observed specificity from the Benin and India Specificity Panels shows that these measured specificity values are to be expected due to the nature of the samples.

This experiment will also be functionally repeated during the QC/QA testing built into the study. 10% of samples throughout the study, including those with lower intensity infections, will be retested between sites, allowing for further quantification of assay performance in samples with wide ranges of intensities.

#### Conclusion

Although there are some items of the Validation Plan that are considered to miss the target set out by the initial plan, upon completion of the 200-sample experiment, collaborators at Quantigen, Smith College, and the University of Washington are confident that the DeWorm3 Assay meets the criteria for final validation. All of the observed results indicate that inaccuracies seen are due to sample heterogeneity and are not a function of the DeWorm3 Assay.

#### Next Steps

Qualification of international labs are still pending completion due to shipping delays and COVID-19 related issues. This includes testing of standards at CMC for multi-site characterization (II.B.iii) and evaluation of specificity using samples from the DeWorm3 Malawi site (II.B.iv). Results from these portions of validation will be reported to all principles upon completion, however, these data are not necessary for verifying that the Quantigen DeWorm3 Assay is validated.

### Appendices

#### A1: Raw Data: *Ascaris suum*-spiked stool Validation (II.A.iii)

**Table A1. 1:** *A. suum Ct Scores for 25 aliquots of the DeWorm3 Ascaris suum-spiked stool (positive extraction control) extracted and qPCR tested by the DeWorm3 Assay for Validation II.A.iii.*

| Sample Name | CT - *Ascaris suum* |
|---|---|
| A1 | 19.26 |
| A1 | 19.41 |
| A2 | 19.71 |
| A2 | 19.82 |
| A3 | 19.81 |
| A3 | 19.95 |
| A4 | 20.75 |
| A4 | 20.92 |
| A5 | 21.20 |
| A5 | 21.30 |
| A6 | 20.90 |
| A6 | 21.13 |
| A7 | 20.27 |
| A7 | 20.19 |
| A8 | 21.22 |
| A8 | 21.21 |
| A9 | 21.47 |
| A9 | 21.63 |
| A10 | 20.70 |
| A10 | 20.75 |
| A11 | 20.85 |
| A11 | 20.74 |
| A12 | 21.14 |
| A12 | 20.99 |
| A13 | 20.49 |
| A13 | 20.45 |
| A14 | 21.35 |
| A14 | 21.49 |
| A15 | 21.42 |
| A15 | 21.33 |
| A16 | 20.95 |
| A16 | 20.98 |
| A17 | 22.20 |
| A17 | 22.32 |
| A18 | 21.09 |
| A18 | 21.12 |
| A19 | 21.44 |
| A19 | 21.25 |
| A20 | 21.99 |
| A20 | 21.00 |
| A21 | 22.47 |
| A21 | 21.90 |
| A22 | 23.06 |
| A22 | 23.31 |
| A23 | 22.02 |
| A23 | 21.83 |
| A24 | 22.07 |
| A24 | 22.48 |
| A25 | 20.93 |
| A25 | 20.81 |

#### A2: Raw Data: *B. atrophaeus* Validation (II.A.iv)

**Table A2. 1**: *B. atrophaeus Ct Scores from 413 STH-Negative Benin and India DeWorm3 Samples. Each sample has two Ct Scores for B. atrophaeus corresponding to each of the two wells/assays constituting the DeWorm3 Assay**. This table extends from Pg. ii - ix, each row denotes a different sample**.*

| Bacillus Ct Assay 1 (Na/Tt/Ba) | Bacillus Ct Assay 2(Ad/Al/Ba) |
|---|---|
| 28.59 | 28.46 |
| 27.52 | 27.61 |
| 30.13 | 29.14 |
| 26.91 | 27.15 |
| 30.21 | 29.84 |
| 28.60 | 28.38 |
| 27.99 | 27.81 |
| 26.74 | 26.84 |
| 27.85 | 28.01 |
| 27.92 | 28.13 |
| 28.68 | 28.56 |
| 27.87 | 27.63 |
| 28.16 | 28.55 |
| 28.22 | 27.67 |
| 28.40 | 28.17 |
| 28.69 | 28.67 |
| 28.62 | 28.39 |
| 28.13 | 27.96 |
| 28.14 | 27.80 |
| 28.02 | 27.97 |
| 27.80 | 28.04 |
| 27.61 | 27.60 |
| 28.29 | 28.28 |
| 28.00 | 27.30 |
| 27.92 | 27.66 |
| 28.56 | 28.31 |
| 28.16 | 28.18 |
| 27.20 | 28.07 |
| 28.38 | 28.31 |
| 27.25 | 27.28 |
| 27.72 | 27.68 |
| 28.28 | 28.40 |
| 28.55 | 28.27 |
| 27.93 | 28.34 |
| 28.18 | 28.38 |
| 27.74 | 27.81 |
| 28.25 | 27.78 |
| 28.26 | 28.26 |
| 27.99 | 27.47 |
| 28.17 | 28.80 |
| 27.36 | 27.48 |
| 27.83 | 27.68 |
| 28.55 | 28.41 |
| 28.77 | 28.73 |
| 27.36 | 27.50 |
| 28.70 | 28.45 |
| 30.30 | 30.67 |
| 28.23 | 28.67 |
| 27.57 | 28.20 |

| Bacillus Ct Assay 1 (Na/Tt/Ba) | Bacillus Ct Assay 2(Ad/Al/Ba) |
|---|---|
| 28.06 | 27.42 |
| 27.59 | 27.93 |
| 28.46 | 27.96 |
| 28.10 | 27.95 |
| 28.64 | 28.70 |
| 27.77 | 27.48 |
| 33.77 | 30.68 |
| 27.85 | 27.78 |
| 29.72 | 29.66 |
| 30.39 | 27.98 |
| 27.83 | 27.76 |
| 28.11 | 28.43 |
| 26.95 | 27.20 |
| 27.50 | 27.67 |
| 27.85 | 28.50 |
| 32.15 | 26.69 |
| 27.86 | 28.04 |
| 30.41 | 31.21 |
| 29.18 | 29.88 |
| 31.18 | 31.19 |
| 31.29 | 30.73 |
| 31.23 | 31.19 |
| 31.74 | 32.54 |
| 30.86 | 31.14 |
| 30.46 | 30.49 |
| 33.85 | 33.80 |
| 30.52 | 30.44 |
| 30.93 | 30.50 |
| 32.62 | 36.24 |
| 27.97 | 28.67 |
| 33.94 | 33.31 |
| 30.25 | 31.12 |
| 31.16 | 31.01 |
| 29.39 | 31.50 |
| 30.31 | 30.59 |
| 30.94 | 30.66 |
| 29.83 | 29.90 |
| 30.25 | 30.52 |
| 29.55 | 30.47 |
| 31.39 | 31.38 |
| 32.33 | 32.11 |
| 31.50 | 31.94 |
| 32.98 | 33.25 |
| 30.64 | 30.47 |
| 31.49 | 31.11 |
| 30.49 | 30.13 |
| 31.62 | 31.57 |
| 31.05 | 30.61 |
| 32.56 | 32.38 |
| 32.44 | 30.39 |

| Bacillus Ct Assay 1 (Na/Tt/Ba) | Bacillus Ct Assay 2(Ad/Al/Ba) |
|---|---|
| 29.27 | 29.78 |
| 30.89 | 31.12 |
| 29.62 | 29.49 |
| 31.43 | 31.95 |
| 31.31 | 38.69 |
| 31.12 | 31.88 |
| 29.42 | 30.91 |
| 30.23 | 30.38 |
| 30.56 | 31.25 |
| 31.12 | 31.90 |
| 30.01 | 30.29 |
| 33.63 | 32.72 |
| 31.23 | 31.47 |
| 29.41 | 30.74 |
| 33.04 | 32.11 |
| 29.31 | 32.57 |
| 29.63 | 29.73 |
| 29.23 | 28.88 |
| 28.67 | 28.92 |
| 29.48 | 29.23 |
| 28.74 | 29.53 |
| 29.00 | 29.16 |
| 29.49 | 29.27 |
| 29.77 | 29.56 |
| 30.67 | 30.15 |
| 29.16 | 29.33 |
| 30.89 | 30.76 |
| 30.82 | 31.25 |
| 30.44 | 30.16 |
| 30.70 | 30.11 |
| 28.95 | 29.13 |
| 29.74 | 29.53 |
| 29.48 | 29.96 |
| 28.61 | 29.29 |
| 27.71 | 27.86 |
| 30.19 | 30.56 |
| 30.68 | 29.80 |
| 28.55 | 28.73 |
| 29.13 | 29.11 |
| 28.76 | 28.40 |
| 29.15 | 28.92 |
| 29.61 | 29.18 |
| 28.94 | 28.70 |
| 29.27 | 29.23 |
| 28.73 | 28.86 |
| 28.68 | 28.49 |
| 29.51 | 30.28 |
| 28.89 | 29.42 |
| 29.53 | 29.41 |
| 29.65 | 29.51 |

| Bacillus Ct Assay 1 (Na/Tt/Ba) | Bacillus Ct Assay 2(Ad/Al/Ba) |
|---|---|
| 30.06 | 30.04 |
| 29.70 | 29.40 |
| 31.77 | 31.79 |
| 30.17 | 29.44 |
| 27.53 | 27.60 |
| 31.45 | 30.69 |
| 29.26 | 29.13 |
| 29.28 | 29.96 |
| 29.38 | 30.04 |
| 31.01 | 31.29 |
| 31.17 | 30.43 |
| 31.48 | 31.03 |
| 39.97 | 32.04 |
| 30.93 | 31.34 |
| 30.63 | 30.75 |
| 36.36 | 31.19 |
| 30.50 | 30.28 |
| 32.46 | 30.51 |
| 30.71 | 31.07 |
| 32.20 | 33.09 |
| 31.21 | 31.49 |
| 31.20 | 31.09 |
| 31.38 | 31.54 |
| 31.53 | 31.28 |
| 34.47 | 31.51 |
| 30.19 | 30.47 |
| 30.12 | 31.08 |
| 30.70 | 30.84 |
| 30.97 | 32.21 |
| 30.98 | 30.69 |
| 31.92 | 32.17 |
| 31.71 | 31.85 |
| 35.36 | 29.32 |
| 30.38 | 30.38 |
| 30.88 | 30.99 |
| 31.07 | 30.93 |
| 33.66 | 31.69 |
| 32.15 | 32.23 |
| 30.62 | 30.93 |
| 27.54 | 27.65 |
| 27.62 | 27.70 |
| 28.00 | 27.83 |
| 27.13 | 27.15 |
| 27.10 | 26.91 |
| 28.23 | 28.53 |
| 30.26 | 31.09 |
| 30.04 | 30.44 |
| 33.21 | 32.27 |
| 30.34 | 30.24 |
| 31.55 | 30.99 |

| Bacillus Ct Assay 1 (Na/Tt/Ba) | Bacillus Ct Assay 2(Ad/Al/Ba) |
|---|---|
| 29.25 | 28.74 |
| 29.35 | 29.36 |
| 28.38 | 28.50 |
| 27.68 | 27.33 |
| 27.71 | 27.51 |
| 28.77 | 28.56 |
| 27.32 | 27.51 |
| 27.60 | 27.64 |
| 28.35 | 28.64 |
| 28.95 | 28.66 |
| 28.29 | 28.31 |
| 27.97 | 27.93 |
| 29.80 | 29.70 |
| 28.12 | 28.26 |
| 27.40 | 27.61 |
| 29.13 | 29.50 |
| 28.17 | 28.19 |
| 28.19 | 28.21 |
| 28.16 | 28.14 |
| 28.75 | 28.85 |
| 28.23 | 27.97 |
| 28.35 | 28.57 |
| 27.88 | 27.82 |
| 28.46 | 28.40 |
| 27.98 | 28.01 |
| 27.51 | 27.85 |
| 28.23 | 28.29 |
| 27.99 | 28.59 |
| 27.38 | 27.98 |
| 26.92 | 27.36 |
| 27.43 | 27.71 |
| 28.01 | 27.79 |
| 28.66 | 29.79 |
| 27.94 | 27.94 |
| 27.56 | 27.56 |
| 27.69 | 27.71 |
| 27.52 | 28.21 |
| 26.83 | 26.74 |
| 27.05 | 27.21 |
| 28.11 | 28.02 |
| 27.44 | 27.62 |
| 28.38 | 28.59 |
| 29.73 | 29.73 |
| 27.73 | 27.98 |
| 28.70 | 28.52 |
| 28.83 | 28.63 |
| 28.24 | 27.98 |
| 27.82 | 27.56 |
| 28.39 | 28.15 |
| 29.08 | 29.20 |

| Bacillus Ct Assay 1 (Na/Tt/Ba) | Bacillus Ct Assay 2(Ad/Al/Ba) |
|---|---|
| 28.32 | 28.34 |
| 28.27 | 28.32 |
| 28.65 | 28.65 |
| 28.96 | 28.94 |
| 28.11 | 28.22 |
| 28.74 | 28.45 |
| 28.15 | 28.21 |
| 27.39 | 27.62 |
| 30.61 | 31.73 |
| 31.55 | 32.07 |
| 31.84 | 31.89 |
| 31.56 | 31.66 |
| 30.43 | 31.26 |
| 31.09 | 32.21 |
| 30.35 | 31.18 |
| 31.86 | 32.47 |
| 30.66 | 31.55 |
| 31.16 | 32.64 |
| 31.22 | 30.91 |
| 30.74 | 31.29 |
| 32.14 | 32.99 |
| 31.77 | 32.00 |
| 30.62 | 31.36 |
| 31.70 | 31.83 |
| 31.08 | 31.35 |
| 30.69 | 31.42 |
| 31.14 | 31.23 |
| 32.41 | 33.65 |
| 31.52 | 31.33 |
| 31.26 | 32.07 |
| 31.77 | 31.86 |
| 31.98 | 31.34 |
| 31.46 | 31.55 |
| 32.17 | 31.89 |
| 31.44 | 31.54 |
| 30.72 | 31.68 |
| 33.78 | 32.44 |
| 31.94 | 32.77 |
| 32.13 | 33.35 |
| 31.44 | 32.44 |
| 31.34 | 32.20 |
| 30.97 | 31.22 |
| 31.52 | 31.62 |
| 31.51 | 32.04 |
| 31.80 | 33.01 |
| 30.85 | 30.96 |
| 31.83 | 32.09 |
| 31.71 | 31.82 |
| 32.80 | 31.84 |
| 31.50 | 32.78 |

| Bacillus Ct Assay 1 (Na/Tt/Ba) | Bacillus Ct Assay 2(Ad/Al/Ba) |
|---|---|
| 31.21 | 31.48 |
| 33.06 | 36.11 |
| 31.49 | 31.83 |
| 32.19 | 32.35 |
| 30.90 | 32.34 |
| 30.97 | 31.84 |
| 31.52 | 31.08 |
| 31.30 | 31.26 |
| 30.79 | 31.51 |
| 31.69 | 32.35 |
| 32.73 | 34.06 |
| 35.21 | 35.32 |
| 34.20 | 34.21 |
| 33.28 | 32.41 |
| 33.07 | 34.90 |
| 33.55 | 32.73 |
| 33.33 | 33.20 |
| 28.39 | 28.50 |
| 27.11 | 26.84 |
| 27.62 | 27.55 |
| 27.75 | 28.19 |
| 26.09 | 26.63 |
| 26.93 | 26.85 |
| 26.29 | 26.22 |
| 27.34 | 27.58 |
| 27.39 | 27.46 |
| 28.15 | 28.21 |
| 27.94 | 27.89 |
| 28.22 | 28.01 |
| 27.10 | 27.37 |
| 27.33 | 27.35 |
| 26.56 | 26.70 |
| 28.00 | 28.01 |
| 27.40 | 27.31 |
| 26.34 | 26.72 |
| 26.91 | 26.99 |
| 27.07 | 26.98 |
| 26.20 | 26.33 |
| 28.20 | 28.01 |
| 27.17 | 27.39 |
| 27.58 | 27.58 |
| 27.85 | 27.85 |
| 26.92 | 26.93 |
| 28.01 | 27.93 |
| 27.46 | 27.81 |
| 27.10 | 26.98 |
| 27.45 | 27.48 |
| 27.19 | 27.27 |
| 26.46 | 26.67 |
| 37.98 | 31.84 |

| Bacillus Ct Assay 1 (Na/Tt/Ba) | Bacillus Ct Assay 2(Ad/Al/Ba) |
|---|---|
| 31.40 | 31.01 |
| 31.78 | 31.11 |
| 32.63 | 32.99 |
| 31.62 | 32.24 |
| 31.44 | 31.67 |
| 31.06 | 31.35 |
| 30.59 | 30.92 |
| 32.96 | 32.56 |
| 30.90 | 31.28 |
| 32.79 | 32.54 |
| 31.98 | 31.85 |
| 31.62 | 32.65 |
| 31.84 | 31.72 |
| 30.90 | 31.82 |
| 32.26 | 31.69 |
| 31.34 | 31.05 |
| 31.57 | 31.41 |
| 30.88 | 31.62 |
| 30.97 | 32.05 |
| 30.98 | 30.97 |
| 31.19 | 31.24 |
| 31.71 | 32.97 |
| 31.79 | 31.62 |
| 31.52 | 31.80 |
| 30.77 | 31.42 |
| 29.15 | 30.39 |
| 30.56 | 31.36 |
| 32.89 | 31.42 |
| 30.17 | 29.91 |
| 31.28 | 31.07 |
| 30.28 | 30.15 |
| 31.12 | 30.74 |
| 30.38 | 30.72 |
| 31.67 | 31.45 |
| 32.60 | 35.30 |
| 31.49 | 31.54 |
| 32.14 | 32.01 |
| 32.58 | 32.90 |
| 32.47 | 32.38 |
| 33.42 | 32.75 |
| 31.50 | 30.76 |
| 31.51 | 31.54 |
| 32.14 | 31.66 |
| 31.77 | 30.89 |
| 31.84 | 31.72 |
| 33.45 | 33.61 |
| 32.08 | 30.97 |
| 31.52 | 30.43 |
| 32.51 | 32.28 |
| 31.56 | 31.44 |
| 31.23 | 31.46 |
| 30.91 | 31.90 |
| 30.63 | 30.78 |
| 32.23 | 31.78 |
| 32.65 | 32.46 |
| 30.99 | 34.77 |
| 31.61 | 31.11 |
| 31.21 | 31.44 |
| 31.89 | 31.44 |
| 31.77 | 31.91 |
| 30.97 | 30.57 |
| 31.16 | 31.23 |
| 32.08 | 31.92 |

#### A3: Raw Data: WGA STH nucleic acid (II.A.v)

**Table A3. 1**: *Validation of 25 aliquots WGA STH nucleic acid (positive qPCR control). Ct Scores for each STH species are displayed.*

| WGA Run Controls | | | | |
|---|---|---|---|---|
| **Sample** | ***A. duodenale*** | ***A. lumbricoides*** | ***N. americanus*** | ***T. trichiura*** |
| gDNA 1 | 28.57 | 32.30 | 30.89 | 29.02 |
| gDNA 2 | 28.90 | 32.56 | 28.73 | 27.21 |
| gDNA 3 | 29.57 | 33.47 | 29.80 | 28.08 |
| gDNA 4 | 29.57 | 33.15 | 29.57 | 27.97 |
| gDNA 5 | 28.86 | 29.13 | 28.49 | 28.17 |
| gDNA 6 | 29.19 | 32.83 | 28.78 | 27.65 |
| gDNA 7 | 29.15 | 32.87 | 27.91 | 27.76 |
| gDNA 8 | 28.62 | 30.48 | 31.27 | 29.20 |
| gDNA 9 | 29.11 | 29.93 | 29.93 | 27.81 |
| gDNA 10 | 29.19 | 31.21 | 30.10 | 28.37 |
| gDNA 11 | 28.26 | 31.47 | 31.19 | 27.57 |
| gDNA 12 | 28.99 | 31.01 | 28.99 | 27.72 |
| gDNA 13 | 37.47 | 37.84 | 29.10 | 26.94 |
| gDNA 14 | 28.80 | 34.49 | 30.61 | 27.69 |
| gDNA 15 | 31.97 | 33.28 | 29.65 | 27.54 |
| gDNA 16 | 28.47 | 32.38 | 28.88 | 28.12 |
| gDNA 17 | 29.01 | 31.68 | 29.63 | 27.37 |
| gDNA 18 | 29.14 | 28.97 | 29.05 | 27.76 |
| gDNA 19 | 28.93 | 31.19 | 30.34 | 27.46 |
| gDNA 20 | 28.56 | 30.56 | 29.56 | 27.99 |
| gDNA 21 | 28.49 | 32.66 | 29.43 | 28.12 |
| gDNA 22 | 28.52 | 33.07 | 27.74 | 27.89 |
| gDNA 23 | 29.77 | 32.45 | 30.33 | 27.34 |
| gDNA 24 | 28.55 | 29.27 | 30.51 | 27.82 |
| gDNA 25 | 29.65 | 32.55 | 31.24 | 27.66 |

#### A4: Raw Data: Assay Accuracy (II.B.ii)

**Table A4. 1:** *Evaluation of Assay Accuracy by standards. (a) Calculation of Assay Accuracy at the Technical Replicate level. (b) Calculation of Assay Accuracy at the Aliquot level. TP = True Positive, TN = True Negative, FN = False Negative, FP = False Positive.*

**(a)**

| Technical Replicate | | | | | | | | | | | | | | | | | | | | |
|---|---|---|---|---|---|---|---|---|---|---|---|---|---|---|---|---|---|---|---|---|
| | | | | | *T. trichiura* | | | | *A. lumbricoides* | | | | *N. americanus* | | | | *A. duodenale* | | | |
| Standard # | STH Status | | | | TP | TN | FN | FP | TP | TN | FN | FP | TP | TN | FN | FP | TP | TN | FN | FP |
| 1 | | Al | Na | | | 12 | | | 12 | | | | 12 | | | | | 12 | | |
| 2 | | | Na | | | 12 | | | | 12 | | | 12 | | | | | 12 | | |
| 7 | | Al | | | | 12 | | | 12 | | | | | 12 | | | | 12 | | |
| 12v2 | | | Na | | | 12 | | | | 12 | | | 12 | | | | | 12 | | |
| 14 | | Al | | | | 12 | | | 12 | | | | | 12 | | | | 12 | | |
| 15 | | Al | Na | | | 12 | | | 12 | | | | 11 | | | | | 12 | | |
| 18 | | Al | | | | 12 | | | 12 | | | | | 12 | | | | 12 | | |
| 19 | Tt | Al | Na | | 11 | | | | 12 | | | | 11 | | | | | 12 | | |
| 21 | | | | | | 12 | | | | 12 | | | | 12 | | | | 12 | | |
| 22 | | | | | | 12 | | | | 12 | | | | 12 | | | | 12 | | |
| 23 | | | | | | 12 | | | | 12 | | | | 12 | | | | 12 | | |
| 24 | | | | | | 12 | | | | 12 | | | | 12 | | | | 12 | | |
| 25 | | | | | | 12 | | | | 12 | | | | 12 | | | | 12 | | |
| 27 | | Al | Na | Ad | | 11 | | 1 | 12 | | | | 12 | | | | 12 | | | |
| NewTt_4 | Tt | (Al) | Na | | 12 | | | | 2 | 10 | | | 12 | | | | | 12 | | |
| NewTt_5 | Tt | Al | Na | | 12 | | | | 12 | | | | 12 | | | | | 12 | | |
| NewTt_6 | Tt | Al | Na | | 12 | | | | 12 | | | | 12 | | | | | 12 | | |
| NewTt_7 | Tt | Al | (Na) | | 12 | | | | 12 | | | | 5 | 7 | | | | 12 | | |
| Totals | | | | | 59 | 155 | 0 | 1 | 122 | 94 | 0 | 0 | 111 | 103 | 0 | 0 | 12 | 204 | 0 | 0 |
| Accuracy (Tech. Replicate) | | | | | 99.5% | | | | 100.0% | | | | 100.0% | | | | 100.0% | | | |

**(b)**

| Individual Extraction | | | | | | | | | | | | | | | | | | | | |
|---|---|---|---|---|---|---|---|---|---|---|---|---|---|---|---|---|---|---|---|---|
| | | | | | *T. trichiura* | | | | *A. lumbricoides* | | | | *N. americanus* | | | | *A. duodenale* | | | |
| Standard # | STH Status | | | | TP | TN | FN | FP | TP | TN | FN | FP | TP | TN | FN | FP | TP | TN | FN | FP |
| 1 | | Al | Na | | | 3 | | | 3 | | | | 3 | | | | | 3 | | |
| 2 | | | Na | | | 3 | | | | 3 | | | 3 | | | | | 3 | | |
| 7 | | Al | | | | 3 | | | 3 | | | | | 3 | | | | 3 | | |
| 12v2 | | | Na | | | 3 | | | | 3 | | | 3 | | | | | 3 | | |
| 14 | | Al | | | | 3 | | | 3 | | | | | 3 | | | | 3 | | |
| 15 | | Al | Na | | | 3 | | | 3 | | | | 3 | | | | | 3 | | |
| 18 | | Al | | | | 3 | | | 3 | | | | | 3 | | | | 3 | | |
| 19 | Tt | Al | Na | | 3 | | | | 3 | | | | 3 | | | | | 3 | | |
| 21 | | | | | | 3 | | | | 3 | | | | 3 | | | | 3 | | |
| 22 | | | | | | 3 | | | | 3 | | | | 3 | | | | 3 | | |
| 23 | | | | | | 3 | | | | 3 | | | | 3 | | | | 3 | | |
| 24 | | | | | | 3 | | | | 3 | | | | 3 | | | | 3 | | |
| 25 | | | | | | 3 | | | | 3 | | | | 3 | | | | 3 | | |
| 27 | | Al | Na | Ad | | 2 | | 1 | 3 | | | | 3 | | | | 3 | | | |
| NewTt_4 | Tt | (Al) | Na | | 3 | | | | 3 | | | | 3 | | | | | 3 | | |
| NewTt_5 | Tt | Al | Na | | 3 | | | | 3 | | | | 3 | | | | | 3 | | |
| NewTt_6 | Tt | Al | Na | | 3 | | | | 3 | | | | 3 | | | | | 3 | | |
| NewTt_7 | Tt | Al | (Na) | | 3 | | | | 3 | | | | 3 | | | | | 3 | | |
| Totals | | | | | 15 | 38 | 0 | 1 | 33 | 21 | 0 | 0 | 30 | 24 | 0 | 0 | 3 | 51 | 0 | 0 |
| Accuracy (Individual Extract) | | | | | 98.1% | | | | 100.0% | | | | 100.0% | | | | 100.0% | | | |

**Table A4.2:** *T. trichiura Ct Scores for all aliquots and technical replicates of the final standard panel tested at Smith and Quantigen. Note where one aliquot is shown no amplification was observed in all other aliquots and replicates.*

| Sample ID | | ***Trichuris trichiura*** | | | | | | | | |
|---|---|---|---|---|---|---|---|---|---|---|
| | | **Smith** | | | | **Quantigen** | | | | |
| | **Aliquot** | Rep #1 | Rep #2 | Rep #3 | Rep #4 | **Aliquot** | Rep #1 | Rep #2 | Rep #3 | Rep #4 |
| **1** | A | | | | | D | | | | |
| **2** | A | | | | | D | | | | |
| **7** | A | | | | | D | | | | |
| **12v2** | A | | | | | D | | | | |
| **14** | A | | | | | D | | | | |
| **15** | A | | | | | D | | | | |
| **18** | A | | | | | D | | | | |
| **19** | A | 32.2 | 30.5 | 32.2 | 30.8 | D | 34.1 | 32.6 | 33.0 | 33.5 |
| | B | 30.4 | 31.3 | 30.5 | 30.6 | E | | 33.1 | 32.5 | 32.7 |
| | C | 32.2 | 31.8 | 30.6 | 31.0 | F | 32.8 | 35.8 | 39.7 | 35.4 |
| **21** | A | | | | | A | | | | |
| **22** | A | | | | | A | | | | |
| **23** | A | | | | | A | | | | |
| **24** | A | | | | | A | | | | |
| **25** | A | | | | | A | | | | |
| **27** | A | | | | | D | | | | |
| | B | | | | | E | | | 36.76244 | |
| | C | | | | | F | | | | |
| **NewTt_4** | A | 26.6 | 26.3 | 27.0 | 26.2 | D | 32.8 | 33.0 | 32.3 | 32.0 |
| | B | 25.9 | 25.6 | 26.0 | 25.7 | E | 30.0 | 30.1 | 30.3 | 30.1 |
| | C | 26.5 | 26.6 | 26.5 | 26.6 | F | 30.2 | 30.1 | 30.7 | 30.3 |
| **NewTt_5** | A | 26.2 | 25.6 | 26.0 | 26.0 | D | 30.4 | 30.2 | 30.0 | 30.9 |
| | B | 25.1 | 25.0 | 25.1 | 24.8 | E | 30.3 | 29.8 | 29.9 | 29.9 |
| | C | 25.3 | 25.5 | 25.5 | 25.4 | F | 31.1 | 31.4 | 31.3 | 31.0 |
| **New Tt_6** | A | 26.1 | 26.0 | 25.9 | 25.9 | D | 31.0 | 31.5 | 30.8 | 30.9 |
| | B | 26.1 | 26.2 | 26.3 | 25.8 | E | 29.4 | 29.3 | 29.2 | 29.2 |
| | C | 26.8 | 26.4 | 26.1 | 26.4 | F | 29.2 | 29.4 | 29.3 | 29.3 |
| **New Tt_7** | A | 26.4 | 26.2 | 26.5 | 26.2 | D | 29.2 | 30.8 | 30.1 | 30.8 |
| | B | 27.3 | 27.4 | 27.1 | 27.3 | E | 32.4 | 32.4 | 32.4 | 32.0 |
| | C | 26.0 | 26.0 | 26.2 | 26.4 | F | 29.8 | 29.9 | 30.0 | 29.8 |

**Table A4.3:** *A. lumbricoides Ct Scores for all aliquots and technical replicates of the final standard panel tested at Smith and Quantigen. Note where one aliquot is shown no amplification was observed in all other aliquots and replicates.*

| Sample ID | | ***Ascaris lumbricoides*** | | | | | | | | |
|---|---|---|---|---|---|---|---|---|---|---|
| | | **Smith** | | | | **Quantigen** | | | | |
| | **Aliquot** | Rep #1 | Rep #2 | Rep #3 | Rep #4 | **Aliquot** | Rep #1 | Rep #2 | Rep #3 | Rep #4 |
| **1** | A | 20.0 | 19.9 | 20.0 | 19.8 | D | 23.4 | 23.4 | 23.6 | 23.3 |
| | B | 21.8 | 21.8 | 22.2 | 21.9 | E | 25.1 | 26.9 | 26.3 | 25.3 |
| | C | 21.1 | 21.2 | 21.1 | 21.7 | F | 24.4 | 24.6 | 24.5 | 24.6 |
| **2** | A | | | | | D | | | | |
| **7** | A | 15.4 | 16.4 | 16.4 | 16.4 | D | 22.7 | 21.1 | 21.3 | 20.9 |
| | B | 16.2 | 16.2 | 16.4 | 16.5 | E | 20.6 | 23.0 | 23.0 | 20.8 |
| | C | 16.4 | 16.4 | 16.1 | 16.5 | F | 22.6 | 22.2 | 21.9 | 21.7 |
| **12v2** | A | | | | | D | | | | |
| **14** | A | 13.7 | 13.6 | 13.6 | 13.7 | D | 17.4 | 17.4 | 17.4 | 17.3 |
| | B | 13.5 | 13.5 | 13.6 | 13.6 | E | 15.6 | 15.7 | 15.7 | 17.5 |
| | C | 13.5 | 13.7 | 13.4 | 13.4 | F | 18.0 | 17.9 | 17.9 | 17.9 |
| **15** | A | 18.8 | 19.0 | 18.9 | 18.9 | D | 21.5 | 21.5 | 21.5 | 21.5 |
| | B | 18.1 | 18.2 | 18.0 | 18.0 | E | 21.4 | 21.5 | 21.4 | 22.5 |
| | C | 18.7 | 18.8 | 18.8 | 18.6 | F | 24.4 | 24.4 | 24.4 | 24.3 |
| **18** | A | 13.9 | 14.0 | 13.9 | 14.0 | D | 18.2 | 18.3 | 18.3 | 17.8 |
| | B | 14.8 | 14.9 | 14.9 | 14.8 | E | 29.5 | 17.7 | 18.0 | 17.8 |
| | C | 14.6 | 14.6 | 14.6 | 14.6 | F | 18.3 | 18.9 | 18.9 | 18.5 |
| **19** | A | 19.4 | 19.3 | 19.3 | 19.2 | D | 20.8 | 20.8 | 20.6 | 20.6 |
| | B | 18.3 | 18.4 | 18.5 | 18.5 | E | 25.7 | 21.0 | 21.2 | 20.8 |
| | C | 19.0 | 19.1 | 19.3 | 19.0 | F | 20.3 | 21.3 | 21.3 | 20.7 |
| **21** | A | | | | | A | | | | |
| **22** | A | | | | | A | | | | |
| **23** | A | | | | | A | | | | |
| **24** | A | | | | | A | | | | |
| **25** | A | | | | | A | | | | |
| **27** | A | 12.8 | 12.8 | 12.8 | 12.8 | D | 17.0 | 17.0 | 17.3 | 17.6 |
| | B | 12.8 | 12.8 | 12.9 | 12.9 | E | 17.9 | 18.0 | 18.1 | 18.2 |
| | C | 13.3 | 13.4 | 13.3 | 13.3 | F | 18.4 | 19.3 | 18.6 | 18.5 |
| **NewTt_4** | A | | | | | D | 36.9 | | | |
| | B | 33.4 | | | | E | | | | 37.2 |
| | C | 33.1 | | | | F | | | | |
| **NewTt_5** | A | 12.5 | 12.5 | 12.7 | 12.5 | D | 16.7 | 16.5 | 16.6 | 16.6 |
| | B | 12.5 | 12.4 | 12.5 | 12.5 | E | 16.2 | 16.0 | 16.3 | 16.1 |
| | C | 12.2 | 12.0 | 12.2 | 12.2 | F | 16.0 | 16.0 | 15.9 | 16.0 |
| **New Tt_6** | A | 13.2 | 13.2 | 13.3 | 13.4 | D | 16.9 | 16.7 | 16.7 | 16.6 |
| | B | 13.0 | 13.0 | 13.0 | 12.9 | E | 15.9 | 16.0 | 16.2 | 16.0 |
| | C | 13.5 | 13.5 | 13.4 | 13.2 | F | 16.8 | 16.9 | 16.8 | 16.9 |
| **New Tt_7** | A | 13.4 | 13.3 | 13.3 | 13.3 | D | 16.2 | 16.3 | 16.2 | 16.1 |
| | B | 12.9 | 13.0 | 12.8 | 12.7 | E | 15.6 | 15.7 | 15.7 | 15.7 |
| | C | 13.5 | 13.3 | 13.3 | 13.5 | F | 16.5 | 16.7 | 16.6 | 16.6 |

**Table A4. 4**: *N. americanus Ct Scores for all aliquots and technical replicates of the final standard panel tested at Smith and Quantigen. Note where one aliquot is shown no amplification was observed in all other aliquots and replicates.*

| Sample ID | | ***Necator americanus*** | | | | | | | | |
|---|---|---|---|---|---|---|---|---|---|---|
| | | **Smith** | | | | **Quantigen** | | | | |
| | **Aliquot** | Rep #1 | Rep #2 | Rep #3 | Rep #4 | **Aliquot** | Rep #1 | Rep #2 | Rep #3 | Rep #4 |
| **1** | A | 22.1 | 22.2 | 22.1 | 22.1 | D | 21.3 | 21.3 | 21.3 | 21.4 |
| | B | 23.5 | 23.6 | 23.6 | 23.8 | E | 21.9 | 21.4 | 21.5 | 21.9 |
| | C | 21.2 | 21.1 | 20.9 | 20.9 | F | 23.1 | 22.8 | 23.1 | 23.0 |
| **2** | A | 23.9 | 23.9 | 24.0 | 23.9 | D | 27.2 | 27.1 | 27.5 | 27.1 |
| | B | 25.9 | 25.2 | 25.4 | 25.4 | E | 29.6 | 29.5 | 29.9 | 29.7 |
| | C | 22.5 | 22.4 | 22.4 | 22.3 | F | 26.0 | 26.5 | 25.8 | 26.2 |
| **7** | A | | | | | D | | | | |
| **12v2** | A | 25.8 | 26.0 | 25.8 | 25.5 | D | 28.5 | 28.1 | 28.3 | 28.2 |
| | B | 25.1 | 25.0 | 24.7 | 25.2 | E | 27.4 | 27.5 | 26.9 | 27.2 |
| | C | 26.0 | 26.7 | 26.0 | 26.3 | F | 27.5 | 27.5 | 27.4 | 27.4 |
| **14** | A | | | | | D | | | | |
| **15** | A | 22.4 | 22.4 | 22.5 | 22.0 | D | 27.5 | 27.5 | 27.4 | 27.5 |
| | B | 25.2 | 25.0 | 25.4 | 25.1 | E | 28.4 | 28.6 | 28.7 | 28.8 |
| | C | 25.7 | 25.8 | 25.8 | 25.5 | F | | 28.7 | 27.9 | 28.4 |
| **18** | A | | | | | D | | | | |
| **19** | A | 20.8 | 21.0 | 20.9 | 20.8 | D | 21.7 | 21.7 | 21.9 | 22.0 |
| | B | 21.8 | 21.9 | 21.8 | 21.8 | E | | 23.6 | 23.4 | 23.4 |
| | C | 20.9 | 21.1 | 20.9 | 21.0 | F | 22.0 | 22.1 | 22.2 | 22.1 |
| **21** | A | | | | | A | | | | |
| **22** | A | | | | | A | | | | |
| **23** | A | | | | | A | | | | |
| **24** | A | | | | | A | | | | |
| **25** | A | | | | | A | | | | |
| **27** | A | 19.3 | 19.7 | 19.7 | 19.6 | D | 27.2 | 27.8 | 27.5 | 27.8 |
| | B | 18.9 | 18.9 | 19.1 | 19.2 | E | 27.4 | 27.5 | 27.3 | 27.5 |
| | C | 19.6 | 19.7 | 19.5 | 19.7 | F | 27.5 | 27.4 | 27.6 | 27.4 |
| **NewTt_4** | A | 17.8 | 17.6 | 17.8 | 17.7 | D | 25.6 | 25.7 | 25.5 | 25.8 |
| | B | 20.4 | 20.3 | 20.3 | 19.8 | E | 21.7 | 21.8 | 21.5 | 21.7 |
| | C | 20.0 | 19.9 | 19.5 | 20.0 | F | 22.9 | 22.9 | 23.0 | 23.3 |
| **NewTt_5** | A | 18.6 | 19.0 | 19.2 | 19.0 | D | 25.1 | 25.1 | 25.2 | 25.3 |
| | B | 18.7 | 18.7 | 19.0 | 18.4 | E | 21.9 | 21.9 | 21.7 | 21.8 |
| | C | 18.1 | 18.1 | 17.7 | 17.8 | F | 21.9 | 21.8 | 22.0 | 22.3 |
| **New Tt_6** | A | 20.1 | 19.6 | 20.0 | 20.1 | D | 22.6 | 22.6 | 22.6 | 22.7 |
| | B | 18.6 | 18.4 | 18.3 | 18.5 | E | 21.2 | 21.3 | 21.2 | 21.2 |
| | C | 18.0 | 18.3 | 17.6 | 17.8 | F | 22.1 | 22.1 | 22.2 | 22.4 |
| **New Tt_7** | A | | | | 34.4 | D | | | | |
| | B | | | | | E | 29.3 | 29.8 | 29.8 | 29.7 |
| | C | | | | | F | | | 37.0 | |

**Table A4. 5:** *A. duodenale Ct Scores for all aliquots and technical replicates of the final standard panel tested at Smith and Quantigen. Note where one aliquot is shown no amplification was observed in all other aliquots and replicates.*

| Sample ID | | *Ancylostoma duodenale* | | | | | | | | |
|---|---|---|---|---|---|---|---|---|---|---|
| | | Smith | | | | Quantigen | | | | |
| | Aliquot | Rep #1 | Rep #2 | Rep #3 | Rep #4 | Aliquot | Rep #1 | Rep #2 | Rep #3 | Rep #4 |
| 1 | A | | | | | D | | | | |
| 2 | A | | | | | D | | | | |
| 7 | A | | | | | D | | | | |
| 12v2 | A | | | | | D | | | | |
| 14 | A | | | | | D | | | | |
| 15 | A | | | | | D | | | | |
| 18 | A | | | | | D | | | | |
| 19 | A | | | | | D | | | | |
| 21 | A | | | | | A | | | | |
| 22 | A | | | | | A | | | | |
| 23 | A | | | | | A | | | | |
| 24 | A | | | | | A | | | | |
| 25 | A | | | | | A | | | | |
| 27 | A | 25.6 | 25.9 | 26.1 | 25.9 | D | 30.4 | 30.4 | 30.2 | 30.3 |
| | B | 26.2 | 26.3 | 26.0 | 26.3 | E | 30.2 | 30.3 | 31.3 | 32.8 |
| | C | 25.4 | 25.9 | 25.8 | 25.9 | F | 31.3 | 31.5 | 39.9 | 36.7 |
| NewTt_4 | A | | | | | D | | | | |
| NewTt_5 | A | | | | | D | | | | |
| New Tt_6 | A | | | | | D | | | | |
| New Tt_7 | A | | | | | D | | | | |

#### A5: Raw Data: Assay Sensitivity/LOD (II.B.v)

***Table A5. 1**: Limit of detection for STH. Each STH standard was serially diluted and qPCR was performed (a) 1:10 Serial Dilution Results (b) 1:2 Serial Dilution Results. Green highlighting indicates that the dilution yielded CT scores for all technical replicates. Red indicates the run contained at least 1 undeterminable Ct score. Yellow highlighting indicates the dilution from which to begin the 1:2 serial. Black indicates that the dilution was not run.*

**(a)**

| First round of dilutions (1:10) | | | | | | |
|---|---|---|---|---|---|---|
| **Sample** | First Dilution Point | 2nd | 3rd | 4th | 5th | 6th |
| **N. americanus** – StdExt1_6_Na | 1.00E-01 | 1.00E-02 | 1.00E-03 | 1.00E-04 | 1.00E-05 | 1.00E-06 |
| **Positives** | 10/10 | 10/10 | 10/10 | 10/10 | 3/10 | 0/10 |
| **A. lumbricoides** – StdExt1_6_Al | 1.00E-01 | 1.00E-02 | 1.00E-03 | 1.00E-04 | 1.00E-05 | 1.00E-06 |
| **Positives** | 10/10 | 10/10 | 10/10 | 8/10 | 2/10 | 1/10 |
| **A. duodenale** – StdExt1_208_Ad | 1.00E-01 | 1.00E-02 | 1.00E-03 | 1.00E-04 | 1.00E-05 | 1.00E-06 |
| **Positives** | 10/10 | 9/10 | 1/10 | 0/10 | 0/10 | 0/10 |
| **T. trichiura** – StdExt2_44_Tt | 1.00E-01 | 1.00E-02 | 1.00E-03 | 1.00E-04 | 1.00E-05 | 1.00E-06 |
| **Positives** | 10/10 | 10/10 | 10/10 | 9/10 | 0/10 | |

| **Key** | |
|---|---|
| *Aliquot ID* | *Standard #* |
| StdExt1_6 | 1 |
| StdExt1_208 | 27 |
| StdExt2_44 | 19 |

**(b)**

| Second round of dilutions (1:2) | | | | | | | |
|---|---|---|---|---|---|---|---|
| **Sample** | First Dilution Point | 2nd | 3rd | 4th | 5th | 6th | 7th |
| **N. americanus** – StdExt1_6_Na | 1.00E-04 | 5.00E-05 | 2.50E-05 | 1.25E-05 | | | |
| **Positives** | 20/20 | 20/20 | 20/20 | 18/20 | | | |
| **A. lumbricoides** – StdExt1_6_Al | 1.00E-03 | 5.00E-04 | 2.50E-04 | 1.25E-04 | 6.25E-05 | 3.13E-05 | 1.56E-05 |
| **Positives** | 20/20 | 20/20 | 20/20 | 20/20 | 17/20 | 4/20 | 4/20 |
| **A. duodenale** – StdExt1_208_Ad | 1.00E-01 | 5.00E-02 | 2.50E-02 | 1.25E-02 | 6.25E-03 | 3.13E-03 | 1.56E-03 |
| **Positives** | 20/20 | 20/20 | 20/20 | 20/20 | 15/20 | 11/20 | 9/20 |
| **T. trichiura** – StdExt2_44_Tt | 1.00E-03 | 5.00E-04 | 2.50E-04 | 1.25E-04 | 6.25E-05 | 3.13E-05 | 1.56E-05 |
| **Positives** | 20/20 | 20/20 | 20/20 | 19/20 | 16/20 | 10/20 | 7/20 |

***Table A5. 2***: *1:10 Serial Dilution Ct Scores for N. americanus. Note there are 10 replicates of each dilution point.*

| First Dilution (1:10) | | | | | |
|---|---|---|---|---|---|
| Well | Sample Name | Target Name | CT | | |
| 1 | StdExt1_6_Na_0.1_1 | N. americanus | 22.596 | | |
| 10 | StdExt1_6_Na_0.1_10 | N. americanus | 21.807 | | |
| 2 | StdExt1_6_Na_0.1_2 | N. americanus | 21.788 | | |
| 3 | StdExt1_6_Na_0.1_3 | N. americanus | 21.742 | | |
| 4 | StdExt1_6_Na_0.1_4 | N. americanus | 22.392 | | |
| 5 | StdExt1_6_Na_0.1_5 | N. americanus | 21.704 | | |
| 6 | StdExt1_6_Na_0.1_6 | N. americanus | 22.286 | | |
| 7 | StdExt1_6_Na_0.1_7 | N. americanus | 21.698 | | |
| 8 | StdExt1_6_Na_0.1_8 | N. americanus | 22.163 | | |
| 9 | StdExt1_6_Na_0.1_9 | N. americanus | 21.784 | 1.00E-01 | 1st Dilution |
| 49 | StdExt1_6_Na_0.01_1 | N. americanus | 25.696 | | |
| 58 | StdExt1_6_Na_0.01_10 | N. americanus | 24.741 | | |
| 50 | StdExt1_6_Na_0.01_2 | N. americanus | 25.620 | | |
| 51 | StdExt1_6_Na_0.01_3 | N. americanus | 25.859 | | |
| 52 | StdExt1_6_Na_0.01_4 | N. americanus | 24.781 | | |
| 53 | StdExt1_6_Na_0.01_5 | N. americanus | 26.013 | | |
| 54 | StdExt1_6_Na_0.01_6 | N. americanus | 24.899 | | |
| 55 | StdExt1_6_Na_0.01_7 | N. americanus | 25.224 | | |
| 56 | StdExt1_6_Na_0.01_8 | N. americanus | 24.996 | | |
| 57 | StdExt1_6_Na_0.01_9 | N. americanus | 25.071 | 1.00E-02 | 2nd Dilution |
| 97 | StdExt1_6_Na_0.001_1 | N. americanus | 28.668 | | |
| 106 | StdExt1_6_Na_0.001_10 | N. americanus | 29.081 | | |
| 98 | StdExt1_6_Na_0.001_2 | N. americanus | 28.961 | | |
| 99 | StdExt1_6_Na_0.001_3 | N. americanus | 28.928 | | |
| 100 | StdExt1_6_Na_0.001_4 | N. americanus | 28.121 | | |
| 101 | StdExt1_6_Na_0.001_5 | N. americanus | 29.189 | | |
| 102 | StdExt1_6_Na_0.001_6 | N. americanus | 28.148 | | |
| 103 | StdExt1_6_Na_0.001_7 | N. americanus | 29.131 | | |
| 104 | StdExt1_6_Na_0.001_8 | N. americanus | 28.790 | | |
| 105 | StdExt1_6_Na_0.001_9 | N. americanus | 27.991 | 1.00E-03 | 3rd Dilution |
| 145 | StdExt1_6_Na_0.0001_1 | N. americanus | 31.092 | | |
| 154 | StdExt1_6_Na_0.0001_10 | N. americanus | 31.348 | | |
| 146 | StdExt1_6_Na_0.0001_2 | N. americanus | 31.89536095 | | |
| 147 | StdExt1_6_Na_0.0001_3 | N. americanus | 31.308 | | |
| 148 | StdExt1_6_Na_0.0001_4 | N. americanus | 35.81274796 | | |
| 149 | StdExt1_6_Na_0.0001_5 | N. americanus | 31.985 | | |
| 150 | StdExt1_6_Na_0.0001_6 | N. americanus | 32.653 | | |
| 151 | StdExt1_6_Na_0.0001_7 | N. americanus | 32.847 | | |
| 152 | StdExt1_6_Na_0.0001_8 | N. americanus | 33.163 | | |
| 153 | StdExt1_6_Na_0.0001_9 | N. americanus | 31.540 | 1.00E-04 | 4th Dilution |
| 193 | StdExt1_6_Na_0.00001_1 | N. americanus | 30.832 | | |
| 202 | StdExt1_6_Na_0.00001_10 | N. americanus | Undetermined | | |
| 194 | StdExt1_6_Na_0.00001_2 | N. americanus | Undetermined | | |
| 195 | StdExt1_6_Na_0.00001_3 | N. americanus | Undetermined | | |
| 196 | StdExt1_6_Na_0.00001_4 | N. americanus | Undetermined | | |
| 197 | StdExt1_6_Na_0.00001_5 | N. americanus | 36.29724503 | | |
| 198 | StdExt1_6_Na_0.00001_6 | N. americanus | Undetermined | | |
| 199 | StdExt1_6_Na_0.00001_7 | N. americanus | Undetermined | | |
| 200 | StdExt1_6_Na_0.00001_8 | N. americanus | 36.0435257 | | |
| 201 | StdExt1_6_Na_0.00001_9 | N. americanus | Undetermined | 1.00E-05 | 5th Dilution |
| 241 | StdExt1_6_Na_0.000001_1 | N. americanus | Undetermined | | |
| 250 | StdExt1_6_Na_0.000001_10 | N. americanus | Undetermined | | |
| 242 | StdExt1_6_Na_0.000001_2 | N. americanus | Undetermined | | |
| 243 | StdExt1_6_Na_0.000001_3 | N. americanus | Undetermined | | |
| 244 | StdExt1_6_Na_0.000001_4 | N. americanus | Undetermined | | |
| 245 | StdExt1_6_Na_0.000001_5 | N. americanus | Undetermined | | |
| 246 | StdExt1_6_Na_0.000001_6 | N. americanus | Undetermined | | |
| 247 | StdExt1_6_Na_0.000001_7 | N. americanus | Undetermined | | |
| 248 | StdExt1_6_Na_0.000001_8 | N. americanus | Undetermined | | |
| 249 | StdExt1_6_Na_0.000001_9 | N. americanus | Undetermined | 1.00E-06 | 6th Dilution |

***Table A5. 3**: 1:2 Serial Dilution Ct Scores for N. americanus. Note there are 20 replicates of each dilution point.*

| Second Dilution (1:2) | | | | | |
|---|---|---|---|---|---|
| Well | Sample Name | Target Name | CT | | |
| 23 | NTC | N. americanus | Undetermined | | |
| 24 | Pos. PCR | N. americanus | 26.950 | | |
| 1 | StdExt1_6_Na_D1_R1 | N. americanus | 30.290 | 1.00E-04 | 1st Dilution |
| 19 | StdExt1_6_Na_D1_R10 | N. americanus | 30.510 | | |
| 2 | StdExt1_6_Na_D1_R11 | N. americanus | 30.302 | | |
| 4 | StdExt1_6_Na_D1_R12 | N. americanus | 30.999 | | |
| 6 | StdExt1_6_Na_D1_R13 | N. americanus | 31.616 | | |
| 8 | StdExt1_6_Na_D1_R14 | N. americanus | 31.217 | | |
| 10 | StdExt1_6_Na_D1_R15 | N. americanus | 31.905 | | |
| 12 | StdExt1_6_Na_D1_R16 | N. americanus | 31.162 | | |
| 14 | StdExt1_6_Na_D1_R17 | N. americanus | 31.200 | | |
| 16 | StdExt1_6_Na_D1_R18 | N. americanus | 30.342 | | |
| 18 | StdExt1_6_Na_D1_R19 | N. americanus | 30.876 | | |
| 3 | StdExt1_6_Na_D1_R2 | N. americanus | 31.453 | | |
| 20 | StdExt1_6_Na_D1_R20 | N. americanus | 31.631 | | |
| 5 | StdExt1_6_Na_D1_R3 | N. americanus | 31.597 | | |
| 7 | StdExt1_6_Na_D1_R4 | N. americanus | 31.240 | | |
| 9 | StdExt1_6_Na_D1_R5 | N. americanus | 30.940 | | |
| 11 | StdExt1_6_Na_D1_R6 | N. americanus | 30.964 | | |
| 13 | StdExt1_6_Na_D1_R7 | N. americanus | 30.592 | | |
| 15 | StdExt1_6_Na_D1_R8 | N. americanus | 31.330 | | |
| 17 | StdExt1_6_Na_D1_R9 | N. americanus | 32.218 | | |
| 49 | StdExt1_6_Na_D2_R1 | N. americanus | 32.909 | 5.00E-05 | 2nd Dilution |
| 67 | StdExt1_6_Na_D2_R10 | N. americanus | 33.023 | | |
| 50 | StdExt1_6_Na_D2_R11 | N. americanus | 32.564 | | |
| 52 | StdExt1_6_Na_D2_R12 | N. americanus | 32.635 | | |
| 54 | StdExt1_6_Na_D2_R13 | N. americanus | 34.877 | | |
| 56 | StdExt1_6_Na_D2_R14 | N. americanus | 31.207 | | |
| 58 | StdExt1_6_Na_D2_R15 | N. americanus | 31.582 | | |
| 60 | StdExt1_6_Na_D2_R16 | N. americanus | 33.777 | | |
| 62 | StdExt1_6_Na_D2_R17 | N. americanus | 32.335 | | |
| 64 | StdExt1_6_Na_D2_R18 | N. americanus | 31.951 | | |
| 66 | StdExt1_6_Na_D2_R19 | N. americanus | 33.287 | | |
| 51 | StdExt1_6_Na_D2_R2 | N. americanus | 32.747 | | |
| 68 | StdExt1_6_Na_D2_R20 | N. americanus | 32.224 | | |
| 53 | StdExt1_6_Na_D2_R3 | N. americanus | 31.441 | | |
| 55 | StdExt1_6_Na_D2_R4 | N. americanus | 33.075 | | |
| 57 | StdExt1_6_Na_D2_R5 | N. americanus | 31.952 | | |
| 59 | StdExt1_6_Na_D2_R6 | N. americanus | 33.297 | | |
| 61 | StdExt1_6_Na_D2_R7 | N. americanus | 32.993 | | |
| 63 | StdExt1_6_Na_D2_R8 | N. americanus | 31.082 | | |
| 65 | StdExt1_6_Na_D2_R9 | N. americanus | 32.950 | | |
| 97 | StdExt1_6_Na_D3_R1 | N. americanus | 32.718 | 2.50E-05 | 3rd Dilution |
| 115 | StdExt1_6_Na_D3_R10 | N. americanus | 33.694 | | |
| 98 | StdExt1_6_Na_D3_R11 | N. americanus | 31.805 | | |
| 100 | StdExt1_6_Na_D3_R12 | N. americanus | 35.434 | | |
| 102 | StdExt1_6_Na_D3_R13 | N. americanus | 32.817 | | |
| 104 | StdExt1_6_Na_D3_R14 | N. americanus | 33.682 | | |
| 106 | StdExt1_6_Na_D3_R15 | N. americanus | 33.135 | | |
| 108 | StdExt1_6_Na_D3_R16 | N. americanus | 34.605 | | |
| 110 | StdExt1_6_Na_D3_R17 | N. americanus | 34.548 | | |
| 112 | StdExt1_6_Na_D3_R18 | N. americanus | 35.765 | | |
| 114 | StdExt1_6_Na_D3_R19 | N. americanus | 32.246 | | |
| 99 | StdExt1_6_Na_D3_R2 | N. americanus | 31.883 | | |
| 116 | StdExt1_6_Na_D3_R20 | N. americanus | 32.658 | | |
| 101 | StdExt1_6_Na_D3_R3 | N. americanus | 33.425 | | |
| 103 | StdExt1_6_Na_D3_R4 | N. americanus | 38.347 | | |
| 105 | StdExt1_6_Na_D3_R5 | N. americanus | 32.610 | | |
| 107 | StdExt1_6_Na_D3_R6 | N. americanus | 33.578 | | |
| 109 | StdExt1_6_Na_D3_R7 | N. americanus | 32.713 | | |
| 111 | StdExt1_6_Na_D3_R8 | N. americanus | 32.915 | | |
| 113 | StdExt1_6_Na_D3_R9 | N. americanus | 33.874 | | |

| Second Dilution (1:2) | | | | | |
|---|---|---|---|---|---|
| Well | Sample Name | Target Name | CT | | |
| 145 | StdExt1_6_Na_D4_R1 | N. americanus | 33.585 | 1.25E-05 | 4th Dilution |
| 163 | StdExt1_6_Na_D4_R10 | N. americanus | 32.114 | | |
| 146 | StdExt1_6_Na_D4_R11 | N. americanus | 35.518 | | |
| 148 | StdExt1_6_Na_D4_R12 | N. americanus | 34.381 | | |
| 150 | StdExt1_6_Na_D4_R13 | N. americanus | 35.530 | | |
| 152 | StdExt1_6_Na_D4_R14 | N. americanus | 34.630 | | |
| 154 | StdExt1_6_Na_D4_R15 | N. americanus | 35.581 | | |
| 156 | StdExt1_6_Na_D4_R16 | N. americanus | 33.914 | | |
| 158 | StdExt1_6_Na_D4_R17 | N. americanus | 36.208 | | |
| 160 | StdExt1_6_Na_D4_R18 | N. americanus | 34.616 | | |
| 162 | StdExt1_6_Na_D4_R19 | N. americanus | 34.376 | | |
| 147 | StdExt1_6_Na_D4_R2 | N. americanus | 33.340 | | |
| 164 | StdExt1_6_Na_D4_R20 | N. americanus | 34.052 | | |
| 149 | StdExt1_6_Na_D4_R3 | N. americanus | Undetermined | | |
| 151 | StdExt1_6_Na_D4_R4 | N. americanus | 32.252 | | |
| 153 | StdExt1_6_Na_D4_R5 | N. americanus | 32.530 | | |
| 155 | StdExt1_6_Na_D4_R6 | N. americanus | 31.538 | | |
| 157 | StdExt1_6_Na_D4_R7 | N. americanus | 35.085 | | |
| 159 | StdExt1_6_Na_D4_R8 | N. americanus | 34.682 | | |
| 161 | StdExt1_6_Na_D4_R9 | N. americanus | Undetermined | | |

**Table A5. 4**: *1:2 Serial Dilution Ct Scores for A. lumbricoides. Note there are 10 replicates of each dilution point.*

| First Dilution (1:10) | | | | | |
|---|---|---|---|---|---|
| Well | Sample Name | Target Name | CT | | |
| 25 | StdExt1_6_Al_0.1_1 | A.lumbricoides | 22.880 | | |
| 34 | StdExt1_6_Al_0.1_10 | A.lumbricoides | 23.029 | | |
| 26 | StdExt1_6_Al_0.1_2 | A.lumbricoides | 23.886 | | |
| 27 | StdExt1_6_Al_0.1_3 | A.lumbricoides | 23.215 | | |
| 28 | StdExt1_6_Al_0.1_4 | A.lumbricoides | 23.744 | | |
| 29 | StdExt1_6_Al_0.1_5 | A.lumbricoides | 23.168 | | |
| 30 | StdExt1_6_Al_0.1_6 | A.lumbricoides | 27.280 | | |
| 31 | StdExt1_6_Al_0.1_7 | A.lumbricoides | 23.156 | | |
| 32 | StdExt1_6_Al_0.1_8 | A.lumbricoides | 23.856 | | |
| 33 | StdExt1_6_Al_0.1_9 | A.lumbricoides | 23.259 | 1.00E-01 | 1st Dilution |
| 73 | StdExt1_6_Al_0.01_1 | A.lumbricoides | 26.410 | | |
| 82 | StdExt1_6_Al_0.01_10 | A.lumbricoides | 26.905 | | |
| 74 | StdExt1_6_Al_0.01_2 | A.lumbricoides | 26.037 | | |
| 75 | StdExt1_6_Al_0.01_3 | A.lumbricoides | 26.340 | | |
| 76 | StdExt1_6_Al_0.01_4 | A.lumbricoides | 25.949 | | |
| 77 | StdExt1_6_Al_0.01_5 | A.lumbricoides | 25.986 | | |
| 78 | StdExt1_6_Al_0.01_6 | A.lumbricoides | 26.292 | | |
| 79 | StdExt1_6_Al_0.01_7 | A.lumbricoides | 25.980 | | |
| 80 | StdExt1_6_Al_0.01_8 | A.lumbricoides | 28.906 | | |
| 81 | StdExt1_6_Al_0.01_9 | A.lumbricoides | 26.505 | 1.00E-02 | 2nd Dilution |
| 121 | StdExt1_6_Al_0.001_1 | A.lumbricoides | 30.009 | | |
| 130 | StdExt1_6_Al_0.001_10 | A.lumbricoides | 30.876 | | |
| 122 | StdExt1_6_Al_0.001_2 | A.lumbricoides | 30.349 | | |
| 123 | StdExt1_6_Al_0.001_3 | A.lumbricoides | 28.970 | | |
| 124 | StdExt1_6_Al_0.001_4 | A.lumbricoides | 29.121 | | |
| 125 | StdExt1_6_Al_0.001_5 | A.lumbricoides | 30.860 | | |
| 126 | StdExt1_6_Al_0.001_6 | A.lumbricoides | 30.264 | | |
| 127 | StdExt1_6_Al_0.001_7 | A.lumbricoides | 30.362 | | |
| 128 | StdExt1_6_Al_0.001_8 | A.lumbricoides | 29.713 | | |
| 129 | StdExt1_6_Al_0.001_9 | A.lumbricoides | 30.745 | 1.00E-03 | 3rd Dilution |
| 169 | StdExt1_6_Na_0.0001_1 | A.lumbricoides | 34.878 | | |
| 178 | StdExt1_6_Na_0.0001_10 | A.lumbricoides | 35.157 | | |
| 170 | StdExt1_6_Na_0.0001_2 | A.lumbricoides | Undetermined | | |
| 171 | StdExt1_6_Na_0.0001_3 | A.lumbricoides | 35.434 | | |
| 172 | StdExt1_6_Na_0.0001_4 | A.lumbricoides | Undetermined | | |
| 173 | StdExt1_6_Na_0.0001_5 | A.lumbricoides | 35.858 | | |
| 174 | StdExt1_6_Na_0.0001_6 | A.lumbricoides | 39.186 | | |
| 175 | StdExt1_6_Na_0.0001_7 | A.lumbricoides | 38.760 | | |
| 176 | StdExt1_6_Na_0.0001_8 | A.lumbricoides | 37.116 | | |
| 177 | StdExt1_6_Na_0.0001_9 | A.lumbricoides | 39.221 | 1.00E-04 | 4th Dilution |
| 217 | StdExt1_6_Al_0.00001_1 | A.lumbricoides | 27.074 | | |
| 226 | StdExt1_6_Al_0.00001_10 | A.lumbricoides | Undetermined | | |
| 218 | StdExt1_6_Al_0.00001_2 | A.lumbricoides | 32.781 | | |
| 219 | StdExt1_6_Al_0.00001_3 | A.lumbricoides | Undetermined | | |
| 220 | StdExt1_6_Al_0.00001_4 | A.lumbricoides | Undetermined | | |
| 221 | StdExt1_6_Al_0.00001_5 | A.lumbricoides | Undetermined | | |
| 222 | StdExt1_6_Al_0.00001_6 | A.lumbricoides | Undetermined | | |
| 223 | StdExt1_6_Al_0.00001_7 | A.lumbricoides | Undetermined | | |
| 224 | StdExt1_6_Al_0.00001_8 | A.lumbricoides | Undetermined | | |
| 225 | StdExt1_6_Al_0.00001_9 | A.lumbricoides | Undetermined | 1.00E-05 | 5th Dilution |
| 265 | StdExt1_6_Al_0.000001_1 | A.lumbricoides | Undetermined | | |
| 274 | StdExt1_6_Al_0.000001_10 | A.lumbricoides | Undetermined | | |
| 266 | StdExt1_6_Al_0.000001_2 | A.lumbricoides | Undetermined | | |
| 267 | StdExt1_6_Al_0.000001_3 | A.lumbricoides | Undetermined | | |
| 268 | StdExt1_6_Al_0.000001_4 | A.lumbricoides | 37.743 | | |
| 269 | StdExt1_6_Al_0.000001_5 | A.lumbricoides | Undetermined | | |
| 270 | StdExt1_6_Al_0.000001_6 | A.lumbricoides | Undetermined | | |
| 271 | StdExt1_6_Al_0.000001_7 | A.lumbricoides | Undetermined | | |
| 272 | StdExt1_6_Al_0.000001_8 | A.lumbricoides | Undetermined | | |
| 273 | StdExt1_6_Al_0.000001_9 | A.lumbricoides | Undetermined | 1.00E-06 | 6th Dilution |

**Table A5. 5**: *1:2 Serial Dilution Ct Scores for A. lumbricoides. Note there are 20 replicates of each dilution point.*

| Second Dilution (1:2) | | | | | |
|---|---|---|---|---|---|
| Well | Sample Name | Target Name | CT | | |
| 25 | StdExt1_6_Al_D1_R1 | A.lumbricoides | 30.474 | | |
| 43 | StdExt1_6_Al_D1_R10 | A.lumbricoides | 30.462 | | |
| 26 | StdExt1_6_Al_D1_R11 | A.lumbricoides | 30.425 | | |
| 28 | StdExt1_6_Al_D1_R12 | A.lumbricoides | 30.024 | | |
| 30 | StdExt1_6_Al_D1_R13 | A.lumbricoides | 29.543 | | |
| 32 | StdExt1_6_Al_D1_R14 | A.lumbricoides | 30.062 | | |
| 34 | StdExt1_6_Al_D1_R15 | A.lumbricoides | 29.836 | | |
| 36 | StdExt1_6_Al_D1_R16 | A.lumbricoides | 30.571 | | |
| 38 | StdExt1_6_Al_D1_R17 | A.lumbricoides | 30.291 | | |
| 40 | StdExt1_6_Al_D1_R18 | A.lumbricoides | 29.929 | | |
| 42 | StdExt1_6_Al_D1_R19 | A.lumbricoides | 30.159 | 1.00E-03 | 1st Dilution |
| 27 | StdExt1_6_Al_D1_R2 | A.lumbricoides | 29.355 | | |
| 44 | StdExt1_6_Al_D1_R20 | A.lumbricoides | 29.844 | | |
| 29 | StdExt1_6_Al_D1_R3 | A.lumbricoides | 29.202 | | |
| 31 | StdExt1_6_Al_D1_R4 | A.lumbricoides | 30.090 | | |
| 33 | StdExt1_6_Al_D1_R5 | A.lumbricoides | 28.853 | | |
| 35 | StdExt1_6_Al_D1_R6 | A.lumbricoides | 29.889 | | |
| 37 | StdExt1_6_Al_D1_R7 | A.lumbricoides | 29.960 | | |
| 39 | StdExt1_6_Al_D1_R8 | A.lumbricoides | 29.007 | | |
| 41 | StdExt1_6_Al_D1_R9 | A.lumbricoides | 30.490 | | |
| 73 | StdExt1_6_Al_D2_R1 | A.lumbricoides | 31.922 | | |
| 91 | StdExt1_6_Al_D2_R10 | A.lumbricoides | 30.870 | | |
| 74 | StdExt1_6_Al_D2_R11 | A.lumbricoides | 31.513 | | |
| 76 | StdExt1_6_Al_D2_R12 | A.lumbricoides | 31.591 | | |
| 78 | StdExt1_6_Al_D2_R13 | A.lumbricoides | 31.125 | | |
| 80 | StdExt1_6_Al_D2_R14 | A.lumbricoides | 31.877 | | |
| 82 | StdExt1_6_Al_D2_R15 | A.lumbricoides | 30.000 | | |
| 84 | StdExt1_6_Al_D2_R16 | A.lumbricoides | 30.874 | | |
| 86 | StdExt1_6_Al_D2_R17 | A.lumbricoides | 30.727 | | |
| 88 | StdExt1_6_Al_D2_R18 | A.lumbricoides | 30.458 | | |
| 90 | StdExt1_6_Al_D2_R19 | A.lumbricoides | 31.793 | 5.00E-04 | 2nd Dilution |
| 75 | StdExt1_6_Al_D2_R2 | A.lumbricoides | 30.998 | | |
| 92 | StdExt1_6_Al_D2_R20 | A.lumbricoides | 29.650 | | |
| 77 | StdExt1_6_Al_D2_R3 | A.lumbricoides | 32.154 | | |
| 79 | StdExt1_6_Al_D2_R4 | A.lumbricoides | 31.356 | | |
| 81 | StdExt1_6_Al_D2_R5 | A.lumbricoides | 32.255 | | |
| 83 | StdExt1_6_Al_D2_R6 | A.lumbricoides | 30.740 | | |
| 85 | StdExt1_6_Al_D2_R7 | A.lumbricoides | 31.506 | | |
| 87 | StdExt1_6_Al_D2_R8 | A.lumbricoides | 31.110 | | |
| 89 | StdExt1_6_Al_D2_R9 | A.lumbricoides | 32.324 | | |
| 121 | StdExt1_6_Al_D3_R1 | A.lumbricoides | 33.847 | | |
| 139 | StdExt1_6_Al_D3_R10 | A.lumbricoides | 31.042 | | |
| 122 | StdExt1_6_Al_D3_R11 | A.lumbricoides | 30.956 | | |
| 124 | StdExt1_6_Al_D3_R12 | A.lumbricoides | 32.504 | | |
| 126 | StdExt1_6_Al_D3_R13 | A.lumbricoides | 33.273 | | |
| 128 | StdExt1_6_Al_D3_R14 | A.lumbricoides | 34.463 | | |
| 130 | StdExt1_6_Al_D3_R15 | A.lumbricoides | 32.057 | | |
| 132 | StdExt1_6_Al_D3_R16 | A.lumbricoides | 31.146 | | |
| 134 | StdExt1_6_Al_D3_R17 | A.lumbricoides | 32.633 | | |
| 136 | StdExt1_6_Al_D3_R18 | A.lumbricoides | 33.740 | | |
| 138 | StdExt1_6_Al_D3_R19 | A.lumbricoides | 32.245 | 2.50E-04 | 3rd Dilution |
| 123 | StdExt1_6_Al_D3_R2 | A.lumbricoides | 33.904 | | |
| 140 | StdExt1_6_Al_D3_R20 | A.lumbricoides | 32.299 | | |
| 125 | StdExt1_6_Al_D3_R3 | A.lumbricoides | 32.407 | | |
| 127 | StdExt1_6_Al_D3_R4 | A.lumbricoides | 31.218 | | |
| 129 | StdExt1_6_Al_D3_R5 | A.lumbricoides | 32.479 | | |
| 131 | StdExt1_6_Al_D3_R6 | A.lumbricoides | 31.007 | | |
| 133 | StdExt1_6_Al_D3_R7 | A.lumbricoides | 32.190 | | |
| 135 | StdExt1_6_Al_D3_R8 | A.lumbricoides | 30.842 | | |
| 137 | StdExt1_6_Al_D3_R9 | A.lumbricoides | 31.322 | | |

| Second Dilution (1:2) | | | | | |
|---|---|---|---|---|---|
| Well | Sample Name | Target Name | CT | | |
| 169 | StdExt1_6_Al_D4_R1 | A.lumbricoides | 35.216 | | |
| 187 | StdExt1_6_Al_D4_R10 | A.lumbricoides | 31.984 | | |
| 170 | StdExt1_6_Al_D4_R11 | A.lumbricoides | 34.237 | | |
| 172 | StdExt1_6_Al_D4_R12 | A.lumbricoides | 33.063 | | |
| 174 | StdExt1_6_Al_D4_R13 | A.lumbricoides | 32.529 | | |
| 176 | StdExt1_6_Al_D4_R14 | A.lumbricoides | 33.823 | | |
| 178 | StdExt1_6_Al_D4_R15 | A.lumbricoides | 33.165 | | |
| 180 | StdExt1_6_Al_D4_R16 | A.lumbricoides | 35.175 | | |
| 182 | StdExt1_6_Al_D4_R17 | A.lumbricoides | 31.635 | | |
| 184 | StdExt1_6_Al_D4_R18 | A.lumbricoides | 33.941 | 1.25E-04 | 4th Dilution |
| 186 | StdExt1_6_Al_D4_R19 | A.lumbricoides | 32.639 | | |
| 171 | StdExt1_6_Al_D4_R2 | A.lumbricoides | 34.165 | | |
| 188 | StdExt1_6_Al_D4_R20 | A.lumbricoides | 33.353 | | |
| 173 | StdExt1_6_Al_D4_R3 | A.lumbricoides | 38.900 | | |
| 175 | StdExt1_6_Al_D4_R4 | A.lumbricoides | 34.765 | | |
| 177 | StdExt1_6_Al_D4_R5 | A.lumbricoides | 38.360 | | |
| 179 | StdExt1_6_Al_D4_R6 | A.lumbricoides | 36.389 | | |
| 181 | StdExt1_6_Al_D4_R7 | A.lumbricoides | 37.786 | | |
| 183 | StdExt1_6_Al_D4_R8 | A.lumbricoides | 33.883 | | |
| 185 | StdExt1_6_Al_D4_R9 | A.lumbricoides | 35.750 | | |
| 25 | StdExt1_6_Al_D5_R1 | A.lumbricoides | Undetermined | | |
| 26 | StdExt1_6_Al_D5_R11 | A.lumbricoides | 37.122 | | |
| 27 | StdExt1_6_Al_D5_R2 | A.lumbricoides | 34.363 | | |
| 28 | StdExt1_6_Al_D5_R12 | A.lumbricoides | 35.149 | | |
| 29 | StdExt1_6_Al_D5_R3 | A.lumbricoides | 35.674 | | |
| 30 | StdExt1_6_Al_D5_R13 | A.lumbricoides | 34.477 | | |
| 31 | StdExt1_6_Al_D5_R4 | A.lumbricoides | 35.099 | | |
| 32 | StdExt1_6_Al_D5_R14 | A.lumbricoides | 34.094 | | |
| 33 | StdExt1_6_Al_D5_R5 | A.lumbricoides | 35.277 | | |
| 34 | StdExt1_6_Al_D5_R15 | A.lumbricoides | 35.843 | 6.25E-05 | 5th Dilution |
| 35 | StdExt1_6_Al_D5_R6 | A.lumbricoides | 35.794 | | |
| 36 | StdExt1_6_Al_D5_R16 | A.lumbricoides | 37.658 | | |
| 37 | StdExt1_6_Al_D5_R7 | A.lumbricoides | Undetermined | | |
| 38 | StdExt1_6_Al_D5_R17 | A.lumbricoides | 34.468 | | |
| 39 | StdExt1_6_Al_D5_R8 | A.lumbricoides | 35.227 | | |
| 40 | StdExt1_6_Al_D5_R18 | A.lumbricoides | Undetermined | | |
| 41 | StdExt1_6_Al_D5_R9 | A.lumbricoides | 36.832 | | |
| 42 | StdExt1_6_Al_D5_R19 | A.lumbricoides | 34.332 | | |
| 43 | StdExt1_6_Al_D5_R10 | A.lumbricoides | 36.050 | | |
| 44 | StdExt1_6_Al_D5_R20 | A.lumbricoides | 35.009 | | |
| 47 | StdExt1_6_Al_D6_R1 | A.lumbricoides | Undetermined | | |
| 48 | StdExt1_6_Al_D6_R11 | A.lumbricoides | Undetermined | | |
| 73 | StdExt1_6_Al_D6_R2 | A.lumbricoides | Undetermined | | |
| 74 | StdExt1_6_Al_D6_R12 | A.lumbricoides | Undetermined | | |
| 75 | StdExt1_6_Al_D6_R3 | A.lumbricoides | Undetermined | | |
| 76 | StdExt1_6_Al_D6_R13 | A.lumbricoides | Undetermined | | |
| 77 | StdExt1_6_Al_D6_R4 | A.lumbricoides | Undetermined | | |
| 78 | StdExt1_6_Al_D6_R14 | A.lumbricoides | 35.311 | | |
| 79 | StdExt1_6_Al_D6_R5 | A.lumbricoides | Undetermined | | |
| 80 | StdExt1_6_Al_D6_R15 | A.lumbricoides | 37.398 | 3.13E-05 | 6th Dilution |
| 81 | StdExt1_6_Al_D6_R6 | A.lumbricoides | 21.451 | | |
| 82 | StdExt1_6_Al_D6_R16 | A.lumbricoides | Undetermined | | |
| 83 | StdExt1_6_Al_D6_R7 | A.lumbricoides | Undetermined | | |
| 84 | StdExt1_6_Al_D6_R17 | A.lumbricoides | Undetermined | | |
| 85 | StdExt1_6_Al_D6_R8 | A.lumbricoides | Undetermined | | |
| 86 | StdExt1_6_Al_D6_R18 | A.lumbricoides | Undetermined | | |
| 87 | StdExt1_6_Al_D6_R9 | A.lumbricoides | Undetermined | | |
| 88 | StdExt1_6_Al_D6_R19 | A.lumbricoides | Undetermined | | |
| 89 | StdExt1_6_Al_D6_R10 | A.lumbricoides | 35.910 | | |
| 90 | StdExt1_6_Al_D6_R20 | A.lumbricoides | Undetermined | | |
| 91 | StdExt1_6_Al_D7_R1 | A.lumbricoides | Undetermined | | |
| 92 | StdExt1_6_Al_D7_R11 | A.lumbricoides | Undetermined | | |
| 121 | StdExt1_6_Al_D7_R2 | A.lumbricoides | Undetermined | | |
| 122 | StdExt1_6_Al_D7_R12 | A.lumbricoides | Undetermined | | |
| 123 | StdExt1_6_Al_D7_R3 | A.lumbricoides | Undetermined | | |
| 124 | StdExt1_6_Al_D7_R13 | A.lumbricoides | Undetermined | | |
| 125 | StdExt1_6_Al_D7_R4 | A.lumbricoides | 35.615 | | |
| 126 | StdExt1_6_Al_D7_R14 | A.lumbricoides | Undetermined | | |
| 127 | StdExt1_6_Al_D7_R5 | A.lumbricoides | Undetermined | | |
| 128 | StdExt1_6_Al_D7_R15 | A.lumbricoides | Undetermined | 1.56E-05 | 7th Dilution |
| 129 | StdExt1_6_Al_D7_R6 | A.lumbricoides | Undetermined | | |
| 130 | StdExt1_6_Al_D7_R16 | A.lumbricoides | Undetermined | | |
| 131 | StdExt1_6_Al_D7_R7 | A.lumbricoides | Undetermined | | |
| 132 | StdExt1_6_Al_D7_R17 | A.lumbricoides | Undetermined | | |
| 133 | StdExt1_6_Al_D7_R8 | A.lumbricoides | 37.860 | | XX |
| 134 | StdExt1_6_Al_D7_R18 | A.lumbricoides | Undetermined | | |
| 135 | StdExt1_6_Al_D7_R9 | A.lumbricoides | Undetermined | | |
| 136 | StdExt1_6_Al_D7_R19 | A.lumbricoides | 38.819 | | |
| 137 | StdExt1_6_Al_D7_R10 | A.lumbricoides | Undetermined | | |
| 138 | StdExt1_6_Al_D7_R20 | A.lumbricoides | 32.877 | | |

**Table A5. 6**: *1:10 Serial Dilution Ct Scores for A. duodenale. Note there are 10 replicates of each dilution point.*

| First Dilution (1:10) | | | | | |
|---|---|---|---|---|---|
| Well | Sample Name | Target Name | CT | | |
| 11 | StdExt1_208_Ad_0.1_1 | A. duodenale | 32.320 | | |
| 20 | StdExt1_208_Ad_0.1_10 | A. duodenale | 32.429 | | |
| 12 | StdExt1_208_Ad_0.1_2 | A. duodenale | 31.835 | | |
| 13 | StdExt1_208_Ad_0.1_3 | A. duodenale | 32.014 | | |
| 14 | StdExt1_208_Ad_0.1_4 | A. duodenale | 31.738 | | |
| 15 | StdExt1_208_Ad_0.1_5 | A. duodenale | 31.590 | | |
| 16 | StdExt1_208_Ad_0.1_6 | A. duodenale | 31.858 | | |
| 17 | StdExt1_208_Ad_0.1_7 | A. duodenale | 31.996 | | |
| 18 | StdExt1_208_Ad_0.1_8 | A. duodenale | 32.508 | | |
| 19 | StdExt1_208_Ad_0.1_9 | A. duodenale | 31.875 | 1.00E-01 | 1st Dilution |
| 83 | StdExt1_208_Ad_0.01_1 | A. duodenale | 35.675 | | |
| 92 | StdExt1_208_Ad_0.01_10 | A. duodenale | 35.728 | | |
| 84 | StdExt1_208_Ad_0.01_2 | A. duodenale | 34.523 | | |
| 85 | StdExt1_208_Ad_0.01_3 | A. duodenale | 36.200 | | |
| 86 | StdExt1_208_Ad_0.01_4 | A. duodenale | 37.116 | | |
| 87 | StdExt1_208_Ad_0.01_5 | A. duodenale | 37.102 | | |
| 88 | StdExt1_208_Ad_0.01_6 | A. duodenale | 34.971 | | |
| 89 | StdExt1_208_Ad_0.01_7 | A. duodenale | 35.025 | | |
| 90 | StdExt1_208_Ad_0.01_8 | A. duodenale | 36.862 | | |
| 91 | StdExt1_208_Ad_0.01_9 | A. duodenale | Undetermined | 1.00E-02 | 2nd Dilution |
| 131 | StdExt1_208_Ad_0.001_1 | A. duodenale | 39.028 | | |
| 140 | StdExt1_208_Ad_0.001_10 | A. duodenale | Undetermined | | |
| 132 | StdExt1_208_Ad_0.001_2 | A. duodenale | Undetermined | | |
| 133 | StdExt1_208_Ad_0.001_3 | A. duodenale | Undetermined | | |
| 134 | StdExt1_208_Ad_0.001_4 | A. duodenale | Undetermined | | |
| 135 | StdExt1_208_Ad_0.001_5 | A. duodenale | Undetermined | | |
| 136 | StdExt1_208_Ad_0.001_6 | A. duodenale | Undetermined | | |
| 137 | StdExt1_208_Ad_0.001_7 | A. duodenale | Undetermined | | |
| 138 | StdExt1_208_Ad_0.001_8 | A. duodenale | Undetermined | | |
| 139 | StdExt1_208_Ad_0.001_9 | A. duodenale | Undetermined | 1.00E-03 | 3rd Dilution |
| 179 | StdExt1_208_Ad_0.0001_1 | A. duodenale | Undetermined | | |
| 188 | StdExt1_208_Ad_0.0001_10 | A. duodenale | Undetermined | | |
| 180 | StdExt1_208_Ad_0.0001_2 | A. duodenale | Undetermined | | |
| 181 | StdExt1_208_Ad_0.0001_3 | A. duodenale | Undetermined | | |
| 182 | StdExt1_208_Ad_0.0001_4 | A. duodenale | Undetermined | | |
| 183 | StdExt1_208_Ad_0.0001_5 | A. duodenale | Undetermined | | |
| 184 | StdExt1_208_Ad_0.0001_6 | A. duodenale | Undetermined | | |
| 185 | StdExt1_208_Ad_0.0001_7 | A. duodenale | Undetermined | | |
| 186 | StdExt1_208_Ad_0.0001_8 | A. duodenale | Undetermined | | |
| 187 | StdExt1_208_Ad_0.0001_9 | A. duodenale | Undetermined | 1.00E-04 | 4th Dilution |
| 227 | StdExt1_208_Ad_0.00001_1 | A. duodenale | Undetermined | | |
| 236 | StdExt1_208_Ad_0.00001_10 | A. duodenale | Undetermined | | |
| 228 | StdExt1_208_Ad_0.00001_2 | A. duodenale | Undetermined | | |
| 229 | StdExt1_208_Ad_0.00001_3 | A. duodenale | Undetermined | | |
| 230 | StdExt1_208_Ad_0.00001_4 | A. duodenale | Undetermined | | |
| 231 | StdExt1_208_Ad_0.00001_5 | A. duodenale | Undetermined | | |
| 232 | StdExt1_208_Ad_0.00001_6 | A. duodenale | Undetermined | | |
| 233 | StdExt1_208_Ad_0.00001_7 | A. duodenale | Undetermined | | |
| 234 | StdExt1_208_Ad_0.00001_8 | A. duodenale | Undetermined | | |
| 235 | StdExt1_208_Ad_0.00001_9 | A. duodenale | Undetermined | 1.00E-05 | 5th Dilution |
| 275 | StdExt1_208_Ad_0.000001_1 | A. duodenale | Undetermined | | |
| 284 | StdExt1_208_Ad_0.000001_10 | A. duodenale | Undetermined | | |
| 276 | StdExt1_208_Ad_0.000001_2 | A. duodenale | Undetermined | | |
| 277 | StdExt1_208_Ad_0.000001_3 | A. duodenale | Undetermined | | |
| 278 | StdExt1_208_Ad_0.000001_4 | A. duodenale | Undetermined | | |
| 279 | StdExt1_208_Ad_0.000001_5 | A. duodenale | Undetermined | | |
| 280 | StdExt1_208_Ad_0.000001_6 | A. duodenale | Undetermined | | |
| 281 | StdExt1_208_Ad_0.000001_7 | A. duodenale | Undetermined | | |
| 282 | StdExt1_208_Ad_0.000001_8 | A. duodenale | Undetermined | | |
| 283 | StdExt1_208_Ad_0.000001_9 | A. duodenale | Undetermined | 1.00E-06 | 6th Dilution |

**Table A5. 7**: *1:2 Serial Dilution Ct Scores for A. duodenale. Note there are 20 replicates of each dilution point.*

| Second Dilution (1:2) | | | | | |
|---|---|---|---|---|---|
| Well | Sample Name | Target Name | CT | | |
| 47 | NTC | A. duodenale | Undetermined | | |
| 48 | Pos. PCR | A. duodenale | 27.999 | | |
| 217 | StdExt1_208_Ad_D1_R1 | A. duodenale | 30.835 | 1.00E-01 | 1st Dilution |
| 235 | StdExt1_208_Ad_D1_R10 | A. duodenale | 31.735 | | |
| 218 | StdExt1_208_Ad_D1_R11 | A. duodenale | 31.079 | | |
| 220 | StdExt1_208_Ad_D1_R12 | A. duodenale | 30.665 | | |
| 222 | StdExt1_208_Ad_D1_R13 | A. duodenale | 31.353 | | |
| 224 | StdExt1_208_Ad_D1_R14 | A. duodenale | 30.747 | | |
| 226 | StdExt1_208_Ad_D1_R15 | A. duodenale | 31.399 | | |
| 228 | StdExt1_208_Ad_D1_R16 | A. duodenale | 31.000 | | |
| 230 | StdExt1_208_Ad_D1_R17 | A. duodenale | 30.721 | | |
| 232 | StdExt1_208_Ad_D1_R18 | A. duodenale | 30.628 | | |
| 234 | StdExt1_208_Ad_D1_R19 | A. duodenale | 31.446 | | |
| 219 | StdExt1_208_Ad_D1_R2 | A. duodenale | 30.857 | | |
| 236 | StdExt1_208_Ad_D1_R20 | A. duodenale | 31.319 | | |
| 221 | StdExt1_208_Ad_D1_R3 | A. duodenale | 30.966 | | |
| 223 | StdExt1_208_Ad_D1_R4 | A. duodenale | 30.512 | | |
| 225 | StdExt1_208_Ad_D1_R5 | A. duodenale | 31.231 | | |
| 227 | StdExt1_208_Ad_D1_R6 | A. duodenale | 31.110 | | |
| 229 | StdExt1_208_Ad_D1_R7 | A. duodenale | 31.095 | | |
| 231 | StdExt1_208_Ad_D1_R8 | A. duodenale | 30.912 | | |
| 233 | StdExt1_208_Ad_D1_R9 | A. duodenale | 31.270 | | |
| 265 | StdExt1_208_Ad_D2_R1 | A. duodenale | 31.749 | 5.00E-02 | 2nd Dilution |
| 283 | StdExt1_208_Ad_D2_R10 | A. duodenale | 31.977 | | |
| 266 | StdExt1_208_Ad_D2_R11 | A. duodenale | 31.307 | | |
| 268 | StdExt1_208_Ad_D2_R12 | A. duodenale | 31.647 | | |
| 270 | StdExt1_208_Ad_D2_R13 | A. duodenale | 31.925 | | |
| 272 | StdExt1_208_Ad_D2_R14 | A. duodenale | 31.940 | | |
| 274 | StdExt1_208_Ad_D2_R15 | A. duodenale | 32.198 | | |
| 276 | StdExt1_208_Ad_D2_R16 | A. duodenale | 33.388 | | |
| 278 | StdExt1_208_Ad_D2_R17 | A. duodenale | 31.587 | | |
| 280 | StdExt1_208_Ad_D2_R18 | A. duodenale | 33.051 | | |
| 282 | StdExt1_208_Ad_D2_R19 | A. duodenale | 31.971 | | |
| 267 | StdExt1_208_Ad_D2_R2 | A. duodenale | 32.638 | | |
| 284 | StdExt1_208_Ad_D2_R20 | A. duodenale | 31.540 | | |
| 269 | StdExt1_208_Ad_D2_R3 | A. duodenale | 32.289 | | |
| 271 | StdExt1_208_Ad_D2_R4 | A. duodenale | 31.475 | | |
| 273 | StdExt1_208_Ad_D2_R5 | A. duodenale | 32.493 | | |
| 275 | StdExt1_208_Ad_D2_R6 | A. duodenale | 32.496 | | |
| 277 | StdExt1_208_Ad_D2_R7 | A. duodenale | 31.644 | | |
| 279 | StdExt1_208_Ad_D2_R8 | A. duodenale | 31.664 | | |
| 281 | StdExt1_208_Ad_D2_R9 | A. duodenale | 32.665 | | |
| 313 | StdExt1_208_Ad_D3_R1 | A. duodenale | 33.788 | 2.50E-02 | 3rd Dilution |
| 331 | StdExt1_208_Ad_D3_R10 | A. duodenale | 34.538 | | |
| 314 | StdExt1_208_Ad_D3_R11 | A. duodenale | 32.405 | | |
| 316 | StdExt1_208_Ad_D3_R12 | A. duodenale | 32.440 | | |
| 318 | StdExt1_208_Ad_D3_R13 | A. duodenale | 32.586 | | |
| 320 | StdExt1_208_Ad_D3_R14 | A. duodenale | 33.110 | | |
| 322 | StdExt1_208_Ad_D3_R15 | A. duodenale | 33.641 | | |
| 324 | StdExt1_208_Ad_D3_R16 | A. duodenale | 32.904 | | |
| 326 | StdExt1_208_Ad_D3_R17 | A. duodenale | 34.067 | | |
| 328 | StdExt1_208_Ad_D3_R18 | A. duodenale | 34.057 | | |
| 330 | StdExt1_208_Ad_D3_R19 | A. duodenale | 32.790 | | |
| 315 | StdExt1_208_Ad_D3_R2 | A. duodenale | 34.918 | | |
| 332 | StdExt1_208_Ad_D3_R20 | A. duodenale | 33.329 | | |
| 317 | StdExt1_208_Ad_D3_R3 | A. duodenale | 32.474 | | |
| 319 | StdExt1_208_Ad_D3_R4 | A. duodenale | 32.488 | | |
| 321 | StdExt1_208_Ad_D3_R5 | A. duodenale | 33.430 | | |
| 323 | StdExt1_208_Ad_D3_R6 | A. duodenale | 32.920 | | |
| 325 | StdExt1_208_Ad_D3_R7 | A. duodenale | 32.355 | | |
| 327 | StdExt1_208_Ad_D3_R8 | A. duodenale | 32.292 | | |
| 329 | StdExt1_208_Ad_D3_R9 | A. duodenale | 32.236 | | |

| Second Dilution (1:2) | | | | | |
|---|---|---|---|---|---|
| Well | Sample Name | Target Name | CT | | |
| 361 | StdExt1_208_Ad_D4_R1 | A. duodenale | 33.459 | 1.25E-02 | 4th Dilution |
| 379 | StdExt1_208_Ad_D4_R10 | A. duodenale | 33.383 | | |
| 362 | StdExt1_208_Ad_D4_R11 | A. duodenale | 34.610 | | |
| 364 | StdExt1_208_Ad_D4_R12 | A. duodenale | 36.116 | | |
| 366 | StdExt1_208_Ad_D4_R13 | A. duodenale | 34.729 | | |
| 368 | StdExt1_208_Ad_D4_R14 | A. duodenale | 33.443 | | |
| 370 | StdExt1_208_Ad_D4_R15 | A. duodenale | 34.368 | | |
| 372 | StdExt1_208_Ad_D4_R16 | A. duodenale | 35.491 | | |
| 374 | StdExt1_208_Ad_D4_R17 | A. duodenale | 33.878 | | |
| 376 | StdExt1_208_Ad_D4_R18 | A. duodenale | 34.646 | | |
| 378 | StdExt1_208_Ad_D4_R19 | A. duodenale | 34.343 | | |
| 363 | StdExt1_208_Ad_D4_R2 | A. duodenale | 34.167 | | |
| 380 | StdExt1_208_Ad_D4_R20 | A. duodenale | 34.457 | | |
| 365 | StdExt1_208_Ad_D4_R3 | A. duodenale | 34.042 | | |
| 367 | StdExt1_208_Ad_D4_R4 | A. duodenale | 34.761 | | |
| 369 | StdExt1_208_Ad_D4_R5 | A. duodenale | 33.673 | | |
| 371 | StdExt1_208_Ad_D4_R6 | A. duodenale | 33.608 | | |
| 373 | StdExt1_208_Ad_D4_R7 | A. duodenale | 33.122 | | |
| 375 | StdExt1_208_Ad_D4_R8 | A. duodenale | 34.628 | | |
| 377 | StdExt1_208_Ad_D4_R9 | A. duodenale | 34.139 | | |
| 217 | StdExt1_208_Ad_D5_R1 | A. duodenale | Undetermined | 6.25E-03 | 5th Dilution |
| 218 | StdExt1_208_Ad_D5_R11 | A. duodenale | Undetermined | | |
| 219 | StdExt1_208_Ad_D5_R2 | A. duodenale | 37.405 | | |
| 220 | StdExt1_208_Ad_D5_R12 | A. duodenale | Undetermined | | |
| 221 | StdExt1_208_Ad_D5_R3 | A. duodenale | 36.288 | | |
| 222 | StdExt1_208_Ad_D5_R13 | A. duodenale | Undetermined | | |
| 223 | StdExt1_208_Ad_D5_R4 | A. duodenale | 35.569 | | |
| 224 | StdExt1_208_Ad_D5_R14 | A. duodenale | 35.962 | | |
| 225 | StdExt1_208_Ad_D5_R5 | A. duodenale | 34.733 | | |
| 226 | StdExt1_208_Ad_D5_R15 | A. duodenale | 35.696 | | |
| 227 | StdExt1_208_Ad_D5_R6 | A. duodenale | 35.196 | | |
| 228 | StdExt1_208_Ad_D5_R16 | A. duodenale | 36.114 | | |
| 229 | StdExt1_208_Ad_D5_R7 | A. duodenale | 35.788 | | |
| 230 | StdExt1_208_Ad_D5_R17 | A. duodenale | 36.334 | | |
| 231 | StdExt1_208_Ad_D5_R8 | A. duodenale | 39.662 | | |
| 232 | StdExt1_208_Ad_D5_R18 | A. duodenale | 36.404 | | |
| 233 | StdExt1_208_Ad_D5_R9 | A. duodenale | Undetermined | | |
| 234 | StdExt1_208_Ad_D5_R19 | A. duodenale | 39.296 | | |
| 235 | StdExt1_208_Ad_D5_R10 | A. duodenale | 36.454 | | |
| 236 | StdExt1_208_Ad_D5_R20 | A. duodenale | 34.920 | | |
| 265 | StdExt1_208_Ad_D6_R1 | A. duodenale | 35.141 | 3.13E-03 | 6th Dilution |
| 266 | StdExt1_208_Ad_D6_R11 | A. duodenale | Undetermined | | |
| 267 | StdExt1_208_Ad_D6_R2 | A. duodenale | 36.431 | | |
| 268 | StdExt1_208_Ad_D6_R12 | A. duodenale | Undetermined | | |
| 269 | StdExt1_208_Ad_D6_R3 | A. duodenale | Undetermined | | |
| 270 | StdExt1_208_Ad_D6_R13 | A. duodenale | 35.366 | | |
| 271 | StdExt1_208_Ad_D6_R4 | A. duodenale | 38.609 | | |
| 272 | StdExt1_208_Ad_D6_R14 | A. duodenale | Undetermined | | |
| 273 | StdExt1_208_Ad_D6_R5 | A. duodenale | Undetermined | | |
| 274 | StdExt1_208_Ad_D6_R15 | A. duodenale | 35.641 | | |
| 275 | StdExt1_208_Ad_D6_R6 | A. duodenale | 38.139 | | |
| 276 | StdExt1_208_Ad_D6_R16 | A. duodenale | 34.706 | | |
| 277 | StdExt1_208_Ad_D6_R7 | A. duodenale | Undetermined | | |
| 278 | StdExt1_208_Ad_D6_R17 | A. duodenale | 39.343 | | |
| 279 | StdExt1_208_Ad_D6_R8 | A. duodenale | 38.058 | | |
| 280 | StdExt1_208_Ad_D6_R18 | A. duodenale | Undetermined | | |
| 281 | StdExt1_208_Ad_D6_R9 | A. duodenale | 36.533 | | |
| 282 | StdExt1_208_Ad_D6_R19 | A. duodenale | Undetermined | | |
| 283 | StdExt1_208_Ad_D6_R10 | A. duodenale | Undetermined | | |
| 284 | StdExt1_208_Ad_D6_R20 | A. duodenale | 36.423 | | |
| 313 | StdExt1_208_Ad_D7_R1 | A. duodenale | 39.310 | 1.56E-03 | 7th Dilution |
| 314 | StdExt1_208_Ad_D7_R11 | A. duodenale | 38.522 | | |
| 315 | StdExt1_208_Ad_D7_R2 | A. duodenale | Undetermined | | |
| 316 | StdExt1_208_Ad_D7_R12 | A. duodenale | Undetermined | | |
| 317 | StdExt1_208_Ad_D7_R3 | A. duodenale | 36.146 | | |
| 318 | StdExt1_208_Ad_D7_R13 | A. duodenale | Undetermined | | |
| 319 | StdExt1_208_Ad_D7_R4 | A. duodenale | 36.495 | | |
| 320 | StdExt1_208_Ad_D7_R14 | A. duodenale | Undetermined | | |
| 321 | StdExt1_208_Ad_D7_R5 | A. duodenale | 35.162 | | |
| 322 | StdExt1_208_Ad_D7_R15 | A. duodenale | Undetermined | | |
| 323 | StdExt1_208_Ad_D7_R6 | A. duodenale | Undetermined | | |
| 324 | StdExt1_208_Ad_D7_R16 | A. duodenale | Undetermined | | |
| 325 | StdExt1_208_Ad_D7_R7 | A. duodenale | 37.771 | | |
| 326 | StdExt1_208_Ad_D7_R17 | A. duodenale | Undetermined | | |
| 327 | StdExt1_208_Ad_D7_R8 | A. duodenale | Undetermined | | |
| 328 | StdExt1_208_Ad_D7_R18 | A. duodenale | Undetermined | | |
| 329 | StdExt1_208_Ad_D7_R9 | A. duodenale | 36.275 | | |
| 330 | StdExt1_208_Ad_D7_R19 | A. duodenale | 38.044 | | |
| 331 | StdExt1_208_Ad_D7_R10 | A. duodenale | Undetermined | | |
| 332 | StdExt1_208_Ad_D7_R20 | A. duodenale | 39.620 | | |

**Table A5. 8**: *1:10 Serial Dilution Ct Scores for T. trichiura. Note there are 10 replicates of each dilution point.*

| First Dilution (1:10) | | | | | |
|---|---|---|---|---|---|
| Well | Sample Name | Target Name | CT | | |
| A1 | StdExt2_44_Tt_D1_R1 | T.trichiura | 26.349 | | |
| A2 | StdExt2_44_Tt_D1_R2 | T.trichiura | 26.527 | | |
| A3 | StdExt2_44_Tt_D1_R3 | T.trichiura | 26.340 | | |
| A4 | StdExt2_44_Tt_D1_R4 | T.trichiura | 26.325 | | |
| A5 | StdExt2_44_Tt_D1_R5 | T.trichiura | 26.568 | | |
| A6 | StdExt2_44_Tt_D1_R6 | T.trichiura | 26.479 | | |
| A7 | StdExt2_44_Tt_D1_R7 | T.trichiura | 26.597 | | |
| A8 | StdExt2_44_Tt_D1_R8 | T.trichiura | 26.482 | | |
| A9 | StdExt2_44_Tt_D1_R9 | T.trichiura | 26.556 | | |
| A10 | StdExt2_44_Tt_D1_R10 | T.trichiura | 26.629 | 1.00E-01 | 1st Dilution |
| C1 | StdExt2_44_Tt_D2_R1 | T.trichiura | 29.823 | | |
| C2 | StdExt2_44_Tt_D2_R2 | T.trichiura | 29.700 | | |
| C3 | StdExt2_44_Tt_D2_R3 | T.trichiura | 29.641 | | |
| C4 | StdExt2_44_Tt_D2_R4 | T.trichiura | 29.797 | | |
| C5 | StdExt2_44_Tt_D2_R5 | T.trichiura | 29.688 | | |
| C6 | StdExt2_44_Tt_D2_R6 | T.trichiura | 29.944 | | |
| C7 | StdExt2_44_Tt_D2_R7 | T.trichiura | 29.749 | | |
| C8 | StdExt2_44_Tt_D2_R8 | T.trichiura | 29.873 | | |
| C9 | StdExt2_44_Tt_D2_R9 | T.trichiura | 29.981 | | |
| C10 | StdExt2_44_Tt_D2_R10 | T.trichiura | 30.079 | 1.00E-02 | 2nd Dilution |
| E1 | StdExt2_44_Tt_D3_R1 | T.trichiura | 33.091 | | |
| E2 | StdExt2_44_Tt_D3_R2 | T.trichiura | 32.781 | | |
| E3 | StdExt2_44_Tt_D3_R3 | T.trichiura | 33.021 | | |
| E4 | StdExt2_44_Tt_D3_R4 | T.trichiura | 33.369 | | |
| E5 | StdExt2_44_Tt_D3_R5 | T.trichiura | 32.837 | | |
| E6 | StdExt2_44_Tt_D3_R6 | T.trichiura | 33.038 | | |
| E7 | StdExt2_44_Tt_D3_R7 | T.trichiura | 32.483 | | |
| E8 | StdExt2_44_Tt_D3_R8 | T.trichiura | 33.102 | | |
| E9 | StdExt2_44_Tt_D3_R9 | T.trichiura | 32.490 | | |
| E10 | StdExt2_44_Tt_D3_R10 | T.trichiura | 33.286 | 1.00E-03 | 3rd Dilution |
| G1 | StdExt2_44_Tt_D4_R1 | T.trichiura | 36.585 | | |
| G2 | StdExt2_44_Tt_D4_R2 | T.trichiura | 35.552 | | |
| G3 | StdExt2_44_Tt_D4_R3 | T.trichiura | 36.66449 | | |
| G4 | StdExt2_44_Tt_D4_R4 | T.trichiura | 38.441 | | |
| G5 | StdExt2_44_Tt_D4_R5 | T.trichiura | 37.228886 | | |
| G6 | StdExt2_44_Tt_D4_R6 | T.trichiura | Undetermined | | |
| G7 | StdExt2_44_Tt_D4_R7 | T.trichiura | 35.310 | | |
| G8 | StdExt2_44_Tt_D4_R8 | T.trichiura | 37.101 | | |
| G9 | StdExt2_44_Tt_D4_R9 | T.trichiura | 37.229 | | |
| G10 | StdExt2_44_Tt_D4_R10 | T.trichiura | 37.217 | 1.00E-04 | 4th Dilution |
| I1 | StdExt2_44_Tt_D5_R1 | T.trichiura | Undetermined | | |
| I2 | StdExt2_44_Tt_D5_R2 | T.trichiura | Undetermined | | |
| I3 | StdExt2_44_Tt_D5_R3 | T.trichiura | Undetermined | | |
| I4 | StdExt2_44_Tt_D5_R4 | T.trichiura | Undetermined | | |
| I5 | StdExt2_44_Tt_D5_R5 | T.trichiura | Undetermined | | |
| I6 | StdExt2_44_Tt_D5_R6 | T.trichiura | Undetermined | | |
| I7 | StdExt2_44_Tt_D5_R7 | T.trichiura | Undetermined | | |
| I8 | StdExt2_44_Tt_D5_R8 | T.trichiura | Undetermined | | |
| I9 | StdExt2_44_Tt_D5_R9 | T.trichiura | Undetermined | | |
| I10 | StdExt2_44_Tt_D5_R10 | T.trichiura | Undetermined | 1.00E-05 | 5th Dilution |

**Table A5. 9**: *1:2 Serial Dilution Ct Scores for T. trichiura. Note there are 20 replicates of each dilution point.*

| Second Dilution (1:2) | | | | | |
|---|---|---|---|---|---|
| Well | Sample Name | Target Name | CT | | |
| A23 | NTC | T.trichiura | Undetermined | | |
| A24 | Pos. PCR | T.trichiura | 28.669 | | |
| A1 | StdExt2_44_Tt_D1_R1 | T.trichiura | 31.790 | 1.00E-03 | 1st Dilution |
| A2 | StdExt2_44_Tt_D1_R2 | T.trichiura | 32.764 | | |
| A3 | StdExt2_44_Tt_D1_R3 | T.trichiura | 32.411 | | |
| A4 | StdExt2_44_Tt_D1_R4 | T.trichiura | 32.039 | | |
| A5 | StdExt2_44_Tt_D1_R5 | T.trichiura | 32.055 | | |
| A6 | StdExt2_44_Tt_D1_R6 | T.trichiura | 32.404 | | |
| A7 | StdExt2_44_Tt_D1_R7 | T.trichiura | 32.443 | | |
| A8 | StdExt2_44_Tt_D1_R8 | T.trichiura | 32.753 | | |
| A9 | StdExt2_44_Tt_D1_R9 | T.trichiura | 32.029 | | |
| A10 | StdExt2_44_Tt_D1_R10 | T.trichiura | 32.185 | | |
| A11 | StdExt2_44_Tt_D1_R11 | T.trichiura | 33.364 | | |
| A12 | StdExt2_44_Tt_D1_R12 | T.trichiura | 32.505 | | |
| A13 | StdExt2_44_Tt_D1_R13 | T.trichiura | 32.119 | | |
| A14 | StdExt2_44_Tt_D1_R14 | T.trichiura | 31.844 | | |
| A15 | StdExt2_44_Tt_D1_R15 | T.trichiura | 32.863 | | |
| A16 | StdExt2_44_Tt_D1_R16 | T.trichiura | 32.717 | | |
| A17 | StdExt2_44_Tt_D1_R17 | T.trichiura | 32.586 | | |
| A18 | StdExt2_44_Tt_D1_R18 | T.trichiura | 32.012 | | |
| A19 | StdExt2_44_Tt_D1_R19 | T.trichiura | 32.257 | | |
| A20 | StdExt2_44_Tt_D1_R20 | T.trichiura | 32.331 | | |
| C1 | StdExt2_44_Tt_D2_R1 | T.trichiura | 33.484 | 5.00E-04 | 2nd Dilution |
| C2 | StdExt2_44_Tt_D2_R2 | T.trichiura | 34.339 | | |
| C3 | StdExt2_44_Tt_D2_R3 | T.trichiura | 32.372 | | |
| C4 | StdExt2_44_Tt_D2_R4 | T.trichiura | Well Failure | | |
| C5 | StdExt2_44_Tt_D2_R5 | T.trichiura | 33.952 | | |
| C6 | StdExt2_44_Tt_D2_R6 | T.trichiura | 33.536 | | |
| C7 | StdExt2_44_Tt_D2_R7 | T.trichiura | 33.064 | | |
| C8 | StdExt2_44_Tt_D2_R8 | T.trichiura | 35.002 | | |
| C9 | StdExt2_44_Tt_D2_R9 | T.trichiura | 33.176 | | |
| C10 | StdExt2_44_Tt_D2_R10 | T.trichiura | 34.306 | | |
| C11 | StdExt2_44_Tt_D2_R11 | T.trichiura | 33.744 | | |
| C12 | StdExt2_44_Tt_D2_R12 | T.trichiura | 35.296 | | |
| C13 | StdExt2_44_Tt_D2_R13 | T.trichiura | 33.319 | | |
| C14 | StdExt2_44_Tt_D2_R14 | T.trichiura | 33.081 | | |
| C15 | StdExt2_44_Tt_D2_R15 | T.trichiura | 33.607 | | |
| C16 | StdExt2_44_Tt_D2_R16 | T.trichiura | 34.105 | | |
| C17 | StdExt2_44_Tt_D2_R17 | T.trichiura | 33.260 | | |
| C18 | StdExt2_44_Tt_D2_R18 | T.trichiura | 35.116 | | |
| C19 | StdExt2_44_Tt_D2_R19 | T.trichiura | 33.547 | | |
| C20 | StdExt2_44_Tt_D2_R20 | T.trichiura | 33.956 | | |
| E1 | StdExt2_44_Tt_D3_R1 | T.trichiura | 34.817 | 2.50E-04 | 3rd Dilution |
| E2 | StdExt2_44_Tt_D3_R2 | T.trichiura | 35.495 | | |
| E3 | StdExt2_44_Tt_D3_R3 | T.trichiura | 34.640 | | |
| E4 | StdExt2_44_Tt_D3_R4 | T.trichiura | 34.201 | | |
| E5 | StdExt2_44_Tt_D3_R5 | T.trichiura | 34.845 | | |
| E6 | StdExt2_44_Tt_D3_R6 | T.trichiura | 35.112 | | |
| E7 | StdExt2_44_Tt_D3_R7 | T.trichiura | 34.524 | | |
| E8 | StdExt2_44_Tt_D3_R8 | T.trichiura | 33.390 | | |
| E9 | StdExt2_44_Tt_D3_R9 | T.trichiura | 33.927 | | |
| E10 | StdExt2_44_Tt_D3_R10 | T.trichiura | 34.316 | | |
| E11 | StdExt2_44_Tt_D3_R11 | T.trichiura | 34.227 | | |
| E12 | StdExt2_44_Tt_D3_R12 | T.trichiura | 34.327 | | |
| E13 | StdExt2_44_Tt_D3_R13 | T.trichiura | 32.972 | | |
| E14 | StdExt2_44_Tt_D3_R14 | T.trichiura | 34.413 | | |
| E15 | StdExt2_44_Tt_D3_R15 | T.trichiura | 33.647 | | |
| E16 | StdExt2_44_Tt_D3_R16 | T.trichiura | 34.444 | | |
| E17 | StdExt2_44_Tt_D3_R17 | T.trichiura | 33.599 | | |
| E18 | StdExt2_44_Tt_D3_R18 | T.trichiura | 35.071 | | |
| E19 | StdExt2_44_Tt_D3_R19 | T.trichiura | 34.542 | | |
| E20 | StdExt2_44_Tt_D3_R20 | T.trichiura | 34.128 | | |

| Second Dilution (1:2) | | | | | |
|---|---|---|---|---|---|
| Well | Sample Name | Target Name | CT | | |
| G1 | StdExt2_44_Tt_D4_R1 | T.trichiura | 34.668 | 1.25E-04 | 4th Dilution |
| G2 | StdExt2_44_Tt_D4_R2 | T.trichiura | 37.239 | | |
| G3 | StdExt2_44_Tt_D4_R3 | T.trichiura | 35.905 | | |
| G4 | StdExt2_44_Tt_D4_R4 | T.trichiura | 34.152 | | |
| G5 | StdExt2_44_Tt_D4_R5 | T.trichiura | 36.237 | | |
| G6 | StdExt2_44_Tt_D4_R6 | T.trichiura | 35.862 | | |
| G7 | StdExt2_44_Tt_D4_R7 | T.trichiura | 35.479 | | |
| G8 | StdExt2_44_Tt_D4_R8 | T.trichiura | Undetermined | | |
| G9 | StdExt2_44_Tt_D4_R9 | T.trichiura | 38.417 | | |
| G10 | StdExt2_44_Tt_D4_R10 | T.trichiura | 34.398 | | |
| G11 | StdExt2_44_Tt_D4_R11 | T.trichiura | 37.293 | | |
| G12 | StdExt2_44_Tt_D4_R12 | T.trichiura | 35.716 | | |
| G13 | StdExt2_44_Tt_D4_R13 | T.trichiura | 35.967 | | |
| G14 | StdExt2_44_Tt_D4_R14 | T.trichiura | 37.410 | | |
| G15 | StdExt2_44_Tt_D4_R15 | T.trichiura | 36.004 | | |
| G16 | StdExt2_44_Tt_D4_R16 | T.trichiura | 37.509 | | |
| G17 | StdExt2_44_Tt_D4_R17 | T.trichiura | 35.416 | | |
| G18 | StdExt2_44_Tt_D4_R18 | T.trichiura | 37.199 | | |
| G19 | StdExt2_44_Tt_D4_R19 | T.trichiura | 36.049 | | |
| G20 | StdExt2_44_Tt_D4_R20 | T.trichiura | 36.510 | | |
| I1 | StdExt2_44_Tt_D5_R1 | T.trichiura | Undetermined | 6.25E-05 | 5th Dilution |
| I2 | StdExt2_44_Tt_D5_R2 | T.trichiura | 35.935963 | | |
| I3 | StdExt2_44_Tt_D5_R3 | T.trichiura | Undetermined | | |
| I4 | StdExt2_44_Tt_D5_R4 | T.trichiura | 36.615772 | | |
| I5 | StdExt2_44_Tt_D5_R5 | T.trichiura | 37.701 | | |
| I6 | StdExt2_44_Tt_D5_R6 | T.trichiura | 34.70892 | | |
| I7 | StdExt2_44_Tt_D5_R7 | T.trichiura | 36.367 | | |
| I8 | StdExt2_44_Tt_D5_R8 | T.trichiura | 36.382 | | |
| I9 | StdExt2_44_Tt_D5_R9 | T.trichiura | 37.301 | | |
| I10 | StdExt2_44_Tt_D5_R10 | T.trichiura | Undetermined | | |
| I11 | StdExt2_44_Tt_D5_R11 | T.trichiura | 37.311 | | |
| I12 | StdExt2_44_Tt_D5_R12 | T.trichiura | 35.766 | | |
| I13 | StdExt2_44_Tt_D5_R13 | T.trichiura | 35.665 | | |
| I14 | StdExt2_44_Tt_D5_R14 | T.trichiura | 34.397 | | |
| I15 | StdExt2_44_Tt_D5_R15 | T.trichiura | 37.209 | | |
| I16 | StdExt2_44_Tt_D5_R16 | T.trichiura | 35.798 | | |
| I17 | StdExt2_44_Tt_D5_R17 | T.trichiura | 37.20936 | | |
| I18 | StdExt2_44_Tt_D5_R18 | T.trichiura | Undetermined | | |
| I19 | StdExt2_44_Tt_D5_R19 | T.trichiura | 34.081 | | |
| I20 | StdExt2_44_Tt_D5_R20 | T.trichiura | 36.526 | | |
| K1 | StdExt2_44_Tt_D6_R1 | T.trichiura | 35.507 | 3.13E-05 | 6th Dilution |
| K2 | StdExt2_44_Tt_D6_R2 | T.trichiura | Undetermined | | |
| K3 | StdExt2_44_Tt_D6_R3 | T.trichiura | Undetermined | | |
| K4 | StdExt2_44_Tt_D6_R4 | T.trichiura | 34.44711 | | |
| K5 | StdExt2_44_Tt_D6_R5 | T.trichiura | 37.265686 | | |
| K6 | StdExt2_44_Tt_D6_R6 | T.trichiura | 36.959 | | |
| K7 | StdExt2_44_Tt_D6_R7 | T.trichiura | Undetermined | | |
| K8 | StdExt2_44_Tt_D6_R8 | T.trichiura | 36.441128 | | |
| K9 | StdExt2_44_Tt_D6_R9 | T.trichiura | Undetermined | | |
| K10 | StdExt2_44_Tt_D6_R10 | T.trichiura | 37.383 | | |
| K11 | StdExt2_44_Tt_D6_R11 | T.trichiura | 37.075 | | |
| K12 | StdExt2_44_Tt_D6_R12 | T.trichiura | Undetermined | | |
| K13 | StdExt2_44_Tt_D6_R13 | T.trichiura | 37.114407 | | |
| K14 | StdExt2_44_Tt_D6_R14 | T.trichiura | Undetermined | | |
| K15 | StdExt2_44_Tt_D6_R15 | T.trichiura | Undetermined | | |
| K16 | StdExt2_44_Tt_D6_R16 | T.trichiura | Undetermined | | |
| K17 | StdExt2_44_Tt_D6_R17 | T.trichiura | Undetermined | | |
| K18 | StdExt2_44_Tt_D6_R18 | T.trichiura | 37.123684 | | |
| K19 | StdExt2_44_Tt_D6_R19 | T.trichiura | 37.234425 | | |
| K20 | StdExt2_44_Tt_D6_R20 | T.trichiura | Undetermined | | |
| M1 | StdExt2_44_Tt_D7_R1 | T.trichiura | Undetermined | 1.56E-05 | 7th Dilution |
| M2 | StdExt2_44_Tt_D7_R2 | T.trichiura | 36.758 | | |
| M3 | StdExt2_44_Tt_D7_R3 | T.trichiura | Undetermined | | |
| M4 | StdExt2_44_Tt_D7_R4 | T.trichiura | 37.67536 | | |
| M5 | StdExt2_44_Tt_D7_R5 | T.trichiura | 37.371 | | |
| M6 | StdExt2_44_Tt_D7_R6 | T.trichiura | Undetermined | | |
| M7 | StdExt2_44_Tt_D7_R7 | T.trichiura | 36.840 | | |
| M8 | StdExt2_44_Tt_D7_R8 | T.trichiura | Undetermined | | |
| M9 | StdExt2_44_Tt_D7_R9 | T.trichiura | Undetermined | | |
| M10 | StdExt2_44_Tt_D7_R10 | T.trichiura | 39.263245 | | |
| M11 | StdExt2_44_Tt_D7_R11 | T.trichiura | Undetermined | | |
| M12 | StdExt2_44_Tt_D7_R12 | T.trichiura | Undetermined | | |
| M13 | StdExt2_44_Tt_D7_R13 | T.trichiura | Undetermined | | |
| M14 | StdExt2_44_Tt_D7_R14 | T.trichiura | Undetermined | | |
| M15 | StdExt2_44_Tt_D7_R15 | T.trichiura | Undetermined | | |
| M16 | StdExt2_44_Tt_D7_R16 | T.trichiura | Undetermined | | |
| M17 | StdExt2_44_Tt_D7_R17 | T.trichiura | Undetermined | | |
| M18 | StdExt2_44_Tt_D7_R18 | T.trichiura | Undetermined | | |
| M19 | StdExt2_44_Tt_D7_R19 | T.trichiura | 35.19326 | | |
| M20 | StdExt2_44_Tt_D7_R20 | T.trichiura | 38.251 | | |

#### A6: Raw Data: Multi-site Characterization of Spiked Stool Standards (II.B.iii)

**Table A6. 1:** Initial Validation Panel, Smith vs. Quantigen Aliquot-Level **T. trichiura Assay.** Note that Smith College Ct Scores provide the standard deviation in parentheses.

| | | T. trichiura Ct (Mean- 4 Replicates) | | | | | |
|---|---|---|---|---|---|---|---|
| Smith Panel ID | Sample Summary Status | Smith College | | | Quantigen | | |
| | | Aliquot A | Aliquot B | Aliquot C | Aliquot Q1 | Aliquot Q2 | Aliquot Q3 |
| 1 | Al, Na | | | | neg | neg | neg |
| 2 | Na | | | | neg | neg | neg |
| 3 | Tt | 32.15 (1.84) | 32.73 (2.36) | 33.23 (2.17) | 34.79 | 36.39 | 33.17 |
| 7 | Al | | | | neg | neg | neg |
| 8 | Tt, Al, Na | 30.71 (0.81) | 29.34(0.75) | 30.41(1.19) | 35.13 | 35.38 | 35.93 |
| 9 | Tt, Al, Na | 27.54 (0.59) | 28.08 (0.58) | 28.95 (0.69) | 32.95 | 31.56 | 34.13 |
| 11 | Tt, Na | 29.35 (0.73) | 30.28 (0.59) | 30.47 (0.86) | 35.15 | 34.10 | 34.28 |
| 12 | Na | | | | neg | neg | neg |
| 13 | Tt | 30.24 (0.86) | 30.29 (0.58) | 29.92 (0.36) | neg | neg | 35.53 |
| 14 | Al | | | | neg | neg | neg |
| 15 | Al, Na | | | | neg | neg | neg |
| 16 | Tt, Al | 29.50 (0.60) | 29.40 (0.59) | 32.88 (2.16) | 30.96 | neg | 34.94 |
| 17 | Na | | | | neg | neg | neg |
| 18 | Al | | | | neg | neg | neg |
| 19 | Tt, Al, Na | 31.42 (0.90) | 30.71 (0.41) | 31.41 (0.77) | 33.30 | 32.79 | 35.93 |
| 20 | Tt, Al, Na | 30.94 (0.33) | 32.33 (0.64) | 30.22 (0.62) | 35.21 | 36.18 | 35.68 |
| 4 v2 | Tt, Al | 32.65 (1.48) | 31.10 (0.84) | 32.23 (1.58) | 36.07 | neg | 36.52 |
| **10 v2** | Na, Ad | | | | neg | neg | neg |
| 27 | Al, Na, Ad | | | | neg | 36.76 | neg |
| 12 v2 | Na | | | | neg | neg | neg |
| 21 | Bioreclamation | | | | neg | neg | neg |
| 22 | Bioreclamation | | | | neg | neg | neg |
| 23 | Bioreclamation | | | | neg | neg | neg |
| 24 | Bioreclamation | | | | neg | neg | neg |
| 25 | Bioreclamation | | | | neg | neg | neg |

**Table A6.2:** *Initial Validation Panel, Smith vs. Quantigen Aliquot-Level* ***A. lumbricoides Assay.*** *Note that Smith College Ct Scores provide the standard deviation in parentheses.*

| | | A. lumbricoides Ct (Mean- 4 Replicates) | | | | | |
|---|---|---|---|---|---|---|---|
| Smith Panel ID | Sample Summary Status | Smith College | | | Quantigen | | |
| | | Aliquot A | Aliquot B | Aliquot C | Aliquot Q1 | Aliquot Q2 | Aliquot Q3 |
| 1 | **Al, Na** | **19.91 (0.10)** | **21.94 (0.15)** | **21.27 (0.27)** | **23.44** | **25.88** | **24.52** |
| 2 | **Na** | | | | **neg** | **neg** | **neg** |
| 3 | **Tt** | | | | **neg** | **neg** | **neg** |
| 7 | **Al** | **16.15 (0.49)** | **16.31 (0.16)** | **16.37 (0.16)** | **21.49** | **21.83** | **22.08** |
| 8 | **Tt, Al, Na** | **17.13 (0.18)** | **17.72 (0.08)** | **18.49 (0.19)** | **21.88** | **21.09** | **22.28** |
| 9 | **Tt, Al, Na** | **14.30 (0.58)** | **16.10 (0.34)** | **14.82 (0.26)** | **20.20** | **20.38** | **20.91** |
| 11 | **Tt, Na** | | | | **neg** | **neg** | **neg** |
| 12 | **Na** | | | | **neg** | **neg** | **neg** |
| 13 | **Tt** | | | | **neg** | **neg** | **neg** |
| 14 | **Al** | **13.64 (0.03)** | **13.58 (0.45)** | **13.51 (0.12)** | **17.37** | **16.11** | **17.92** |
| 15 | **Al, Na** | **18.90 (0.08)** | **18.07 (0.11)** | **18.72 (0.07)** | **21.50** | **21.71** | **24.37** |
| 16 | **Tt, Al** | **12.04 (0.09)** | **11.97 (0.15)** | **12.24 (0.09)** | **15.88** | **16.01** | **17.30** |
| 17 | **Na** | | | | **neg** | **neg** | **neg** |
| 18 | **Al** | **13.95 (0.05)** | **24.85 (0.05)** | **14.59 (0.03)** | **18.15** | **20.74** | **18.64** |
| 19 | **Tt, Al, Na** | **19.28 (0.07)** | **18.43 (0.08)** | **19.10 (0.12)** | **20.68** | **22.18** | **20.89** |
| 20 | **Tt, Al, Na** | **18.35 (0.06)** | **18.84 (0.03)** | **18.31 (0.06)** | **20.57** | **20.53** | **20.44** |
| 4 v2 | **Tt, Al** | **17.10 (0.07)** | **17.94 (0.05)** | **16.66 (0.06)** | **20.05** | **21.83** | **20.57** |
| **10 v2** | **Na, Ad** | | | | **neg** | **neg** | **neg** |
| 27 | **Al, Na, Ad** | **12.80 (0.03)** | **12.86 (0.03)** | **13.34 (0.01)** | **17.23** | **18.05** | **18.71** |
| 12 v2 | **Na** | | | | **neg** | **neg** | **neg** |
| 21 | **Bioreclamation** | | | | **neg** | **neg** | **neg** |
| 22 | **Bioreclamation** | | | | **neg** | **neg** | **neg** |
| 23 | **Bioreclamation** | | | | **neg** | **neg** | **neg** |
| 24 | **Bioreclamation** | | | | **neg** | **neg** | **neg** |
| 25 | **Bioreclamation** | | | | **neg** | **neg** | **neg** |

**Table A6. 3:** *Initial Validation Panel, Smith vs. Quantigen Aliquot-Level* ***N. americanus Assay*** *Note that Smith College Ct Scores provide the standard deviation in parentheses.*

| | | N. americanus Ct (Mean- 4 Replicates | | | | | |
|---|---|---|---|---|---|---|---|
| Smith Panel ID | Sample Summary Status | Smith College | | | Quantigen | | |
| | | Aliquot A | Aliquot B | Aliquot C | Aliquot Q1 | Aliquot Q2 | Aliquot Q3 |
| 1 | **Al, Na** | **22.13 (0.08)** | **23.61 (0.11)** | **20.99 (0.14)** | **21.31** | **21.68** | **23.00** |
| 2 | **Na** | **23.93 (0.03)** | **25.48 (0.29)** | **22.43 (0.06)** | **27.21** | **29.68** | **26.13** |
| 3 | **Tt** | | | | **neg** | **neg** | **neg** |
| 7 | **Al** | | | | **neg** | **neg** | **neg** |
| 8 | **Tt, Al, Na** | **24.13 (0.03)** | **21.04 (0.27)** | **26.17 (0.21)** | **22.79** | **32.59** | **29.36** |
| 9 | **Tt, Al, Na** | **24.97 (1.06)** | **25.32 (0.31)** | **23.11 (0.44)** | **30.12** | **26.38** | **29.61** |
| 11 | **Tt, Na** | **23.72 (0.04)** | **25.76 (0.14)** | **25.25 (0.18)** | **29.92** | **25.57** | **31.64** |
| 12 | **Na** | **28.17 (0.10)** | **28.88 (0.10)** | **35.05 (2.55)** | **33.88** | **34.44** | **34.97** |
| 13 | **Tt** | | | | **neg** | **neg** | **neg** |
| 14 | **Al** | | | | **neg** | **neg** | **neg** |
| 15 | **Al, Na** | **22.33 (0.21)** | **25.19 (0.16)** | **25.72 (0.14)** | **27.46** | **28.63** | **28.30** |
| 16 | **Tt, Al** | | | | **neg** | **neg** | **neg** |
| 17 | **Na** | **35.35 (2.17)** | **24.38 (0.14)** | **26.48 (0.07)** | **27.05** | **36.51** | **37.85** |
| 18 | **Al** | | | | **neg** | **neg** | **neg** |
| 19 | **Tt, Al, Na** | **20.90 (0.10)** | **21.81 (0.09)** | **20.97 (0.10)** | **21.81** | **23.48** | **22.12** |
| 20 | **Tt, Al, Na** | **22.52 (0.08)** | **22.65 (0.09)** | **23.31 (0.12)** | **24.29** | **26.49** | **26.06** |
| 4 v2 | **Tt, Al** | | | | **neg** | **neg** | **neg** |
| **10 v2** | Na, Ad | **22.20 (0.24)** | **22.11 (0.21)** | **19.75 (0.06)** | **25.02** | **26.43** | **26.79** |
| 27 | **Al, Na, Ad** | **19.58 (0.19)** | **19.03 (0.15)** | **19.63 (0.11)** | **27.60** | **27.43** | **27.47** |
| 12 v2 | Na | **25.8 (0.17)** | **25.0 (0.21)** | **26.2 (0.32)** | **28.29** | **27.24** | **27.44** |
| 21 | **Bioreclamation** | | | | **neg** | **neg** | **neg** |
| 22 | **Bioreclamation** | | | | **neg** | **neg** | **neg** |
| 23 | **Bioreclamation** | | | | **neg** | **neg** | **neg** |
| 24 | **Bioreclamation** | | | | **neg** | **neg** | **neg** |
| 25 | **Bioreclamation** | | | | **neg** | **neg** | **neg** |

**Table A6.4:** *Initial Validation Panel, Smith vs. Quantigen Aliquot-Level* ***A. duodenale Assay*** *Note that Smith College Ct Scores provide the standard deviation in parentheses.*

| | | A. duodenale Ct (Mean- 4 Replicates) | | | | | |
|---|---|---|---|---|---|---|---|
| Smith Panel ID | Sample Summary Status | Smith College | | | Quantigen | | |
| | | Aliquot A | Aliquot B | Aliquot C | Aliquot Q1 | Aliquot Q2 | Aliquot Q3 |
| 1 | Al, Na | | | | neg | neg | neg |
| 2 | Na | | | | neg | neg | neg |
| 3 | Tt | | | | neg | neg | neg |
| 7 | Al | | | | neg | neg | neg |
| 8 | Tt, Al, Na | | | | neg | neg | neg |
| 9 | Tt, Al, Na | | | | neg | neg | neg |
| 11 | Tt, Na | | | | neg | neg | neg |
| 12 | Na | | | | neg | neg | neg |
| 13 | Tt | | | | neg | neg | neg |
| 14 | Al | | | | neg | neg | neg |
| 15 | Al, Na | | | | neg | neg | neg |
| 16 | Tt, Al | | | | neg | neg | neg |
| 17 | Na | | | | neg | neg | neg |
| 18 | Al | | | | neg | neg | neg |
| 19 | Tt, Al, Na | | | | neg | neg | neg |
| 20 | Tt, Al, Na | | | | neg | neg | neg |
| 4 v2 | Tt, Al | | | | neg | neg | neg |
| **10 v2** | Na, Ad | 30.70 (0.39) | 30.85 (0.87) | 30.98 (0.20) | 34.23 | 37.73 | 38.29 |
| 27 | Al, Na, Ad | 25.86 (0.19) | 26.20 (0.16) | 25.75 (0.23) | 30.35 | 30.24 | 31.73 |
| 12 v2 | Na | | | | neg | neg | neg |
| 21 | Bioreclamation | | | | neg | neg | neg |
| 22 | Bioreclamation | | | | neg | neg | neg |
| 23 | Bioreclamation | | | | neg | neg | neg |
| 24 | Bioreclamation | | | | neg | neg | neg |
| 25 | Bioreclamation | | | | neg | neg | neg |

**Table A6. 5***: Initial Validation Panel Candidates Quantigen data (Technical Replicates),* ***T. trichiura Assay****.*

| | T. trichiura | | | | | | | | | | | |
|---|---|---|---|---|---|---|---|---|---|---|---|---|
| | Ct Scores | | | | | | | | | | | |
| Smith Panel ID | Extraction #1 | | | | Extraction #2 | | | | Extraction #3 | | | |
| 1 | Undetermined | Undetermined | Undetermined | Undetermined | Undetermined | Undetermined | Undetermined | Undetermined | Undetermined | Undetermined | Undetermined | Undetermined |
| 2 | Undetermined | Undetermined | Undetermined | Undetermined | Undetermined | Undetermined | Undetermined | Undetermined | Undetermined | Undetermined | Undetermined | Undetermined |
| 3 | 40.00 | 32.05 | 37.80 | 34.52 | 35.68 | 36.93 | Well Failure | 36.55 | 32.59 | 33.72 | 35.41 | 30.97 |
| 7 | Well Failure | Undetermined | Undetermined | Undetermined | Undetermined | Undetermined | Undetermined | Undetermined | Undetermined | Undetermined | Undetermined | Undetermined |
| 8 | 38.53 | 34.36 | 33.57 | 34.07 | 35.48 | 36.28 | 34.31 | 35.47 | 35.14 | 40.00 | 35.97 | 36.69 |
| 9 | 40.00 | 32.31 | 33.46 | 33.07 | 33.56 | 40.00 | 33.11 | 28.00 | 35.03 | 32.92 | 33.59 | 35.00 |
| 11 | 35.36 | 36.18 | 40.00 | 33.91 | 35.52 | 33.79 | 33.34 | 33.76 | 40.00 | 34.48 | 35.43 | 32.94 |
| 12 | Undetermined | Undetermined | Undetermined | Undetermined | Undetermined | Undetermined | Undetermined | Undetermined | Undetermined | Undetermined | Undetermined | Undetermined |
| 13 | 40.00 | 40.00 | 40.00 | 40.00 | 40.00 | 40.00 | 40.00 | 40.00 | 35.93 | 34.44 | 36.22 | 40.00 |
| 14 | Undetermined | Undetermined | Undetermined | Undetermined | Undetermined | Undetermined | Undetermined | Well Failure | Undetermined | Undetermined | Undetermined | Undetermined |
| 15 | Undetermined | Undetermined | Undetermined | Undetermined | Undetermined | Undetermined | Undetermined | Undetermined | Well Failure | Undetermined | Undetermined | Undetermined |
| 16 | 30.84 | 30.63 | 30.76 | 31.60 | 40.00 | 40.00 | 40.00 | 40.00 | 35.04 | 33.99 | 35.29 | 35.44 |
| 17 | Undetermined | Undetermined | Undetermined | Undetermined | Undetermined | Undetermined | Undetermined | Undetermined | Undetermined | Undetermined | Undetermined | Undetermined |
| 18 | Undetermined | Undetermined | Undetermined | Undetermined | Well Failure | Undetermined | Undetermined | Undetermined | Undetermined | Undetermined | Undetermined | Undetermined |
| 19 | 34.08 | 32.61 | 32.99 | 33.54 | Well Failure | 33.10 | 32.54 | 32.73 | 32.81 | 35.78 | 39.73 | 35.38 |
| 20 | 35.30 | 34.83 | 35.05 | 35.65 | 38.35 | 35.67 | 34.97 | 35.71 | 40.00 | 40.00 | 35.35 | 36.00 |
| 4.2 | 36.46 | 35.53 | 36.57 | 35.70 | 40.00 | 40.00 | 40.00 | 40.00 | 40.00 | 36.52 | 40.00 | 40.00 |
| 10.2 | Undetermined | Undetermined | Undetermined | Undetermined | Undetermined | Undetermined | Undetermined | Undetermined | Undetermined | Undetermined | Undetermined | Undetermined |
| 27 | Undetermined | Undetermined | Undetermined | Undetermined | Undetermined | Undetermined | 36.76 | Undetermined | Undetermined | Undetermined | Undetermined | Undetermined |
| 12.2 | Undetermined | Undetermined | Undetermined | Undetermined | Undetermined | Undetermined | Undetermined | Undetermined | Undetermined | Undetermined | Undetermined | Undetermined |
| 21 | Undetermined | Undetermined | Undetermined | Undetermined | Undetermined | Undetermined | Undetermined | Undetermined | Undetermined | Undetermined | Undetermined | Undetermined |
| 22 | Undetermined | Undetermined | Undetermined | Undetermined | Undetermined | Undetermined | Undetermined | Undetermined | Undetermined | Undetermined | Undetermined | Undetermined |
| 23 | Undetermined | Undetermined | Undetermined | Undetermined | Undetermined | Undetermined | Undetermined | Undetermined | Undetermined | Undetermined | Undetermined | Undetermined |
| 24 | Undetermined | Undetermined | Undetermined | Undetermined | Undetermined | Undetermined | Undetermined | Undetermined | Undetermined | Undetermined | Undetermined | Undetermined |
| 25 | Undetermined | Undetermined | Undetermined | Undetermined | Undetermined | Undetermined | Undetermined | Undetermined | Undetermined | Undetermined | Undetermined | Undetermined |

**Table A6.6:** *Initial Validation Panel Candidates Quantigen data (Technical Replicates),* ***A. lumbricoides Assay****.*

| A. lumbricoides | | | | | | | | | | | | |
|---|---|---|---|---|---|---|---|---|---|---|---|---|
| | Ct Scores | | | | | | | | | | | |
| Smith Panel ID | Extraction #1 | | | | Extraction #2 | | | | Extraction #3 | | | |
| 1 | 23.44 | 23.42 | 23.60 | 23.27 | 25.10 | 26.91 | 26.26 | 25.26 | 24.44 | 24.59 | 24.49 | 24.57 |
| 2 | Undetermined | Undetermined | Undetermined | Undetermined | Undetermined | Undetermined | Undetermined | Undetermined | Undetermined | Undetermined | Undetermined | Undetermined |
| 3 | Undetermined | Undetermined | Undetermined | Undetermined | Undetermined | Undetermined | Undetermined | Undetermined | Undetermined | Undetermined | Undetermined | Undetermined |
| 7 | 22.67 | 21.07 | 21.34 | 20.88 | 20.58 | 22.99 | 22.98 | 20.79 | 22.57 | 22.18 | 21.86 | 21.70 |
| 8 | 22.55 | 21.71 | 21.33 | 21.93 | 21.00 | 21.46 | 20.75 | 21.16 | 22.28 | 22.24 | 22.32 | 22.29 |
| 9 | 20.94 | 20.13 | 19.77 | 19.97 | 19.34 | 21.96 | 19.31 | 20.93 | 21.22 | 20.79 | 20.83 | 20.82 |
| 11 | Undetermined | Undetermined | Undetermined | Undetermined | Undetermined | Undetermined | Undetermined | Undetermined | Undetermined | Undetermined | Undetermined | Undetermined |
| 12 | Undetermined | Undetermined | Undetermined | Undetermined | Undetermined | Undetermined | Undetermined | Undetermined | Undetermined | Undetermined | Undetermined | Undetermined |
| 13 | Undetermined | Undetermined | Undetermined | Undetermined | Undetermined | Undetermined | Undetermined | Undetermined | Undetermined | Undetermined | Undetermined | Undetermined |
| 14 | 17.40 | 17.38 | 17.39 | 17.30 | 15.59 | 15.68 | 15.66 | 17.52 | 18.01 | 17.94 | 17.88 | 17.87 |
| 15 | 21.45 | 21.54 | 21.48 | 21.53 | 21.45 | 21.48 | 21.41 | 22.52 | 24.42 | 24.36 | 24.36 | 24.34 |
| 16 | 15.86 | 15.74 | 15.97 | 15.93 | 15.85 | 15.80 | 15.63 | 16.77 | 17.43 | 17.71 | 16.98 | 17.06 |
| 17 | Undetermined | Undetermined | Undetermined | Undetermined | Undetermined | Undetermined | Undetermined | Undetermined | Undetermined | Undetermined | Undetermined | Undetermined |
| 18 | 18.21 | 18.35 | 18.27 | 17.78 | 29.50 | 17.66 | 17.97 | 17.84 | 18.26 | 18.87 | 18.90 | 18.52 |
| 19 | 20.79 | 20.76 | 20.61 | 20.57 | 25.74 | 21.00 | 21.23 | 20.75 | 20.35 | 21.27 | 21.27 | 20.69 |
| 20 | 20.45 | 20.55 | 20.51 | 20.75 | 21.40 | 20.24 | 20.24 | 20.25 | 20.30 | 20.62 | 20.43 | 20.39 |
| 4.2 | 19.79 | 20.06 | 20.00 | 20.36 | 22.10 | 22.09 | 21.53 | 21.62 | 20.67 | 20.60 | 20.46 | 20.56 |
| 10.2 | Undetermined | Undetermined | Undetermined | Undetermined | Undetermined | Undetermined | Undetermined | Undetermined | Undetermined | Undetermined | Undetermined | Undetermined |
| 27 | 17.00 | 17.03 | 17.32 | 17.56 | 17.94 | 18.02 | 18.05 | 18.18 | 18.39 | 19.28 | 18.63 | 18.55 |
| 12.2 | Undetermined | Undetermined | Undetermined | Undetermined | Undetermined | Undetermined | Undetermined | Undetermined | Undetermined | Undetermined | Undetermined | Undetermined |
| 21 | Undetermined | Undetermined | Undetermined | Undetermined | Undetermined | Undetermined | Undetermined | Undetermined | Undetermined | Undetermined | Undetermined | Undetermined |
| 22 | Undetermined | Undetermined | Undetermined | Undetermined | Undetermined | Undetermined | Undetermined | Undetermined | Undetermined | Undetermined | Undetermined | Undetermined |
| 23 | Undetermined | Undetermined | Undetermined | Undetermined | Undetermined | Undetermined | Undetermined | Undetermined | Undetermined | Undetermined | Undetermined | Undetermined |
| 24 | Undetermined | Undetermined | Undetermined | Undetermined | Undetermined | Undetermined | Undetermined | Undetermined | Undetermined | Undetermined | Undetermined | Undetermined |
| 25 | Undetermined | Undetermined | Undetermined | Undetermined | Undetermined | Undetermined | Undetermined | Undetermined | Undetermined | Undetermined | Undetermined | Undetermined |

**Table A6. 7**: *Initial Validation Panel Candidates Quantigen data (Technical Replicates), N. americanus Assay. Standard #17 was dropped from the final DeWorm3 Spiked Stool Standards Panel due to inconsistent Ct Scores across Smith and Quantigen Data; Smith data is summarized in Table A6.3.*

| N. americanus | | | | | | | | | | | | |
|---|---|---|---|---|---|---|---|---|---|---|---|---|
| | Ct Scores | | | | | | | | | | | |
| Smith Panel ID | Extraciton #1 | | | | Extraction #2 | | | | Extraction #3 | | | |
| 1 | 21.27 | 21.31 | 21.27 | 21.39 | 21.86 | 21.43 | 21.52 | 21.93 | 23.06 | 22.82 | 23.10 | 23.02 |
| 2 | 27.17 | 27.06 | 27.54 | 27.06 | 29.62 | 29.54 | 29.89 | 29.68 | 26.03 | 26.47 | 25.82 | 26.21 |
| 3 | Undetermined | Undetermined | Undetermined | Undetermined | Undetermined | Undetermined | Well Failure | Undetermined | Undetermined | Undetermined | Undetermined | Undetermined |
| 7 | Well Failure | Undetermined | Undetermined | Undetermined | Undetermined | Undetermined | Undetermined | Undetermined | Undetermined | Undetermined | Undetermined | Undetermined |
| 8 | 18.63 | 24.16 | 24.05 | 24.30 | 32.94 | 31.81 | 32.63 | 32.97 | 29.37 | 29.50 | 29.25 | 29.32 |
| 9 | 31.08 | 29.89 | 29.78 | 29.73 | 24.71 | 31.29 | 24.80 | 24.74 | 29.46 | 30.02 | 29.50 | 29.46 |
| 11 | 30.10 | 29.78 | 29.91 | 29.88 | 25.60 | 25.50 | 25.66 | 25.54 | 36.30 | 29.75 | 30.39 | 30.13 |
| 12 | 40.00 | 35.65 | 32.84 | 33.16 | 33.02 | 34.49 | 32.93 | 37.33 | 40.00 | 35.59 | 34.22 | 35.08 |
| 13 | Undetermined | Undetermined | Undetermined | Undetermined | Undetermined | Undetermined | Undetermined | Undetermined | Undetermined | Undetermined | Undetermined | Undetermined |
| 14 | Undetermined | Undetermined | Undetermined | Undetermined | Undetermined | Undetermined | Undetermined | Well Failure | Undetermined | Undetermined | Undetermined | Undetermined |
| 15 | 27.45 | 27.48 | 27.44 | 27.46 | 28.40 | 28.57 | 28.71 | 28.85 | Well Failure | 28.68 | 27.87 | 28.35 |
| 16 | Undetermined | Undetermined | Undetermined | Undetermined | Undetermined | Undetermined | Undetermined | Undetermined | Undetermined | Undetermined | Undetermined | Undetermined |
| 17 | 26.88 | 26.94 | 27.05 | 27.34 | 36.35 | 40.00 | 40.00 | 36.66 | 37.85 | 40.00 | 40.00 | 40.00 |
| 18 | Undetermined | Undetermined | Undetermined | Undetermined | Well Failure | Undetermined | Undetermined | Undetermined | Undetermined | Undetermined | Undetermined | Undetermined |
| 19 | 21.72 | 21.66 | 21.87 | 21.99 | Well Failure | 23.58 | 23.43 | 23.43 | 22.04 | 22.14 | 22.17 | 22.14 |
| 20 | 23.77 | 24.51 | 24.35 | 24.53 | 26.27 | 26.56 | 26.44 | 26.69 | 25.95 | 25.98 | 26.25 | 26.06 |
| 4.2 | Undetermined | Undetermined | Undetermined | Undetermined | Undetermined | Undetermined | Undetermined | Undetermined | Undetermined | Undetermined | Undetermined | Undetermined |
| 10.2 | 25.23 | 25.04 | 24.74 | 25.07 | 26.49 | 26.61 | 26.41 | 26.23 | 26.81 | 26.65 | 26.92 | 26.80 |
| 27 | 27.23 | 27.80 | 27.53 | 27.82 | 27.39 | 27.48 | 27.35 | 27.50 | 27.52 | 27.43 | 27.56 | 27.37 |
| 12.2 | 28.47 | 28.15 | 28.34 | 28.20 | 27.38 | 27.51 | 26.89 | 27.19 | 27.50 | 27.47 | 27.41 | 27.40 |
| 21 | Undetermined | Undetermined | Undetermined | Undetermined | Undetermined | Undetermined | Undetermined | Undetermined | Undetermined | Undetermined | Undetermined | Undetermined |
| 22 | Undetermined | Undetermined | Undetermined | Undetermined | Undetermined | Undetermined | Undetermined | Undetermined | Undetermined | Undetermined | Undetermined | Undetermined |
| 23 | Undetermined | Undetermined | Undetermined | Undetermined | Undetermined | Undetermined | Undetermined | Undetermined | Undetermined | Undetermined | Undetermined | Undetermined |
| 24 | Undetermined | Undetermined | Undetermined | Undetermined | Undetermined | Undetermined | Undetermined | Undetermined | Undetermined | Undetermined | Undetermined | Undetermined |
| 25 | Undetermined | Undetermined | Undetermined | Undetermined | Undetermined | Undetermined | Undetermined | Undetermined | Undetermined | Undetermined | Undetermined | Undetermined |

***Table A6. 8***: *Initial Validation Panel Candidates Quantigen data (Technical Replicates),* ***A. duodenale Assay****. Standard #10.2 was dropped from the final DeWorm3 Spiked Stool Standards Panel due to the drop-out of technical replicates observed during DeWorm3 Validation; it is suspected that the gDNA spiked into naïve stool at low-concentration led to degradation of the DNA.*

| | A. duodenale | | | | | | | | | | | |
|---|---|---|---|---|---|---|---|---|---|---|---|---|
| | Ct Scores | | | | | | | | | | | |
| Smith Panel ID | Extraction #1 | | | | Extraction #2 | | | | Extraction #3 | | | |
| 1 | Undetermined | Undetermined | Undetermined | Undetermined | Undetermined | Undetermined | Undetermined | Undetermined | Undetermined | Undetermined | Undetermined | Undetermined |
| 2 | Undetermined | Undetermined | Undetermined | Undetermined | Undetermined | Undetermined | Undetermined | Undetermined | Undetermined | Undetermined | Undetermined | Undetermined |
| 3 | Undetermined | Undetermined | Undetermined | Undetermined | Undetermined | Undetermined | Undetermined | Undetermined | Undetermined | Undetermined | Undetermined | Undetermined |
| 7 | Undetermined | Undetermined | Undetermined | Undetermined | Undetermined | Undetermined | Undetermined | Undetermined | Undetermined | Undetermined | Undetermined | Undetermined |
| 8 | Undetermined | Undetermined | Undetermined | Undetermined | Undetermined | Undetermined | Undetermined | Undetermined | Undetermined | Undetermined | Undetermined | Undetermined |
| 9 | Undetermined | Undetermined | Undetermined | Undetermined | Undetermined | Undetermined | Undetermined | Undetermined | Undetermined | Undetermined | Undetermined | Undetermined |
| 11 | Undetermined | Undetermined | Undetermined | Undetermined | Undetermined | Undetermined | Undetermined | Undetermined | Undetermined | Undetermined | Undetermined | Undetermined |
| 12 | Undetermined | Undetermined | Undetermined | Undetermined | Undetermined | Undetermined | Undetermined | Undetermined | Undetermined | Undetermined | Undetermined | Undetermined |
| 13 | Undetermined | Undetermined | Undetermined | Undetermined | Undetermined | Undetermined | Undetermined | Undetermined | Undetermined | Undetermined | Undetermined | Undetermined |
| 14 | Undetermined | Undetermined | Undetermined | Undetermined | Undetermined | Undetermined | Undetermined | Undetermined | Undetermined | Undetermined | Undetermined | Undetermined |
| 15 | Undetermined | Undetermined | Undetermined | Undetermined | Undetermined | Undetermined | Undetermined | Undetermined | Undetermined | Undetermined | Undetermined | Undetermined |
| 16 | Undetermined | Undetermined | Undetermined | Undetermined | Undetermined | Undetermined | Undetermined | Undetermined | Undetermined | Undetermined | Undetermined | Undetermined |
| 17 | Undetermined | Undetermined | Undetermined | Undetermined | Undetermined | Undetermined | Undetermined | Undetermined | Undetermined | Undetermined | Undetermined | Undetermined |
| 18 | Undetermined | Undetermined | Undetermined | Undetermined | Undetermined | Undetermined | Undetermined | Undetermined | Undetermined | Undetermined | Undetermined | Undetermined |
| 19 | Undetermined | Undetermined | Undetermined | Undetermined | Undetermined | Undetermined | Undetermined | Undetermined | Undetermined | Undetermined | Undetermined | Undetermined |
| 20 | Undetermined | Undetermined | Undetermined | Undetermined | Undetermined | Undetermined | Undetermined | Undetermined | Undetermined | Undetermined | Undetermined | Undetermined |
| 4.2 | Undetermined | Undetermined | Undetermined | Undetermined | Undetermined | Undetermined | Undetermined | Undetermined | Undetermined | Undetermined | Undetermined | Undetermined |
| 10.2 | 34.10 | 34.28 | 34.47 | 34.06 | 38.79 | 40.00 | 36.67 | 40.00 | 40.00 | 40.00 | 39.88 | 36.71 |
| 27 | 30.59 | 30.00 | 30.41 | 30.40 | 30.16 | 30.28 | 30.19 | 30.34 | 31.34 | 32.76 | 31.28 | 31.55 |
| 12.2 | Undetermined | Undetermined | Undetermined | Undetermined | Undetermined | Undetermined | Undetermined | Undetermined | Undetermined | Undetermined | Undetermined | Undetermined |
| 21 | Undetermined | Undetermined | Undetermined | Undetermined | Undetermined | Undetermined | Undetermined | Undetermined | Undetermined | Undetermined | Undetermined | Undetermined |
| 22 | Undetermined | Undetermined | Undetermined | Undetermined | Undetermined | Undetermined | Undetermined | Undetermined | Undetermined | Undetermined | Undetermined | Undetermined |
| 23 | Undetermined | Undetermined | Undetermined | Undetermined | Undetermined | Undetermined | Undetermined | Undetermined | Undetermined | Undetermined | Undetermined | Undetermined |
| 24 | Undetermined | Undetermined | Undetermined | Undetermined | Undetermined | Undetermined | Undetermined | Undetermined | Undetermined | Undetermined | Undetermined | Undetermined |
| 25 | Undetermined | Undetermined | Undetermined | Undetermined | Undetermined | Undetermined | Undetermined | Undetermined | Undetermined | Undetermined | Undetermined | Undetermined |

**Table A6. 9:** *Investigation of inconsistent T. trichiura containing standards: Comparison of qPCR procedures. Each of the three Quantigen extracts from standards #13, #4.2, and #16 were tested by Smith College qPCR Assay in quadruplicate. Comparing the DeWorm3 qPCR Assay results "Quantigen" and the Smith College Results "Smith," suggests the discordance of these standard is not due to significant differences in detection by qPCR procedure.*

| T. trichiura | | | | | | | | | | | | | |
|---|---|---|---|---|---|---|---|---|---|---|---|---|---|
| | Ct Scores | | | | | | | | | | | | |
| Smith Panel ID | | Extraction #1 | | | | Extraction #2 | | | | Extraction #3 | | | |
| 13 | Quantigen | 40.00 | 40.00 | 40.00 | 40.00 | 40.00 | 40.00 | 40.00 | 40.00 | 35.93 | 34.44 | 36.22 | 40.00 |
| | Smith | 40.00 | 40.00 | 40.00 | 40.00 | 40.00 | 40.00 | 40.00 | 40.00 | 33.26 | 33.26 | 40.00 | 40.00 |
| 4.2 | Quantigen | 36.46 | 35.53 | 36.57 | 35.70 | 40.00 | 40.00 | 40.00 | 40.00 | 40.00 | 36.52 | 40.00 | 40.00 |
| | Smith | 35.89 | 35.89 | 40.00 | 40.00 | 40.00 | 40.00 | 40.00 | 40.00 | 40.00 | 40.00 | 40.00 | 40.00 |
| 16 | Quantigen | 30.84 | 30.63 | 30.76 | 31.60 | 40.00 | 40.00 | 40.00 | 40.00 | 35.04 | 33.99 | 35.29 | 35.44 |
| | Smith | 32.75 | 33.59 | 31.12 | 33.18 | 40.00 | 40.00 | 40.00 | 40.00 | 37.06 | 35.71 | 34.20 | 35.19 |

**Table A6. 10:** *Investigation of inconsistent T. trichiura containing standards: Comparison of DNA Extraction, Bead beating procedures. Note all Ct scores represent a single aliquot and qPCR replicate (i.e., each bead type was tested with a unique aliquot of each Standard). Smith College Beadbeating mix is "MP Bio" and the DeWorm3 Assay Beadbeating mix "Omni".*

| ID | Standard Status | N. americanus | | | | T. trichiura | | | | A. lumbricoides | | | |
|---|---|---|---|---|---|---|---|---|---|---|---|---|---|
| | | 3 min | | 30 min | | 3 min | | 30 min | | 3 min | | 30 min | |
| | | MP Bio | Omni | MP Bio | Omni | MP Bio | Omni | MP Bio | Omni | MP Bio | Omni | MP Bio | Omni |
| Std 8 | Tt, Al, Na | 29.70 | 24.73 | 25.75 | 22.24 | 35.08 | 34.95 | 33.59 | 33.67 | 21.23 | 21.57 | 20.82 | 19.96 |
| Std 11 | Tt, Na | 26.42 | 27.32 | 31.59 | 27.12 | 33.40 | UD | 33.82 | 31.91 | 35.61 | UD | 27.90 | UD |
| Std 13 | Tt | UD | UD | UD | UD | UD | 33.04 | UD | 37.61 | UD | UD | UD | 38.86 |
| Std 16 | Tt, Al | UD | UD | UD | UD | 32.48 | 31.70 | 29.87 | 31.55 | 14.88 | 14.77 | 14.23 | 14.00 |

**Table A6. 11:** *Investigation of inconsistent T. trichiura containing standards. Smith College extracted 6 additional aliquots of Standard #11 with the Smith College Beadbeating mix (MP Bio "Mp", n=3) and the DeWorm3 Assay Beadbeating mix (Omni "O", n=3). Samples were qPCR tested in quadruplicate and tested on two qPCR plates (Run 1 & Run 2), see Panel 1. The same experiment was repeated in Panel 2 with 6 additional aliquots of Standard #11.*

Panel 1: Initial Experiment

| | qPCR Run 1 | qPCR Run 2 |
|---|---|---|
| **Sample Name** | **Tt Cт** | **Tt Cт** |
| Mp #1 | 31.08 | 37.75 |
| Mp #1 | UD | UD |
| Mp #1 | 29.90 | 35.97 |
| Mp #1 | 33.69 | 39.78 |
| Mp #2 | UD | 33.05 |
| Mp #2 | 31.16 | 34.29 |
| Mp #2 | UD | 30.80 |
| Mp #2 | UD | 31.12 |
| Mp #3 | 32.50 | 30.83 |
| Mp #3 | 31.05 | 30.96 |
| Mp #3 | 29.92 | 30.57 |
| Mp #3 | 29.84 | 29.42 |
| O #1 | 33.84 | 32.57 |
| O #1 | 34.00 | 30.60 |
| O #1 | 34.25 | UD |
| O #1 | 31.94 | 30.85 |
| O #2 | 30.63 | 30.44 |
| O #2 | 30.21 | 29.59 |
| O #2 | 30.34 | 30.22 |
| O #2 | 34.09 | 29.63 |
| O #3 | 34.58 | UD |
| O #3 | 33.02 | 32.14 |
| O #3 | 34.44 | 31.43 |
| O #3 | 33.09 | 31.76 |

Panel 2: Repeat

| Sample Name | Tt Cт |
|---|---|
| MP v2 #1 | 32.71 |
| MP v2 #1 | 29.43 |
| MP v2 #1 | 29.53 |
| MP v2 #1 | 33.25 |
| MP v2 #2 | 29.66 |
| MP v2 #2 | 29.32 |
| MP v2 #2 | 28.54 |
| MP v2 #2 | 27.75 |
| MP v2 #3 | 34.57 |
| MP v2 #3 | 30.33 |
| MP v2 #3 | 34.39 |
| MP v2 #3 | 30.05 |
| O v2 #1 | 32.03 |
| O v2 #1 | 32.46 |
| O v2 #1 | 34.01 |
| O v2 #1 | 37.24 |
| O v2 #2 | 29.81 |
| O v2 #2 | 29.42 |
| O v2 #2 | 29.95 |
| O v2 #2 | 32.19 |
| O v2 #3 | UD |
| O v2 #3 | UD |
| O v2 #3 | 35.20 |
| O v2 #3 | UD |

#### A7: Evaluation of Assay Specificity (II.B.iv)

**Figure A7. 1:** *Raw Ct Scores for four Benin Specificity Panel Samples identified STH-positive by Smith College DNA extraction and qPCR testing. Note each sample was tested in duplicate by qPCR.*

| Original Smith Data (Ran in duplicate) | | | | |
|---|---|---|---|---|
| Sample ID | N. americanus | T. trichiura | A. lumbricoides | A. duodenale |
| 1 | Undetermined | Undetermined | 31.0 | Undetermined |
| | Undetermined | Undetermined | 30.5 | Undetermined |
| 2 | 27.8 | Undetermined | Undetermined | Undetermined |
| | 28.2 | Undetermined | Undetermined | Undetermined |
| 3 | 21.4 | Undetermined | Undetermined | Undetermined |
| | 21.2 | Undetermined | Undetermined | Undetermined |

**Figure A7. 2:** *(a)* ***DeWorm3 Liquid Assay testing of Smith College DNA extracts*** *of the four Benin Specificity Panel Samples identified STH-positive by the Smith College Assay.* ***(b) DeWorm3 Lyophilized Assay testing of Smith College DNA extracts*** *of the four Benin Specificity Panel Samples identified STH-positive by the Smith College Assay.*

(a)

| Experiment 1: Smith and Quantigen Extracts Ran in Triplicate on Liquid SMITH DATA ONLY | | | | |
|---|---|---|---|---|
| Sample ID | N. americanus | T. trichiura | A. lumbricoides | A. duodenale |
| 1 | Undetermined | Undetermined | 31.8 | Undetermined |
| | Undetermined | Undetermined | 32.2 | Undetermined |
| | Undetermined | Undetermined | 32.1 | Undetermined |
| 2 | 29.7 | Undetermined | Undetermined | Undetermined |
| | 30.0 | Undetermined | Undetermined | Undetermined |
| | 29.6 | Undetermined | Undetermined | Undetermined |
| 3 | 22.4 | Undetermined | Undetermined | Undetermined |
| | 22.4 | Undetermined | Undetermined | Undetermined |
| | 22.4 | Undetermined | Undetermined | Undetermined |

(b)

| Experiment 2: Smith and Quantigen Extracts Ran in Triplicate on Lyo SMITH DATA ONLY | | | | |
|---|---|---|---|---|
| Sample ID | N. americanus | T. trichiura | A. lumbricoides | A. duodenale |
| 1 | Undetermined | Undetermined | 32.5 | Undetermined |
| | Undetermined | Undetermined | 33.8 | Undetermined |
| | Undetermined | Undetermined | 33.9 | Undetermined |
| 2 | 29.6 | Undetermined | Undetermined | Undetermined |
| | 29.1 | Undetermined | Undetermined | Undetermined |
| | 30.1 | Undetermined | Undetermined | Undetermined |
| 3 | 23.0 | Undetermined | Undetermined | Undetermined |
| | 22.4 | Undetermined | Undetermined | Undetermined |
| | 22.9 | Undetermined | Undetermined | Undetermined |

**Figure A7. 3:** ***(a) Re-testing by DeWorm3 Liquid Assay of the Quantigen Extracts*** *of the three Benin Specificity Panel Samples identified STH-positive by the Smith College Assay.* ***(b) Re-testing by DeWorm3 Lyophilized Assay of the Quantigen Extracts*** *of the three Benin Specificity Panel Samples identified STH-positive by the Smith College Assay.*

(a)

| Experiment 1: Smith and Quantigen Extracts Ran in Triplicate on Liquid QUANTIGEN DATA ONLY | | | | |
|---|---|---|---|---|
| **Sample ID** | **N. americanus** | **T. trichiura** | **A. lumbricoides** | **A. duodenale** |
| 1 | Undetermined | Undetermined | 34.0 | Undetermined |
| | Undetermined | Undetermined | 36.2 | Undetermined |
| | Undetermined | Undetermined | 37.5 | Undetermined |
| 2 | Undetermined | Undetermined | Undetermined | Undetermined |
| | Undetermined | Undetermined | Undetermined | Undetermined |
| | Undetermined | Undetermined | Undetermined | Undetermined |
| 3 | Undetermined | Undetermined | Undetermined | Undetermined |
| | Undetermined | Undetermined | Undetermined | Undetermined |
| | Undetermined | Undetermined | Undetermined | Undetermined |

(b)

| Experiment 2: Smith and Quantigen Extracts Ran in Triplicate on Lyo QUANTIGEN DATA ONLY | | | | |
|---|---|---|---|---|
| **Sample ID** | **N. americanus** | **T. trichiura** | **A. lumbricoides** | **A. duodenale** |
| 1 | Undetermined | Undetermined | 37.2 | Undetermined |
| | Undetermined | Undetermined | 38.7 | Undetermined |
| | Undetermined | Undetermined | 36.8 | Undetermined |
| 2 | Undetermined | Undetermined | Undetermined | Undetermined |
| | Undetermined | Undetermined | Undetermined | Undetermined |
| | Undetermined | Undetermined | Undetermined | Undetermined |
| 3 | Undetermined | Undetermined | Undetermined | Undetermined |
| | Undetermined | Undetermined | Undetermined | Undetermined |
| | Undetermined | Undetermined | Undetermined | Undetermined |

**Figure A7. 4***: Re-extraction and qPCR testing by the DeWorm3 procedures at Quantigen of the three Benin Specificity Panel Samples identified STH-positive by Smith College Assay. Note three new aliquots from each sample were extracted in singlet qPCR testing, thus each row represents a separate aliquot.*

| Experiment 3: Original Samples Re-extracted in Triplicate (Quantigen Ext'n Method) | | | | |
|---|---|---|---|---|
| **Sample ID** | **N. americanus** | **T. trichirua** | **A. lumbricoides** | **A. duodenale** |
| 1 | Undetermined | Undetermined | Undetermined | Undetermined |
| | Undetermined | Undetermined | 36.9 | Undetermined |
| | Undetermined | Undetermined | 34.3 | Undetermined |
| 2 | 31.3 | Undetermined | Undetermined | Undetermined |
| | Undetermined | Undetermined | Undetermined | Undetermined |
| | 31.3 | Undetermined | Undetermined | Undetermined |
| 3 | 30.1 | Undetermined | Undetermined | Undetermined |
| | 28.2 | Undetermined | Undetermined | Undetermined |
| | Undetermined | Undetermined | Undetermined | Undetermined |

**Figure A7. 5**: *Re-testing of the five discordant DNA extracts from CMC by the Lyophilized DeWorm3 Assay.*

| Re-qPCR of Original CMC DNA at Quantigen | | | | | |
|---|---|---|---|---|---|
| ***Sample ID*** | ***Replicate*** | ***N. americanus*** | ***T. trichiura*** | ***A. lumbricoides*** | ***A. duodenale*** |
| **1** | **a** | 21.6 | Undetermined | Undetermined | Undetermined |
| | **b** | 21.2 | Undetermined | Undetermined | Undetermined |
| | **c** | 21.3 | Undetermined | Undetermined | Undetermined |
| **2** | **a** | 19.6 | Undetermined | Undetermined | Undetermined |
| | **b** | 20.0 | Undetermined | Undetermined | Undetermined |
| | **c** | 19.8 | Undetermined | Undetermined | Undetermined |
| **3** | **a** | 31.0 | Undetermined | Undetermined | Undetermined |
| | **b** | 31.2 | Undetermined | Undetermined | Undetermined |
| | **c** | 31.0 | Undetermined | Undetermined | Undetermined |
| **4** | **a** | 36.9 | Undetermined | Undetermined | Undetermined |
| | **b** | Undetermined | Undetermined | Undetermined | Undetermined |
| | **c** | 32.7 | Undetermined | Undetermined | Undetermined |
| **5** | **a** | 25.0 | Undetermined | Undetermined | Undetermined |
| | **b** | 25.2 | Undetermined | Undetermined | Undetermined |
| | **c** | 26.6 | Undetermined | Undetermined | Undetermined |

**Figure A7. 6**: *Re-testing of the five discordant Quantigen DNA extracts by the Lyophilized DeWorm3 Assay.*

| Re-qPCR of Original Quantigen DNA | | | | | |
|---|---|---|---|---|---|
| ***Sample ID*** | ***Replicate*** | ***N. americanus*** | ***T. trichiura*** | ***A. lumbricoides*** | ***A. duodenale*** |
| **1** | **a** | Undetermined | Undetermined | Undetermined | Undetermined |
| | **b** | Undetermined | Undetermined | Undetermined | Undetermined |
| | **c** | Undetermined | Undetermined | Undetermined | Undetermined |
| **2** | **a** | Undetermined | Undetermined | Undetermined | Undetermined |
| | **b** | Undetermined | Undetermined | Undetermined | Undetermined |
| | **c** | Undetermined | Undetermined | Undetermined | Undetermined |
| **3** | **a** | Undetermined | Undetermined | Undetermined | Undetermined |
| | **b** | Undetermined | Undetermined | Undetermined | Undetermined |
| | **c** | Undetermined | Undetermined | Undetermined | Undetermined |
| **4** | **a** | Undetermined | Undetermined | Undetermined | Undetermined |
| | **b** | Undetermined | Undetermined | Undetermined | Undetermined |
| | **c** | Undetermined | Undetermined | Undetermined | Undetermined |
| **5** | **a** | Undetermined | Undetermined | Undetermined | Undetermined |
| | **b** | Undetermined | Undetermined | Undetermined | Undetermined |
| | **c** | Undetermined | Undetermined | Undetermined | Undetermined |

**Figure A7. 7:** *Re-extraction of the 5 discordant India samples at Quantigen. Each sample extract was testing in duplicate.*

| Re-Extraction of Samples at Quantigen | | | | | | |
|---|---|---|---|---|---|---|
| ***Sample ID*** | ***Extraction*** | ***Replicate*** | ***N. americanus*** | ***T. trichiura*** | ***A. lumbricoides*** | ***A. duodenale*** |
| **1** | 1 | a | **26.3** | Undetermined | Undetermined | Undetermined |
| | | b | **26.7** | Undetermined | Undetermined | Undetermined |
| | 2 | a | **N/A** | Undetermined | Undetermined | Undetermined |
| | | b | **N/A** | Undetermined | Undetermined | Undetermined |
| **2** | 1 | a | **27.2** | Undetermined | Undetermined | Undetermined |
| | | b | **26.9** | Undetermined | Undetermined | Undetermined |
| | 2 | a | **25.4** | Undetermined | Undetermined | Undetermined |
| | | b | **24.8** | Undetermined | Undetermined | Undetermined |
| **3** | 1 | a | **Undetermined** | Undetermined | Undetermined | Undetermined |
| | | b | **Undetermined** | Undetermined | Undetermined | Undetermined |
| | 2 | a | **29.1** | Undetermined | Undetermined | Undetermined |
| | | b | **29.3** | Undetermined | Undetermined | Undetermined |
| **4** | 1 | a | **36.7** | Undetermined | Undetermined | Undetermined |
| | | b | **36.0** | Undetermined | Undetermined | Undetermined |
| | 2 | a | **N/A** | Undetermined | Undetermined | Undetermined |
| | | b | **N/A** | Undetermined | Undetermined | Undetermined |
| **5** | 1 | a | **Undetermined** | Undetermined | Undetermined | Undetermined |
| | | b | **Undetermined** | Undetermined | Undetermined | Undetermined |
| | 2 | a | **30.3** | Undetermined | Undetermined | Undetermined |
| | | b | **30.2** | Undetermined | Undetermined | Undetermined |

#### A8: 200 Sample Experiment Data

**Figure A8.1:** *The 200 sample experiment found 48 positive samples across 4 extractions. Within these samples, there was discordance, both between sites and within sites.*

| | | Quantigen | | Smith | | | |
|---|---|---|---|---|---|---|---|
| | **Species** | **Extract 1** | **Extract 2** | **Extract 3** | **Extract 4** | **Quantigen** | **Smith** |
| 1 | *N. americanus* | 25.3 | 24.2 | 18.13 | 18.41 | Positive | Positive |
| | *T.trichiura* | Undetermined | Undetermined | Undetermined | Undetermined | | |
| | *A. lumbricoides* | Undetermined | Undetermined | Undetermined | Undetermined | | |
| 2 | *N. americanus* | 36.3 | 35.9 | 32.4 | 26.7 | Positive | Positive |
| | *T.trichiura* | Undetermined | Undetermined | Undetermined | Undetermined | | |
| | *A. lumbricoides* | Undetermined | Undetermined | Undetermined | Undetermined | | |
| 3 | *N. americanus* | 23.6 | 23.5 | 21.11 | 17.29 | Positive | Positive |
| | *T.trichiura* | Undetermined | Undetermined | Undetermined | Undetermined | | |
| | *A. lumbricoides* | Undetermined | Undetermined | Undetermined | Undetermined | | |
| 4 | *N. americanus* | 28.4 | 28.6 | 23.91 | 32.14 | Positive | Positive |
| | *T.trichiura* | Undetermined | Undetermined | Undetermined | Undetermined | | |
| | *A. lumbricoides* | Undetermined | Undetermined | Undetermined | Undetermined | | |
| 5 | *N. americanus* | 28.3 | 27.2 | 23.68 | 24.36 | Positive | Positive |
| | *T.trichiura* | Undetermined | Undetermined | Undetermined | Undetermined | | |
| | *A. lumbricoides* | Undetermined | Undetermined | Undetermined | Undetermined | | |
| 6 | *N. americanus* | 26.9 | 28.5 | 20.77 | 22.83 | Positive | Positive |
| | *T.trichiura* | Undetermined | Undetermined | Undetermined | Undetermined | | |
| | *A. lumbricoides* | Undetermined | Undetermined | Undetermined | Undetermined | | |
| 7 | *N. americanus* | 36.7 | 36.7 | 36.2 | 29.1 | Positive | Positive |
| | *T.trichiura* | Undetermined | Undetermined | Undetermined | Undetermined | | |
| | *A. lumbricoides* | Undetermined | Undetermined | Undetermined | Undetermined | | |
| 8 | *N. americanus* | 33.3 | 34.1 | 30.46 | 29.98 | Positive | Positive |
| | *T.trichiura* | Undetermined | Undetermined | Undetermined | Undetermined | | |
| | *A. lumbricoides* | Undetermined | Undetermined | Undetermined | Undetermined | | |
| 9 | *N. americanus* | Undetermined | Undetermined | Undetermined | Undetermined | Positive | Positive |
| | *T.trichiura* | Undetermined | Undetermined | Undetermined | Undetermined | | |
| | *A. lumbricoides* | 19.8 | 20.8 | 15.6 | 15.2 | | |
| 10 | *N. americanus* | 24.7 | 22.9 | 17.30 | 20.57 | Positive | Positive |
| | *T.trichiura* | Undetermined | Undetermined | Undetermined | Undetermined | | |
| | *A. lumbricoides* | Undetermined | Undetermined | Undetermined | Undetermined | | |

| | | | | | | | |
|---|---|---|---|---|---|---|---|
| 11 | *N. americanus* | Undetermined | Undetermined | Undetermined | Undetermined | Positive | Positive |
| | *T.trichiura* | Undetermined | Undetermined | Undetermined | Undetermined | | |
| | *A. lumbricoides* | 18.5 | 19.1 | 14.4 | 14.9 | | |
| 12 | *N. americanus* | Undetermined | Undetermined | Undetermined | Undetermined | Positive | Positive |
| | *T.trichiura* | Undetermined | Undetermined | Undetermined | Undetermined | | |
| | *A. lumbricoides* | 34.7 | 37.0 | 32.6 | 32.1 | | |
| 13 | *N. americanus* | 25.1 | 31.0 | 19.69 | 18.99 | Positive | Positive |
| | *T.trichiura* | Undetermined | Undetermined | Undetermined | Undetermined | | |
| | *A. lumbricoides* | Undetermined | Undetermined | Undetermined | Undetermined | | |
| 14 | *N. americanus* | Undetermined | Undetermined | Undetermined | Undetermined | Positive | Positive |
| | *T.trichiura* | Undetermined | Undetermined | Undetermined | Undetermined | | |
| | *A. lumbricoides* | 11.7 | 12.3 | 7.8 | 9.3 | | |
| 15 | *N. americanus* | Undetermined | Undetermined | Undetermined | Undetermined | Positive | Positive |
| | *T.trichiura* | Undetermined | Undetermined | Undetermined | Undetermined | | |
| | *A. lumbricoides* | 18.3 | 17.7 | 14.1 | 15.0 | | |
| 16 | *N. americanus* | Undetermined | Undetermined | Undetermined | Undetermined | Positive | Positive |
| | *T.trichiura* | Undetermined | Undetermined | Undetermined | Undetermined | | |
| | *A. lumbricoides* | 18.0 | 19.8 | 14.5 | 14.3 | | |
| 17 | *N. americanus* | Undetermined | Undetermined | Undetermined | Undetermined | Positive | Positive |
| | *T.trichiura* | Undetermined | Undetermined | Undetermined | Undetermined | | |
| | *A. lumbricoides* | 12.6 | 12.5 | 9.0 | 8.5 | | |
| 18 | *N. americanus* | Undetermined | Undetermined | Undetermined | Undetermined | Positive | Positive |
| | *T.trichiura* | Undetermined | Undetermined | Undetermined | Undetermined | | |
| | *A. lumbricoides* | 15.1 | 15.7 | 10.4 | 9.9 | | |
| 19 | *N. americanus* | Undetermined | Undetermined | Undetermined | Undetermined | Positive | Positive |
| | *T.trichiura* | Undetermined | Undetermined | Undetermined | Undetermined | | |
| | *A. lumbricoides* | 19.5 | 21.8 | 14.5 | 14.7 | | |
| 20 | *N. americanus* | Undetermined | Undetermined | Undetermined | Undetermined | Positive | Positive |
| | *T. trichiura* | Undetermined | Undetermined | 28.6 | 26.8 | | |
| | *A. lumbricoides* | 13.9 | 16.6 | 9.9 | 10.2 | | |
| 21 | *N. americanus* | 31.3 | Undetermined | 23.8 | 30.7 | Positive | Positive |
| | *T. trichiura* | Undetermined | Undetermined | Undetermined | Undetermined | | |
| | *A. lumbricoides* | Undetermined | 36.2 | Undetermined | Undetermined | | |
| 22 | *N. americanus* | Undetermined | Undetermined | Undetermined | Undetermined | Positive | Positive |
| | *T.trichiura* | Undetermined | Undetermined | Undetermined | Undetermined | | |
| | *A. lumbricoides* | 13.2 | 15.2 | 9.8 | 10.3 | | |
| 23 | *N. americanus* | 22.2 | 23.7 | 17.91 | 18.65 | Positive | Positive |
| | *T.trichiura* | Undetermined | Undetermined | Undetermined | Undetermined | | |
| | *A. lumbricoides* | 19.3 | 19.2 | 15.1 | 15.4 | | |

| | | | | | | | |
|---|---|---|---|---|---|---|---|
| **24** | ***N. americanus*** | **Undetermined** | **Undetermined** | **Undetermined** | **Undetermined** | **Positive** | **Positive** |
| | ***T.trichiura*** | **Undetermined** | **Undetermined** | **Undetermined** | **Undetermined** | | |
| | ***A. lumbricoides*** | **13.1** | **13.0** | **8.8** | **8.9** | | |
| **25** | ***N. americanus*** | **27.6** | **Undetermined** | **18.7** | **20.6** | **Positive** | **Positive** |
| | ***T.trichiura*** | **Undetermined** | **Undetermined** | **Undetermined** | **Undetermined** | | |
| | ***A. lumbricoides*** | **18.8** | **19.2** | **11.3** | **12.4** | | |
| **26** | ***N. americanus*** | **Undetermined** | **Undetermined** | **Undetermined** | **Undetermined** | **Positive** | **Positive** |
| | ***T.trichiura*** | **Undetermined** | **Undetermined** | **Undetermined** | **Undetermined** | | |
| | ***A. lumbricoides*** | **34.4** | **31.1** | **31.6** | **26.6** | | |
| **27** | ***N. americanus*** | **26.2** | **26.1** | **18.93** | **19.07** | **Positive** | **Positive** |
| | ***T.trichiura*** | **Undetermined** | **Undetermined** | **Undetermined** | **Undetermined** | | |
| | ***A. lumbricoides*** | **Undetermined** | **Undetermined** | **Undetermined** | **Undetermined** | | |
| **28** | ***N. americanus*** | **23.4** | **25.9** | **16.65** | **17.80** | **Positive** | **Positive** |
| | ***T.trichiura*** | **Undetermined** | **Undetermined** | **28.7** | **29.4** | | |
| | ***A. lumbricoides*** | **Undetermined** | **Undetermined** | **Undetermined** | **Undetermined** | | |
| **29** | ***N. americanus*** | **22.8** | **23.7** | **17.79** | **17.40** | **Positive** | **Positive** |
| | ***T.trichiura*** | **Undetermined** | **Undetermined** | **Undetermined** | **Undetermined** | | |
| | ***A. lumbricoides*** | **Undetermined** | **Undetermined** | **Undetermined** | **Undetermined** | | |
| **30** | ***N. americanus*** | **18.5** | **17.8** | **13.89** | **14.34** | **Positive** | **Positive** |
| | ***T.trichiura*** | **Undetermined** | **Undetermined** | **Undetermined** | **Undetermined** | | |
| | ***A. lumbricoides*** | **Undetermined** | **31.2** | **Undetermined** | **32.4** | | |
| **31** | ***N. americanus*** | **Undetermined** | **Undetermined** | **Undetermined** | **Undetermined** | **Positive** | **Positive** |
| | ***T.trichiura*** | **37.1** | **34.1** | **Undetermined** | **32.3** | | |
| | ***A. lumbricoides*** | **Undetermined** | **Undetermined** | **Undetermined** | **Undetermined** | | |
| **32** | ***N. americanus*** | **27.4** | **25.6** | **23.55** | **27.21** | **Positive** | **Positive** |
| | ***T.trichiura*** | **Undetermined** | **Undetermined** | **Undetermined** | **Undetermined** | | |
| | ***A. lumbricoides*** | **Undetermined** | **Undetermined** | **Undetermined** | **Undetermined** | | |
| **33** | ***N. americanus*** | **Undetermined** | **Undetermined** | **Undetermined** | **Undetermined** | **Positive** | **Positive** |
| | ***T.trichiura*** | **Undetermined** | **Undetermined** | **Undetermined** | **Undetermined** | | |
| | ***A. lumbricoides*** | **19.0** | **22.9** | **14.9** | **14.7** | | |
| **34** | ***N. americanus*** | **22.5** | **27.1** | **20.49** | **17.30** | **Positive** | **Positive** |
| | ***T.trichiura*** | **Undetermined** | **Undetermined** | **Undetermined** | **Undetermined** | | |
| | ***A. lumbricoides*** | **Undetermined** | **Undetermined** | **Undetermined** | **Undetermined** | | |
| **35** | ***N. americanus*** | **Undetermined** | **Undetermined** | **Undetermined** | **Undetermined** | **Negative** | **Positive** |
| | ***T.trichiura*** | **Undetermined** | **Undetermined** | **Undetermined** | **Undetermined** | | |
| | ***A. lumbricoides*** | **Undetermined** | **Undetermined** | **33.9** | **32.2** | | |
| **36** | ***N. americanus*** | **Undetermined** | **Undetermined** | **Undetermined** | **Undetermined** | **Positive (1/2)** | **Negative** |
| | ***T.trichiura*** | **Undetermined** | **Undetermined** | **Undetermined** | **Undetermined** | | |
| | ***A. lumbricoides*** | **Undetermined** | **34.2** | **Undetermined** | **Undetermined** | | |

| | | | | | | | |
|---|---|---|---|---|---|---|---|
| **37** | ***N. americanus*** | **Undetermined** | **Undetermined** | **Undetermined** | **Undetermined** | **Negative** | **Positive (1/2)** |
| | ***T.trichiura*** | **Undetermined** | **Undetermined** | **Undetermined** | **Undetermined** | | |
| | ***A. lumbricoides*** | **Undetermined** | **Undetermined** | **Undetermined** | **31.9** | | |
| **38** | ***N. americanus*** | **28.8** | **25.7** | **28.4** | **Undetermined** | **Positive** | **Positive (1/2)** |
| | ***T.trichiura*** | **Undetermined** | **Undetermined** | **Undetermined** | **Undetermined** | | |
| | ***A. lumbricoides*** | **Undetermined** | **Undetermined** | **Undetermined** | **Undetermined** | | |
| **39** | ***N. americanus*** | **Undetermined** | **36.9** | **Undetermined** | **Undetermined** | **Positive (1/2)** | **Negative** |
| | ***T.trichiura*** | **Undetermined** | **Undetermined** | **Undetermined** | **Undetermined** | | |
| | ***A. lumbricoides*** | **Undetermined** | **Undetermined** | **Undetermined** | **Undetermined** | | |
| **40** | ***N. americanus*** | **Undetermined** | **Undetermined** | **Undetermined** | **Undetermined** | **Positive (1/2)** | **Negative** |
| | ***T.trichiura*** | **Undetermined** | **Undetermined** | **Undetermined** | **Undetermined** | | |
| | ***A. lumbricoides*** | **Undetermined** | **36.2** | **Undetermined** | **Undetermined** | | |
| **41** | ***N. americanus*** | **Undetermined** | **Undetermined** | **Undetermined** | **Undetermined** | **Positive (1/2)** | **Negative** |
| | ***T.trichiura*** | **Undetermined** | **Undetermined** | **Undetermined** | **Undetermined** | | |
| | ***A. lumbricoides*** | **Undetermined** | **33.8** | **Undetermined** | **Undetermined** | | |
| **42** | ***N. americanus*** | **Undetermined** | **32.5** | **Undetermined** | **Undetermined** | **Positive (1/2)** | **Negative** |
| | ***T.trichiura*** | **Undetermined** | **Undetermined** | **Undetermined** | **Undetermined** | | |
| | ***A. lumbricoides*** | **Undetermined** | **Undetermined** | **Undetermined** | **Undetermined** | | |
| **43** | ***N. americanus*** | **36.5** | **Undetermined** | **Undetermined** | **Undetermined** | **Positive (1/2)** | **Negative** |
| | ***T.trichiura*** | **Undetermined** | **Undetermined** | **Undetermined** | **Undetermined** | | |
| | ***A. lumbricoides*** | **Undetermined** | **Undetermined** | **Undetermined** | **Undetermined** | | |
| **44** | ***N. americanus*** | **Undetermined** | **Undetermined** | **Undetermined** | **Undetermined** | **Positive (1/2)** | **Negative** |
| | ***T.trichiura*** | **Undetermined** | **Undetermined** | **Undetermined** | **Undetermined** | | |
| | ***A. lumbricoides*** | **Undetermined** | **31.5** | **Undetermined** | **Undetermined** | | |
| **45** | ***N. americanus*** | **Undetermined** | **Undetermined** | **Undetermined** | **Undetermined** | **Positive (1/2)** | **Negative** |
| | ***T.trichiura*** | **Undetermined** | **Undetermined** | **Undetermined** | **Undetermined** | | |
| | ***A. lumbricoides*** | **Undetermined** | **20.0** | **Undetermined** | **Undetermined** | | |
| **46** | ***N. americanus*** | **Undetermined** | **Undetermined** | **33.2** | **Undetermined** | **Negative** | **Positive (1/2)** |
| | ***T.trichiura*** | **Undetermined** | **Undetermined** | **Undetermined** | **Undetermined** | | |
| | ***A. lumbricoides*** | **Undetermined** | **Undetermined** | **Undetermined** | **Undetermined** | | |
| **47** | ***N. americanus*** | **Undetermined** | **Undetermined** | **Undetermined** | **Undetermined** | **Negative** | **Positive (1/2)** |
| | ***T.trichiura*** | **Undetermined** | **Undetermined** | **Undetermined** | **Undetermined** | | |
| | ***A. lumbricoides*** | **Undetermined** | **Undetermined** | **Undetermined** | **35.2** | | |
| **48** | ***N. americanus*** | **Undetermined** | **36.9** | **Undetermined** | **Undetermined** | **Positive(1/2)** | **Negative** |
| | ***T.trichiura*** | **Undetermined** | **Undetermined** | **Undetermined** | **Undetermined** | | |
| | ***A. lumbricoides*** | **Undetermined** | **Undetermined** | **Undetermined** | **Undetermined** | | |

**Figure A8.2:** *All line data for the 200 sample experiment extractions*

| | | Quantigen | | Smith | |
|---|---|---|---|---|---|
| **Sample ID** | **Species** | **Extract 1** | **Extract 2** | **Extract 3** | **Extract 4** |
| 1 | *N. americanus* | 25.3 | 24.2 | 18.13 | 18.41 |
| | *T.trichiura* | Undetermined | Undetermined | Undetermined | Undetermined |
| | *A. lumbricoides* | Undetermined | Undetermined | Undetermined | Undetermined |
| 2 | *N. americanus* | 36.3 | 35.9 | 32.4 | 26.7 |
| | *T.trichiura* | Undetermined | Undetermined | Undetermined | Undetermined |
| | *A. lumbricoides* | Undetermined | Undetermined | Undetermined | Undetermined |
| 3 | *N. americanus* | 23.6 | 23.5 | 21.11 | 17.29 |
| | *T.trichiura* | Undetermined | Undetermined | Undetermined | Undetermined |
| | *A. lumbricoides* | Undetermined | Undetermined | Undetermined | Undetermined |
| 4 | *N. americanus* | 28.4 | 28.6 | 23.91 | 32.14 |
| | *T.trichiura* | Undetermined | Undetermined | Undetermined | Undetermined |
| | *A. lumbricoides* | Undetermined | Undetermined | Undetermined | Undetermined |
| 5 | *N. americanus* | 28.3 | 27.2 | 23.68 | 24.36 |
| | *T.trichiura* | Undetermined | Undetermined | Undetermined | Undetermined |
| | *A. lumbricoides* | Undetermined | Undetermined | Undetermined | Undetermined |
| 6 | *N. americanus* | 26.9 | 28.5 | 20.77 | 22.83 |
| | *T.trichiura* | Undetermined | Undetermined | Undetermined | Undetermined |
| | *A. lumbricoides* | Undetermined | Undetermined | Undetermined | Undetermined |
| 7 | *N. americanus* | 36.7 | 36.7 | 36.2 | 29.1 |
| | *T.trichiura* | Undetermined | Undetermined | Undetermined | Undetermined |
| | *A. lumbricoides* | Undetermined | Undetermined | Undetermined | Undetermined |
| 8 | *N. americanus* | 33.3 | 34.1 | 30.46 | 29.98 |
| | *T.trichiura* | Undetermined | Undetermined | Undetermined | Undetermined |
| | *A. lumbricoides* | Undetermined | Undetermined | Undetermined | Undetermined |
| 9 | *N. americanus* | Undetermined | Undetermined | Undetermined | Undetermined |
| | *T.trichiura* | Undetermined | Undetermined | Undetermined | Undetermined |
| | *A. lumbricoides* | 19.8 | 20.8 | 15.6 | 15.2 |
| 10 | *N. americanus* | 24.7 | 22.9 | 17.30 | 20.57 |
| | *T.trichiura* | Undetermined | Undetermined | Undetermined | Undetermined |
| | *A. lumbricoides* | Undetermined | Undetermined | Undetermined | Undetermined |
| 11 | *N. americanus* | Undetermined | Undetermined | Undetermined | Undetermined |
| | *T.trichiura* | Undetermined | Undetermined | Undetermined | Undetermined |
| | *A. lumbricoides* | 18.5 | 19.1 | 14.4 | 14.9 |
| 12 | *N. americanus* | Undetermined | Undetermined | Undetermined | Undetermined |
| | *T.trichiura* | Undetermined | Undetermined | Undetermined | Undetermined |
| | *A. lumbricoides* | 34.7 | 37.0 | 32.6 | 32.1 |

| | | | | | |
|---|---|---|---|---|---|
| **13** | ***N. americanus*** | **25.1** | **31.0** | **19.69** | **18.99** |
| | ***T.trichiura*** | **Undetermined** | **Undetermined** | **Undetermined** | **Undetermined** |
| | ***A. lumbricoides*** | **Undetermined** | **Undetermined** | **Undetermined** | **Undetermined** |
| **14** | ***N. americanus*** | **Undetermined** | **Undetermined** | **Undetermined** | **Undetermined** |
| | ***T.trichiura*** | **Undetermined** | **Undetermined** | **Undetermined** | **Undetermined** |
| | ***A. lumbricoides*** | **11.7** | **12.3** | **7.8** | **9.3** |
| **15** | ***N. americanus*** | **Undetermined** | **Undetermined** | **Undetermined** | **Undetermined** |
| | ***T.trichiura*** | **Undetermined** | **Undetermined** | **Undetermined** | **Undetermined** |
| | ***A. lumbricoides*** | **18.3** | **17.7** | **14.1** | **15.0** |
| **16** | ***N. americanus*** | **Undetermined** | **Undetermined** | **Undetermined** | **Undetermined** |
| | ***T.trichiura*** | **Undetermined** | **Undetermined** | **Undetermined** | **Undetermined** |
| | ***A. lumbricoides*** | **18.0** | **19.8** | **14.5** | **14.3** |
| **17** | ***N. americanus*** | **Undetermined** | **Undetermined** | **Undetermined** | **Undetermined** |
| | ***T.trichiura*** | **Undetermined** | **Undetermined** | **Undetermined** | **Undetermined** |
| | ***A. lumbricoides*** | **12.6** | **12.5** | **9.0** | **8.5** |
| **18** | ***N. americanus*** | **Undetermined** | **Undetermined** | **Undetermined** | **Undetermined** |
| | ***T.trichiura*** | **Undetermined** | **Undetermined** | **Undetermined** | **Undetermined** |
| | ***A. lumbricoides*** | **15.1** | **15.7** | **10.4** | **9.9** |
| **19** | ***N. americanus*** | **Undetermined** | **Undetermined** | **Undetermined** | **Undetermined** |
| | ***T.trichiura*** | **Undetermined** | **Undetermined** | **Undetermined** | **Undetermined** |
| | ***A. lumbricoides*** | **19.5** | **21.8** | **14.5** | **14.7** |
| **20** | ***N. americanus*** | **Undetermined** | **Undetermined** | **Undetermined** | **Undetermined** |
| | ***T.trichiura*** | **Undetermined** | **Undetermined** | **28.6** | **26.8** |
| | ***A. lumbricoides*** | **13.9** | **16.6** | **9.9** | **10.2** |
| **21** | ***N. americanus*** | **31.3** | **Undetermined** | **23.8** | **30.7** |
| | ***T.trichiura*** | **Undetermined** | **Undetermined** | **Undetermined** | **Undetermined** |
| | ***A. lumbricoides*** | **Undetermined** | **36.2** | **Undetermined** | **Undetermined** |
| **22** | ***N. americanus*** | **Undetermined** | **Undetermined** | **Undetermined** | **Undetermined** |
| | ***T.trichiura*** | **Undetermined** | **Undetermined** | **Undetermined** | **Undetermined** |
| | ***A. lumbricoides*** | **13.2** | **15.2** | **9.8** | **10.3** |
| **23** | ***N. americanus*** | **22.2** | **23.7** | **17.91** | **18.65** |
| | ***T.trichiura*** | **Undetermined** | **Undetermined** | **Undetermined** | **Undetermined** |
| | ***A. lumbricoides*** | **19.3** | **19.2** | **15.1** | **15.4** |
| **24** | ***N. americanus*** | **Undetermined** | **Undetermined** | **Undetermined** | **Undetermined** |
| | ***T.trichiura*** | **Undetermined** | **Undetermined** | **Undetermined** | **Undetermined** |
| | ***A. lumbricoides*** | **13.1** | **13.0** | **8.8** | **8.9** |
| **25** | ***N. americanus*** | **27.6** | **Undetermined** | **18.7** | **20.6** |
| | ***T.trichiura*** | **Undetermined** | **Undetermined** | **Undetermined** | **Undetermined** |
| | ***A. lumbricoides*** | **18.8** | **19.2** | **11.3** | **12.4** |
| **26** | ***N. americanus*** | **Undetermined** | **Undetermined** | **Undetermined** | **Undetermined** |
| | ***T.trichiura*** | **Undetermined** | **Undetermined** | **Undetermined** | **Undetermined** |
| | ***A. lumbricoides*** | **34.4** | **31.1** | **31.6** | **26.6** |

| | | | | | |
|---|---|---|---|---|---|
| **27** | ***N. americanus*** | **26.2** | **26.1** | **18.93** | **19.07** |
| | ***T.trichiura*** | **Undetermined** | **Undetermined** | **Undetermined** | **Undetermined** |
| | ***A. lumbricoides*** | **Undetermined** | **Undetermined** | **Undetermined** | **Undetermined** |
| **28** | ***N. americanus*** | **23.4** | **25.9** | **16.65** | **17.80** |
| | ***T.trichiura*** | **Undetermined** | **Undetermined** | **28.7** | **29.4** |
| | ***A. lumbricoides*** | **Undetermined** | **Undetermined** | **Undetermined** | **Undetermined** |
| **29** | ***N. americanus*** | **22.8** | **23.7** | **17.79** | **17.40** |
| | ***T.trichiura*** | **Undetermined** | **Undetermined** | **Undetermined** | **Undetermined** |
| | ***A. lumbricoides*** | **Undetermined** | **Undetermined** | **Undetermined** | **Undetermined** |
| **30** | ***N. americanus*** | **18.5** | **17.8** | **13.89** | **14.34** |
| | ***T.trichiura*** | **Undetermined** | **Undetermined** | **Undetermined** | **Undetermined** |
| | ***A. lumbricoides*** | **Undetermined** | **31.2** | **Undetermined** | **32.4** |
| **31** | ***N. americanus*** | **Undetermined** | **Undetermined** | **Undetermined** | **Undetermined** |
| | ***T.trichiura*** | **37.1** | **34.1** | **Undetermined** | **32.3** |
| | ***A. lumbricoides*** | **Undetermined** | **Undetermined** | **Undetermined** | **Undetermined** |
| **32** | ***N. americanus*** | **27.4** | **25.6** | **23.55** | **27.21** |
| | ***T.trichiura*** | **Undetermined** | **Undetermined** | **Undetermined** | **Undetermined** |
| | ***A. lumbricoides*** | **Undetermined** | **Undetermined** | **Undetermined** | **Undetermined** |
| **33** | ***N. americanus*** | **Undetermined** | **Undetermined** | **Undetermined** | **Undetermined** |
| | ***T.trichiura*** | **Undetermined** | **Undetermined** | **Undetermined** | **Undetermined** |
| | ***A. lumbricoides*** | **19.0** | **22.9** | **14.9** | **14.7** |
| **34** | ***N. americanus*** | **22.5** | **27.1** | **20.49** | **17.30** |
| | ***T.trichiura*** | **Undetermined** | **Undetermined** | **Undetermined** | **Undetermined** |
| | ***A. lumbricoides*** | **Undetermined** | **Undetermined** | **Undetermined** | **Undetermined** |
| **35** | ***N. americanus*** | **Undetermined** | **Undetermined** | **Undetermined** | **Undetermined** |
| | ***T.trichiura*** | **Undetermined** | **Undetermined** | **Undetermined** | **Undetermined** |
| | ***A. lumbricoides*** | **Undetermined** | **Undetermined** | **33.9** | **32.2** |
| **36** | ***N. americanus*** | **Undetermined** | **Undetermined** | **Undetermined** | **Undetermined** |
| | ***T.trichiura*** | **Undetermined** | **Undetermined** | **Undetermined** | **Undetermined** |
| | ***A. lumbricoides*** | **Undetermined** | **34.2** | **Undetermined** | **Undetermined** |
| **37** | ***N. americanus*** | **Undetermined** | **Undetermined** | **Undetermined** | **Undetermined** |
| | ***T.trichiura*** | **Undetermined** | **Undetermined** | **Undetermined** | **Undetermined** |
| | ***A. lumbricoides*** | **Undetermined** | **Undetermined** | **Undetermined** | **31.9** |
| **38** | ***N. americanus*** | **28.8** | **25.7** | **28.4** | **Undetermined** |
| | ***T.trichiura*** | **Undetermined** | **Undetermined** | **Undetermined** | **Undetermined** |
| | ***A. lumbricoides*** | **Undetermined** | **Undetermined** | **Undetermined** | **Undetermined** |
| **39** | ***N. americanus*** | **Undetermined** | **36.9** | **Undetermined** | **Undetermined** |
| | ***T.trichiura*** | **Undetermined** | **Undetermined** | **Undetermined** | **Undetermined** |
| | ***A. lumbricoides*** | **Undetermined** | **Undetermined** | **Undetermined** | **Undetermined** |

| | | | | | |
|---|---|---|---|---|---|
| **40** | ***N. americanus*** | **Undetermined** | **Undetermined** | **Undetermined** | **Undetermined** |
| | ***T.trichiura*** | **Undetermined** | **Undetermined** | **Undetermined** | **Undetermined** |
| | ***A. lumbricoides*** | **Undetermined** | **36.2** | **Undetermined** | **Undetermined** |
| **41** | ***N. americanus*** | **Undetermined** | **Undetermined** | **Undetermined** | **Undetermined** |
| | ***T.trichiura*** | **Undetermined** | **Undetermined** | **Undetermined** | **Undetermined** |
| | ***A. lumbricoides*** | **Undetermined** | **33.8** | **Undetermined** | **Undetermined** |
| **42** | ***N. americanus*** | **Undetermined** | **32.5** | **Undetermined** | **Undetermined** |
| | ***T.trichiura*** | **Undetermined** | **Undetermined** | **Undetermined** | **Undetermined** |
| | ***A. lumbricoides*** | **Undetermined** | **Undetermined** | **Undetermined** | **Undetermined** |
| **43** | ***N. americanus*** | **36.5** | **Undetermined** | **Undetermined** | **Undetermined** |
| | ***T.trichiura*** | **Undetermined** | **Undetermined** | **Undetermined** | **Undetermined** |
| | ***A. lumbricoides*** | **Undetermined** | **Undetermined** | **Undetermined** | **Undetermined** |
| **44** | ***N. americanus*** | **Undetermined** | **Undetermined** | **Undetermined** | **Undetermined** |
| | ***T.trichiura*** | **Undetermined** | **Undetermined** | **Undetermined** | **Undetermined** |
| | ***A. lumbricoides*** | **Undetermined** | **31.5** | **Undetermined** | **Undetermined** |
| **45** | ***N. americanus*** | **Undetermined** | **Undetermined** | **Undetermined** | **Undetermined** |
| | ***T.trichiura*** | **Undetermined** | **Undetermined** | **Undetermined** | **Undetermined** |
| | ***A. lumbricoides*** | **Undetermined** | **20.0** | **Undetermined** | **Undetermined** |
| **46** | ***N. americanus*** | **Undetermined** | **Undetermined** | **33.2** | **Undetermined** |
| | ***T.trichiura*** | **Undetermined** | **Undetermined** | **Undetermined** | **Undetermined** |
| | ***A. lumbricoides*** | **Undetermined** | **Undetermined** | **Undetermined** | **Undetermined** |
| **47** | ***N. americanus*** | **Undetermined** | **Undetermined** | **Undetermined** | **Undetermined** |
| | ***T.trichiura*** | **Undetermined** | **Undetermined** | **Undetermined** | **Undetermined** |
| | ***A. lumbricoides*** | **Undetermined** | **Undetermined** | **Undetermined** | **35.2** |
| **48** | ***N. americanus*** | **Undetermined** | **36.9** | **Undetermined** | **Undetermined** |
| | ***T.trichiura*** | **Undetermined** | **Undetermined** | **Undetermined** | **Undetermined** |
| | ***A. lumbricoides*** | **Undetermined** | **Undetermined** | **Undetermined** | **Undetermined** |
| **49** | ***N. americanus*** | **Undetermined** | **Undetermined** | **Undetermined** | **Undetermined** |
| | ***T.trichiura*** | **Undetermined** | **Undetermined** | **Undetermined** | **Undetermined** |
| | ***A. lumbricoides*** | **Undetermined** | **Undetermined** | **Undetermined** | **Undetermined** |
| **50** | ***N. americanus*** | **Undetermined** | **Undetermined** | **Undetermined** | **Undetermined** |
| | ***T.trichiura*** | **Undetermined** | **Undetermined** | **Undetermined** | **Undetermined** |
| | ***A. lumbricoides*** | **Undetermined** | **Undetermined** | **Undetermined** | **Undetermined** |
| **51** | ***N. americanus*** | **Undetermined** | **Undetermined** | **Undetermined** | **Undetermined** |
| | ***T.trichiura*** | **Undetermined** | **Undetermined** | **Undetermined** | **Undetermined** |
| | ***A. lumbricoides*** | **Undetermined** | **Undetermined** | **Undetermined** | **Undetermined** |
| **52** | ***N. americanus*** | **Undetermined** | **Undetermined** | **Undetermined** | **Undetermined** |
| | ***T.trichiura*** | **Undetermined** | **Undetermined** | **Undetermined** | **Undetermined** |
| | ***A. lumbricoides*** | **Undetermined** | **Undetermined** | **Undetermined** | **Undetermined** |

| | | | | | |
|---|---|---|---|---|---|
| **53** | ***N. americanus*** | **Undetermined** | **Undetermined** | **Undetermined** | **Undetermined** |
| | ***T.trichiura*** | **Undetermined** | **Undetermined** | **Undetermined** | **Undetermined** |
| | ***A. lumbricoides*** | **Undetermined** | **Undetermined** | **Undetermined** | **Undetermined** |
| **54** | ***N. americanus*** | **Undetermined** | **Undetermined** | **Undetermined** | **Undetermined** |
| | ***T.trichiura*** | **Undetermined** | **Undetermined** | **Undetermined** | **Undetermined** |
| | ***A. lumbricoides*** | **Undetermined** | **Undetermined** | **Undetermined** | **Undetermined** |
| **55** | ***N. americanus*** | **Undetermined** | **Undetermined** | **Undetermined** | **Undetermined** |
| | ***T.trichiura*** | **Undetermined** | **Undetermined** | **Undetermined** | **Undetermined** |
| | ***A. lumbricoides*** | **Undetermined** | **Undetermined** | **Undetermined** | **Undetermined** |
| **56** | ***N. americanus*** | **Undetermined** | **Undetermined** | **Undetermined** | **Undetermined** |
| | ***T.trichiura*** | **Undetermined** | **Undetermined** | **Undetermined** | **Undetermined** |
| | ***A. lumbricoides*** | **Undetermined** | **Undetermined** | **Undetermined** | **Undetermined** |
| **57** | ***N. americanus*** | **Undetermined** | **Undetermined** | **Undetermined** | **Undetermined** |
| | ***T.trichiura*** | **Undetermined** | **Undetermined** | **Undetermined** | **Undetermined** |
| | ***A. lumbricoides*** | **Undetermined** | **Undetermined** | **Undetermined** | **Undetermined** |
| **58** | ***N. americanus*** | **Undetermined** | **Undetermined** | **Undetermined** | **Undetermined** |
| | ***T.trichiura*** | **Undetermined** | **Undetermined** | **Undetermined** | **Undetermined** |
| | ***A. lumbricoides*** | **Undetermined** | **Undetermined** | **Undetermined** | **Undetermined** |
| **59** | ***N. americanus*** | **Undetermined** | **Undetermined** | **Undetermined** | **Undetermined** |
| | ***T.trichiura*** | **Undetermined** | **Undetermined** | **Undetermined** | **Undetermined** |
| | ***A. lumbricoides*** | **Undetermined** | **Undetermined** | **Undetermined** | **Undetermined** |
| **60** | ***N. americanus*** | **Undetermined** | **Undetermined** | **Undetermined** | **Undetermined** |
| | ***T.trichiura*** | **Undetermined** | **Undetermined** | **Undetermined** | **Undetermined** |
| | ***A. lumbricoides*** | **Undetermined** | **Undetermined** | **Undetermined** | **Undetermined** |
| **61** | ***N. americanus*** | **Undetermined** | **Undetermined** | **Undetermined** | **Undetermined** |
| | ***T.trichiura*** | **Undetermined** | **Undetermined** | **Undetermined** | **Undetermined** |
| | ***A. lumbricoides*** | **Undetermined** | **Undetermined** | **Undetermined** | **Undetermined** |
| **62** | ***N. americanus*** | **Undetermined** | **Undetermined** | **Undetermined** | **Undetermined** |
| | ***T.trichiura*** | **Undetermined** | **Undetermined** | **Undetermined** | **Undetermined** |
| | ***A. lumbricoides*** | **Undetermined** | **Undetermined** | **Undetermined** | **Undetermined** |
| **63** | ***N. americanus*** | **Undetermined** | **Undetermined** | **Undetermined** | **Undetermined** |
| | ***T.trichiura*** | **Undetermined** | **Undetermined** | **Undetermined** | **Undetermined** |
| | ***A. lumbricoides*** | **Undetermined** | **Undetermined** | **Undetermined** | **Undetermined** |
| **64** | ***N. americanus*** | **Undetermined** | **Undetermined** | **Undetermined** | **Undetermined** |
| | ***T.trichiura*** | **Undetermined** | **Undetermined** | **Undetermined** | **Undetermined** |
| | ***A. lumbricoides*** | **Undetermined** | **Undetermined** | **Undetermined** | **Undetermined** |
| **65** | ***N. americanus*** | **Undetermined** | **Undetermined** | **Undetermined** | **Undetermined** |
| | ***T.trichiura*** | **Undetermined** | **Undetermined** | **Undetermined** | **Undetermined** |
| | ***A. lumbricoides*** | **Undetermined** | **Undetermined** | **Undetermined** | **Undetermined** |

| | | | | | |
|---|---|---|---|---|---|
| 66 | *N. americanus* | Undetermined | Undetermined | Undetermined | Undetermined |
| | *T.trichiura* | Undetermined | Undetermined | Undetermined | Undetermined |
| | *A. lumbricoides* | Undetermined | Undetermined | Undetermined | Undetermined |
| 67 | *N. americanus* | Undetermined | Undetermined | Undetermined | Undetermined |
| | *T.trichiura* | Undetermined | Undetermined | Undetermined | Undetermined |
| | *A. lumbricoides* | Undetermined | Undetermined | Undetermined | Undetermined |
| 68 | *N. americanus* | Undetermined | Undetermined | Undetermined | Undetermined |
| | *T.trichiura* | Undetermined | Undetermined | Undetermined | Undetermined |
| | *A. lumbricoides* | Undetermined | Undetermined | Undetermined | Undetermined |
| 69 | *N. americanus* | Undetermined | Undetermined | Undetermined | Undetermined |
| | *T.trichiura* | Undetermined | Undetermined | Undetermined | Undetermined |
| | *A. lumbricoides* | Undetermined | Undetermined | Undetermined | Undetermined |
| 70 | *N. americanus* | Undetermined | Undetermined | Undetermined | Undetermined |
| | *T.trichiura* | Undetermined | Undetermined | Undetermined | Undetermined |
| | *A. lumbricoides* | Undetermined | Undetermined | Undetermined | Undetermined |
| 71 | *N. americanus* | Undetermined | Undetermined | Undetermined | Undetermined |
| | *T.trichiura* | Undetermined | Undetermined | Undetermined | Undetermined |
| | *A. lumbricoides* | Undetermined | Undetermined | Undetermined | Undetermined |
| 72 | *N. americanus* | Undetermined | Undetermined | Undetermined | Undetermined |
| | *T.trichiura* | Undetermined | Undetermined | Undetermined | Undetermined |
| | *A. lumbricoides* | Undetermined | Undetermined | Undetermined | Undetermined |
| 73 | *N. americanus* | Undetermined | Undetermined | Undetermined | Undetermined |
| | *T.trichiura* | Undetermined | Undetermined | Undetermined | Undetermined |
| | *A. lumbricoides* | Undetermined | Undetermined | Undetermined | Undetermined |
| 74 | *N. americanus* | Undetermined | Undetermined | Undetermined | Undetermined |
| | *T.trichiura* | Undetermined | Undetermined | Undetermined | Undetermined |
| | *A. lumbricoides* | Undetermined | Undetermined | Undetermined | Undetermined |
| 75 | *N. americanus* | Undetermined | Undetermined | Undetermined | Undetermined |
| | *T.trichiura* | Undetermined | Undetermined | Undetermined | Undetermined |
| | *A. lumbricoides* | Undetermined | Undetermined | Undetermined | Undetermined |
| 76 | *N. americanus* | Undetermined | Undetermined | Undetermined | Undetermined |
| | *T.trichiura* | Undetermined | Undetermined | Undetermined | Undetermined |
| | *A. lumbricoides* | Undetermined | Undetermined | Undetermined | Undetermined |
| 77 | *N. americanus* | Undetermined | Undetermined | Undetermined | Undetermined |
| | *T.trichiura* | Undetermined | Undetermined | Undetermined | Undetermined |
| | *A. lumbricoides* | Undetermined | Undetermined | Undetermined | Undetermined |
| 78 | *N. americanus* | Undetermined | Undetermined | Undetermined | Undetermined |
| | *T.trichiura* | Undetermined | Undetermined | Undetermined | Undetermined |
| | *A. lumbricoides* | Undetermined | Undetermined | Undetermined | Undetermined |

| | | | | | |
|---|---|---|---|---|---|
| **79** | ***N. americanus*** | **Undetermined** | **Undetermined** | **Undetermined** | **Undetermined** |
| | ***T.trichiura*** | **Undetermined** | **Undetermined** | **Undetermined** | **Undetermined** |
| | ***A. lumbricoides*** | **Undetermined** | **Undetermined** | **Undetermined** | **Undetermined** |
| **80** | ***N. americanus*** | **Undetermined** | **Undetermined** | **Undetermined** | **Undetermined** |
| | ***T.trichiura*** | **Undetermined** | **Undetermined** | **Undetermined** | **Undetermined** |
| | ***A. lumbricoides*** | **Undetermined** | **Undetermined** | **Undetermined** | **Undetermined** |
| **81** | ***N. americanus*** | **Undetermined** | **Undetermined** | **Undetermined** | **Undetermined** |
| | ***T.trichiura*** | **Undetermined** | **Undetermined** | **Undetermined** | **Undetermined** |
| | ***A. lumbricoides*** | **Undetermined** | **Undetermined** | **Undetermined** | **Undetermined** |
| **82** | ***N. americanus*** | **Undetermined** | **Undetermined** | **Undetermined** | **Undetermined** |
| | ***T.trichiura*** | **Undetermined** | **Undetermined** | **Undetermined** | **Undetermined** |
| | ***A. lumbricoides*** | **Undetermined** | **Undetermined** | **Undetermined** | **Undetermined** |
| **83** | ***N. americanus*** | **Undetermined** | **Undetermined** | **Undetermined** | **Undetermined** |
| | ***T.trichiura*** | **Undetermined** | **Undetermined** | **Undetermined** | **Undetermined** |
| | ***A. lumbricoides*** | **Undetermined** | **Undetermined** | **Undetermined** | **Undetermined** |
| **84** | ***N. americanus*** | **Undetermined** | **Undetermined** | **Undetermined** | **Undetermined** |
| | ***T.trichiura*** | **Undetermined** | **Undetermined** | **Undetermined** | **Undetermined** |
| | ***A. lumbricoides*** | **Undetermined** | **Undetermined** | **Undetermined** | **Undetermined** |
| **85** | ***N. americanus*** | **Undetermined** | **Undetermined** | **Undetermined** | **Undetermined** |
| | ***T.trichiura*** | **Undetermined** | **Undetermined** | **Undetermined** | **Undetermined** |
| | ***A. lumbricoides*** | **Undetermined** | **Undetermined** | **Undetermined** | **Undetermined** |
| **86** | ***N. americanus*** | **Undetermined** | **Undetermined** | **Undetermined** | **Undetermined** |
| | ***T.trichiura*** | **Undetermined** | **Undetermined** | **Undetermined** | **Undetermined** |
| | ***A. lumbricoides*** | **Undetermined** | **Undetermined** | **Undetermined** | **Undetermined** |
| **87** | ***N. americanus*** | **Undetermined** | **Undetermined** | **Undetermined** | **Undetermined** |
| | ***T.trichiura*** | **Undetermined** | **Undetermined** | **Undetermined** | **Undetermined** |
| | ***A. lumbricoides*** | **Undetermined** | **Undetermined** | **Undetermined** | **Undetermined** |
| **88** | ***N. americanus*** | **Undetermined** | **Undetermined** | **Undetermined** | **Undetermined** |
| | ***T.trichiura*** | **Undetermined** | **Undetermined** | **Undetermined** | **Undetermined** |
| | ***A. lumbricoides*** | **Undetermined** | **Undetermined** | **Undetermined** | **Undetermined** |
| **89** | ***N. americanus*** | **Undetermined** | **Undetermined** | **Undetermined** | **Undetermined** |
| | ***T.trichiura*** | **Undetermined** | **Undetermined** | **Undetermined** | **Undetermined** |
| | ***A. lumbricoides*** | **Undetermined** | **Undetermined** | **Undetermined** | **Undetermined** |
| **90** | ***N. americanus*** | **Undetermined** | **Undetermined** | **Undetermined** | **Undetermined** |
| | ***T.trichiura*** | **Undetermined** | **Undetermined** | **Undetermined** | **Undetermined** |
| | ***A. lumbricoides*** | **Undetermined** | **Undetermined** | **Undetermined** | **Undetermined** |
| **91** | ***N. americanus*** | **Undetermined** | **Undetermined** | **Undetermined** | **Undetermined** |
| | ***T.trichiura*** | **Undetermined** | **Undetermined** | **Undetermined** | **Undetermined** |
| | ***A. lumbricoides*** | **Undetermined** | **Undetermined** | **Undetermined** | **Undetermined** |

| | | | | | |
|---|---|---|---|---|---|
| 92 | ***N. americanus*** | **Undetermined** | **Undetermined** | **Undetermined** | **Undetermined** |
| | ***T.trichiura*** | **Undetermined** | **Undetermined** | **Undetermined** | **Undetermined** |
| | ***A. lumbricoides*** | **Undetermined** | **Undetermined** | **Undetermined** | **Undetermined** |
| 93 | ***N. americanus*** | **Undetermined** | **Undetermined** | **Undetermined** | **Undetermined** |
| | ***T.trichiura*** | **Undetermined** | **Undetermined** | **Undetermined** | **Undetermined** |
| | ***A. lumbricoides*** | **Undetermined** | **Undetermined** | **Undetermined** | **Undetermined** |
| 94 | ***N. americanus*** | **Undetermined** | **Undetermined** | **Undetermined** | **Undetermined** |
| | ***T.trichiura*** | **Undetermined** | **Undetermined** | **Undetermined** | **Undetermined** |
| | ***A. lumbricoides*** | **Undetermined** | **Undetermined** | **Undetermined** | **Undetermined** |
| 95 | ***N. americanus*** | **Undetermined** | **Undetermined** | **Undetermined** | **Undetermined** |
| | ***T.trichiura*** | **Undetermined** | **Undetermined** | **Undetermined** | **Undetermined** |
| | ***A. lumbricoides*** | **Undetermined** | **Undetermined** | **Undetermined** | **Undetermined** |
| 96 | ***N. americanus*** | **Undetermined** | **Undetermined** | **Undetermined** | **Undetermined** |
| | ***T.trichiura*** | **Undetermined** | **Undetermined** | **Undetermined** | **Undetermined** |
| | ***A. lumbricoides*** | **Undetermined** | **Undetermined** | **Undetermined** | **Undetermined** |
| 97 | ***N. americanus*** | **Undetermined** | **Undetermined** | **Undetermined** | **Undetermined** |
| | ***T.trichiura*** | **Undetermined** | **Undetermined** | **Undetermined** | **Undetermined** |
| | ***A. lumbricoides*** | **Undetermined** | **Undetermined** | **Undetermined** | **Undetermined** |
| 98 | ***N. americanus*** | **Undetermined** | **Undetermined** | **Undetermined** | **Undetermined** |
| | ***T.trichiura*** | **Undetermined** | **Undetermined** | **Undetermined** | **Undetermined** |
| | ***A. lumbricoides*** | **Undetermined** | **Undetermined** | **Undetermined** | **Undetermined** |
| 99 | ***N. americanus*** | **Undetermined** | **Undetermined** | **Undetermined** | **Undetermined** |
| | ***T.trichiura*** | **Undetermined** | **Undetermined** | **Undetermined** | **Undetermined** |
| | ***A. lumbricoides*** | **Undetermined** | **Undetermined** | **Undetermined** | **Undetermined** |
| 100 | ***N. americanus*** | **Undetermined** | **Undetermined** | **Undetermined** | **Undetermined** |
| | ***T.trichiura*** | **Undetermined** | **Undetermined** | **Undetermined** | **Undetermined** |
| | ***A. lumbricoides*** | **Undetermined** | **Undetermined** | **Undetermined** | **Undetermined** |
| 101 | ***N. americanus*** | **Undetermined** | **Undetermined** | **Undetermined** | **Undetermined** |
| | ***T.trichiura*** | **Undetermined** | **Undetermined** | **Undetermined** | **Undetermined** |
| | ***A. lumbricoides*** | **Undetermined** | **Undetermined** | **Undetermined** | **Undetermined** |
| 102 | ***N. americanus*** | **Undetermined** | **Undetermined** | **Undetermined** | **Undetermined** |
| | ***T.trichiura*** | **Undetermined** | **Undetermined** | **Undetermined** | **Undetermined** |
| | ***A. lumbricoides*** | **Undetermined** | **Undetermined** | **Undetermined** | **Undetermined** |
| 103 | ***N. americanus*** | **Undetermined** | **Undetermined** | **Undetermined** | **Undetermined** |
| | ***T.trichiura*** | **Undetermined** | **Undetermined** | **Undetermined** | **Undetermined** |
| | ***A. lumbricoides*** | **Undetermined** | **Undetermined** | **Undetermined** | **Undetermined** |
| 104 | ***N. americanus*** | **Undetermined** | **Undetermined** | **Undetermined** | **Undetermined** |
| | ***T.trichiura*** | **Undetermined** | **Undetermined** | **Undetermined** | **Undetermined** |
| | ***A. lumbricoides*** | **Undetermined** | **Undetermined** | **Undetermined** | **Undetermined** |

| | | | | | |
|---|---|---|---|---|---|
| **105** | ***N. americanus*** | **Undetermined** | **Undetermined** | **Undetermined** | **Undetermined** |
| | ***T.trichiura*** | **Undetermined** | **Undetermined** | **Undetermined** | **Undetermined** |
| | ***A. lumbricoides*** | **Undetermined** | **Undetermined** | **Undetermined** | **Undetermined** |
| **106** | ***N. americanus*** | **Undetermined** | **Undetermined** | **Undetermined** | **Undetermined** |
| | ***T.trichiura*** | **Undetermined** | **Undetermined** | **Undetermined** | **Undetermined** |
| | ***A. lumbricoides*** | **Undetermined** | **Undetermined** | **Undetermined** | **Undetermined** |
| **107** | ***N. americanus*** | **Undetermined** | **Undetermined** | **Undetermined** | **Undetermined** |
| | ***T.trichiura*** | **Undetermined** | **Undetermined** | **Undetermined** | **Undetermined** |
| | ***A. lumbricoides*** | **Undetermined** | **Undetermined** | **Undetermined** | **Undetermined** |
| **108** | ***N. americanus*** | **Undetermined** | **Undetermined** | **Undetermined** | **Undetermined** |
| | ***T.trichiura*** | **Undetermined** | **Undetermined** | **Undetermined** | **Undetermined** |
| | ***A. lumbricoides*** | **Undetermined** | **Undetermined** | **Undetermined** | **Undetermined** |
| **109** | ***N. americanus*** | **Undetermined** | **Undetermined** | **Undetermined** | **Undetermined** |
| | ***T.trichiura*** | **Undetermined** | **Undetermined** | **Undetermined** | **Undetermined** |
| | ***A. lumbricoides*** | **Undetermined** | **Undetermined** | **Undetermined** | **Undetermined** |
| **110** | ***N. americanus*** | **Undetermined** | **Undetermined** | **Undetermined** | **Undetermined** |
| | ***T.trichiura*** | **Undetermined** | **Undetermined** | **Undetermined** | **Undetermined** |
| | ***A. lumbricoides*** | **Undetermined** | **Undetermined** | **Undetermined** | **Undetermined** |
| **111** | ***N. americanus*** | **Undetermined** | **Undetermined** | **Undetermined** | **Undetermined** |
| | ***T.trichiura*** | **Undetermined** | **Undetermined** | **Undetermined** | **Undetermined** |
| | ***A. lumbricoides*** | **Undetermined** | **Undetermined** | **Undetermined** | **Undetermined** |
| **112** | ***N. americanus*** | **Undetermined** | **Undetermined** | **Undetermined** | **Undetermined** |
| | ***T.trichiura*** | **Undetermined** | **Undetermined** | **Undetermined** | **Undetermined** |
| | ***A. lumbricoides*** | **Undetermined** | **Undetermined** | **Undetermined** | **Undetermined** |
| **113** | ***N. americanus*** | **Undetermined** | **Undetermined** | **Undetermined** | **Undetermined** |
| | ***T.trichiura*** | **Undetermined** | **Undetermined** | **Undetermined** | **Undetermined** |
| | ***A. lumbricoides*** | **Undetermined** | **Undetermined** | **Undetermined** | **Undetermined** |
| **114** | ***N. americanus*** | **Undetermined** | **Undetermined** | **Undetermined** | **Undetermined** |
| | ***T.trichiura*** | **Undetermined** | **Undetermined** | **Undetermined** | **Undetermined** |
| | ***A. lumbricoides*** | **Undetermined** | **Undetermined** | **Undetermined** | **Undetermined** |
| **115** | ***N. americanus*** | **Undetermined** | **Undetermined** | **Undetermined** | **Undetermined** |
| | ***T.trichiura*** | **Undetermined** | **Undetermined** | **Undetermined** | **Undetermined** |
| | ***A. lumbricoides*** | **Undetermined** | **Undetermined** | **Undetermined** | **Undetermined** |
| **116** | ***N. americanus*** | **Undetermined** | **Undetermined** | **Undetermined** | **Undetermined** |
| | ***T.trichiura*** | **Undetermined** | **Undetermined** | **Undetermined** | **Undetermined** |
| | ***A. lumbricoides*** | **Undetermined** | **Undetermined** | **Undetermined** | **Undetermined** |
| **117** | ***N. americanus*** | **Undetermined** | **Undetermined** | **Undetermined** | **Undetermined** |
| | ***T.trichiura*** | **Undetermined** | **Undetermined** | **Undetermined** | **Undetermined** |
| | ***A. lumbricoides*** | **Undetermined** | **Undetermined** | **Undetermined** | **Undetermined** |

| | | | | | |
|---|---|---|---|---|---|
| **118** | ***N. americanus*** | **Undetermined** | **Undetermined** | **Undetermined** | **Undetermined** |
| | ***T.trichiura*** | **Undetermined** | **Undetermined** | **Undetermined** | **Undetermined** |
| | ***A. lumbricoides*** | **Undetermined** | **Undetermined** | **Undetermined** | **Undetermined** |
| **119** | ***N. americanus*** | **Undetermined** | **Undetermined** | **Undetermined** | **Undetermined** |
| | ***T.trichiura*** | **Undetermined** | **Undetermined** | **Undetermined** | **Undetermined** |
| | ***A. lumbricoides*** | **Undetermined** | **Undetermined** | **Undetermined** | **Undetermined** |
| **120** | ***N. americanus*** | **Undetermined** | **Undetermined** | **Undetermined** | **Undetermined** |
| | ***T.trichiura*** | **Undetermined** | **Undetermined** | **Undetermined** | **Undetermined** |
| | ***A. lumbricoides*** | **Undetermined** | **Undetermined** | **Undetermined** | **Undetermined** |
| **121** | ***N. americanus*** | **Undetermined** | **Undetermined** | **Undetermined** | **Undetermined** |
| | ***T.trichiura*** | **Undetermined** | **Undetermined** | **Undetermined** | **Undetermined** |
| | ***A. lumbricoides*** | **Undetermined** | **Undetermined** | **Undetermined** | **Undetermined** |
| **122** | ***N. americanus*** | **Undetermined** | **Undetermined** | **Undetermined** | **Undetermined** |
| | ***T.trichiura*** | **Undetermined** | **Undetermined** | **Undetermined** | **Undetermined** |
| | ***A. lumbricoides*** | **Undetermined** | **Undetermined** | **Undetermined** | **Undetermined** |
| **123** | ***N. americanus*** | **Undetermined** | **Undetermined** | **Undetermined** | **Undetermined** |
| | ***T.trichiura*** | **Undetermined** | **Undetermined** | **Undetermined** | **Undetermined** |
| | ***A. lumbricoides*** | **Undetermined** | **Undetermined** | **Undetermined** | **Undetermined** |
| **124** | ***N. americanus*** | **Undetermined** | **Undetermined** | **Undetermined** | **Undetermined** |
| | ***T.trichiura*** | **Undetermined** | **Undetermined** | **Undetermined** | **Undetermined** |
| | ***A. lumbricoides*** | **Undetermined** | **Undetermined** | **Undetermined** | **Undetermined** |
| **125** | ***N. americanus*** | **Undetermined** | **Undetermined** | **Undetermined** | **Undetermined** |
| | ***T.trichiura*** | **Undetermined** | **Undetermined** | **Undetermined** | **Undetermined** |
| | ***A. lumbricoides*** | **Undetermined** | **Undetermined** | **Undetermined** | **Undetermined** |
| **126** | ***N. americanus*** | **Undetermined** | **Undetermined** | **Undetermined** | **Undetermined** |
| | ***T.trichiura*** | **Undetermined** | **Undetermined** | **Undetermined** | **Undetermined** |
| | ***A. lumbricoides*** | **Undetermined** | **Undetermined** | **Undetermined** | **Undetermined** |
| **127** | ***N. americanus*** | **Undetermined** | **Undetermined** | **Undetermined** | **Undetermined** |
| | ***T.trichiura*** | **Undetermined** | **Undetermined** | **Undetermined** | **Undetermined** |
| | ***A. lumbricoides*** | **Undetermined** | **Undetermined** | **Undetermined** | **Undetermined** |
| **128** | ***N. americanus*** | **Undetermined** | **Undetermined** | **Undetermined** | **Undetermined** |
| | ***T.trichiura*** | **Undetermined** | **Undetermined** | **Undetermined** | **Undetermined** |
| | ***A. lumbricoides*** | **Undetermined** | **Undetermined** | **Undetermined** | **Undetermined** |
| **129** | ***N. americanus*** | **Undetermined** | **Undetermined** | **Undetermined** | **Undetermined** |
| | ***T.trichiura*** | **Undetermined** | **Undetermined** | **Undetermined** | **Undetermined** |
| | ***A. lumbricoides*** | **Undetermined** | **Undetermined** | **Undetermined** | **Undetermined** |
| **130** | ***N. americanus*** | **Undetermined** | **Undetermined** | **Undetermined** | **Undetermined** |
| | ***T.trichiura*** | **Undetermined** | **Undetermined** | **Undetermined** | **Undetermined** |
| | ***A. lumbricoides*** | **Undetermined** | **Undetermined** | **Undetermined** | **Undetermined** |

| | | | | | |
|---|---|---|---|---|---|
| **131** | ***N. americanus*** | **Undetermined** | **Undetermined** | **Undetermined** | **Undetermined** |
| | ***T.trichiura*** | **Undetermined** | **Undetermined** | **Undetermined** | **Undetermined** |
| | ***A. lumbricoides*** | **Undetermined** | **Undetermined** | **Undetermined** | **Undetermined** |
| **132** | ***N. americanus*** | **Undetermined** | **Undetermined** | **Undetermined** | **Undetermined** |
| | ***T.trichiura*** | **Undetermined** | **Undetermined** | **Undetermined** | **Undetermined** |
| | ***A. lumbricoides*** | **Undetermined** | **Undetermined** | **Undetermined** | **Undetermined** |
| **133** | ***N. americanus*** | **Undetermined** | **Undetermined** | **Undetermined** | **Undetermined** |
| | ***T.trichiura*** | **Undetermined** | **Undetermined** | **Undetermined** | **Undetermined** |
| | ***A. lumbricoides*** | **Undetermined** | **Undetermined** | **Undetermined** | **Undetermined** |
| **134** | ***N. americanus*** | **Undetermined** | **Undetermined** | **Undetermined** | **Undetermined** |
| | ***T.trichiura*** | **Undetermined** | **Undetermined** | **Undetermined** | **Undetermined** |
| | ***A. lumbricoides*** | **Undetermined** | **Undetermined** | **Undetermined** | **Undetermined** |
| **135** | ***N. americanus*** | **Undetermined** | **Undetermined** | **Undetermined** | **Undetermined** |
| | ***T.trichiura*** | **Undetermined** | **Undetermined** | **Undetermined** | **Undetermined** |
| | ***A. lumbricoides*** | **Undetermined** | **Undetermined** | **Undetermined** | **Undetermined** |
| **136** | ***N. americanus*** | **Undetermined** | **Undetermined** | **Undetermined** | **Undetermined** |
| | ***T.trichiura*** | **Undetermined** | **Undetermined** | **Undetermined** | **Undetermined** |
| | ***A. lumbricoides*** | **Undetermined** | **Undetermined** | **Undetermined** | **Undetermined** |
| **137** | ***N. americanus*** | **Undetermined** | **Undetermined** | **Undetermined** | **Undetermined** |
| | ***T.trichiura*** | **Undetermined** | **Undetermined** | **Undetermined** | **Undetermined** |
| | ***A. lumbricoides*** | **Undetermined** | **Undetermined** | **Undetermined** | **Undetermined** |
| **138** | ***N. americanus*** | **Undetermined** | **Undetermined** | **Undetermined** | **Undetermined** |
| | ***T.trichiura*** | **Undetermined** | **Undetermined** | **Undetermined** | **Undetermined** |
| | ***A. lumbricoides*** | **Undetermined** | **Undetermined** | **Undetermined** | **Undetermined** |
| **139** | ***N. americanus*** | **Undetermined** | **Undetermined** | **Undetermined** | **Undetermined** |
| | ***T.trichiura*** | **Undetermined** | **Undetermined** | **Undetermined** | **Undetermined** |
| | ***A. lumbricoides*** | **Undetermined** | **Undetermined** | **Undetermined** | **Undetermined** |
| **140** | ***N. americanus*** | **Undetermined** | **Undetermined** | **Undetermined** | **Undetermined** |
| | ***T.trichiura*** | **Undetermined** | **Undetermined** | **Undetermined** | **Undetermined** |
| | ***A. lumbricoides*** | **Undetermined** | **Undetermined** | **Undetermined** | **Undetermined** |
| **141** | ***N. americanus*** | **Undetermined** | **Undetermined** | **Undetermined** | **Undetermined** |
| | ***T.trichiura*** | **Undetermined** | **Undetermined** | **Undetermined** | **Undetermined** |
| | ***A. lumbricoides*** | **Undetermined** | **Undetermined** | **Undetermined** | **Undetermined** |
| **142** | ***N. americanus*** | **Undetermined** | **Undetermined** | **Undetermined** | **Undetermined** |
| | ***T.trichiura*** | **Undetermined** | **Undetermined** | **Undetermined** | **Undetermined** |
| | ***A. lumbricoides*** | **Undetermined** | **Undetermined** | **Undetermined** | **Undetermined** |
| **143** | ***N. americanus*** | **Undetermined** | **Undetermined** | **Undetermined** | **Undetermined** |
| | ***T.trichiura*** | **Undetermined** | **Undetermined** | **Undetermined** | **Undetermined** |
| | ***A. lumbricoides*** | **Undetermined** | **Undetermined** | **Undetermined** | **Undetermined** |

| | | | | | |
|---|---|---|---|---|---|
| **144** | ***N. americanus*** | **Undetermined** | **Undetermined** | **Undetermined** | **Undetermined** |
| | ***T.trichiura*** | **Undetermined** | **Undetermined** | **Undetermined** | **Undetermined** |
| | ***A. lumbricoides*** | **Undetermined** | **Undetermined** | **Undetermined** | **Undetermined** |
| **145** | ***N. americanus*** | **Undetermined** | **Undetermined** | **Undetermined** | **Undetermined** |
| | ***T.trichiura*** | **Undetermined** | **Undetermined** | **Undetermined** | **Undetermined** |
| | ***A. lumbricoides*** | **Undetermined** | **Undetermined** | **Undetermined** | **Undetermined** |
| **146** | ***N. americanus*** | **Undetermined** | **Undetermined** | **Undetermined** | **Undetermined** |
| | ***T.trichiura*** | **Undetermined** | **Undetermined** | **Undetermined** | **Undetermined** |
| | ***A. lumbricoides*** | **Undetermined** | **Undetermined** | **Undetermined** | **Undetermined** |
| **147** | ***N. americanus*** | **Undetermined** | **Undetermined** | **Undetermined** | **Undetermined** |
| | ***T.trichiura*** | **Undetermined** | **Undetermined** | **Undetermined** | **Undetermined** |
| | ***A. lumbricoides*** | **Undetermined** | **Undetermined** | **Undetermined** | **Undetermined** |
| **148** | ***N. americanus*** | **Undetermined** | **Undetermined** | **Undetermined** | **Undetermined** |
| | ***T.trichiura*** | **Undetermined** | **Undetermined** | **Undetermined** | **Undetermined** |
| | ***A. lumbricoides*** | **Undetermined** | **Undetermined** | **Undetermined** | **Undetermined** |
| **149** | ***N. americanus*** | **Undetermined** | **Undetermined** | **Undetermined** | **Undetermined** |
| | ***T.trichiura*** | **Undetermined** | **Undetermined** | **Undetermined** | **Undetermined** |
| | ***A. lumbricoides*** | **Undetermined** | **Undetermined** | **Undetermined** | **Undetermined** |
| **150** | ***N. americanus*** | **Undetermined** | **Undetermined** | **Undetermined** | **Undetermined** |
| | ***T.trichiura*** | **Undetermined** | **Undetermined** | **Undetermined** | **Undetermined** |
| | ***A. lumbricoides*** | **Undetermined** | **Undetermined** | **Undetermined** | **Undetermined** |
| **151** | ***N. americanus*** | **Undetermined** | **Undetermined** | **Undetermined** | **Undetermined** |
| | ***T.trichiura*** | **Undetermined** | **Undetermined** | **Undetermined** | **Undetermined** |
| | ***A. lumbricoides*** | **Undetermined** | **Undetermined** | **Undetermined** | **Undetermined** |
| **152** | ***N. americanus*** | **Undetermined** | **Undetermined** | **Undetermined** | **Undetermined** |
| | ***T.trichiura*** | **Undetermined** | **Undetermined** | **Undetermined** | **Undetermined** |
| | ***A. lumbricoides*** | **Undetermined** | **Undetermined** | **Undetermined** | **Undetermined** |
| **153** | ***N. americanus*** | **Undetermined** | **Undetermined** | **Undetermined** | **Undetermined** |
| | ***T.trichiura*** | **Undetermined** | **Undetermined** | **Undetermined** | **Undetermined** |
| | ***A. lumbricoides*** | **Undetermined** | **Undetermined** | **Undetermined** | **Undetermined** |
| **154** | ***N. americanus*** | **Undetermined** | **Undetermined** | **Undetermined** | **Undetermined** |
| | ***T.trichiura*** | **Undetermined** | **Undetermined** | **Undetermined** | **Undetermined** |
| | ***A. lumbricoides*** | **Undetermined** | **Undetermined** | **Undetermined** | **Undetermined** |
| **155** | ***N. americanus*** | **Undetermined** | **Undetermined** | **Undetermined** | **Undetermined** |
| | ***T.trichiura*** | **Undetermined** | **Undetermined** | **Undetermined** | **Undetermined** |
| | ***A. lumbricoides*** | **Undetermined** | **Undetermined** | **Undetermined** | **Undetermined** |
| **156** | ***N. americanus*** | **Undetermined** | **Undetermined** | **Undetermined** | **Undetermined** |
| | ***T.trichiura*** | **Undetermined** | **Undetermined** | **Undetermined** | **Undetermined** |
| | ***A. lumbricoides*** | **Undetermined** | **Undetermined** | **Undetermined** | **Undetermined** |

| | | | | | |
|---|---|---|---|---|---|
| **157** | ***N. americanus*** | **Undetermined** | **Undetermined** | **Undetermined** | **Undetermined** |
| | ***T.trichiura*** | **Undetermined** | **Undetermined** | **Undetermined** | **Undetermined** |
| | ***A. lumbricoides*** | **Undetermined** | **Undetermined** | **Undetermined** | **Undetermined** |
| **158** | ***N. americanus*** | **Undetermined** | **Undetermined** | **Undetermined** | **Undetermined** |
| | ***T.trichiura*** | **Undetermined** | **Undetermined** | **Undetermined** | **Undetermined** |
| | ***A. lumbricoides*** | **Undetermined** | **Undetermined** | **Undetermined** | **Undetermined** |
| **159** | ***N. americanus*** | **Undetermined** | **Undetermined** | **Undetermined** | **Undetermined** |
| | ***T.trichiura*** | **Undetermined** | **Undetermined** | **Undetermined** | **Undetermined** |
| | ***A. lumbricoides*** | **Undetermined** | **Undetermined** | **Undetermined** | **Undetermined** |
| **160** | ***N. americanus*** | **Undetermined** | **Undetermined** | **Undetermined** | **Undetermined** |
| | ***T.trichiura*** | **Undetermined** | **Undetermined** | **Undetermined** | **Undetermined** |
| | ***A. lumbricoides*** | **Undetermined** | **Undetermined** | **Undetermined** | **Undetermined** |
| **161** | ***N. americanus*** | **Undetermined** | **Undetermined** | **Undetermined** | **Undetermined** |
| | ***T.trichiura*** | **Undetermined** | **Undetermined** | **Undetermined** | **Undetermined** |
| | ***A. lumbricoides*** | **Undetermined** | **Undetermined** | **Undetermined** | **Undetermined** |
| **162** | ***N. americanus*** | **Undetermined** | **Undetermined** | **Undetermined** | **Undetermined** |
| | ***T.trichiura*** | **Undetermined** | **Undetermined** | **Undetermined** | **Undetermined** |
| | ***A. lumbricoides*** | **Undetermined** | **Undetermined** | **Undetermined** | **Undetermined** |
| **163** | ***N. americanus*** | **Undetermined** | **Undetermined** | **Undetermined** | **Undetermined** |
| | ***T.trichiura*** | **Undetermined** | **Undetermined** | **Undetermined** | **Undetermined** |
| | ***A. lumbricoides*** | **Undetermined** | **Undetermined** | **Undetermined** | **Undetermined** |
| **164** | ***N. americanus*** | **Undetermined** | **Undetermined** | **Undetermined** | **Undetermined** |
| | ***T.trichiura*** | **Undetermined** | **Undetermined** | **Undetermined** | **Undetermined** |
| | ***A. lumbricoides*** | **Undetermined** | **Undetermined** | **Undetermined** | **Undetermined** |
| **165** | ***N. americanus*** | **Undetermined** | **Undetermined** | **Undetermined** | **Undetermined** |
| | ***T.trichiura*** | **Undetermined** | **Undetermined** | **Undetermined** | **Undetermined** |
| | ***A. lumbricoides*** | **Undetermined** | **Undetermined** | **Undetermined** | **Undetermined** |
| **166** | ***N. americanus*** | **Undetermined** | **Undetermined** | **Undetermined** | **Undetermined** |
| | ***T.trichiura*** | **Undetermined** | **Undetermined** | **Undetermined** | **Undetermined** |
| | ***A. lumbricoides*** | **Undetermined** | **Undetermined** | **Undetermined** | **Undetermined** |
| **167** | ***N. americanus*** | **Undetermined** | **Undetermined** | **Undetermined** | **Undetermined** |
| | ***T.trichiura*** | **Undetermined** | **Undetermined** | **Undetermined** | **Undetermined** |
| | ***A. lumbricoides*** | **Undetermined** | **Undetermined** | **Undetermined** | **Undetermined** |
| **168** | ***N. americanus*** | **Undetermined** | **Undetermined** | **Undetermined** | **Undetermined** |
| | ***T.trichiura*** | **Undetermined** | **Undetermined** | **Undetermined** | **Undetermined** |
| | ***A. lumbricoides*** | **Undetermined** | **Undetermined** | **Undetermined** | **Undetermined** |
| **169** | ***N. americanus*** | **Undetermined** | **Undetermined** | **Undetermined** | **Undetermined** |
| | ***T.trichiura*** | **Undetermined** | **Undetermined** | **Undetermined** | **Undetermined** |
| | ***A. lumbricoides*** | **Undetermined** | **Undetermined** | **Undetermined** | **Undetermined** |

| | | | | | |
|---|---|---|---|---|---|
| **170** | ***N. americanus*** | **Undetermined** | **Undetermined** | **Undetermined** | **Undetermined** |
| | ***T.trichiura*** | **Undetermined** | **Undetermined** | **Undetermined** | **Undetermined** |
| | ***A. lumbricoides*** | **Undetermined** | **Undetermined** | **Undetermined** | **Undetermined** |
| **171** | ***N. americanus*** | **Undetermined** | **Undetermined** | **Undetermined** | **Undetermined** |
| | ***T.trichiura*** | **Undetermined** | **Undetermined** | **Undetermined** | **Undetermined** |
| | ***A. lumbricoides*** | **Undetermined** | **Undetermined** | **Undetermined** | **Undetermined** |
| **172** | ***N. americanus*** | **Undetermined** | **Undetermined** | **Undetermined** | **Undetermined** |
| | ***T.trichiura*** | **Undetermined** | **Undetermined** | **Undetermined** | **Undetermined** |
| | ***A. lumbricoides*** | **Undetermined** | **Undetermined** | **Undetermined** | **Undetermined** |
| **173** | ***N. americanus*** | **Undetermined** | **Undetermined** | **Undetermined** | **Undetermined** |
| | ***T.trichiura*** | **Undetermined** | **Undetermined** | **Undetermined** | **Undetermined** |
| | ***A. lumbricoides*** | **Undetermined** | **Undetermined** | **Undetermined** | **Undetermined** |
| **174** | ***N. americanus*** | **Undetermined** | **Undetermined** | **Undetermined** | **Undetermined** |
| | ***T.trichiura*** | **Undetermined** | **Undetermined** | **Undetermined** | **Undetermined** |
| | ***A. lumbricoides*** | **Undetermined** | **Undetermined** | **Undetermined** | **Undetermined** |
| **175** | ***N. americanus*** | **Undetermined** | **Undetermined** | **Undetermined** | **Undetermined** |
| | ***T.trichiura*** | **Undetermined** | **Undetermined** | **Undetermined** | **Undetermined** |
| | ***A. lumbricoides*** | **Undetermined** | **Undetermined** | **Undetermined** | **Undetermined** |
| **176** | ***N. americanus*** | **Undetermined** | **Undetermined** | **Undetermined** | **Undetermined** |
| | ***T.trichiura*** | **Undetermined** | **Undetermined** | **Undetermined** | **Undetermined** |
| | ***A. lumbricoides*** | **Undetermined** | **Undetermined** | **Undetermined** | **Undetermined** |
| **177** | ***N. americanus*** | **Undetermined** | **Undetermined** | **Undetermined** | **Undetermined** |
| | ***T.trichiura*** | **Undetermined** | **Undetermined** | **Undetermined** | **Undetermined** |
| | ***A. lumbricoides*** | **Undetermined** | **Undetermined** | **Undetermined** | **Undetermined** |
| **178** | ***N. americanus*** | **Undetermined** | **Undetermined** | **Undetermined** | **Undetermined** |
| | ***T.trichiura*** | **Undetermined** | **Undetermined** | **Undetermined** | **Undetermined** |
| | ***A. lumbricoides*** | **Undetermined** | **Undetermined** | **Undetermined** | **Undetermined** |
| **179** | ***N. americanus*** | **Undetermined** | **Undetermined** | **Undetermined** | **Undetermined** |
| | ***T.trichiura*** | **Undetermined** | **Undetermined** | **Undetermined** | **Undetermined** |
| | ***A. lumbricoides*** | **Undetermined** | **Undetermined** | **Undetermined** | **Undetermined** |
| **180** | ***N. americanus*** | **Undetermined** | **Undetermined** | **Undetermined** | **Undetermined** |
| | ***T.trichiura*** | **Undetermined** | **Undetermined** | **Undetermined** | **Undetermined** |
| | ***A. lumbricoides*** | **Undetermined** | **Undetermined** | **Undetermined** | **Undetermined** |
| **181** | ***N. americanus*** | **Undetermined** | **Undetermined** | **Undetermined** | **Undetermined** |
| | ***T.trichiura*** | **Undetermined** | **Undetermined** | **Undetermined** | **Undetermined** |
| | ***A. lumbricoides*** | **Undetermined** | **Undetermined** | **Undetermined** | **Undetermined** |
| **182** | ***N. americanus*** | **Undetermined** | **Undetermined** | **Undetermined** | **Undetermined** |
| | ***T.trichiura*** | **Undetermined** | **Undetermined** | **Undetermined** | **Undetermined** |
| | ***A. lumbricoides*** | **Undetermined** | **Undetermined** | **Undetermined** | **Undetermined** |

| | | | | | |
|---|---|---|---|---|---|
| **183** | ***N. americanus*** | **Undetermined** | **Undetermined** | **Undetermined** | **Undetermined** |
| | ***T.trichiura*** | **Undetermined** | **Undetermined** | **Undetermined** | **Undetermined** |
| | ***A. lumbricoides*** | **Undetermined** | **Undetermined** | **Undetermined** | **Undetermined** |
| **184** | ***N. americanus*** | **Undetermined** | **Undetermined** | **Undetermined** | **Undetermined** |
| | ***T.trichiura*** | **Undetermined** | **Undetermined** | **Undetermined** | **Undetermined** |
| | ***A. lumbricoides*** | **Undetermined** | **Undetermined** | **Undetermined** | **Undetermined** |
| **185** | ***N. americanus*** | **Undetermined** | **Undetermined** | **Undetermined** | **Undetermined** |
| | ***T.trichiura*** | **Undetermined** | **Undetermined** | **Undetermined** | **Undetermined** |
| | ***A. lumbricoides*** | **Undetermined** | **Undetermined** | **Undetermined** | **Undetermined** |
| **186** | ***N. americanus*** | **Undetermined** | **Undetermined** | **Undetermined** | **Undetermined** |
| | ***T.trichiura*** | **Undetermined** | **Undetermined** | **Undetermined** | **Undetermined** |
| | ***A. lumbricoides*** | **Undetermined** | **Undetermined** | **Undetermined** | **Undetermined** |
| **187** | ***N. americanus*** | **Undetermined** | **Undetermined** | **Undetermined** | **Undetermined** |
| | ***T.trichiura*** | **Undetermined** | **Undetermined** | **Undetermined** | **Undetermined** |
| | ***A. lumbricoides*** | **Undetermined** | **Undetermined** | **Undetermined** | **Undetermined** |
| **188** | ***N. americanus*** | **Undetermined** | **Undetermined** | **Undetermined** | **Undetermined** |
| | ***T.trichiura*** | **Undetermined** | **Undetermined** | **Undetermined** | **Undetermined** |
| | ***A. lumbricoides*** | **Undetermined** | **Undetermined** | **Undetermined** | **Undetermined** |
| **189** | ***N. americanus*** | **Undetermined** | **Undetermined** | **Undetermined** | **Undetermined** |
| | ***T.trichiura*** | **Undetermined** | **Undetermined** | **Undetermined** | **Undetermined** |
| | ***A. lumbricoides*** | **Undetermined** | **Undetermined** | **Undetermined** | **Undetermined** |
| **190** | ***N. americanus*** | **Undetermined** | **Undetermined** | **Undetermined** | **Undetermined** |
| | ***T.trichiura*** | **Undetermined** | **Undetermined** | **Undetermined** | **Undetermined** |
| | ***A. lumbricoides*** | **Undetermined** | **Undetermined** | **Undetermined** | **Undetermined** |
| **191** | ***N. americanus*** | **Undetermined** | **Undetermined** | **Undetermined** | **Undetermined** |
| | ***T.trichiura*** | **Undetermined** | **Undetermined** | **Undetermined** | **Undetermined** |
| | ***A. lumbricoides*** | **Undetermined** | **Undetermined** | **Undetermined** | **Undetermined** |
| **192** | ***N. americanus*** | **Undetermined** | **Undetermined** | **Undetermined** | **Undetermined** |
| | ***T.trichiura*** | **Undetermined** | **Undetermined** | **Undetermined** | **Undetermined** |
| | ***A. lumbricoides*** | **Undetermined** | **Undetermined** | **Undetermined** | **Undetermined** |
| **193** | ***N. americanus*** | **Undetermined** | **Undetermined** | **Undetermined** | **Undetermined** |
| | ***T.trichiura*** | **Undetermined** | **Undetermined** | **Undetermined** | **Undetermined** |
| | ***A. lumbricoides*** | **Undetermined** | **Undetermined** | **Undetermined** | **Undetermined** |
| **194** | ***N. americanus*** | **Undetermined** | **Undetermined** | **Undetermined** | **Undetermined** |
| | ***T.trichiura*** | **Undetermined** | **Undetermined** | **Undetermined** | **Undetermined** |
| | ***A. lumbricoides*** | **Undetermined** | **Undetermined** | **Undetermined** | **Undetermined** |
| **195** | ***N. americanus*** | **Undetermined** | **Undetermined** | **Undetermined** | **Undetermined** |
| | ***T.trichiura*** | **Undetermined** | **Undetermined** | **Undetermined** | **Undetermined** |
| | ***A. lumbricoides*** | **Undetermined** | **Undetermined** | **Undetermined** | **Undetermined** |

| | | | | | |
|---|---|---|---|---|---|
| **196** | ***N. americanus*** | **Undetermined** | **Undetermined** | **Undetermined** | **Undetermined** |
| | ***T.trichiura*** | **Undetermined** | **Undetermined** | **Undetermined** | **Undetermined** |
| | ***A. lumbricoides*** | **Undetermined** | **Undetermined** | **Undetermined** | **Undetermined** |
| **197** | ***N. americanus*** | **Undetermined** | **Undetermined** | **Undetermined** | **Undetermined** |
| | ***T.trichiura*** | **Undetermined** | **Undetermined** | **Undetermined** | **Undetermined** |
| | ***A. lumbricoides*** | **Undetermined** | **Undetermined** | **Undetermined** | **Undetermined** |
| **198** | ***N. americanus*** | **Undetermined** | **Undetermined** | **Undetermined** | **Undetermined** |
| | ***T.trichiura*** | **Undetermined** | **Undetermined** | **Undetermined** | **Undetermined** |
| | ***A. lumbricoides*** | **Undetermined** | **Undetermined** | **Undetermined** | **Undetermined** |
| **199** | ***N. americanus*** | **Undetermined** | **Undetermined** | **Undetermined** | **Undetermined** |
| | ***T.trichiura*** | **Undetermined** | **Undetermined** | **Undetermined** | **Undetermined** |
| | ***A. lumbricoides*** | **Undetermined** | **Undetermined** | **Undetermined** | **Undetermined** |
| **200** | ***N. americanus*** | **Undetermined** | **Undetermined** | **Undetermined** | **Undetermined** |
| | ***T.trichiura*** | **Undetermined** | **Undetermined** | **Undetermined** | **Undetermined** |
| | ***A. lumbricoides*** | **Undetermined** | **Undetermined** | **Undetermined** | **Undetermined** |

**Figure A8.3:** *DNA line data for N. americanus at Quantigen and Smith*

| | Extract 1 | | Extract 2 | | Extract 3 | | Extract 4 | |
|---|---|---|---|---|---|---|---|---|
| Sample ID | Quantigen | Smith | Quantigen | Smith | Smith | Quantigen | Smith | Quantigen |
| 1 | Undetermined | Undetermined | Undetermined | Undetermined | Undetermined | Undetermined | Undetermined | Undetermined |
| 2 | Undetermined | Undetermined | Undetermined | Undetermined | Undetermined | Undetermined | Undetermined | Undetermined |
| 3 | Undetermined | Undetermined | Undetermined | Undetermined | Undetermined | Undetermined | Undetermined | Undetermined |
| 4 | Undetermined | Undetermined | Undetermined | Undetermined | Undetermined | Undetermined | Undetermined | Undetermined |
| 5 | Undetermined | Undetermined | Undetermined | Undetermined | Undetermined | Undetermined | Undetermined | Undetermined |
| 6 | 25.3 | 24.90 | 24.2 | 22.83 | 18.13 | 18.83 | 18.41 | 18.73 |
| 7 | Undetermined | Undetermined | Undetermined | Undetermined | Undetermined | Undetermined | Undetermined | Undetermined |
| 8 | Undetermined | Undetermined | Undetermined | Undetermined | Undetermined | Undetermined | Undetermined | Undetermined |
| 9 | Undetermined | Undetermined | Undetermined | Undetermined | Undetermined | 36.7 | Undetermined | Undetermined |
| 10 | Undetermined | Undetermined | Undetermined | Undetermined | Undetermined | Undetermined | Undetermined | Undetermined |
| 11 | Undetermined | Undetermined | Undetermined | Undetermined | Undetermined | Undetermined | Undetermined | Undetermined |
| 12 | 36.3 | Undetermined | 35.9 | Undetermined | 32.4 | 32.1 | 26.7 | 27.6 |
| 13 | Undetermined | Undetermined | Undetermined | Undetermined | Undetermined | Undetermined | Undetermined | Undetermined |
| 14 | 28.8 | 27.7 | 25.7 | 25.7 | 28.4 | 27.8 | Undetermined | Undetermined |
| 15 | 23.6 | 23.13 | 23.5 | 21.57 | 21.11 | 16.90 | 17.29 | 21.13 |
| 16 | 28.4 | 28.09 | 28.6 | 28.45 | 23.91 | 26.17 | 32.14 | 35.48 |
| 17 | Undetermined | Undetermined | Undetermined | Undetermined | Undetermined | Undetermined | Undetermined | Undetermined |
| 18 | Undetermined | Undetermined | Undetermined | Undetermined | Undetermined | Undetermined | Undetermined | Undetermined |
| 19 | Undetermined | Undetermined | Undetermined | Undetermined | Undetermined | Undetermined | Undetermined | Undetermined |
| 20 | Undetermined | Undetermined | Undetermined | Undetermined | Undetermined | Undetermined | Undetermined | Undetermined |
| 21 | Undetermined | Undetermined | Undetermined | Undetermined | Undetermined | Undetermined | Undetermined | Undetermined |
| 22 | Undetermined | Undetermined | Undetermined | Undetermined | Undetermined | Undetermined | Undetermined | Undetermined |
| 23 | 28.3 | 27.89 | 27.2 | 26.10 | 23.68 | 24.33 | 24.36 | 25.50 |
| 24 | Undetermined | Undetermined | Undetermined | Undetermined | Undetermined | Undetermined | Undetermined | Undetermined |
| 25 | Undetermined | Undetermined | Undetermined | Undetermined | Undetermined | Undetermined | Undetermined | Undetermined |

| | | | | | | | | |
|---|---|---|---|---|---|---|---|---|
| 26 | Undetermined | Undetermined | Undetermined | Undetermined | Undetermined | Undetermined | Undetermined | Undetermined |
| 27 | Undetermined | Undetermined | Undetermined | Undetermined | Undetermined | Undetermined | Undetermined | Undetermined |
| 28 | Undetermined | Undetermined | Undetermined | Undetermined | Undetermined | Undetermined | Undetermined | Undetermined |
| 29 | Undetermined | Undetermined | Undetermined | Undetermined | Undetermined | Undetermined | Undetermined | Undetermined |
| 30 | Undetermined | Undetermined | Undetermined | Undetermined | Undetermined | Undetermined | Undetermined | Undetermined |
| 31 | Undetermined | Undetermined | Undetermined | Undetermined | Undetermined | Undetermined | Undetermined | Undetermined |
| 32 | Undetermined | Undetermined | Undetermined | Undetermined | Undetermined | Undetermined | Undetermined | Undetermined |
| 33 | Undetermined | 32.0 | Undetermined | Undetermined | Undetermined | Undetermined | Undetermined | Undetermined |
| 34 | Undetermined | Undetermined | Undetermined | Undetermined | Undetermined | Undetermined | Undetermined | Undetermined |
| 35 | Undetermined | Undetermined | Undetermined | Undetermined | Undetermined | Undetermined | Undetermined | 36.6 |
| 36 | Undetermined | Undetermined | Undetermined | Undetermined | Undetermined | Undetermined | Undetermined | Undetermined |
| 37 | Undetermined | Undetermined | Undetermined | Undetermined | Undetermined | Undetermined | Undetermined | Undetermined |
| 38 | Undetermined | Undetermined | Undetermined | Undetermined | Undetermined | Undetermined | Undetermined | Undetermined |
| 39 | 26.9 | 25.92 | 28.5 | 26.25 | 20.77 | 24.00 | 22.83 | 25.96 |
| 40 | Undetermined | Undetermined | 32.5 | 30.1 | Undetermined | Undetermined | Undetermined | 29.5 |
| 41 | 36.5 | 35.2 | Undetermined | Undetermined | Undetermined | Undetermined | Undetermined | Undetermined |
| 42 | Undetermined | Undetermined | Undetermined | Undetermined | Undetermined | Undetermined | Undetermined | Undetermined |
| 43 | Undetermined | Undetermined | Undetermined | Undetermined | Undetermined | Undetermined | Undetermined | Undetermined |
| 44 | Undetermined | Undetermined | Undetermined | Undetermined | Undetermined | Undetermined | Undetermined | Undetermined |
| 45 | Undetermined | Undetermined | Undetermined | Undetermined | Undetermined | Undetermined | Undetermined | Undetermined |
| 46 | Undetermined | Undetermined | Undetermined | Undetermined | Undetermined | Undetermined | Undetermined | Undetermined |
| 47 | Undetermined | Undetermined | Undetermined | Undetermined | Undetermined | Undetermined | Undetermined | Undetermined |
| 48 | 36.7 | Undetermined | 36.7 | Undetermined | 36.2 | Undetermined | 29.1 | 30.5 |
| 49 | Undetermined | Undetermined | Undetermined | Undetermined | Undetermined | Undetermined | Undetermined | Undetermined |
| 50 | 33.3 | 32.53 | 34.1 | 32.31 | 30.46 | 31.16 | 29.98 | 31.48 |
| 51 | Undetermined | Undetermined | Undetermined | Undetermined | Undetermined | Undetermined | Undetermined | Undetermined |
| 52 | Undetermined | Undetermined | Undetermined | Undetermined | Undetermined | Undetermined | Undetermined | Undetermined |
| 53 | Undetermined | Undetermined | Undetermined | Undetermined | Undetermined | Undetermined | Undetermined | Undetermined |
| 54 | Undetermined | Undetermined | Undetermined | Undetermined | Undetermined | Undetermined | Undetermined | Undetermined |
| 55 | Undetermined | Undetermined | Undetermined | Undetermined | Undetermined | Undetermined | Undetermined | Undetermined |

| | | | | | | | | |
|---|---|---|---|---|---|---|---|---|
| 56 | Undetermined | Undetermined | Undetermined | Undetermined | Undetermined | Undetermined | Undetermined | Undetermined |
| 57 | Undetermined | Undetermined | Undetermined | Undetermined | Undetermined | Undetermined | Undetermined | Undetermined |
| 58 | Undetermined | Undetermined | Undetermined | Undetermined | Undetermined | Undetermined | Undetermined | Undetermined |
| 59 | Undetermined | Undetermined | Undetermined | Undetermined | Undetermined | Undetermined | Undetermined | Undetermined |
| 60 | 24.7 | 24.90 | 22.9 | 21.26 | 17.30 | 18.24 | 20.57 | 21.22 |
| 61 | Undetermined | Undetermined | Undetermined | Undetermined | Undetermined | Undetermined | Undetermined | Undetermined |
| 62 | Undetermined | Undetermined | Undetermined | Undetermined | Undetermined | Undetermined | Undetermined | Undetermined |
| 63 | Undetermined | Undetermined | Undetermined | Undetermined | Undetermined | Undetermined | Undetermined | Undetermined |
| 64 | Undetermined | Undetermined | Undetermined | Undetermined | Undetermined | Undetermined | Undetermined | Undetermined |
| 65 | Undetermined | Undetermined | Undetermined | Undetermined | Undetermined | Undetermined | Undetermined | Undetermined |
| 66 | Undetermined | Undetermined | Undetermined | Undetermined | Undetermined | Undetermined | Undetermined | Undetermined |
| 67 | Undetermined | Undetermined | Undetermined | Undetermined | Undetermined | Undetermined | Undetermined | Undetermined |
| 68 | 25.1 | 25.03 | 31.0 | 29.63 | 19.69 | 19.32 | 18.99 | 19.94 |
| 69 | Undetermined | Undetermined | Undetermined | Undetermined | Undetermined | Undetermined | Undetermined | Undetermined |
| 70 | Undetermined | Undetermined | Undetermined | Undetermined | Undetermined | Undetermined | Undetermined | Undetermined |
| 71 | Undetermined | Undetermined | Undetermined | Undetermined | Undetermined | Undetermined | Undetermined | Undetermined |
| 72 | Undetermined | Undetermined | Undetermined | Undetermined | Undetermined | Undetermined | Undetermined | Undetermined |
| 73 | Undetermined | Undetermined | Undetermined | Undetermined | Undetermined | Undetermined | Undetermined | Undetermined |
| 74 | Undetermined | Undetermined | Undetermined | Undetermined | Undetermined | Undetermined | Undetermined | Undetermined |
| 75 | Undetermined | Undetermined | Undetermined | Undetermined | Undetermined | Undetermined | Undetermined | Undetermined |
| 76 | Undetermined | Undetermined | Undetermined | Undetermined | Undetermined | Undetermined | Undetermined | Undetermined |
| 77 | Undetermined | Undetermined | Undetermined | Undetermined | Undetermined | Undetermined | Undetermined | Undetermined |
| 78 | Undetermined | Undetermined | Undetermined | Undetermined | Undetermined | Undetermined | Undetermined | Undetermined |
| 79 | Undetermined | Undetermined | Undetermined | Undetermined | Undetermined | Undetermined | Undetermined | Undetermined |
| 80 | Undetermined | Undetermined | Undetermined | Undetermined | Undetermined | Undetermined | Undetermined | Undetermined |
| 81 | Undetermined | Undetermined | Undetermined | Undetermined | Undetermined | Undetermined | Undetermined | Undetermined |
| 82 | Undetermined | Undetermined | Undetermined | Undetermined | Undetermined | Undetermined | Undetermined | Undetermined |
| 83 | Undetermined | Undetermined | Undetermined | Undetermined | Undetermined | Undetermined | Undetermined | Undetermined |
| 84 | Undetermined | Undetermined | Undetermined | Undetermined | Undetermined | Undetermined | Undetermined | Undetermined |
| 85 | Undetermined | Undetermined | Undetermined | Undetermined | Undetermined | Undetermined | Undetermined | Undetermined |

| | | | | | | | | |
|---|---|---|---|---|---|---|---|---|
| 86 | Undetermined | Undetermined | Undetermined | Undetermined | Undetermined | Undetermined | Undetermined | Undetermined |
| 87 | Undetermined | Undetermined | Undetermined | Undetermined | Undetermined | Undetermined | Undetermined | Undetermined |
| 88 | Undetermined | Undetermined | Undetermined | Undetermined | Undetermined | Undetermined | Undetermined | Undetermined |
| 89 | Undetermined | Undetermined | Undetermined | Undetermined | Undetermined | Undetermined | Undetermined | Undetermined |
| 90 | Undetermined | Undetermined | Undetermined | Undetermined | Undetermined | Undetermined | Undetermined | Undetermined |
| 91 | Undetermined | Undetermined | Undetermined | Undetermined | Undetermined | Undetermined | Undetermined | Undetermined |
| 92 | Undetermined | Undetermined | Undetermined | Undetermined | Undetermined | Undetermined | Undetermined | Undetermined |
| 93 | Undetermined | Undetermined | Undetermined | Undetermined | Undetermined | Undetermined | Undetermined | Undetermined |
| 94 | Undetermined | Undetermined | Undetermined | Undetermined | Undetermined | Undetermined | Undetermined | Undetermined |
| 95 | Undetermined | Undetermined | Undetermined | Undetermined | Undetermined | Undetermined | Undetermined | Undetermined |
| 96 | Undetermined | Undetermined | Undetermined | Undetermined | Undetermined | Undetermined | Undetermined | Undetermined |
| 97 | Undetermined | Undetermined | Undetermined | Undetermined | Undetermined | Undetermined | Undetermined | Undetermined |
| 98 | Undetermined | Undetermined | Undetermined | Undetermined | Undetermined | Undetermined | Undetermined | Undetermined |
| 99 | Undetermined | Undetermined | Undetermined | Undetermined | Undetermined | Undetermined | Undetermined | Undetermined |
| 100 | Undetermined | Undetermined | 36.9 | Undetermined | Undetermined | Undetermined | Undetermined | Undetermined |
| 101 | Undetermined | Undetermined | Undetermined | Undetermined | Undetermined | Undetermined | Undetermined | Undetermined |
| 102 | Undetermined | Undetermined | Undetermined | Undetermined | Undetermined | Undetermined | Undetermined | Undetermined |
| 103 | Undetermined | Undetermined | Undetermined | Undetermined | Undetermined | Undetermined | Undetermined | Undetermined |
| 104 | Undetermined | Undetermined | Undetermined | Undetermined | Undetermined | Undetermined | Undetermined | Undetermined |
| 105 | Undetermined | Undetermined | Undetermined | Undetermined | Undetermined | Undetermined | Undetermined | Undetermined |
| 106 | Undetermined | Undetermined | Undetermined | Undetermined | Undetermined | Undetermined | Undetermined | Undetermined |
| 107 | Undetermined | Undetermined | Undetermined | Undetermined | Undetermined | Undetermined | Undetermined | Undetermined |
| 108 | Undetermined | Undetermined | Undetermined | Undetermined | Undetermined | Undetermined | Undetermined | Undetermined |
| 109 | Undetermined | Undetermined | Undetermined | Undetermined | Undetermined | Undetermined | Undetermined | Undetermined |
| 110 | Undetermined | Undetermined | Undetermined | Undetermined | Undetermined | Undetermined | Undetermined | Undetermined |
| 111 | Undetermined | Undetermined | Undetermined | Undetermined | Undetermined | Undetermined | Undetermined | Undetermined |
| 112 | Undetermined | Undetermined | Undetermined | Undetermined | Undetermined | Undetermined | Undetermined | Undetermined |
| 113 | Undetermined | Undetermined | Undetermined | Undetermined | Undetermined | Undetermined | Undetermined | Undetermined |
| 114 | Undetermined | Undetermined | Undetermined | Undetermined | Undetermined | Undetermined | Undetermined | Undetermined |
| 115 | Undetermined | Undetermined | Undetermined | Undetermined | Undetermined | Undetermined | Undetermined | Undetermined |

| | | | | | | | | |
|---|---|---|---|---|---|---|---|---|
| 116 | Undetermined | Undetermined | Undetermined | Undetermined | Undetermined | Undetermined | Undetermined | Undetermined |
| 117 | Undetermined | Undetermined | Undetermined | Undetermined | Undetermined | Undetermined | Undetermined | Undetermined |
| 118 | 31.3 | 30.3 | Undetermined | Undetermined | 23.8 | 25.5 | 30.7 | 33.2 |
| 119 | Undetermined | Undetermined | Undetermined | Undetermined | Undetermined | Undetermined | Undetermined | Undetermined |
| 120 | Undetermined | Undetermined | Undetermined | Undetermined | Undetermined | Undetermined | Undetermined | Undetermined |
| 121 | Undetermined | Undetermined | Undetermined | Undetermined | Undetermined | Undetermined | Undetermined | Undetermined |
| 122 | Undetermined | Undetermined | Undetermined | Undetermined | Undetermined | Undetermined | Undetermined | Undetermined |
| 123 | Undetermined | Undetermined | Undetermined | Undetermined | Undetermined | Undetermined | Undetermined | Undetermined |
| 124 | Undetermined | Undetermined | Undetermined | Undetermined | Undetermined | Undetermined | Undetermined | Undetermined |
| 125 | 22.2 | 21.77 | 23.7 | 22.56 | 17.91 | 19.61 | 18.65 | 20.15 |
| 126 | Undetermined | Undetermined | Undetermined | Undetermined | Undetermined | Undetermined | Undetermined | Undetermined |
| 127 | Undetermined | Undetermined | Undetermined | Undetermined | Undetermined | Undetermined | Undetermined | Undetermined |
| 128 | Undetermined | Undetermined | Undetermined | Undetermined | Undetermined | Undetermined | Undetermined | Undetermined |
| 129 | 27.6 | 27.2 | Undetermined | Undetermined | 18.7 | 20.0 | 20.6 | 18.6 |
| 130 | Undetermined | Undetermined | Undetermined | Undetermined | Undetermined | Undetermined | Undetermined | Undetermined |
| 131 | Undetermined | Undetermined | Undetermined | Undetermined | Undetermined | Undetermined | Undetermined | Undetermined |
| 132 | Undetermined | Undetermined | Undetermined | Undetermined | Undetermined | Undetermined | Undetermined | Undetermined |
| 133 | Undetermined | Undetermined | Undetermined | Undetermined | Undetermined | Undetermined | Undetermined | Undetermined |
| 134 | Undetermined | Undetermined | Undetermined | Undetermined | Undetermined | Undetermined | Undetermined | Undetermined |
| 135 | Undetermined | Undetermined | Undetermined | Undetermined | Undetermined | Undetermined | Undetermined | Undetermined |
| 136 | Undetermined | Undetermined | Undetermined | Undetermined | Undetermined | Undetermined | Undetermined | Undetermined |
| 137 | Undetermined | Undetermined | Undetermined | Undetermined | Undetermined | Undetermined | Undetermined | Undetermined |
| 138 | Undetermined | Undetermined | Undetermined | Undetermined | Undetermined | Undetermined | Undetermined | Undetermined |
| 139 | Undetermined | Undetermined | Undetermined | Undetermined | Undetermined | Undetermined | Undetermined | Undetermined |
| 140 | Undetermined | Undetermined | Undetermined | Undetermined | Undetermined | Undetermined | Undetermined | Undetermined |
| 141 | Undetermined | Undetermined | Undetermined | Undetermined | Undetermined | Undetermined | Undetermined | Undetermined |
| 142 | Undetermined | Undetermined | Undetermined | Undetermined | Undetermined | Undetermined | Undetermined | Undetermined |
| 143 | Undetermined | Undetermined | Undetermined | Undetermined | Undetermined | Undetermined | Undetermined | Undetermined |
| 144 | Undetermined | Undetermined | Undetermined | Undetermined | Undetermined | Undetermined | Undetermined | Undetermined |
| 145 | Undetermined | Undetermined | Undetermined | Undetermined | Undetermined | Undetermined | Undetermined | Undetermined |

| | | | | | | | | |
|---|---|---|---|---|---|---|---|---|
| 146 | Undetermined | Undetermined | Undetermined | Undetermined | Undetermined | Undetermined | Undetermined | Undetermined |
| 147 | Undetermined | Undetermined | Undetermined | Undetermined | Undetermined | Undetermined | Undetermined | Undetermined |
| 148 | Undetermined | Undetermined | Undetermined | Undetermined | 33.2 | Undetermined | Undetermined | Undetermined |
| 149 | Undetermined | Undetermined | Undetermined | Undetermined | Undetermined | Undetermined | Undetermined | Undetermined |
| 150 | Undetermined | Undetermined | Undetermined | Undetermined | Undetermined | Undetermined | Undetermined | Undetermined |
| 151 | Undetermined | Undetermined | Undetermined | Undetermined | Undetermined | Undetermined | Undetermined | Undetermined |
| 152 | Undetermined | Undetermined | Undetermined | Undetermined | Undetermined | Undetermined | Undetermined | Undetermined |
| 153 | 26.2 | 28.80 | 26.1 | 23.87 | 18.93 | 19.29 | 19.07 | 19.22 |
| 154 | Undetermined | Undetermined | Undetermined | Undetermined | Undetermined | Undetermined | Undetermined | Undetermined |
| 155 | Undetermined | Undetermined | Undetermined | Undetermined | Undetermined | Undetermined | Undetermined | Undetermined |
| 156 | Undetermined | Undetermined | Undetermined | Undetermined | Undetermined | Undetermined | Undetermined | Undetermined |
| 157 | Undetermined | Undetermined | Undetermined | Undetermined | Undetermined | Undetermined | Undetermined | Undetermined |
| 158 | Undetermined | Undetermined | Undetermined | Undetermined | Undetermined | Undetermined | Undetermined | Undetermined |
| 159 | Undetermined | Undetermined | Undetermined | Undetermined | Undetermined | Undetermined | Undetermined | Undetermined |
| 160 | Undetermined | Undetermined | Undetermined | Undetermined | Undetermined | Undetermined | Undetermined | Undetermined |
| 161 | Undetermined | Undetermined | Undetermined | Undetermined | Undetermined | Undetermined | Undetermined | Undetermined |
| 162 | Undetermined | Undetermined | Undetermined | Undetermined | Undetermined | Undetermined | Undetermined | Undetermined |
| 163 | Undetermined | Undetermined | Undetermined | Undetermined | Undetermined | Undetermined | Undetermined | Undetermined |
| 164 | Undetermined | Undetermined | Undetermined | Undetermined | Undetermined | Undetermined | Undetermined | Undetermined |
| 165 | Undetermined | Undetermined | Undetermined | Undetermined | Undetermined | Undetermined | Undetermined | Undetermined |
| 166 | Undetermined | Undetermined | Undetermined | Undetermined | Undetermined | Undetermined | Undetermined | Undetermined |
| 167 | 23.4 | 23.87 | 25.9 | 22.86 | 16.65 | 18.19 | 17.80 | 17.34 |
| 168 | 22.8 | 22.13 | 23.7 | 22.72 | 17.79 | 18.63 | 17.40 | 18.13 |
| 169 | 18.5 | 17.41 | 17.8 | 17.71 | 13.89 | 14.58 | 14.34 | 14.95 |
| 170 | Undetermined | Undetermined | Undetermined | Undetermined | Undetermined | Undetermined | Undetermined | Undetermined |
| 171 | Undetermined | Undetermined | Undetermined | Undetermined | Undetermined | Undetermined | Undetermined | Undetermined |
| 172 | Undetermined | Undetermined | Undetermined | Undetermined | Undetermined | Undetermined | Undetermined | Undetermined |
| 173 | Undetermined | Undetermined | Undetermined | Undetermined | Undetermined | Undetermined | Undetermined | Undetermined |
| 174 | Undetermined | Undetermined | Undetermined | Undetermined | Undetermined | Undetermined | Undetermined | Undetermined |
| 175 | Undetermined | Undetermined | Undetermined | Undetermined | Undetermined | Undetermined | Undetermined | Undetermined |

| | | | | | | | | |
|---|---|---|---|---|---|---|---|---|
| 176 | 27.4 | 27.29 | 25.6 | 25.16 | 23.55 | 24.70 | 27.21 | 28.20 |
| 177 | Undetermined | Undetermined | Undetermined | Undetermined | Undetermined | Undetermined | Undetermined | Undetermined |
| 178 | Undetermined | Undetermined | Undetermined | Undetermined | Undetermined | Undetermined | Undetermined | Undetermined |
| 179 | Undetermined | Undetermined | Undetermined | Undetermined | Undetermined | Undetermined | Undetermined | Undetermined |
| 180 | Undetermined | Undetermined | Undetermined | Undetermined | Undetermined | Undetermined | Undetermined | Undetermined |
| 181 | Undetermined | Undetermined | Undetermined | Undetermined | Undetermined | Undetermined | Undetermined | Undetermined |
| 182 | Undetermined | Undetermined | Undetermined | Undetermined | Undetermined | Undetermined | Undetermined | Undetermined |
| 183 | Undetermined | Undetermined | Undetermined | Undetermined | Undetermined | Undetermined | Undetermined | Undetermined |
| 184 | Undetermined | Undetermined | Undetermined | Undetermined | Undetermined | Undetermined | Undetermined | Undetermined |
| 185 | Undetermined | Undetermined | Undetermined | Undetermined | Undetermined | Undetermined | Undetermined | Undetermined |
| 186 | Undetermined | Undetermined | Undetermined | Undetermined | Undetermined | Undetermined | Undetermined | Undetermined |
| 187 | Undetermined | Undetermined | Undetermined | Undetermined | Undetermined | Undetermined | Undetermined | Undetermined |
| 188 | Undetermined | Undetermined | Undetermined | Undetermined | Undetermined | Undetermined | Undetermined | Undetermined |
| 189 | Undetermined | Undetermined | Undetermined | Undetermined | Undetermined | Undetermined | Undetermined | Undetermined |
| 190 | Undetermined | Undetermined | Undetermined | Undetermined | Undetermined | Undetermined | Undetermined | Undetermined |
| 191 | Undetermined | Undetermined | 36.9 | Undetermined | Undetermined | Undetermined | Undetermined | Undetermined |
| 192 | 22.5 | 22.84 | 27.1 | 23.91 | 20.49 | 21.65 | 17.30 | 18.89 |
| 193 | Undetermined | Undetermined | Undetermined | Undetermined | Undetermined | Undetermined | Undetermined | Undetermined |
| 194 | Undetermined | Undetermined | Undetermined | Undetermined | Undetermined | Undetermined | Undetermined | Undetermined |
| 195 | Undetermined | Undetermined | Undetermined | Undetermined | Undetermined | Undetermined | Undetermined | Undetermined |
| 196 | Undetermined | Undetermined | Undetermined | Undetermined | Undetermined | Undetermined | Undetermined | Undetermined |
| 197 | Undetermined | Undetermined | Undetermined | Undetermined | Undetermined | Undetermined | Undetermined | Undetermined |
| 198 | Undetermined | Undetermined | Undetermined | Undetermined | Undetermined | Undetermined | Undetermined | Undetermined |
| 199 | Undetermined | Undetermined | Undetermined | Undetermined | Undetermined | Undetermined | Undetermined | Undetermined |
| 200 | Undetermined | Undetermined | Undetermined | Undetermined | Undetermined | Undetermined | Undetermined | Undetermined |

**Figure A8.4:** *DNA line data for T. trichiura at Quantigen and Smith*

| | Extract 1 | | Extract 2 | | Extract 3 | | Extract 4 | |
|---|---|---|---|---|---|---|---|---|
| **Sample ID** | **Quantigen** | **Smith** | **Quantigen** | **Smith** | **Smith** | **Quantigen** | **Smith** | **Quantigen** |
| 1 | Undetermined | Undetermined | Undetermined | Undetermined | Undetermined | Undetermined | Undetermined | Undetermined |
| 2 | Undetermined | Undetermined | Undetermined | Undetermined | Undetermined | Undetermined | Undetermined | Undetermined |
| 3 | Undetermined | Undetermined | Undetermined | Undetermined | Undetermined | Undetermined | Undetermined | Undetermined |
| 4 | Undetermined | Undetermined | Undetermined | Undetermined | Undetermined | Undetermined | Undetermined | Undetermined |
| 5 | Undetermined | Undetermined | Undetermined | Undetermined | Undetermined | Undetermined | Undetermined | Undetermined |
| 6 | Undetermined | Undetermined | Undetermined | Undetermined | Undetermined | Undetermined | Undetermined | Undetermined |
| 7 | Undetermined | Undetermined | Undetermined | Undetermined | Undetermined | Undetermined | Undetermined | Undetermined |
| 8 | Undetermined | Undetermined | Undetermined | Undetermined | Undetermined | Undetermined | Undetermined | Undetermined |
| 9 | Undetermined | Undetermined | Undetermined | Undetermined | Undetermined | Undetermined | Undetermined | Undetermined |
| 10 | Undetermined | Undetermined | Undetermined | Undetermined | Undetermined | Undetermined | Undetermined | Undetermined |
| 11 | Undetermined | Undetermined | Undetermined | Undetermined | Undetermined | Undetermined | Undetermined | Undetermined |
| 12 | Undetermined | Undetermined | Undetermined | Undetermined | Undetermined | Undetermined | Undetermined | Undetermined |
| 13 | Undetermined | Undetermined | Undetermined | Undetermined | Undetermined | Undetermined | Undetermined | Undetermined |
| 14 | Undetermined | Undetermined | Undetermined | Undetermined | Undetermined | Undetermined | Undetermined | Undetermined |
| 15 | Undetermined | Undetermined | Undetermined | Undetermined | Undetermined | Undetermined | Undetermined | Undetermined |
| 16 | Undetermined | Undetermined | Undetermined | Undetermined | Undetermined | Undetermined | Undetermined | Undetermined |
| 17 | Undetermined | Undetermined | Undetermined | Undetermined | Undetermined | Undetermined | Undetermined | Undetermined |
| 18 | Undetermined | Undetermined | Undetermined | Undetermined | Undetermined | Undetermined | Undetermined | Undetermined |
| 19 | Undetermined | Undetermined | Undetermined | Undetermined | Undetermined | Undetermined | Undetermined | Undetermined |
| 20 | Undetermined | Undetermined | Undetermined | Undetermined | Undetermined | Undetermined | Undetermined | Undetermined |
| 21 | Undetermined | Undetermined | Undetermined | Undetermined | Undetermined | Undetermined | Undetermined | Undetermined |
| 22 | Undetermined | Undetermined | Undetermined | Undetermined | Undetermined | Undetermined | Undetermined | Undetermined |
| 23 | Undetermined | Undetermined | Undetermined | Undetermined | Undetermined | Undetermined | Undetermined | Undetermined |
| 24 | Undetermined | Undetermined | Undetermined | Undetermined | Undetermined | Undetermined | Undetermined | Undetermined |
| 25 | Undetermined | Undetermined | Undetermined | Undetermined | Undetermined | Undetermined | Undetermined | Undetermined |

| | | | | | | | | |
|---|---|---|---|---|---|---|---|---|
| 26 | Undetermined | Undetermined | Undetermined | Undetermined | Undetermined | Undetermined | Undetermined | Undetermined |
| 27 | Undetermined | Undetermined | Undetermined | Undetermined | Undetermined | Undetermined | Undetermined | Undetermined |
| 28 | Undetermined | Undetermined | Undetermined | Undetermined | Undetermined | Undetermined | Undetermined | Undetermined |
| 29 | Undetermined | Undetermined | Undetermined | Undetermined | Undetermined | Undetermined | Undetermined | Undetermined |
| 30 | Undetermined | Undetermined | Undetermined | Undetermined | Undetermined | Undetermined | Undetermined | Undetermined |
| 31 | Undetermined | Undetermined | Undetermined | Undetermined | Undetermined | Undetermined | Undetermined | Undetermined |
| 32 | Undetermined | Undetermined | Undetermined | Undetermined | Undetermined | Undetermined | Undetermined | Undetermined |
| 33 | Undetermined | Undetermined | Undetermined | Undetermined | Undetermined | Undetermined | Undetermined | Undetermined |
| 34 | Undetermined | Undetermined | Undetermined | Undetermined | Undetermined | Undetermined | Undetermined | Undetermined |
| 35 | Undetermined | Undetermined | Undetermined | Undetermined | Undetermined | Undetermined | Undetermined | Undetermined |
| 36 | Undetermined | Undetermined | Undetermined | Undetermined | Undetermined | Undetermined | Undetermined | Undetermined |
| 37 | Undetermined | Undetermined | Undetermined | Undetermined | Undetermined | Undetermined | Undetermined | Undetermined |
| 38 | Undetermined | Undetermined | Undetermined | Undetermined | Undetermined | Undetermined | Undetermined | Undetermined |
| 39 | Undetermined | Undetermined | Undetermined | Undetermined | Undetermined | Undetermined | Undetermined | Undetermined |
| 40 | Undetermined | Undetermined | Undetermined | Undetermined | Undetermined | Undetermined | Undetermined | Undetermined |
| 41 | Undetermined | Undetermined | Undetermined | Undetermined | Undetermined | Undetermined | Undetermined | Undetermined |
| 42 | Undetermined | Undetermined | Undetermined | Undetermined | Undetermined | Undetermined | Undetermined | Undetermined |
| 43 | Undetermined | Undetermined | Undetermined | Undetermined | Undetermined | Undetermined | Undetermined | Undetermined |
| 44 | Undetermined | Undetermined | Undetermined | Undetermined | Undetermined | Undetermined | Undetermined | Undetermined |
| 45 | Undetermined | Undetermined | Undetermined | Undetermined | Undetermined | Undetermined | Undetermined | Undetermined |
| 46 | Undetermined | Undetermined | Undetermined | Undetermined | Undetermined | Undetermined | Undetermined | Undetermined |
| 47 | Undetermined | Undetermined | Undetermined | Undetermined | Undetermined | Undetermined | Undetermined | Undetermined |
| 48 | Undetermined | Undetermined | Undetermined | Undetermined | Undetermined | Undetermined | Undetermined | Undetermined |
| 49 | Undetermined | Undetermined | Undetermined | Undetermined | Undetermined | Undetermined | Undetermined | Undetermined |
| 50 | Undetermined | Undetermined | Undetermined | Undetermined | Undetermined | Undetermined | Undetermined | Undetermined |
| 51 | Undetermined | Undetermined | Undetermined | Undetermined | Undetermined | Undetermined | Undetermined | Undetermined |
| 52 | Undetermined | Undetermined | Undetermined | Undetermined | Undetermined | Undetermined | Undetermined | Undetermined |
| 53 | Undetermined | Undetermined | Undetermined | Undetermined | Undetermined | Undetermined | Undetermined | Undetermined |
| 54 | Undetermined | Undetermined | Undetermined | Undetermined | Undetermined | Undetermined | Undetermined | Undetermined |
| 55 | Undetermined | Undetermined | Undetermined | Undetermined | Undetermined | Undetermined | Undetermined | Undetermined |

| | | | | | | | | |
|---|---|---|---|---|---|---|---|---|
| 56 | Undetermined | Undetermined | Undetermined | Undetermined | Undetermined | Undetermined | Undetermined | Undetermined |
| 57 | Undetermined | Undetermined | Undetermined | Undetermined | Undetermined | Undetermined | Undetermined | Undetermined |
| 58 | Undetermined | Undetermined | Undetermined | Undetermined | Undetermined | Undetermined | Undetermined | Undetermined |
| 59 | Undetermined | Undetermined | Undetermined | Undetermined | Undetermined | Undetermined | Undetermined | Undetermined |
| 60 | Undetermined | Undetermined | Undetermined | Undetermined | Undetermined | Undetermined | Undetermined | Undetermined |
| 61 | Undetermined | Undetermined | Undetermined | Undetermined | Undetermined | Undetermined | Undetermined | Undetermined |
| 62 | Undetermined | Undetermined | Undetermined | Undetermined | Undetermined | Undetermined | Undetermined | Undetermined |
| 63 | Undetermined | Undetermined | Undetermined | Undetermined | Undetermined | Undetermined | Undetermined | Undetermined |
| 64 | Undetermined | Undetermined | Undetermined | Undetermined | Undetermined | Undetermined | Undetermined | Undetermined |
| 65 | Undetermined | Undetermined | Undetermined | Undetermined | Undetermined | Undetermined | Undetermined | Undetermined |
| 66 | Undetermined | Undetermined | Undetermined | Undetermined | Undetermined | Undetermined | Undetermined | Undetermined |
| 67 | Undetermined | Undetermined | Undetermined | Undetermined | Undetermined | Undetermined | Undetermined | Undetermined |
| 68 | Undetermined | Undetermined | Undetermined | Undetermined | Undetermined | Undetermined | Undetermined | Undetermined |
| 69 | Undetermined | Undetermined | Undetermined | Undetermined | Undetermined | Undetermined | Undetermined | Undetermined |
| 70 | Undetermined | Undetermined | Undetermined | Undetermined | Undetermined | Undetermined | Undetermined | Undetermined |
| 71 | Undetermined | Undetermined | Undetermined | Undetermined | Undetermined | Undetermined | Undetermined | Undetermined |
| 72 | Undetermined | Undetermined | Undetermined | Undetermined | Undetermined | Undetermined | Undetermined | Undetermined |
| 73 | Undetermined | Undetermined | Undetermined | Undetermined | Undetermined | Undetermined | Undetermined | Undetermined |
| 74 | Undetermined | Undetermined | Undetermined | Undetermined | Undetermined | Undetermined | Undetermined | Undetermined |
| 75 | Undetermined | Undetermined | Undetermined | Undetermined | Undetermined | Undetermined | Undetermined | Undetermined |
| 76 | Undetermined | Undetermined | Undetermined | Undetermined | Undetermined | Undetermined | Undetermined | Undetermined |
| 77 | Undetermined | Undetermined | Undetermined | Undetermined | Undetermined | Undetermined | Undetermined | Undetermined |
| 78 | Undetermined | Undetermined | Undetermined | Undetermined | Undetermined | Undetermined | Undetermined | Undetermined |
| 79 | Undetermined | Undetermined | Undetermined | Undetermined | Undetermined | Undetermined | Undetermined | Undetermined |
| 80 | Undetermined | Undetermined | Undetermined | Undetermined | Undetermined | Undetermined | Undetermined | Undetermined |
| 81 | Undetermined | Undetermined | Undetermined | Undetermined | Undetermined | Undetermined | Undetermined | Undetermined |
| 82 | Undetermined | Undetermined | Undetermined | Undetermined | Undetermined | Undetermined | Undetermined | Undetermined |
| 83 | Undetermined | Undetermined | Undetermined | Undetermined | Undetermined | Undetermined | Undetermined | Undetermined |
| 84 | Undetermined | Undetermined | Undetermined | Undetermined | Undetermined | Undetermined | Undetermined | Undetermined |
| 85 | Undetermined | Undetermined | Undetermined | Undetermined | Undetermined | Undetermined | Undetermined | Undetermined |

| | | | | | | | | |
|---|---|---|---|---|---|---|---|---|
| 86 | Undetermined | Undetermined | Undetermined | Undetermined | Undetermined | Undetermined | Undetermined | Undetermined |
| 87 | Undetermined | Undetermined | Undetermined | Undetermined | Undetermined | Undetermined | Undetermined | Undetermined |
| 88 | Undetermined | Undetermined | Undetermined | Undetermined | Undetermined | Undetermined | Undetermined | Undetermined |
| 89 | Undetermined | Undetermined | Undetermined | Undetermined | Undetermined | Undetermined | Undetermined | Undetermined |
| 90 | Undetermined | Undetermined | Undetermined | Undetermined | Undetermined | Undetermined | Undetermined | Undetermined |
| 91 | Undetermined | Undetermined | Undetermined | Undetermined | Undetermined | Undetermined | Undetermined | Undetermined |
| 92 | Undetermined | Undetermined | Undetermined | Undetermined | Undetermined | Undetermined | Undetermined | Undetermined |
| 93 | Undetermined | Undetermined | Undetermined | Undetermined | Undetermined | Undetermined | Undetermined | Undetermined |
| 94 | Undetermined | Undetermined | Undetermined | Undetermined | Undetermined | Undetermined | Undetermined | Undetermined |
| 95 | Undetermined | Undetermined | Undetermined | Undetermined | Undetermined | Undetermined | Undetermined | Undetermined |
| 96 | Undetermined | Undetermined | Undetermined | Undetermined | Undetermined | Undetermined | Undetermined | Undetermined |
| 97 | Undetermined | Undetermined | Undetermined | Undetermined | Undetermined | Undetermined | Undetermined | Undetermined |
| 98 | Undetermined | Undetermined | Undetermined | Undetermined | Undetermined | Undetermined | Undetermined | Undetermined |
| 99 | Undetermined | Undetermined | Undetermined | Undetermined | Undetermined | Undetermined | Undetermined | Undetermined |
| 100 | Undetermined | Undetermined | Undetermined | Undetermined | Undetermined | Undetermined | Undetermined | Undetermined |
| 101 | Undetermined | Undetermined | Undetermined | Undetermined | Undetermined | Undetermined | Undetermined | Undetermined |
| 102 | Undetermined | Undetermined | Undetermined | Undetermined | Undetermined | Undetermined | Undetermined | Undetermined |
| 103 | Undetermined | Undetermined | Undetermined | Undetermined | Undetermined | Undetermined | Undetermined | Undetermined |
| 104 | Undetermined | Undetermined | Undetermined | Undetermined | 28.6 | 31.3 | 26.8 | 28.1 |
| 105 | Undetermined | Undetermined | Undetermined | Undetermined | Undetermined | Undetermined | Undetermined | Undetermined |
| 106 | Undetermined | Undetermined | Undetermined | Undetermined | Undetermined | Undetermined | Undetermined | Undetermined |
| 107 | Undetermined | Undetermined | Undetermined | Undetermined | Undetermined | Undetermined | Undetermined | Undetermined |
| 108 | Undetermined | Undetermined | Undetermined | Undetermined | Undetermined | Undetermined | Undetermined | Undetermined |
| 109 | Undetermined | Undetermined | Undetermined | Undetermined | Undetermined | Undetermined | Undetermined | Undetermined |
| 110 | Undetermined | Undetermined | Undetermined | Undetermined | Undetermined | Undetermined | Undetermined | Undetermined |
| 111 | Undetermined | Undetermined | Undetermined | Undetermined | Undetermined | Undetermined | Undetermined | Undetermined |
| 112 | Undetermined | Undetermined | Undetermined | Undetermined | Undetermined | Undetermined | Undetermined | Undetermined |
| 113 | Undetermined | Undetermined | Undetermined | Undetermined | Undetermined | Undetermined | Undetermined | Undetermined |
| 114 | Undetermined | Undetermined | Undetermined | Undetermined | Undetermined | Undetermined | Undetermined | Undetermined |
| 115 | Undetermined | Undetermined | Undetermined | Undetermined | Undetermined | Undetermined | Undetermined | Undetermined |

| | | | | | | | | | |
|---|---|---|---|---|---|---|---|---|---|
| 116 | Undetermined | Undetermined | Undetermined | Undetermined | Undetermined | Undetermined | Undetermined | Undetermined | Undetermined |
| 117 | Undetermined | Undetermined | Undetermined | Undetermined | Undetermined | Undetermined | Undetermined | Undetermined | Undetermined |
| 118 | Undetermined | Undetermined | Undetermined | Undetermined | Undetermined | Undetermined | Undetermined | Undetermined | Undetermined |
| 119 | Undetermined | Undetermined | Undetermined | Undetermined | Undetermined | Undetermined | Undetermined | Undetermined | Undetermined |
| 120 | Undetermined | Undetermined | Undetermined | Undetermined | Undetermined | Undetermined | Undetermined | Undetermined | Undetermined |
| 121 | Undetermined | Undetermined | Undetermined | Undetermined | Undetermined | Undetermined | Undetermined | Undetermined | Undetermined |
| 122 | Undetermined | Undetermined | Undetermined | Undetermined | Undetermined | Undetermined | Undetermined | Undetermined | Undetermined |
| 123 | Undetermined | Undetermined | Undetermined | Undetermined | Undetermined | Undetermined | Undetermined | Undetermined | Undetermined |
| 124 | Undetermined | Undetermined | Undetermined | Undetermined | Undetermined | Undetermined | Undetermined | Undetermined | Undetermined |
| 125 | Undetermined | Undetermined | Undetermined | Undetermined | Undetermined | Undetermined | Undetermined | Undetermined | Undetermined |
| 126 | Undetermined | Undetermined | Undetermined | Undetermined | Undetermined | Undetermined | Undetermined | Undetermined | Undetermined |
| 127 | Undetermined | Undetermined | Undetermined | Undetermined | Undetermined | Undetermined | Undetermined | Undetermined | Undetermined |
| 128 | Undetermined | Undetermined | Undetermined | Undetermined | Undetermined | Undetermined | Undetermined | Undetermined | Undetermined |
| 129 | Undetermined | Undetermined | Undetermined | Undetermined | Undetermined | Undetermined | Undetermined | Undetermined | Undetermined |
| 130 | Undetermined | Undetermined | Undetermined | Undetermined | Undetermined | Undetermined | Undetermined | Undetermined | Undetermined |
| 131 | Undetermined | Undetermined | Undetermined | Undetermined | Undetermined | Undetermined | Undetermined | Undetermined | Undetermined |
| 132 | Undetermined | Undetermined | Undetermined | Undetermined | Undetermined | Undetermined | Undetermined | Undetermined | Undetermined |
| 133 | Undetermined | Undetermined | Undetermined | Undetermined | Undetermined | Undetermined | Undetermined | Undetermined | Undetermined |
| 134 | Undetermined | Undetermined | Undetermined | Undetermined | Undetermined | Undetermined | Undetermined | Undetermined | Undetermined |
| 135 | Undetermined | Undetermined | Undetermined | Undetermined | Undetermined | Undetermined | Undetermined | Undetermined | Undetermined |
| 136 | Undetermined | Undetermined | Undetermined | Undetermined | Undetermined | Undetermined | Undetermined | Undetermined | Undetermined |
| 137 | Undetermined | Undetermined | Undetermined | Undetermined | Undetermined | Undetermined | Undetermined | Undetermined | Undetermined |
| 138 | Undetermined | Undetermined | Undetermined | Undetermined | Undetermined | Undetermined | Undetermined | Undetermined | Undetermined |
| 139 | Undetermined | Undetermined | Undetermined | Undetermined | Undetermined | Undetermined | Undetermined | Undetermined | Undetermined |
| 140 | Undetermined | Undetermined | Undetermined | Undetermined | Undetermined | Undetermined | Undetermined | Undetermined | Undetermined |
| 141 | Undetermined | Undetermined | Undetermined | Undetermined | Undetermined | Undetermined | Undetermined | Undetermined | Undetermined |
| 142 | Undetermined | Undetermined | Undetermined | Undetermined | Undetermined | Undetermined | Undetermined | Undetermined | Undetermined |
| 143 | Undetermined | Undetermined | Undetermined | Undetermined | Undetermined | Undetermined | Undetermined | Undetermined | Undetermined |
| 144 | Undetermined | Undetermined | Undetermined | Undetermined | Undetermined | Undetermined | Undetermined | Undetermined | Undetermined |
| 145 | Undetermined | Undetermined | Undetermined | Undetermined | Undetermined | Undetermined | Undetermined | Undetermined | Undetermined |

| | | | | | | | | |
|---|---|---|---|---|---|---|---|---|
| 146 | Undetermined | Undetermined | Undetermined | Undetermined | Undetermined | Undetermined | Undetermined | Undetermined |
| 147 | Undetermined | Undetermined | Undetermined | Undetermined | Undetermined | Undetermined | Undetermined | Undetermined |
| 148 | Undetermined | Undetermined | Undetermined | Undetermined | Undetermined | Undetermined | Undetermined | Undetermined |
| 149 | Undetermined | Undetermined | Undetermined | Undetermined | Undetermined | Undetermined | Undetermined | Undetermined |
| 150 | Undetermined | Undetermined | Undetermined | Undetermined | Undetermined | Undetermined | Undetermined | Undetermined |
| 151 | Undetermined | Undetermined | Undetermined | Undetermined | Undetermined | Undetermined | Undetermined | Undetermined |
| 152 | Undetermined | Undetermined | Undetermined | Undetermined | Undetermined | Undetermined | Undetermined | Undetermined |
| 153 | Undetermined | Undetermined | Undetermined | Undetermined | Undetermined | Undetermined | Undetermined | Undetermined |
| 154 | Undetermined | Undetermined | Undetermined | Undetermined | Undetermined | Undetermined | Undetermined | Undetermined |
| 155 | Undetermined | Undetermined | Undetermined | Undetermined | Undetermined | Undetermined | Undetermined | Undetermined |
| 156 | Undetermined | Undetermined | Undetermined | Undetermined | Undetermined | Undetermined | Undetermined | Undetermined |
| 157 | Undetermined | Undetermined | Undetermined | Undetermined | Undetermined | Undetermined | Undetermined | Undetermined |
| 158 | Undetermined | Undetermined | Undetermined | Undetermined | Undetermined | Undetermined | Undetermined | Undetermined |
| 159 | Undetermined | Undetermined | Undetermined | Undetermined | Undetermined | Undetermined | Undetermined | Undetermined |
| 160 | Undetermined | Undetermined | Undetermined | Undetermined | Undetermined | Undetermined | Undetermined | Undetermined |
| 161 | Undetermined | Undetermined | Undetermined | Undetermined | Undetermined | Undetermined | Undetermined | Undetermined |
| 162 | Undetermined | Undetermined | Undetermined | Undetermined | Undetermined | Undetermined | Undetermined | Undetermined |
| 163 | Undetermined | Undetermined | Undetermined | Undetermined | Undetermined | Undetermined | Undetermined | Undetermined |
| 164 | Undetermined | Undetermined | Undetermined | Undetermined | Undetermined | Undetermined | Undetermined | Undetermined |
| 165 | Undetermined | Undetermined | Undetermined | Undetermined | Undetermined | Undetermined | Undetermined | Undetermined |
| 166 | Undetermined | Undetermined | Undetermined | Undetermined | Undetermined | Undetermined | Undetermined | Undetermined |
| 167 | Undetermined | Undetermined | Undetermined | Undetermined | 28.7 | 30.3 | 29.4 | 30.5 |
| 168 | Undetermined | Undetermined | Undetermined | Undetermined | Undetermined | Undetermined | Undetermined | Undetermined |
| 169 | Undetermined | Undetermined | Undetermined | Undetermined | Undetermined | Undetermined | Undetermined | Undetermined |
| 170 | Undetermined | Undetermined | Undetermined | Undetermined | Undetermined | Undetermined | Undetermined | Undetermined |
| 171 | Undetermined | Undetermined | Undetermined | Undetermined | Undetermined | Undetermined | Undetermined | Undetermined |
| 172 | Undetermined | Undetermined | Undetermined | Undetermined | Undetermined | Undetermined | Undetermined | Undetermined |
| 173 | Undetermined | Undetermined | Undetermined | Undetermined | Undetermined | Undetermined | Undetermined | Undetermined |
| 174 | 37.1 | 32.8 | 34.1 | 38.1 | Undetermined | Undetermined | 32.3 | 35.7 |
| 175 | Undetermined | Undetermined | Undetermined | Undetermined | Undetermined | Undetermined | Undetermined | Undetermined |

| | | | | | | | | |
|---|---|---|---|---|---|---|---|---|
| 176 | Undetermined | Undetermined | Undetermined | Undetermined | Undetermined | Undetermined | Undetermined | Undetermined |
| 177 | Undetermined | Undetermined | Undetermined | Undetermined | Undetermined | Undetermined | Undetermined | Undetermined |
| 178 | Undetermined | Undetermined | Undetermined | Undetermined | Undetermined | Undetermined | Undetermined | Undetermined |
| 179 | Undetermined | Undetermined | Undetermined | Undetermined | Undetermined | Undetermined | Undetermined | Undetermined |
| 180 | Undetermined | Undetermined | Undetermined | Undetermined | Undetermined | Undetermined | Undetermined | Undetermined |
| 181 | Undetermined | Undetermined | Undetermined | Undetermined | Undetermined | Undetermined | Undetermined | Undetermined |
| 182 | Undetermined | Undetermined | Undetermined | Undetermined | Undetermined | Undetermined | Undetermined | Undetermined |
| 183 | Undetermined | Undetermined | Undetermined | Undetermined | Undetermined | Undetermined | Undetermined | Undetermined |
| 184 | Undetermined | Undetermined | Undetermined | Undetermined | Undetermined | Undetermined | Undetermined | Undetermined |
| 185 | Undetermined | Undetermined | Undetermined | Undetermined | Undetermined | Undetermined | Undetermined | Undetermined |
| 186 | Undetermined | Undetermined | Undetermined | Undetermined | Undetermined | Undetermined | Undetermined | Undetermined |
| 187 | Undetermined | Undetermined | Undetermined | Undetermined | Undetermined | Undetermined | Undetermined | Undetermined |
| 188 | Undetermined | Undetermined | Undetermined | Undetermined | Undetermined | Undetermined | Undetermined | Undetermined |
| 189 | Undetermined | Undetermined | Undetermined | Undetermined | Undetermined | Undetermined | Undetermined | Undetermined |
| 190 | Undetermined | Undetermined | Undetermined | Undetermined | Undetermined | Undetermined | Undetermined | Undetermined |
| 191 | Undetermined | Undetermined | Undetermined | Undetermined | Undetermined | Undetermined | Undetermined | Undetermined |
| 192 | Undetermined | Undetermined | Undetermined | Undetermined | Undetermined | Undetermined | Undetermined | Undetermined |
| 193 | Undetermined | Undetermined | Undetermined | Undetermined | Undetermined | Undetermined | Undetermined | Undetermined |
| 194 | Undetermined | Undetermined | Undetermined | Undetermined | Undetermined | Undetermined | Undetermined | Undetermined |
| 195 | Undetermined | Undetermined | Undetermined | Undetermined | Undetermined | Undetermined | Undetermined | Undetermined |
| 196 | Undetermined | Undetermined | Undetermined | Undetermined | Undetermined | Undetermined | Undetermined | Undetermined |
| 197 | Undetermined | Undetermined | Undetermined | Undetermined | Undetermined | Undetermined | Undetermined | Undetermined |
| 198 | Undetermined | Undetermined | Undetermined | Undetermined | Undetermined | Undetermined | Undetermined | Undetermined |
| 199 | Undetermined | Undetermined | Undetermined | Undetermined | Undetermined | Undetermined | Undetermined | Undetermined |
| 200 | Undetermined | Undetermined | Undetermined | Undetermined | Undetermined | Undetermined | Undetermined | Undetermined |

**Figure A8.4:** *DNA line data for A. lumbricoides at Quantigen and Smith*

| | Extract 1 | | Extract 2 | | Extract 3 | | Extract 4 | |
|---|---|---|---|---|---|---|---|---|
| Sample ID | Quantigen | Smith | Quantigen | Smith | Smith | Quantigen | Smith | Quantigen |
| 1 | Undetermined | Undetermined | Undetermined | Undetermined | Undetermined | Undetermined | Undetermined | Undetermined |
| 2 | Undetermined | Undetermined | Undetermined | Undetermined | Undetermined | Undetermined | Undetermined | Undetermined |
| 3 | Undetermined | Undetermined | Undetermined | Undetermined | Undetermined | Undetermined | Undetermined | Undetermined |
| 4 | Undetermined | Undetermined | Undetermined | Undetermined | Undetermined | Undetermined | Undetermined | Undetermined |
| 5 | Undetermined | Undetermined | Undetermined | Undetermined | Undetermined | Undetermined | Undetermined | Undetermined |
| 6 | Undetermined | Undetermined | Undetermined | Undetermined | Undetermined | Undetermined | Undetermined | Undetermined |
| 7 | Undetermined | Undetermined | Undetermined | Undetermined | Undetermined | Undetermined | Undetermined | Undetermined |
| 8 | Undetermined | Undetermined | 36.2 | Undetermined | Undetermined | Undetermined | Undetermined | Undetermined |
| 9 | Undetermined | Undetermined | Undetermined | Undetermined | Undetermined | Undetermined | Undetermined | Undetermined |
| 10 | Undetermined | Undetermined | Undetermined | Undetermined | Undetermined | Undetermined | Undetermined | Undetermined |
| 11 | Undetermined | Undetermined | Undetermined | Undetermined | Undetermined | Undetermined | Undetermined | Undetermined |
| 12 | Undetermined | Undetermined | Undetermined | Undetermined | Undetermined | Undetermined | Undetermined | Undetermined |
| 13 | Undetermined | Undetermined | Undetermined | Undetermined | Undetermined | Undetermined | Undetermined | Undetermined |
| 14 | Undetermined | Undetermined | Undetermined | Undetermined | Undetermined | Undetermined | Undetermined | Undetermined |
| 15 | Undetermined | Undetermined | Undetermined | Undetermined | Undetermined | Undetermined | Undetermined | Undetermined |
| 16 | Undetermined | Undetermined | Undetermined | Undetermined | Undetermined | Undetermined | Undetermined | Undetermined |
| 17 | Undetermined | Undetermined | Undetermined | Undetermined | Undetermined | Undetermined | Undetermined | Undetermined |
| 18 | Undetermined | Undetermined | Undetermined | Undetermined | Undetermined | Undetermined | Undetermined | Undetermined |
| 19 | Undetermined | Undetermined | Undetermined | Undetermined | Undetermined | Undetermined | Undetermined | Undetermined |
| 20 | Undetermined | Undetermined | Undetermined | Undetermined | Undetermined | Undetermined | Undetermined | Undetermined |
| 21 | Undetermined | Undetermined | Undetermined | Undetermined | Undetermined | Undetermined | Undetermined | Undetermined |
| 22 | Undetermined | Undetermined | Undetermined | Undetermined | Undetermined | Undetermined | Undetermined | Undetermined |
| 23 | Undetermined | Undetermined | Undetermined | Undetermined | Undetermined | Undetermined | Undetermined | Undetermined |
| 24 | Undetermined | Undetermined | Undetermined | Undetermined | Undetermined | Undetermined | Undetermined | Undetermined |
| 25 | Undetermined | Undetermined | Undetermined | Undetermined | Undetermined | Undetermined | Undetermined | Undetermined |

| | | | | | | | | |
|---|---|---|---|---|---|---|---|---|
| 26 | Undetermined | Undetermined | Undetermined | Undetermined | Undetermined | Undetermined | Undetermined | Undetermined |
| 27 | Undetermined | Undetermined | Undetermined | Undetermined | Undetermined | Undetermined | Undetermined | Undetermined |
| 28 | Undetermined | Undetermined | Undetermined | Undetermined | Undetermined | Undetermined | Undetermined | Undetermined |
| 29 | Undetermined | Undetermined | Undetermined | Undetermined | Undetermined | Undetermined | Undetermined | Undetermined |
| 30 | Undetermined | Undetermined | Undetermined | Undetermined | Undetermined | Undetermined | Undetermined | Undetermined |
| 31 | Undetermined | Undetermined | Undetermined | Undetermined | Undetermined | Undetermined | Undetermined | Undetermined |
| 32 | Undetermined | Undetermined | Undetermined | Undetermined | Undetermined | Undetermined | Undetermined | Undetermined |
| 33 | Undetermined | 22.6 | Undetermined | Undetermined | Undetermined | Undetermined | Undetermined | 28.6 |
| 34 | Undetermined | Undetermined | Undetermined | Undetermined | Undetermined | Undetermined | Undetermined | Undetermined |
| 35 | Undetermined | Undetermined | Undetermined | Undetermined | Undetermined | Undetermined | Undetermined | Undetermined |
| 36 | Undetermined | Undetermined | Undetermined | Undetermined | Undetermined | Undetermined | Undetermined | Undetermined |
| 37 | Undetermined | Undetermined | Undetermined | Undetermined | Undetermined | Undetermined | Undetermined | Undetermined |
| 38 | Undetermined | Undetermined | Undetermined | Undetermined | Undetermined | Undetermined | Undetermined | Undetermined |
| 39 | Undetermined | Undetermined | Undetermined | Undetermined | Undetermined | Undetermined | Undetermined | Undetermined |
| 40 | Undetermined | Undetermined | Undetermined | Undetermined | Undetermined | Undetermined | Undetermined | Undetermined |
| 41 | Undetermined | Undetermined | Undetermined | Undetermined | Undetermined | Undetermined | Undetermined | Undetermined |
| 42 | Undetermined | Undetermined | Undetermined | Undetermined | Undetermined | Undetermined | Undetermined | Undetermined |
| 43 | Undetermined | Undetermined | Undetermined | Undetermined | Undetermined | Undetermined | Undetermined | Undetermined |
| 44 | Undetermined | Undetermined | Undetermined | Undetermined | Undetermined | Undetermined | Undetermined | Undetermined |
| 45 | Undetermined | Undetermined | Undetermined | Undetermined | Undetermined | Undetermined | Undetermined | Undetermined |
| 46 | Undetermined | Undetermined | Undetermined | Undetermined | Undetermined | Undetermined | Undetermined | Undetermined |
| 47 | Undetermined | Undetermined | Undetermined | Undetermined | Undetermined | Undetermined | Undetermined | Undetermined |
| 48 | Undetermined | Undetermined | Undetermined | Undetermined | Undetermined | Undetermined | Undetermined | Undetermined |
| 49 | Undetermined | Undetermined | Undetermined | Undetermined | Undetermined | Undetermined | Undetermined | Undetermined |
| 50 | Undetermined | Undetermined | Undetermined | Undetermined | Undetermined | Undetermined | Undetermined | Undetermined |
| 51 | Undetermined | Undetermined | Undetermined | Undetermined | Undetermined | Undetermined | Undetermined | Undetermined |
| 52 | Undetermined | Undetermined | Undetermined | Undetermined | Undetermined | Undetermined | Undetermined | Undetermined |
| 53 | Undetermined | Undetermined | Undetermined | Undetermined | Undetermined | Undetermined | Undetermined | Undetermined |
| 54 | Undetermined | Undetermined | Undetermined | Undetermined | Undetermined | Undetermined | Undetermined | Undetermined |
| 55 | Undetermined | Undetermined | Undetermined | Undetermined | Undetermined | Undetermined | Undetermined | Undetermined |

| | | | | | | | | |
|---|---|---|---|---|---|---|---|---|
| 56 | Undetermined | Undetermined | Undetermined | Undetermined | Undetermined | Undetermined | Undetermined | Undetermined |
| 57 | 19.8 | 19.2 | 20.8 | 19.6 | 15.6 | 17.9 | 15.2 | 18.0 |
| 58 | Undetermined | Undetermined | Undetermined | Undetermined | Undetermined | Undetermined | Undetermined | Undetermined |
| 59 | Undetermined | Undetermined | Undetermined | Undetermined | Undetermined | Undetermined | Undetermined | Undetermined |
| 60 | Undetermined | Undetermined | Undetermined | Undetermined | Undetermined | Undetermined | Undetermined | Undetermined |
| 61 | Undetermined | Undetermined | Undetermined | Undetermined | Undetermined | Undetermined | Undetermined | Undetermined |
| 62 | Undetermined | Undetermined | Undetermined | Undetermined | Undetermined | Undetermined | Undetermined | Undetermined |
| 63 | 18.5 | 17.7 | 19.1 | 18.3 | 14.4 | 16.4 | 14.9 | 15.9 |
| 64 | Undetermined | Undetermined | Undetermined | Undetermined | Undetermined | Undetermined | Undetermined | Undetermined |
| 65 | Undetermined | Undetermined | Undetermined | Undetermined | Undetermined | Undetermined | Undetermined | Undetermined |
| 66 | Undetermined | Undetermined | Undetermined | Undetermined | Undetermined | Undetermined | Undetermined | Undetermined |
| 67 | 34.7 | 33.8 | 37.0 | 34.2 | 32.6 | 33.3 | 32.1 | 32.7 |
| 68 | Undetermined | Undetermined | Undetermined | Undetermined | Undetermined | Undetermined | Undetermined | Undetermined |
| 69 | Undetermined | Undetermined | Undetermined | Undetermined | Undetermined | 33.5 | Undetermined | 35.6 |
| 70 | 11.7 | 11.4 | 12.3 | 10.9 | 7.8 | 9.2 | 9.3 | 10.4 |
| 71 | 18.3 | 17.2 | 17.7 | 16.9 | 14.1 | 15.5 | 15.0 | 14.3 |
| 72 | Undetermined | Undetermined | Undetermined | Undetermined | Undetermined | Undetermined | Undetermined | Undetermined |
| 73 | Undetermined | Undetermined | Undetermined | Undetermined | Undetermined | Undetermined | Undetermined | Undetermined |
| 74 | Undetermined | Undetermined | Undetermined | Undetermined | Undetermined | Undetermined | Undetermined | Undetermined |
| 75 | Undetermined | Undetermined | Undetermined | Undetermined | Undetermined | Undetermined | Undetermined | Undetermined |
| 76 | Undetermined | Undetermined | Undetermined | Undetermined | Undetermined | Undetermined | Undetermined | Undetermined |
| 77 | Undetermined | Undetermined | Undetermined | Undetermined | Undetermined | Undetermined | Undetermined | Undetermined |
| 78 | Undetermined | Undetermined | Undetermined | Undetermined | Undetermined | Undetermined | Undetermined | Undetermined |
| 79 | 18.0 | 17.2 | 19.8 | 16.9 | 14.5 | 14.2 | 14.3 | 14.3 |
| 80 | Undetermined | Undetermined | Undetermined | Undetermined | Undetermined | Undetermined | Undetermined | Undetermined |
| 81 | Undetermined | Undetermined | 34.2 | 30.8 | Undetermined | Undetermined | Undetermined | Undetermined |
| 82 | Undetermined | Undetermined | Undetermined | Undetermined | Undetermined | Undetermined | Undetermined | Undetermined |
| 83 | 12.6 | 11.8 | 12.5 | 11.7 | 9.0 | 10.3 | 8.5 | 9.8 |
| 84 | Undetermined | 33.4 | Undetermined | Undetermined | Undetermined | 32.5 | 31.9 | 36.4 |
| 85 | 15.1 | 13.7 | 15.7 | 13.7 | 10.4 | 10.4 | 9.9 | 10.4 |

| | | | | | | | | |
|---|---|---|---|---|---|---|---|---|
| 86 | Undetermined | Undetermined | Undetermined | Undetermined | Undetermined | Undetermined | Undetermined | Undetermined |
| 87 | Undetermined | Undetermined | Undetermined | Undetermined | Undetermined | Undetermined | Undetermined | Undetermined |
| 88 | Undetermined | Undetermined | Undetermined | Undetermined | Undetermined | Undetermined | Undetermined | Undetermined |
| 89 | 19.5 | 18.4 | 21.8 | 20.4 | 14.5 | 15.2 | 14.7 | 15.8 |
| 90 | Undetermined | Undetermined | Undetermined | Undetermined | Undetermined | Undetermined | Undetermined | Undetermined |
| 91 | Undetermined | Undetermined | Undetermined | Undetermined | Undetermined | Undetermined | Undetermined | Undetermined |
| 92 | Undetermined | Undetermined | Undetermined | Undetermined | Undetermined | Undetermined | Undetermined | Undetermined |
| 93 | Undetermined | Undetermined | Undetermined | Undetermined | Undetermined | Undetermined | Undetermined | Undetermined |
| 94 | Undetermined | Undetermined | Undetermined | Undetermined | Undetermined | 34.9 | Undetermined | Undetermined |
| 95 | Undetermined | Undetermined | Undetermined | Undetermined | Undetermined | Undetermined | Undetermined | Undetermined |
| 96 | Undetermined | Undetermined | Undetermined | Undetermined | Undetermined | Undetermined | Undetermined | Undetermined |
| 97 | Undetermined | Undetermined | Undetermined | Undetermined | Undetermined | Undetermined | Undetermined | Undetermined |
| 98 | Undetermined | Undetermined | Undetermined | Undetermined | Undetermined | Undetermined | Undetermined | Undetermined |
| 99 | Undetermined | Undetermined | Undetermined | Undetermined | Undetermined | Undetermined | Undetermined | 35.9 |
| 100 | Undetermined | Undetermined | Undetermined | Undetermined | Undetermined | Undetermined | Undetermined | Undetermined |
| 101 | Undetermined | Undetermined | Undetermined | Undetermined | Undetermined | Undetermined | Undetermined | Undetermined |
| 102 | Undetermined | Undetermined | Undetermined | Undetermined | Undetermined | Undetermined | Undetermined | Undetermined |
| 103 | Undetermined | Undetermined | Undetermined | Undetermined | Undetermined | Undetermined | Undetermined | Undetermined |
| 104 | 13.9 | 13.4 | 16.6 | 15.6 | 9.9 | 11.9 | 10.2 | 11.9 |
| 105 | Undetermined | Undetermined | Undetermined | Undetermined | Undetermined | Undetermined | Undetermined | Undetermined |
| 106 | Undetermined | Undetermined | Undetermined | Undetermined | Undetermined | Undetermined | Undetermined | Undetermined |
| 107 | Undetermined | Undetermined | Undetermined | Undetermined | Undetermined | Undetermined | Undetermined | Undetermined |
| 108 | Undetermined | Undetermined | Undetermined | Undetermined | Undetermined | Undetermined | Undetermined | Undetermined |
| 109 | Undetermined | Undetermined | Undetermined | Undetermined | Undetermined | Undetermined | Undetermined | Undetermined |
| 110 | Undetermined | Undetermined | Undetermined | Undetermined | Undetermined | Undetermined | 35.2 | 37.1 |
| 111 | Undetermined | Undetermined | Undetermined | 34.7 | Undetermined | Undetermined | Undetermined | Undetermined |
| 112 | Undetermined | Undetermined | Undetermined | Undetermined | Undetermined | Undetermined | Undetermined | Undetermined |
| 113 | Undetermined | Undetermined | Undetermined | Undetermined | Undetermined | Undetermined | Undetermined | Undetermined |
| 114 | Undetermined | Undetermined | Undetermined | Undetermined | Undetermined | Undetermined | Undetermined | Undetermined |
| 115 | Undetermined | Undetermined | Undetermined | Undetermined | Undetermined | Undetermined | Undetermined | Undetermined |

| | | | | | | | | |
|---|---|---|---|---|---|---|---|---|
| 116 | Undetermined | Undetermined | Undetermined | Undetermined | Undetermined | Undetermined | Undetermined | Undetermined |
| 117 | Undetermined | Undetermined | Undetermined | Undetermined | Undetermined | Undetermined | Undetermined | Undetermined |
| 118 | Undetermined | Undetermined | 36.2 | Undetermined | Undetermined | 36.6 | Undetermined | Undetermined |
| 119 | Undetermined | Undetermined | Undetermined | Undetermined | Undetermined | Undetermined | Undetermined | Undetermined |
| 120 | Undetermined | Undetermined | Undetermined | Undetermined | Undetermined | Undetermined | Undetermined | Undetermined |
| 121 | Undetermined | Undetermined | Undetermined | Undetermined | Undetermined | Undetermined | Undetermined | Undetermined |
| 122 | Undetermined | Undetermined | Undetermined | Undetermined | Undetermined | Undetermined | Undetermined | Undetermined |
| 123 | 13.2 | 12.5 | 15.2 | 13.1 | 9.8 | 12.3 | 10.3 | 11.3 |
| 124 | Undetermined | Undetermined | Undetermined | Undetermined | Undetermined | Undetermined | Undetermined | Undetermined |
| 125 | Undetermined | Undetermined | Undetermined | Undetermined | Undetermined | Undetermined | Undetermined | Undetermined |
| 126 | 19.3 | 16.7 | 19.2 | 17.3 | 15.1 | 15.6 | 15.4 | 16.0 |
| 127 | 13.1 | 12.8 | 13.0 | 11.9 | 8.8 | 9.7 | 8.9 | 10.8 |
| 128 | Undetermined | Undetermined | Undetermined | Undetermined | Undetermined | Undetermined | Undetermined | Undetermined |
| 129 | Undetermined | Undetermined | Undetermined | Undetermined | Undetermined | Undetermined | Undetermined | Undetermined |
| 130 | 18.8 | 18.3 | 19.2 | 18.7 | 11.3 | 12.1 | 12.4 | 11.2 |
| 131 | Undetermined | Undetermined | Undetermined | 34.3 | Undetermined | 33.8 | Undetermined | Undetermined |
| 132 | 34.4 | 30.5 | 31.1 | 27.8 | 31.6 | 29.9 | 26.6 | 26.8 |
| 133 | Undetermined | Undetermined | Undetermined | Undetermined | Undetermined | Undetermined | Undetermined | Undetermined |
| 134 | Undetermined | Undetermined | Undetermined | Undetermined | Undetermined | Undetermined | Undetermined | Undetermined |
| 135 | Undetermined | Undetermined | Undetermined | Undetermined | Undetermined | Undetermined | Undetermined | Undetermined |
| 136 | Undetermined | Undetermined | Undetermined | Undetermined | Undetermined | Undetermined | Undetermined | Undetermined |
| 137 | Undetermined | Undetermined | Undetermined | Undetermined | Undetermined | Undetermined | Undetermined | Undetermined |
| 138 | Undetermined | Undetermined | Undetermined | Undetermined | Undetermined | Undetermined | Undetermined | Undetermined |
| 139 | Undetermined | Undetermined | Undetermined | Undetermined | Undetermined | Undetermined | Undetermined | Undetermined |
| 140 | Undetermined | Undetermined | Undetermined | Undetermined | Undetermined | Undetermined | Undetermined | Undetermined |
| 141 | Undetermined | Undetermined | Undetermined | Undetermined | Undetermined | Undetermined | Undetermined | Undetermined |
| 142 | Undetermined | Undetermined | Undetermined | Undetermined | Undetermined | Undetermined | Undetermined | Undetermined |
| 143 | Undetermined | Undetermined | Undetermined | Undetermined | Undetermined | Undetermined | Undetermined | Undetermined |
| 144 | Undetermined | Undetermined | Undetermined | Undetermined | Undetermined | Undetermined | Undetermined | Undetermined |
| 145 | Undetermined | Undetermined | Undetermined | Undetermined | Undetermined | Undetermined | Undetermined | Undetermined |

| | | | | | | | | |
|---|---|---|---|---|---|---|---|---|
| 146 | Undetermined | Undetermined | Undetermined | Undetermined | Undetermined | Undetermined | Undetermined | Undetermined |
| 147 | Undetermined | Undetermined | Undetermined | Undetermined | Undetermined | Undetermined | Undetermined | Undetermined |
| 148 | Undetermined | Undetermined | Undetermined | 34.7 | Undetermined | Undetermined | Undetermined | Undetermined |
| 149 | Undetermined | Undetermined | Undetermined | Undetermined | Undetermined | Undetermined | Undetermined | Undetermined |
| 150 | Undetermined | Undetermined | Undetermined | Undetermined | Undetermined | Undetermined | Undetermined | Undetermined |
| 151 | Undetermined | Undetermined | Undetermined | Undetermined | Undetermined | Undetermined | Undetermined | Undetermined |
| 152 | Undetermined | Undetermined | Undetermined | Undetermined | Undetermined | Undetermined | Undetermined | Undetermined |
| 153 | Undetermined | Undetermined | Undetermined | Undetermined | Undetermined | Undetermined | Undetermined | Undetermined |
| 154 | Undetermined | Undetermined | Undetermined | Undetermined | Undetermined | Undetermined | Undetermined | Undetermined |
| 155 | Undetermined | Undetermined | 31.5 | 37.0 | Undetermined | Undetermined | Undetermined | Undetermined |
| 156 | Undetermined | Undetermined | Undetermined | Undetermined | Undetermined | Undetermined | Undetermined | Undetermined |
| 157 | Undetermined | Undetermined | Undetermined | Undetermined | Undetermined | Undetermined | Undetermined | Undetermined |
| 158 | Undetermined | Undetermined | 20.0 | 19.0 | Undetermined | Undetermined | Undetermined | Undetermined |
| 159 | Undetermined | Undetermined | Undetermined | Undetermined | Undetermined | Undetermined | Undetermined | Undetermined |
| 160 | Undetermined | Undetermined | Undetermined | Undetermined | Undetermined | Undetermined | Undetermined | Undetermined |
| 161 | Undetermined | Undetermined | Undetermined | Undetermined | Undetermined | Undetermined | Undetermined | Undetermined |
| 162 | Undetermined | Undetermined | Undetermined | Undetermined | Undetermined | Undetermined | Undetermined | Undetermined |
| 163 | Undetermined | Undetermined | Undetermined | Undetermined | Undetermined | Undetermined | Undetermined | Undetermined |
| 164 | Undetermined | Undetermined | Undetermined | Undetermined | Undetermined | Undetermined | Undetermined | Undetermined |
| 165 | Undetermined | Undetermined | Undetermined | Undetermined | Undetermined | Undetermined | Undetermined | Undetermined |
| 166 | Undetermined | Undetermined | Undetermined | Undetermined | Undetermined | Undetermined | Undetermined | Undetermined |
| 167 | Undetermined | Undetermined | Undetermined | Undetermined | Undetermined | Undetermined | Undetermined | Undetermined |
| 168 | Undetermined | Undetermined | Undetermined | Undetermined | Undetermined | Undetermined | Undetermined | Undetermined |
| 169 | Undetermined | Undetermined | 31.2 | 28.6 | Undetermined | Undetermined | 32.4 | 33.5 |
| 170 | Undetermined | Undetermined | Undetermined | Undetermined | Undetermined | Undetermined | Undetermined | Undetermined |
| 171 | Undetermined | Undetermined | Undetermined | Undetermined | Undetermined | Undetermined | Undetermined | Undetermined |
| 172 | Undetermined | Undetermined | Undetermined | Undetermined | Undetermined | Undetermined | Undetermined | Undetermined |
| 173 | Undetermined | Undetermined | 33.8 | 30.2 | Undetermined | Undetermined | Undetermined | Undetermined |
| 174 | Undetermined | Undetermined | Undetermined | Undetermined | Undetermined | Undetermined | Undetermined | Undetermined |
| 175 | Undetermined | Undetermined | Undetermined | Undetermined | Undetermined | Undetermined | Undetermined | Undetermined |

| | | | | | | | | |
|---|---|---|---|---|---|---|---|---|
| 176 | Undetermined | Undetermined | Undetermined | Undetermined | Undetermined | Undetermined | Undetermined | Undetermined |
| 177 | Undetermined | Undetermined | Undetermined | Undetermined | Undetermined | Undetermined | Undetermined | Undetermined |
| 178 | Undetermined | Undetermined | Undetermined | Undetermined | Undetermined | Undetermined | Undetermined | Undetermined |
| 179 | Undetermined | Undetermined | Undetermined | Undetermined | Undetermined | Undetermined | Undetermined | Undetermined |
| 180 | Undetermined | Undetermined | Undetermined | Undetermined | Undetermined | Undetermined | Undetermined | Undetermined |
| 181 | Undetermined | Undetermined | Undetermined | Undetermined | Undetermined | Undetermined | Undetermined | Undetermined |
| 182 | Undetermined | Undetermined | Undetermined | Undetermined | Undetermined | Undetermined | Undetermined | Undetermined |
| 183 | 19.0 | 18.5 | 22.9 | 21.8 | 14.9 | 15.4 | 14.7 | 15.7 |
| 184 | Undetermined | Undetermined | Undetermined | Undetermined | Undetermined | Undetermined | Undetermined | Undetermined |
| 185 | Undetermined | Undetermined | Undetermined | Undetermined | Undetermined | Undetermined | Undetermined | Undetermined |
| 186 | Undetermined | Undetermined | Undetermined | Undetermined | Undetermined | Undetermined | Undetermined | Undetermined |
| 187 | Undetermined | Undetermined | Undetermined | Undetermined | Undetermined | Undetermined | Undetermined | Undetermined |
| 188 | Undetermined | Undetermined | Undetermined | Undetermined | Undetermined | Undetermined | Undetermined | Undetermined |
| 189 | Undetermined | Undetermined | Undetermined | Undetermined | Undetermined | Undetermined | Undetermined | Undetermined |
| 190 | Undetermined | Undetermined | Undetermined | Undetermined | Undetermined | Undetermined | Undetermined | Undetermined |
| 191 | Undetermined | Undetermined | Undetermined | Undetermined | Undetermined | Undetermined | Undetermined | Undetermined |
| 192 | Undetermined | Undetermined | Undetermined | Undetermined | Undetermined | Undetermined | Undetermined | Undetermined |
| 193 | Undetermined | Undetermined | Undetermined | Undetermined | Undetermined | Undetermined | Undetermined | Undetermined |
| 194 | Undetermined | Undetermined | Undetermined | Undetermined | 33.9 | 32.9 | 32.2 | 34.1 |
| 195 | Undetermined | Undetermined | Undetermined | Undetermined | Undetermined | Undetermined | Undetermined | Undetermined |
| 196 | Undetermined | Undetermined | Undetermined | Undetermined | Undetermined | Undetermined | Undetermined | Undetermined |
| 197 | Undetermined | Undetermined | Undetermined | Undetermined | Undetermined | Undetermined | Undetermined | Undetermined |
| 198 | Undetermined | Undetermined | Undetermined | Undetermined | Undetermined | Undetermined | Undetermined | Undetermined |
| 199 | Undetermined | Undetermined | Undetermined | Undetermined | Undetermined | Undetermined | Undetermined | Undetermined |
| 200 | Undetermined | Undetermined | Undetermined | Undetermined | Undetermined | Undetermined | Undetermined | Undetermined |

#### A9: Control Proposal

**Purpose**

The purpose of this proposal is to clearly outline the proposed changes to the current DeWorm3 Assay, specifically with respect to the controls included and how they are analyzed. This proposal will detail each control, the current standard, the new proposal, and the data and evidence to support the new proposal.

**Internal Process Control: *Bacillus atrophaeus***

*Current*

The DeWorm3 Assay *B. atrophaeus* control is spiked into each sample at the time of extraction and serves as an internal process control. The *B. atrophaeus* Ct cut off is defined as 2 standard deviations from the mean Ct defined in the validation report (Ct $\leq$ 34, in the absence of STH amplification). Per the validation report, the official Ct score range for the whole of the DeWorm3 study will be defined by 2 standard deviations from all baseline samples (DeWorm3 Assay Validation Report, *Verification II.A.iv*).

*B. atrophaeus Proposal*

In the interest of solidifying a procedure for determining the *B. atrophaeus* Ct score range, the following proposal has been formulated:

(i) Use the 6,133 LMC baseline samples to define the mean, median, and standard deviation.
(ii) Quantigen to re-extract all samples with *B. atrophaeus* Ct Scores that fall outside of 2 standard deviations from the mean, determined in (i), in one or both assays/wells.
(iii) Comparative analysis of STH-status flips at 2, 3, and 4 standard deviations
    (a) This will allow a standard to be set for the whole study, and subsequent studies in the field, as to how to evaluate Ct cut offs for an internal process control.

Finally, for a sample to have official results, *B. atrophaeus* or an STH target must amplify within the defined range in both sample wells. All samples that are queued for repeat based on *B. atrophaeus* status will be repeated at the completion of initial testing.

*Data in support of B. atrophaeus Proposal*

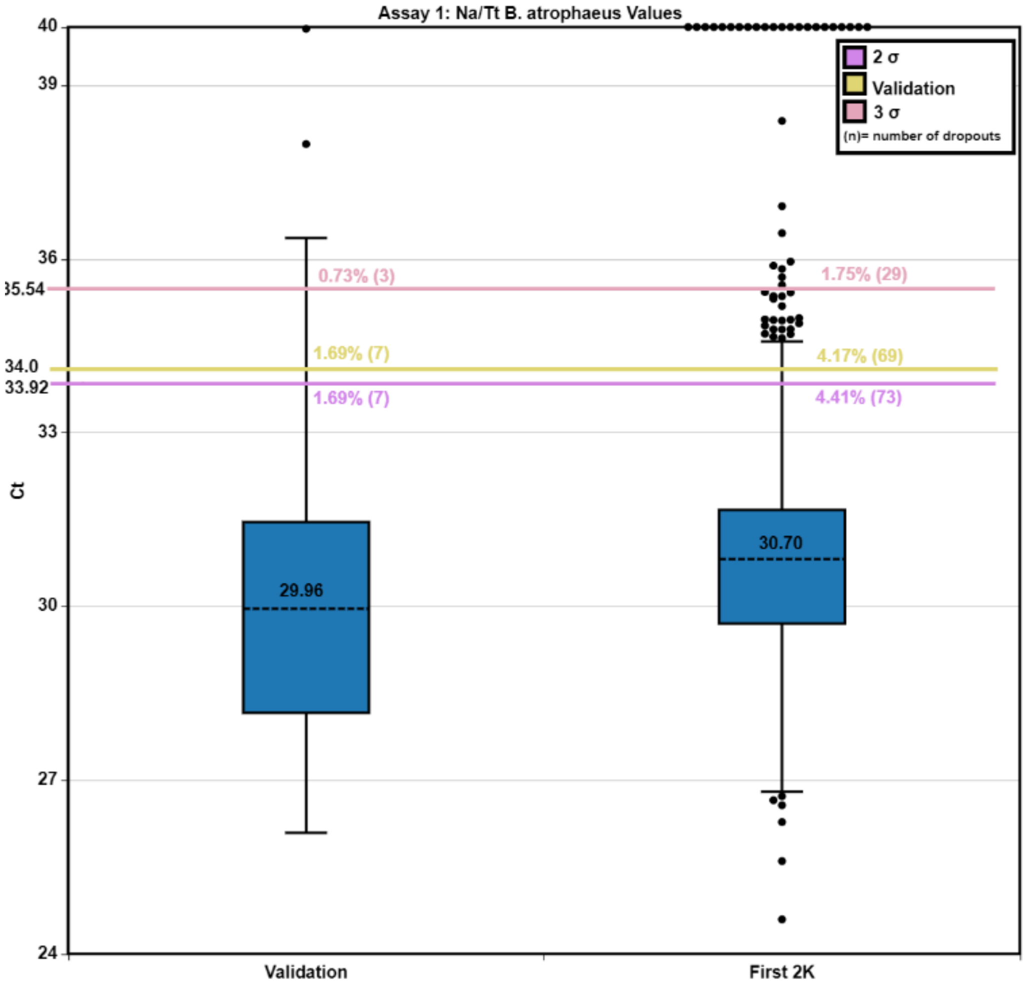

*Figure A9.1 8: Assay 1 (Na/Tt) B. atrophaeus data from Validation and the First 2k, indicating repeats based on Validation cut off, 2 and 3 standard deviation cut offs (from the mean of the first 2,000 samples).*

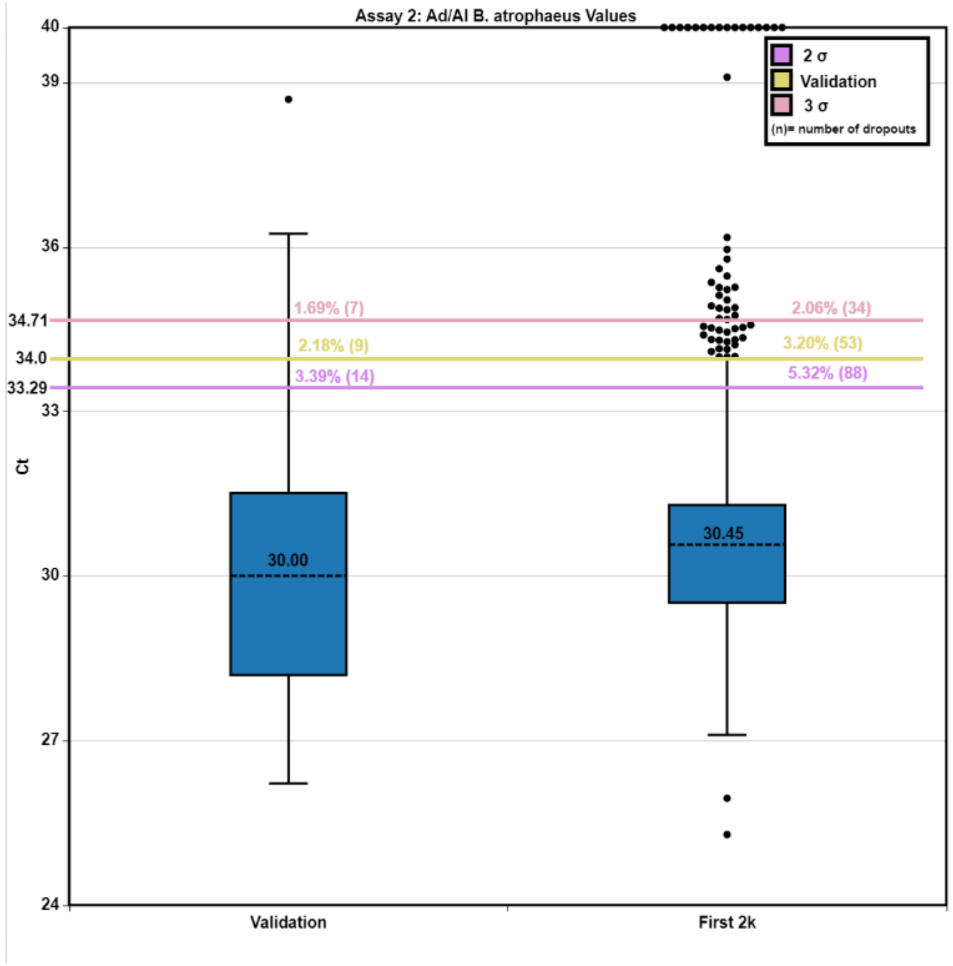

*Figure A9.29: Assay 2 (Ad/Al) B. atrophaeus data from Validation and the First 2k, indicating repeats based on the Validation cut off, 2 and 3 standard deviation cut offs (from the mean of the first 2,000 samples).*

*Table A9. 4: Sample Repeats by Bacillus Cut Offs*

| Bacillus Cut Off Breakdown | | | |
|---|---|---|---|
| | **Well** | **Extraction** | **Total Failures** |
| **2 σ** | 71 | 42 | **113** |
| **3 σ** | 43 | 10 | **53** |
| **Validation** | 58 | 32 | **90** |
| **Undetermined** | 29 | 5 | **34** |

In Figure A9.1 and Figure A9.2, the differences in means (indicated by the dashed black line) between the *B. atrophaeus* scores in Validation (n=413) vs. the First 2,000 (n= 1653) samples can be seen. The colored lines indicate the Ct score cut offs at Validation (yellow line; ≥ 34), 2 standard deviations (in reference to the first 2,000 samples) (purple line; ≥ 33.94 Assay 1, ≥ 33.28 Assay 2), and 3 standard deviations (pink line; ≥ 35.57 Assay 1, ≥ 34.68 Assay 2). Table 1. outlines the number of samples which require re-testing due to an unacceptable *B. atrophaeus* Ct score. Of importance, these data clearly show: (1) a shift in *B. atrophaeus* Ct score averages in the First 2k sample data (30.7 ± 1.61 Assay 1, 30.45 ± 1.42 Assay 2) compared to the validation (29.96 ± 2.02 Assay 1, 30.00 ± 2.00 Assay 2), and (2) the drastic drop in repeats when evaluating at a 3 standard deviation cut off (35.54 Assay 1, 34.71 Assay 2), compared to a 2 standard deviation cut off (33.92 Assay 1, 33.29 Assay 2) or the current validation cut off (34).

These data suggest that the means and cut offs for the whole DeWorm3 study will be better informed by all the Benin LMC data. These data also further support the piece of the proposal, outlined in B.II.ii-iii. As there is currently no gold-standard for how to evaluate internal controls in the STH field, the DeWorm3 project with its scope, sits in a unique position to inform the community at which deviation and thus, Ct value cut off is the most appropriate to pass or fail a sample. Evaluating all baseline sample data will set up a firm standard for the DeWorm3 project, the STH scientific community, and save money and resources by reducing the number of frivolous repeats, which occurs when cut offs are decided arbitrarily.

##### *Supporting Material for the B. atrophaeus Proposal*

Previous studies from the Williams Lab at Smith College provide evidence for how to determine Ct cut offs to repeat samples. In this study[8], it was found that repeating samples at a 3 standard deviation cut-off was most appropriate. This was decided after evaluating the STH status 'flips' at 2, 3, and 4 standard deviations. While this paper was able to determine this after all samples (n=2,800) were run, this is not feasible for the DeWorm3 project, both in terms of data evaluation and the likely high number of repeats at 2 standard deviations.

Again, this study was not done at the same scale of DeWorm3, but it indicates that setting of a Ct score cut off after all – or in the case of DeWorm3, some – samples have been run, as well as a comparative analysis of STH flips, is the current optimal way deal with internal controls cut offs. It may be important to note that this study used a plasmid-based control (IAC), compared to the DeWorm3 Assay, which uses a spiked bacterial control.

Other studies, again using IAC, did not set a Ct cut-off, but rather repeated samples based on whether the IAC was detected at all or whether it was simply 'Undetermined.' This

[8] Benjamin-Chung J, Pilotte N, Ercumen A, Grant JR, Maasch JRMA, et al. (2020) Comparison of multi-parallel qPCR and double-slide Kato-Katz for detection of soil-transmitted helminth infection among children in rural Bangladesh. PLOS Neglected Tropical Diseases 14(4): e0008087. https://doi.org/10.1371/journal.pntd.0008087

paper[9] provides some evidence for not setting a Ct at all, but rather only repeated those samples that show up 'Undetermined.'

These differences are most important when considering the number of repeats across the study. Within the first 2,000 samples, there would be a total of 113 samples repeated, based on 2 standard deviation cut offs; maintaining the Validation cutoff would result in 90 sample repeats. The most compelling data is the drop in the number of samples that are repeated when switching to a 3 standard deviation cut off or a pass/fail cut off. If 3 standard deviations were to be used, then 53 samples would be repeated, while if *B. atrophaeus* was evaluated on a pass/fail basis, only 34 samples would need to be repeated.

**Extraction Control: *A. suum***

##### *Current A. suum Control*

Currently the DeWorm3 Assay, the *A. suum* control is contrived by spiking eggs into naïve stool. Laid out in the validation plan, the *A. suum* extraction control must have a Ct of $\leq$ 23. An *A. suum* extraction control failure (Ct >23) results in an extraction plate failure.

##### *A. suum Proposal*

The new proposal for determining the *A. suum* cut off is as follows:

***(i)*** The Ct cut off for each lot of *A. suum* will be determined after all aliquots from that lot have been ran (~70/lot).[10]

***(ii)*** The Ct cut off will be defined by 2 or 3 standard deviations away from the mean of that lot.[11]

***(iii)*** Plates will be officially passed/failed based on what falls outside of that range.

---

[9] Dunn, J.C., Papaiakovou, M., Han, K.T. *et al.* The increased sensitivity of qPCR in comparison to Kato-Katz is required for the accurate assessment of the prevalence of soil-transmitted helminth infection in settings that have received multiple rounds of mass drug administration. *Parasites Vectors* **13,** 324 (2020). https://doi.org/10.1186/s13071-020-04197-w

[10] A Batch of 70 was a rough estimate by Dr. Pilotte and supported by Dr. Williams. Both agreed this number, while smaller, will provide the opportunity for better results.

[11] The standard deviation number will be determined upon comparative analyses of these, like the other controls.

#### *Data in support of A. suum Proposal*

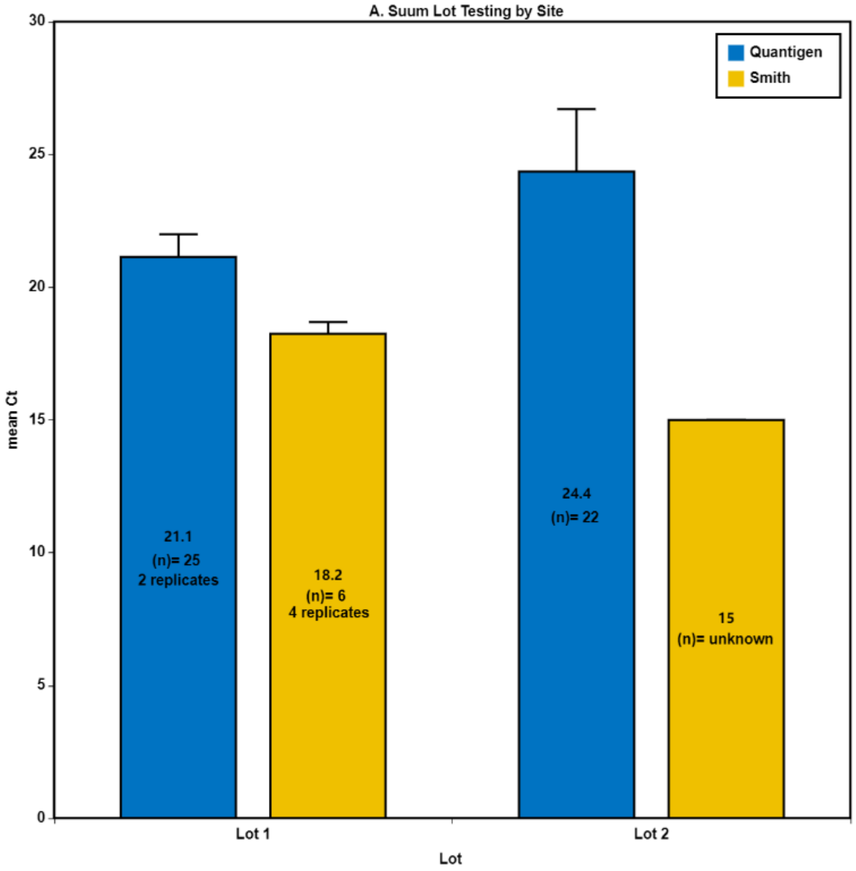

*Figure A9.310: Mean comparison between A. suum lots. Results separated by Smith and Quantigen. Smith and Quantigen threshold are different (0.03 and 0.06) and can cause a 3 Ct shift.*

*Table A9.5: A. suum Lot-to-Lot Variation*

| A. suum (Pos. Extract) | | | | |
|---|---|---|---|---|
| ***Lot #*** | ***1*** | | ***2*** | |
| | **Validation** | **Smith** | **First 2K** | **Smith** |
| ***n*** | *50.0* | *24* | *22* | *x* |
| **Max** | 23.3 | 18.9 | 29.8 | n/a |
| **Min** | 19.3 | 17.5 | 20.7 | n/a |
| **Mean** | 21.1 | 18.3 | 24.4 | 15.0 |
| **σ** | 0.86 | 0.44 | 2.46 | n/a |

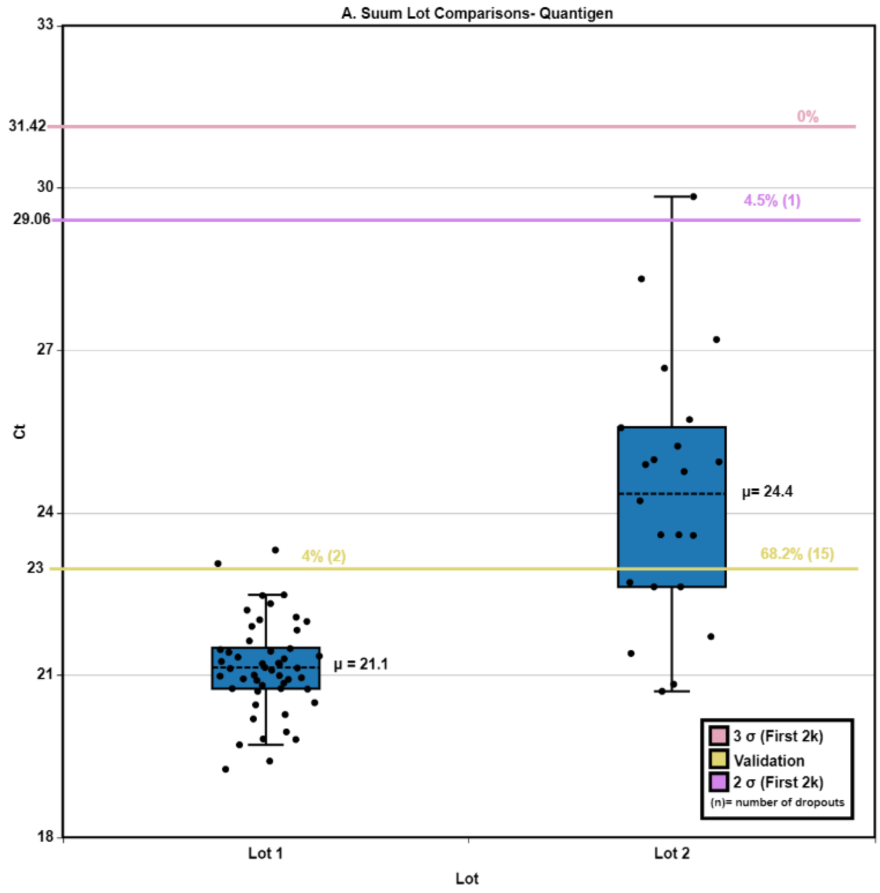

*Figure A9.411: A. suum Lot 1 vs. Lot 2 Box Plot (Quantigen Data Only). Includes Cut Offs based on Validation (Ct $\leq$ 23; yellow line), 2 standard deviations (Lot 2 data Only) (Ct $\leq$ 29.06; purple line), and 3 standard deviations (Ct $\leq$ 31.42; orange line)*

*Table A9.6: First 2k A. Suum Failures*

| ***A. Suum* Cut Off Breakdown (First 2k)** | |
|---|---|
| | **Total Failures** |
| **2 σ** | **1** |
| **3 σ** | **0** |
| **Validation** | **15** |

Looking at all the above figures, the lot-to-lot differences become very apparent. All Validation samples were from Lot 1 while the First 2k samples were from Lot 2. The differences between Quantigen and Smith Ct scores for the *A. suum* are not of concern as the specific testing methods and data analysis is different and have previously shown to produce a shift in Ct scores.

The lot-to-lot differences also speak to the variability of the naïve stool that can be acquired and used. The nature of the sample type and the composition of each stool sample produces differences that further confound this control and dispels the idea that all lots of *A. suum* should be treated the same as the initial Validation lot.

In addition, the control is contrived and created by hand with variable results. Looking at the mean differences between Lots 1 and 2, the mean of Lot 2 is shifted above the Validation Cut Off. This clearly indicates that the mean, range, standard deviation, and cut off that was appropriate for Lot 1 will not be applicable to subsequent lots and will lead to an unsustainable repeat rate.

Further emphasizing this point in Figure 4, there are incredibly high repeat rates (n=15) for the first 22 plates, if the Validation Cut off and previous control standard, is to be followed. Readjusting the data, based on the proposal in C.II, the number of repeat rates drops to 1 with an adjusted 2 standard deviation cut off and drops to 0 with the adjusted 3 standard deviation cut off.

##### *Supporting Material for the A. suum Proposal*

An alternative to the proposal at hand could be to introduce rigorous lot testing which would include testing many samples. However, this is not a feasible solution for the contrived control. The amount of work that goes into creating the control limits the number of controls that can be made per batch. Additionally, the number of aliquots that would be required for extensive lot testing, to ensure that a true range is represented, would introduce the need to pool different aliquots of naïve stool, increasing the amount of variability in composition, and subsequently, Ct scores.

Additionally, the lack of lot testing at one site is not a concern in terms of the stability of the control itself. While there is no stability data on this control and egg, previous studies[12] have shown that even if the eggs break, the DNA itself is stable.

While setting a Ct score cut-off for batches after using a full lot of *A. suum* control is not ideal, the fact that this control is contrived and is not being produced in a QC manner must remain as part of the conversation. There is not a strong and proven precedent for using contrived controls in this manner, and there is certainly no evidence supporting using the first lot of a contrived control to set cut-off ranges for subsequent lots. This standard would be appropriate if the controls had been produced through a Quality Control Process, but as this is not possible for the STH community at this time, this is the best alternative.

[12] Papaiakovou M, Pilotte N, Baumer B, et al. A comparative analysis of preservation techniques for the optimal molecular detection of hookworm DNA in a human fecal specimen. *PLoS Negl Trop Dis*. 2018;12(1):e0006130. Published 2018 Jan 18. doi:10.1371/journal.pntd.0006130

#### Negative Controls: No Sample Control and No Template Control

##### *Current Controls*

###### *No Sample Control (NSC)*

The NSC control is an all-reagent control with no stool added, but with *B. atrophaeus* spiked in. This NSC serves as a negative process control, and the addition of *B. atrophaeus* serves as an extraction control. If the NSC *B. atrophaeus* score falls outside of the determined range (see 0, or any STH is amplified, then this results in an extraction plate failure (DeWorm3 Assay Validation Plan, *I.K. Description, Preparation, and Validation of Run Controls)*.

###### ***No Template Control (NTC)***

The NTC control contains no stool or *B. atrophaeus* and is added directly to the qPCR plate. This is a PCR control to ensure that there is no contamination. If there is any amplification of STH or *B. atrophaeus*, then the NTC fails which results in a qPCR plate failure.

##### *NSC/NTC Proposal*

As there are both an NSC and NTC that serve the function of a contamination control, the general proposal is to eliminate the NTC, making the NSC the primary contamination control on the plate.

The more detailed proposal is as follows:

***a)*** NSC will be “reagent-only,” no *B. atrophaeus* spiked in.
***b)*** Will double as an NTC in qPCR

##### *Data to support NSC/NTC Proposal*

Across the 22 plates of the first 2k data, there have been no NSC failures due to STH contamination. Additionally, there have been no NTC failures due to contamination.

Looking at Figure A9.5: Plate Repeats for the first 2k samples based on NSC failures in Assay 1 (Na/Tt), there are various NSC repeats based on *B. atrophaeus* failures in Assay 1 (Na/Tt); 5 when using the Validation and 2 standard deviation cut off, and 1 using the 3 standard deviation cut off. In Figure 6, there are additional NSC repeats based purely on *B. atrophaeus* failures in Assay 2 (Ad/Al); 9 when using 2 standard deviations, 3 when using the Validation, and 1 using 3 standard deviations.

If this were a sample, there would be either 1 sample repeat or 5 samples repeats, but because this is a control that is tied to the status of the whole plate, all 92 samples would be failed and queued for repeat, thus wasting enormous amounts of resources and time. A visual of these repeats can be seen in Table 4.

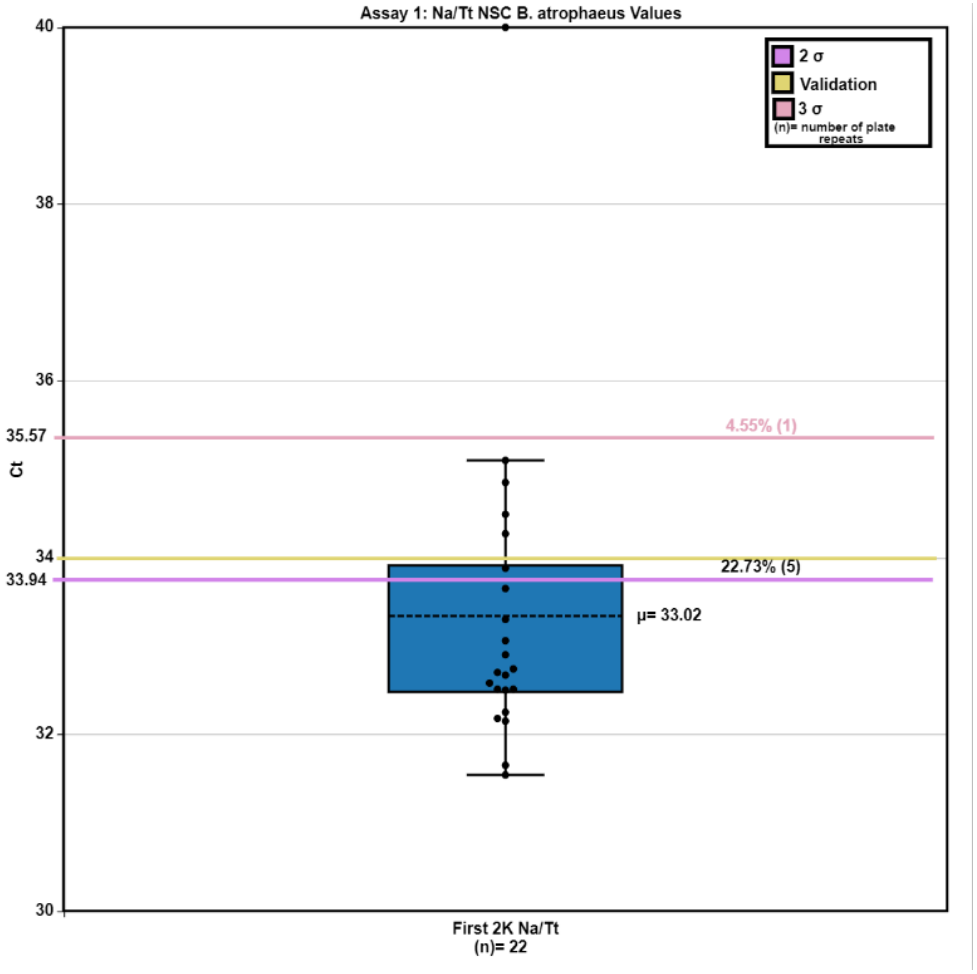

*Figure A9.5 12: Plate Repeats for the first 2k samples based on NSC failures in Assay 1 (Na/Tt)*

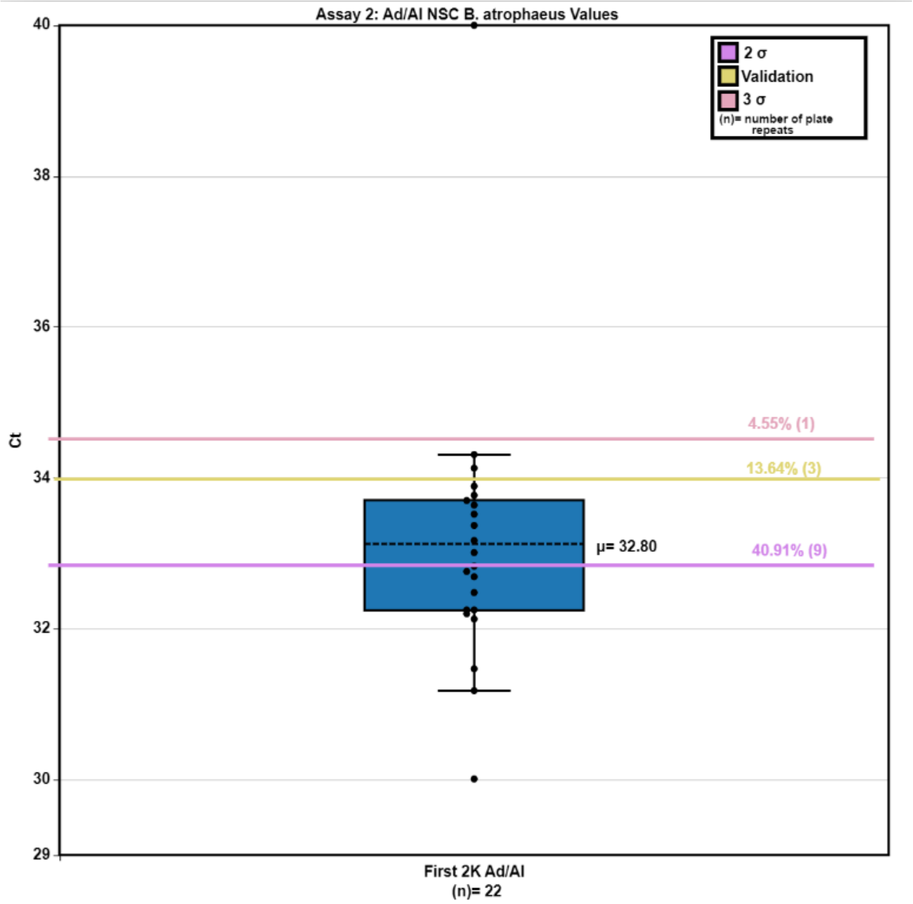

*Figure A9.613: Plate Repeats for the first 2k samples based on NSC failures in Assay 2 (Ad/Al)*

*Table A9. 7: Number of Samples Repeated due to NSC B. atrophaeus failures*

| Number of Sample Repeats from NSC Failures | | |
|---|---|---|
| | **Plates Failed** | **Samples Queued for Repeat** |
| **2 σ** | 14 | 1288 |
| **3 σ** | 2 | 184 |
| **Validation** | 8 | 736 |

To reiterate, all failures indicated in Table A9.4 are failures due strictly to *B. atrophaeus*. There have not been any NSC failures due to STH amplification.

##### *Evidence to support NSC/NTC Proposal*

Other assays do not have both NSC and NTC controls on a plate. As both of these are controls that are intended to monitor contamination, having both are redundant, and as seen in 0, there would be frivolous repeats by including both of these controls. The standard for other developed assays follows this and should be replicated in the DeWorm3 assay.

Furthermore, as the DeWorm3 Assay Plate is lyophilized, the primary opportunity to introduce contaminants (i.e., when the MasterMix is being prepared) has been eliminated. In typical liquid assays, the NTC would test for such contamination, but as the assay is lyophilized this control, again, becomes superfluous and ends up testing for the same contamination that the NSC could.

#### Positive PCR

##### *Current Positive PCR Standard*

Per the Validation Report, the current passing qualification for the DeWorm3 Positive PCR WGA STH Control is within 2 standard deviations from the validation-defined mean (DeWorm3 Assay Validation Report, *II.A.v Validation of Run Control: Whole genome-amplified (WGA) STH nucleic acid)*.

This control is a contrived control that was pooled and aliquoted out, as to limit any lot-to-lot deviations. Although there is technically only one lot of this control, the cocktail of WGA DNA was created in such a way as to target similar Cts across all the STH species. This creation considered the number of repeat sequences and adjusted the amount of input material to generate a particular Ct score. This concept has led to the creation of a control in which there is less WGA material for *N. americanus* and *A. lumbricoides* than the other targets.

##### *Positive PCR Proposal*

###### ***Transition to Plasmids***

Quantigen proposes that the DeWorm3 Assay moves away from using a WGA cocktail, rather using a cocktail of single copy plasmids per target.

***(i)*** If this proposal is accepted, Quantigen will validate this control to set a 3 standard deviation cut off to pass/fail the batch

***(ii)*** If this proposal is accepted, Quantigen secondarily proposes that there are only two wells of positive PCR per 384 well batch (i.e. only on one of the 96-well plates will there be a reserved spot for positive PCR).

***(iii)*** Quantigen will run the plasmid cocktail at 4 pg/PCR reaction (1 pg of plasmid/target).

**Continue with WGA, with altered passing qualifications.**

***(iv)*** If one of the two control wells amplifies all 4 targets in one batch, the batch will pass regardless of the status of the other Pos. PCR control

***(v)*** Ct range cut off to be expanded to include Cts seen in validation and First 2k data

***(vi)*** Alternatively, we could create a new batch of WGA gDNA which is not titrated to specific Ct score

#### *Data to Support Positive PCR Proposal*

While conducting lot testing on sets of aliquots sent from Smith, *A. lumbricoides* exhibited fall out in 2/3 lots, with high Ct score ranges in 2/3 lots. This is summarized in Table 6 and illustrated in Figure A9.7.

Table A9.5 fall out numbers were calculated by using the validation cut offs. These are as follows: *N. americanus ≤31.6, T. trichiura ≤ 28.8, A. duodenale ≤ 32.7, A. lumbricoides ≤ 35.5.* Within these, there are 4 replicates of *A. lumbricoides* where there was no amplification; these Undetermined scores are distributed across all the lots.

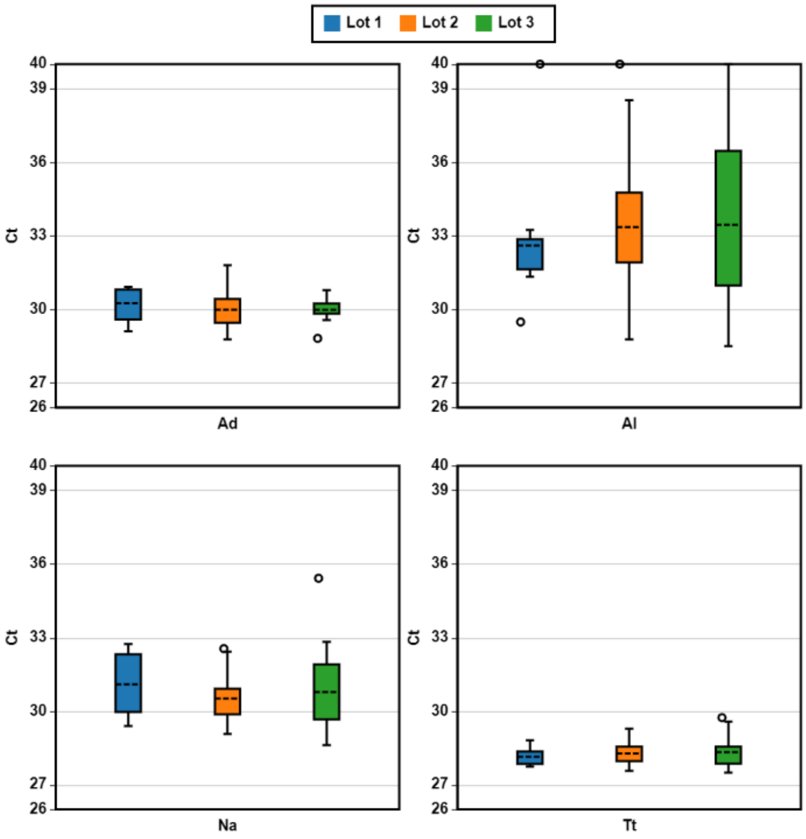

*Figure A9.7 14: WGA STH gDNA Lot Comparisons*

*Table A9.8: Positive PCR Lot Testing*

| Comparison of Averages Across New Lots (1:4 Dilution) | | | | | | | | | | | | | |
|---|---|---|---|---|---|---|---|---|---|---|---|---|---|
| **Lot #** | **Tech. Reps.** | **Na** | | | **Tt** | | | **Ad** | | | **Al** | | |
| | | *Avg.* | *σ* | *# Fall Out* | *Avg.* | *σ* | *# Fall Out* | *Avg.* | *σ* | *# Fall Out* | *Avg.* | *σ* | *# Fall Out* |
| **Lot 1** | **12** | **31.10** | 1.19 | 4 | **28.14** | 0.33 | 1 | **30.26** | 0.65 | 0 | **32.61** | 2.52 | 1 |
| **Lot 2** | **24** | **30.52** | 0.91 | 3 | **28.28** | 0.46 | 4 | **29.99** | 0.70 | 0 | **33.36** | 2.81 | 4 |
| **Lot 3** | **24** | **30.78** | 1.61 | 7 | **28.33** | 0.57 | 4 | **29.98** | 0.40 | 0 | **33.45** | 3.42 | 7 |
| **All Lots** | | **30.80** | 1.28 | **14** | **28.25** | 0.49 | **9** | **30.08** | 0.59 | **0** | **33.14** | 2.98 | **12** |

From these data, the unreliability of the current Positive PCR standard is seen. When the control performs well, the averages for each target are consistent, however, the positive PCR control should not allow for any fall out from the range.

In Table A9.5, it is clearly seen there is pervasive fall out across 3 of 4 targets, with each instance of fall out triggering a qPCR repeat.

A transition to plasmids will provide a more controlled spike-in quantity, in addition to increased confidence in the amount of each target that is spiked in.

If there is the desire to maintain use of WGA gDNA, considering this data, it can be clearly seen that there would be frivolous batch-wide repeats, due to the contrived, titrated nature of the controls. It is believed that this could be ameliorated if the control is no longer titrated.

It is also important to note, that while there was no fall out across the first 11 batches, all these batches were testing with Lot 1 of Positive PCR, which had much tighter ranges for all targets.

##### *Evidence to Support Positive PCR Proposal*

Other assays that have been developed for testing of STH species have used plasmids as the positive PCR control. By using a plasmid as a positive PCR control, it would more closely align with the current standard within the field.

While there has been concerns about increased contamination when using plasmids, but this contamination concern is only present when the plasmids are growing. This can be alleviated by only growing plasmids at Smith or Quantigen, and then exporting the plasmid DNA to the other sites for testing.
